# Genome-wide association studies of binge-eating behaviour and anorexia nervosa yield insights into the unique and shared biology of eating disorder phenotypes

**DOI:** 10.1101/2025.01.31.25321397

**Authors:** Jet D Termorshuizen, Helena L Davies, Sang-Hyuck Lee, Jessica K Dennis, Christopher Hübel, Jessica S Johnson, Yi Lu, Melissa A Munn-Chernoff, Triinu Peters, Baiyu Qi, Katherine E Schaumberg, Rebecca H Signer, Karanvir Singh, Abigail R ter Kuile, Laura M Thornton, Jiayi Xu, Shuyang Yao, Zeynep Yilmaz, Ruyue Zhang, Johan Zvrskovec, Mohamed Abdulkadir, Ziada Ayorech, Elizabeth C Corfield, Alexandra Havdahl, Kristi Krebs, Taralynn M Mack, Maria Niarchou, Teemu Palviainen, Julia M Sealock, Jessica H Baker, Andrew W Bergen, Andreas Birgegård, Vesna Boraska Perica, Katharina Bühren, Roland Burghardt, Matteo Cassina, Giovanni Castellini, Enrico Collantoni, James J Crowley, Unna N Danner, Franziska Degenhardt, Janiece E DeSocio, Christian Dina, Monika Dmitrzak-Węglarz, Laramie E Duncan, Karin M Egberts, Lenka Foretova, Ina Giegling, Fragiskos Gonidakis, Scott D Gordon, Jakob Grove, Sébastien Guillaume, Jerry D Guintivano, Annette M Hartmann, Konstantinos Hatzikotoulas, Stefan Herms, Hartmut Imgart, Susana Jiménez-Murcia, Antonio Julià, Gursharan Kalsi, Deborah Kaminská, Leila J Karhunen, Hannah L. Kennedy, Kirsty M Kiezebrink, Theresa Kolb, Janne T Larsen, Dong Li, Lisa Lilenfeld, Mario Maj, Morten Mattingsdal, Paolo Meneguzzo, Allison L Miller, Karen S Mitchell, Alessio Maria Monteleone, Catherine M Olsen, Leonid Padyukov, Richard Parker, Michaela A. Pettie, Dalila Pinto, Anu Raevuori, Samuli Ripatti, Marion E Roberts, Paolo Santonastaso, Androula Savva, Ulrike H Schmidt, Alexandra Schosser, Jochen Seitz, Lenka LS Slachtova, Agnieszka Slopien, Sandro Sorbi, Peter S Straub, Jin P Szatkiewicz, Friederike I Tam, Elena Tenconi, Alfonso Tortorella, Artemis Tsitsika, Annemarie A van Elburg, Gudrun Wagner, Hunna J Watson, Roger AH Adan, Lars Alfredsson, Ole A Andreassen, Helga Ask, Anders D. Børglum, Harry A Brandt, David Collier, Steven Crawford, Scott Crow, Lea K Davis, Martina de Zwaan, George Dedoussis, Danielle M Dick, Stefan Ehrlich, Xavier Estivill, Angela Favaro, Fernando Fernández-Aranda, Krista Fischer, Andreas J Forstner, Philip Gorwood, Hakon Hakonarson, Johannes Hebebrand, Beate Herpertz-Dahlmann, Anke Hinney, James I Hudson, Craig Johnson, Jennifer Jordan, Allan S Kaplan, Jaakko Kaprio, Andreas FK Karwautz, Martien JH Kas, Walter H Kaye, James L Kennedy, Martin A Kennedy, Anna Keski-Rahkonen, Youl-Ri Kim, Kelly L Klump, Mikael Landén, Stéphanie Le Hellard, Kelli Lehto, Qingqin S. Li, Jolanta Lissowska, Jurjen J. Luykx, Sarah L Maguire, Nicholas G Martin, Manuel Mattheisen, Sarah E Medland, Philip Mehler, Nadia Micali, James E Mitchell, Palmiero Monteleone, Preben Bo Mortensen, Benedetta Nacmias, Roel A Ophoff, Hana Papezova, Nancy L Pedersen, Liselotte V Petersen, Luisa S Rajcsanyi, Nicolas Ramoz, Ted Reichborn-Kjennerud, Valdo Ricca, Stephan Ripke, Dan Rujescu, Filip Rybakowski, Stephen W Scherer, Margarita CT Slof-Op ‘t Landt, Howard Steiger, Patrick F Sullivan, Beata Świątkowska, Eric F van Furth, Tracey D Wade, Thomas Werge, David C Whiteman, D. Blake Woodside, Ya-Ke Wu, Stephan Zipfel, Eating Disorders Working Group of the Psychiatric Genomics Consortium, Estonian Biobank (EstBB), Cynthia M Bulik, Laura M Huckins, Gerome Breen, Jonathan RI Coleman

## Abstract

Eating disorders—including anorexia nervosa (AN), bulimia nervosa, and binge eating disorder—are clinically distinct but exhibit symptom overlap and diagnostic crossover. Genomic analyses have mostly examined AN. We conducted the first genomic meta-analysis of binge-eating behaviour (BE; 39,279 cases, 1,227,436 controls), alongside new analyses of AN (24,223 cases, 1,243,971 controls) and its subtypes (all European ancestries). We identified six loci associated with BE, including loci associated with higher body mass index (BMI) and impulse-control behaviours. AN GWAS yielded eight loci, validating six loci. Subsequent polygenic risk score analysis demonstrated an association with AN in two East Asian ancestry cohorts. BE and AN exhibited similar positive genetic correlations with psychiatric disorders, but opposing genetic correlations with anthropometric traits. Most of the genetic signal in BE and AN was not shared with BMI. We have extended eating disorder genomics beyond AN; future work will incorporate multiple diagnoses and global ancestries.

---

Eating disorders include anorexia nervosa (AN), bulimia nervosa, and binge eating disorder, among others. They are diagnostically distinct, yet show considerable overlap in symptoms, as reflected in diagnostic migration over time ^1–3^. AN is characterised by low weight, fear of weight gain, and an inability to recognise the seriousness of the low weight. It has two subtypes, which both exhibit low weight achieved by caloric restriction and increased energy expenditure. In the binge eating/purging subtype (AN-BP), this is coupled with binge eating (BE) and/or purging behaviours, whereas the restricting subtype (AN-R) lacks these features. Bulimia nervosa occurs in individuals at normal or high weight and is characterised by the combination of BE and compensatory behaviours (e.g., fasting, self-induced vomiting, laxative use, diuretic use). Binge eating disorder shares the BE component of bulimia nervosa and also occurs at both normal and high weights, but lacks recurrent compensatory behaviours ^1^.

Genome-wide association studies (GWASs) of eating disorders have focused primarily on AN ^4–7^, in part due to its elevated mortality ^8^, with only a few GWASs of other eating disorder-related phenotypes to date ^9,10^. The most recent AN GWAS ^6^ included 16,992 cases and identified eight genome-wide significant loci. Genetic correlations based on common single nucleotide polymorphisms (SNP-*r*_*g*_s) showed high correlations with other psychiatric disorders, and suggested that metabolic and anthropometric factors might also underlie AN pathophysiology ^6,7^. The metabolic aspect of AN is reflected by a positive SNP-*r*_*g*_ with high-density lipoprotein cholesterol and negative SNP-*r*_*g*_s with insulin resistance, leptin, and type 2 diabetes. Importantly, these SNP-*r*_*g*_s were independent of body mass index (BMI), a significant finding given that low BMI is a defining feature of AN.

To fully understand the genetic landscape of eating disorders, it is essential to advance beyond AN. A substantial genetic correlation between AN and bulimia nervosa has been shown in family and twin studies ^11,12^, suggesting that these phenotypes share genetic risk. This may partially reflect the presence of BE as a transdiagnostic symptom, common both to bulimia nervosa and to AN-BP. Psychiatric diagnoses defined by the DSM and ICD have been criticised for failing to capture patient experiences or biological realities, given that disorders often exist on a continuum, overlap, co-occur, present heterogeneously, and morph over time ^13^. Studying only those who meet diagnostic thresholds can bias research toward the most severe cases and excludes individuals who have significant pathology but do not meet strict diagnostic criteria. Alternative, symptom-based approaches have been proposed to complement diagnostic approaches and advance the field of eating disorders ^14^. Here, we present the first GWAS of BE across multiple cohorts. We additionally conducted an AN GWAS meta-analysis with augmented sample size and increased statistical power, and the first GWASs of explicitly defined AN subtypes (AN-R and AN-BP). We identified genetic commonalities and differences between BE and AN.

## RESULTS

### Summary of phenotypes

We operationalised five phenotypes (**Table 1**): broadly-defined BE (BE-BROAD) and narrowly-defined BE (BE-NARROW), and AN, AN-R, and AN-BP. We primarily report results for BE-BROAD (which captures the common genetic component of BE with greater statistical power than BE-NARROW; Methods) and for AN. Analyses of BE-NARROW, AN-R, and AN-BP are reported in Supplementary Results. We contrast our BE-BROAD results with those of a previous publication that assessed a model-derived BE phenotype in the Million Veteran Program ^10^ (Supplementary Material).

**Table 1:** Phenotype definitions. Diagnostic codes are shown with coding system in round brackets, and equivalent codes in square brackets. Control definitions are shown in order of preference. Footnotes: *If assessed with terms such as “psychological overeating”, “compulsive eating”, etc., these individuals will not be included as BE-BROAD cases. **If Other Specified Feeding or Eating Disorder or Eating Disorder Not Otherwise Specified is diagnosed, these are included only if clearly stated ‘subthreshold bulimia nervosa’ or ‘subthreshold binge eating disorder’.

| Phenotype | Inclusion criteria:<br>Self-report or questionnaires | Inclusion criteria:<br>Diagnostic codes |
| --- | --- | --- |
| BE-BROAD<br>*Binge eating broadly defined*<br>(incl. BE-NARROW) | 1. Eating an unusually large amount of food in a short period of time with loss of control. Frequency and duration criteria not required, or;<br>2. If assessed by a single item like “Have you ever experienced binge eating?”*, or;<br>3. A self-reported diagnosis of bulimia nervosa or binge eating disorder, confirmation by healthcare professional not required**. | 307.51<br>(ICD-9, DSM-III-R, DSM-IV)<br>F50.2, F50.3<br>(ICD-10)<br>307.51 [F50.2]<br>(DSM-5)<br>307.51 [F50.3]<br>(DSM-5)<br>F50.81<br>(ICD-10-CM) |
| BE-NARROW<br>*Binge eating narrowly defined* | 1. Eating an unusually large amount of food in a short period of time with loss of control, and these episodes occurred on average at least once a week for at least three months, or;<br>2. A clinical diagnosis of bulimia nervosa or binge eating disorder via hospital/register records or a structured clinical interview, or;<br>3. A self-reported diagnosis of bulimia nervosa or binge eating disorder with confirmation by a healthcare professional. | 307.51<br>(ICD-9, DSM-III-R, DSM-IV)<br>F50.2<br>(ICD-10)<br>F50.81<br>(ICD-10-CM)<br>307.51 [F50.2]<br>(DSM-5) |
| AN<br>*AN - no subtype specified*<br>(incl. AN-R and AN-BP) | 1. A clinical diagnosis of AN / AN binge eating/purging subtype / AN restricting subtype via hospital/register records, structured clinical interviews, or online questionnaires based on standardised criteria, or;<br>2. A self-reported diagnosis of AN / AN binge eating/purging subtype / AN restricting subtype, confirmation by healthcare professional not required.<br>Note: atypical AN (without significantly low body weight) was not specifically excluded from our analyses, but has been historically ill-defined and was not ascertained for in most cohorts. As such, our AN data primarily reflect typical AN. | 306.50<br>(ICD-8)<br>307.1<br>(ICD-9, DSM-III-R, DSM-IV)<br>F50.0<br>(ICD-10)<br>F50.1<br>(ICD-10)<br>307.1 [F50.1]<br>(DSM-5) |
| AN-R<br>*AN - restricting subtype* | 1. A clinical diagnosis of AN restricting subtype via hospital/register records, structured clinical interviews, or online questionnaires based on standardised criteria, or;<br>2. A self-reported diagnosis of AN restricting subtype, confirmation by healthcare professional not required | F50.01<br>(ICD-10)<br>307.1 [F50.01]<br>(DSM-5) |
| AN-BP<br>*AN - binge eating/ purging subtype* | 1. A clinical diagnosis of AN binge eating/purging subtype via hospital/register records, structured clinical interviews, or online questionnaires based on standardised criteria, or;<br>2. A self-reported diagnosis of AN binge eating/purging subtype, confirmation by healthcare professional not required | F50.02<br>(ICD-10)<br>307.1 [F50.02]<br>(DSM-5) |
| Controls | 1. No history of any eating disorder and no history of binge eating (broadly or narrowly defined), or;<br>2. No history of any eating disorder, or;<br>3. Unscreened (i.e., these individuals may have had an eating disorder or binge eating). |  |

### GWAS meta-analyses

Our BE-BROAD GWAS included 17 European-ancestry datasets with 39,279 cases and 1,227,436 controls, assessing 6,244,919 common (minor allele frequency≥1%), high-confidence (imputation INFO score>0.6), autosomal single nucleotide polymorphisms (SNPs; Supplementary Table 1). Conditional and joint analyses confirmed six independently associated loci (Figure 1; Table 2; Supplementary Figures 1-6; Supplementary Table 2). The liability-scale SNP-based heritability for BE-BROAD was 5% (SE=0.4%, assuming population prevalence of 4.5% ^15^). The intercept was 1.03, significantly >1 and the attenuation ratio was 0.14 (SE=0.04) (Supplementary Table 3). While an intercept >1 can indicate confounding, this ratio suggests that inflation was primarily due to polygenicity ^16^.

**Figure 1:**
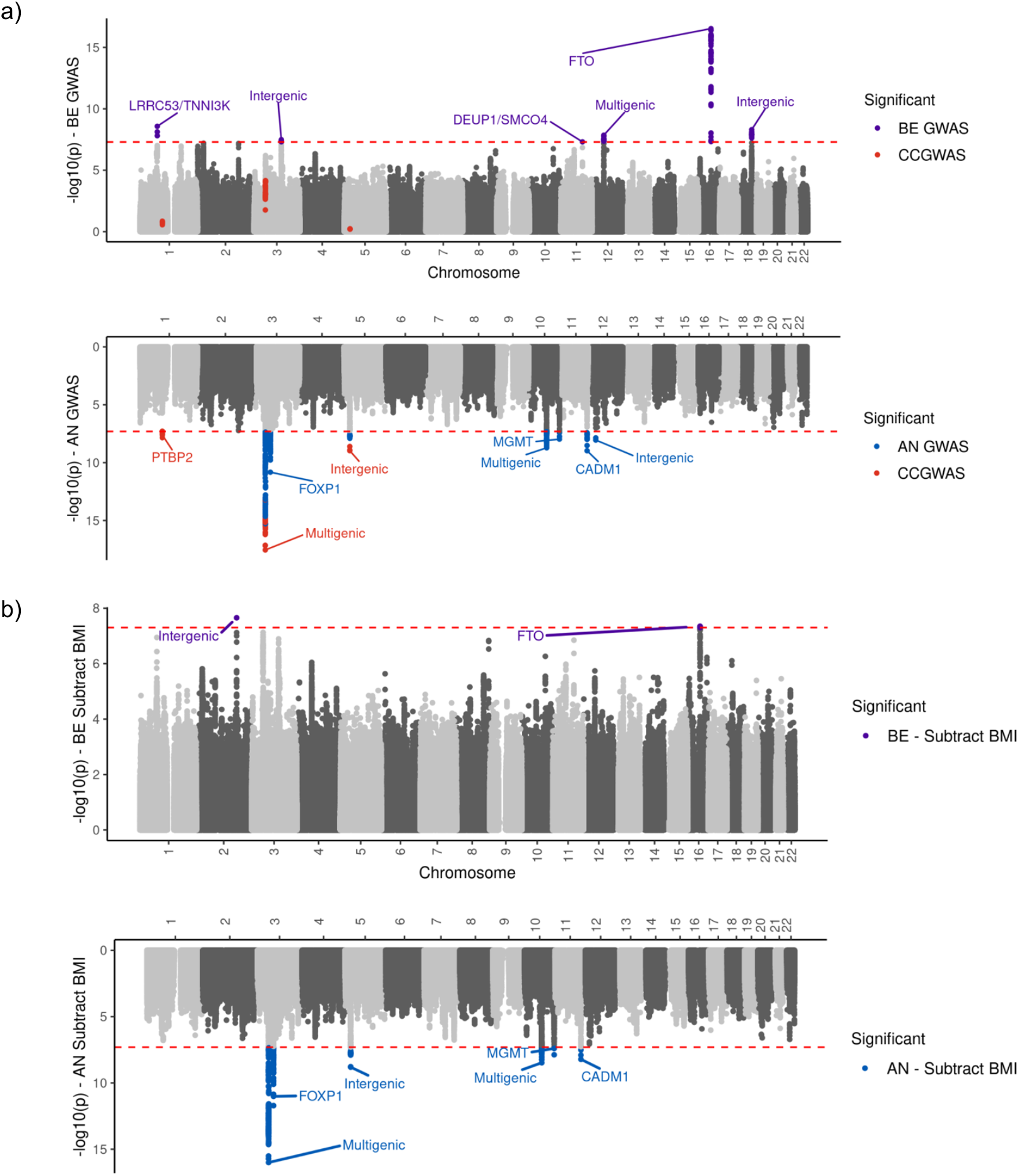
Miami plots showing results from the BE-BROAD (top) and AN (bottom) meta-analyses. The dotted red line is the genome-wide significance threshold (*P*≤5x10^-8^). **a**. Main GWAS analyses, with variants reaching genome-wide significance coloured in blue if significant in the anorexia nervosa GWAS and in purple if significant in the binge eating broad GWAS. Variants reaching genome-wide significance in case-case GWAS of BE-BROAD vs AN are coloured in red. **b**. GWAS-by-subtraction analyses, showing results from the non-BMI genetic component. Variants reaching genome-wide significance coloured in blue for anorexia nervosa non-BMI component and purple in binge eating non-BMI component.

For AN, we identified eight independently associated loci in 26 European-ancestry datasets with 24,223 cases and 1,243,971 controls, across 6,926,820 common, high-confidence, autosomal SNPs (Figure 1; Table 2; Supplementary Figures 7-14; Supplementary Tables 1 and 2). Six of these loci were detected in a previous AN GWAS ^6^ and two were newly identified. Three previously significant loci (on chromosome 2 and 3 from Watson et al ^6^, and on chromosome 12 from Duncan et al ^7^) did not reach genome-wide significance (*P*=2x10^-7^–6x10^-7^). The liability-scale SNP-based heritability was 13% (SE=0.7%, assuming population prevalence of 1.5% ^17^), with an intercept of 1.02, significantly >1 and an attenuation ratio of 0.07 (SE=0.03), suggesting inflation was largely due to polygenicity (Supplementary Table 3).

**Table 2:** Association statistics for genome-wide significant loci for BE-BROAD and AN. Loci are numbered sequentially within-analysis from chromosome 1 to chromosome X. Base pair start and end positions correspond to hg19. Odds ratios (OR) are given relevant to the ALT allele. Genes are listed if at least one transcript in GENCODE version 47 lies at least partially within the locus, with >3 genes defined as multigenic.

| Phenotype | Locus | CHR | Locus Start | Locus End | Lead SNP | P | REF/<br>ALT | OR | SE | ALT<br>Freq<br>Cases | ALT Freq<br>Controls | INFO | Previously<br>associated | Genes |
| --- | --- | --- | --- | --- | --- | --- | --- | --- | --- | --- | --- | --- | --- | --- |
| BE-BROAD | 1 | 1 | 74,977,295 | 75,014,362 | rs7541513 | 2.65x10<sup>-9</sup> | G/A | 1.05 | 0.009 | 0.427 | 0.425 | 0.952 | - | *LRRC53 / TNNI3K* |
|  | 2 | 3 | 117,420,697 | 117,953,697 | rs1456193 | 3.25x10<sup>-8</sup> | T/C | 1.06 | 0.011 | 0.808 | 0.802 | 0.996 | - | Intergenic |
|  | 3 | 11 | 93,171,140 | 93,231,940 | rs2925354 | 4.91x10<sup>-8</sup> | G/A | 0.93 | 0.013 | 0.107 | 0.112 | 0.995 | - | *DEUP1 / SMCO4* |
|  | 4 | 12 | 50,169,080 | 50,285,780 | rs7953534 | 1.40x10<sup>-8</sup> | G/A | 1.05 | 0.009 | 0.351 | 0.333 | 0.985 | - | Multigenic |
|  | 5 | 16 | 53,797,904 | 53,845,494 | rs11642015 | 2.97x10<sup>-17</sup> | C/T | 1.07 | 0.009 | 0.429 | 0.417 | 1.000 | - | *FTO* |
|  | 6 | 18 | 57,730,978 | 57,914,978 | rs66723169 | 5.02x10<sup>-9</sup> | C/A | 1.06 | 0.010 | 0.254 | 0.209 | 0.995 | - | Intergenic |
| AN | 1 | 1 | 96,700,455 | 97,285,455 | rs10747478 | 1.43x10<sup>-8</sup> | T/G | 0.94 | 0.011 | 0.570 | 0.587 | 0.998 | Yes<sup>20</sup> | *PTBP2* |
|  | 2 | 3 | 47,501,450 | 51,741,450 | rs113519699 | 2.86x10<sup>-18</sup> | A/C | 1.18 | 0.019 | 0.102 | 0.087 | 0.997 | Yes<sup>20</sup> | Multigenic |
|  | 3 | 3 | 70,602,234 | 71,074,242 | rs9310201 | 3.12x10<sup>-12</sup> | A/T | 1.08 | 0.011 | 0.427 | 0.408 | 0.982 | Yes<sup>20</sup> | *FOXP1* |
|  | 4 | 5 | 24,868,440 | 25,300,440 | rs6872919 | 1.07x10<sup>-9</sup> | A/C | 1.07 | 0.011 | 0.570 | 0.532 | 0.997 | Yes<sup>20</sup> | Intergenic |
|  | 5 | 10 | 75,850,972 | 76,524,972 | rs10740439 | 1.91x10<sup>-9</sup> | C/T | 0.93 | 0.012 | 0.711 | 0.716 | 0.994 | No | Multigenic |
|  | 6 | 10 | 131,274,055 | 131,459,255 | rs12762024 | 3.28x10<sup>-9</sup> | G/C | 1.07 | 0.011 | 0.447 | 0.455 | 0.995 | Yes<sup>20</sup> | *MGMT* |
|  | 7 | 11 | 114,997,256 | 115,284,956 | rs6589488 | 9.80x10<sup>-9</sup> | A/T | 0.91 | 0.016 | 0.846 | 0.875 | 0.990 | Yes<sup>20</sup> | *CADM1* |
|  | 8 | 12 | 17,635,314 | 17,965,314 | rs12817084 | 8.93x10<sup>-9</sup> | T/C | 1.10 | 0.011 | 0.115 | 0.117 | 0.991 | No | Intergenic |

We also conducted analyses on chromosome X for a subset of studies with available data (Supplementary Table 4). No genome-wide significant loci were identified for BE-BROAD nor AN, although one genome-wide significant locus was identified for BE-NARROW (Supplementary Results, Supplementary Figure 15, Supplementary Table 5).

Given known sex differences in eating disorders, and mostly female cases in our data (96% in BE-BROAD, 94% in AN), we carried out female-only GWASs as sensitivity analyses. Results were similar to the main analyses, with differences attributable to the reduction in sample size (Supplementary Results, Supplementary Table 6).

### Genetic relationship between eating phenotypes and other traits

We assessed the genetic similarity of BE-BROAD and AN through examining their SNP-*r*_*g*_ with each other and with other traits, via univariate and bivariate causal mixture modelling (“MiXeR analysis”) and via case-case GWAS ^18,19^. The SNP-*r*_*g*_ between BE-BROAD and AN was 0.46 (SE=0.04, *P=*3.44x10^-30^), indicating moderate genetic overlap (Supplementary Table 7). MiXeR analysis resulted in a similar estimate of SNP-*r*_*g*_ (0.49, SE=0.01). Bivariate MiXeR indicated an overlap of 2654 causal variants between BE-BROAD and BE-AN (83% of all BE-BROAD causal variants) with highly concordant effects (95%; Figure 2a). However, the bivariate MiXeR model was not distinguishable from the minimal possible overlap model, which requires an overlap >2450 variants given the genetic correlation observed (Supplementary Results, Supplementary Table 8). Case-case GWAS leverages a genetic distance measure representing the average squared difference in allele frequency at causal SNPs ^18^. The genetic distance was highest between individuals with AN and controls (0.46), similar to that between case groups (0.40), whereas the genetic distance between individuals with BE-BROAD and controls was smaller (0.25, Figure 2b). We identified case-divergent loci on chromosomes 1, 3, and 5, overlapping with independent loci identified in the AN GWAS (Figure 1), suggesting that some loci differentiating AN from controls also differentiate AN from BE-BROAD.

**Figure 2:**
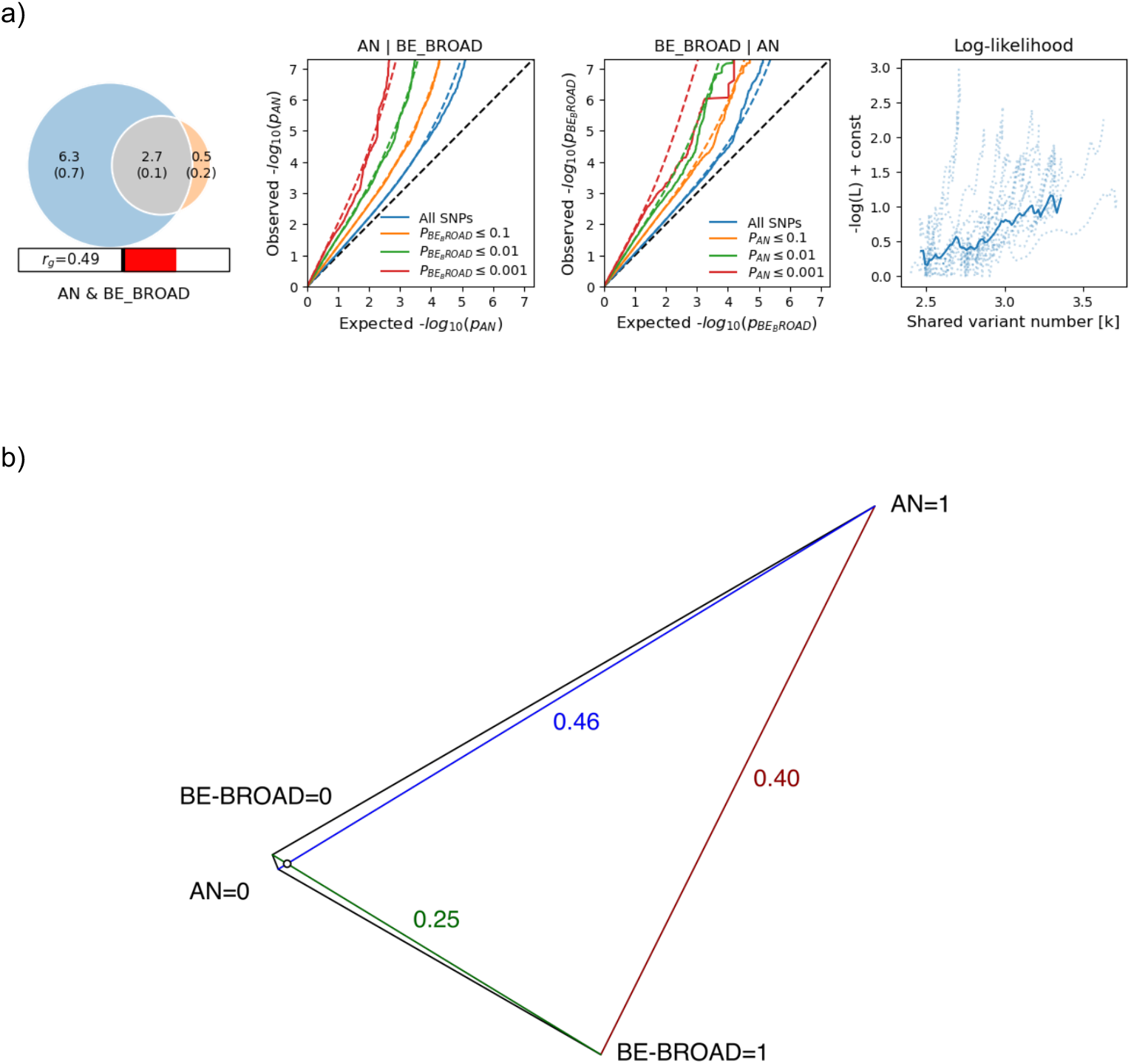
**a)** Bivariate causal mixture modelling results from MiXeR. (Left) Venn diagram showing overlap of putative causal variants in AN (blue) and BE BROAD (orange), demonstrating high overlap (grey), and moderate genetic correlation (bar beneath). (Middle) Q-Q plots of variant effects for (left) AN conditional on BE BROAD and (right) BE BROAD conditional on AN, demonstrating enrichment for associations with each trait conditional on the other. (Right) Log-likelihood plot of model fit under differing models, from the minimal overlap model (leftmost) to the full overlap model (rightmost), showing best model fit (lowest y-axis value) is not distinguishable from the minimal overlap. **b)** Genetic distance between cases and controls of BE-BROAD and AN estimated by case-case GWAS. The genetic distance,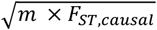 is calculated by taking the square root of the product of *m*, the number of independent causal variants estimated here as 10,000 based on the polygenic nature of BE-BROAD and AN, and *F*_*ST*,*casual*_, the average normalized squared differences in allele frequencies, derived based on the SNP-based heritabilities, genetic correlations, and population prevalences of the two traits.

We used LDSC to calculate pairwise SNP-*r*_*g*_ for both BE-BROAD and AN with 225 traits including psychiatric, personality, metabolic, and anthropometric traits (Table 3, Supplementary Table 9). We generally observed positive SNP-*r*_*g*_ of BE-BROAD with psychiatric traits and anthropometric phenotypes, except for persistent thinness and pubertal growth which displayed negative SNP-*r*_*g*_. In contrast, significant SNP-*r*_*g*_ with metabolic traits were absent for BE-BROAD except for BMI-adjusted fasting insulin. We validated and extended previously observed SNP-*r*_*g*_ patterns with AN ^6^ (Table 3, Supplementary Table 9). We found positive SNP-*r*_*g*_ across psychiatric disorders, as well as with neuroticism, educational attainment, and physical activity. We observed negative SNP-*r*_*g*_ across anthropometric traits, and notably, a non-significant SNP-*r*_*g*_ with persistent thinness. Metabolic SNP-*r*_*g*_ mirrored Watson et al., with predominantly negative SNP-*r*_*g*_, except for total cholesterol in high-density lipoprotein ^6^.

**Table 3:** A representative subset of significant genetic correlations of BE-BROAD and AN with external traits. *Note*. The full set of genetic correlations can be found in Supplementary Table 9. PMID = PubMed ID of GWAS for external trait, BMI = body mass index, adj = adjusted, ADHD = attention deficit/hyperactivity disorder, MDD = major depressive disorder, OCD = obsessive-compulsive disorder, PTSD = post-traumatic stress disorder, AUDIT-P = Alcohol Use Disorder Identification Test Problem items.

| Trait | Category | PMID | rg<br>(AN) | SE<br>(AN) | P<br>(AN) | rg<br>(BE) | SE<br>(BE) | P<br>(BE) |
| --- | --- | --- | --- | --- | --- | --- | --- | --- |
| BMI | Anthropometric | 30239722 | -0.31 | 0.020 | **2.37x10<sup>-56</sup>** | 0.36 | 0.032 | **1.94x10<sup>-28</sup>** |
| Body fat % | Anthropometric | 30593698 | -0.35 | 0.028 | **2.71x10<sup>-36</sup>** | 0.20 | 0.042 | **2.65x10<sup>-6</sup>** |
| Fat free mass | Anthropometric | 30593698 | -0.15 | 0.022 | **2.93x10<sup>-11</sup>** | 0.30 | 0.034 | **6.92x10<sup>-19</sup>** |
| Fat mass | Anthropometric | 31852892 | -0.32 | 0.023 | **3.63x10<sup>-43</sup>** | 0.31 | 0.035 | **1.36x10<sup>-18</sup>** |
| Persistent thinness | Anthropometric | 30677029 | 0.11 | 0.074 | 0.123 | -0.46 | 0.110 | **2.37x10<sup>-5</sup>** |
| Pubertal growth (height diff, 8 years-adult) | Anthropometric | 23449627 | -0.03 | 0.053 | 0.606 | -0.37 | 0.079 | **3.08x10<sup>-6</sup>** |
| Severe early-onset obesity | Anthropometric | 30677029 | -0.11 | 0.059 | 0.0587 | 0.48 | 0.080 | **1.40x10<sup>-9</sup>** |
| Waist-hip ratio | Anthropometric | 30239722 | -0.25 | 0.021 | **1.72x10<sup>-34</sup>** | 0.19 | 0.034 | **1.37x10<sup>-8</sup>** |
| Waist-hip ratio (BMI-adj) | Anthropometric | 30239722 | -0.09 | 0.020 | **9.33x10<sup>-6</sup>** | -0.05 | 0.033 | 0.119 |
| Fasting insulin (age- & sex-adj) | Metabolism | 33402679 | -0.30 | 0.048 | **2.79x10<sup>-10</sup>** | 0.15 | 0.069 | 0.0292 |
| Fasting insulin (BMI-adj) | Metabolism | 34059833 | -0.17 | 0.035 | **9.62x10<sup>-7</sup>** | -0.29 | 0.047 | **6.29x10<sup>-10</sup>** |
| HbA1c | Metabolism | 28898252 | -0.14 | 0.042 | 6.00x10<sup>-4</sup> | -0.03 | 0.053 | 0.5242 |
| HbA1c (BMI-adj) | Metabolism | 34059833 | -0.16 | 0.034 | **5.22x10<sup>-6</sup>** | 0.01 | 0.044 | 0.765 |
| Type 2 diabetes | Metabolism | 30297969 | -0.18 | 0.024 | **1.76x10<sup>-14</sup>** | 0.11 | 0.040 | 6.10x10<sup>-3</sup> |
| ADHD | Psychiatric | 36702997 | 0.07 | 0.029 | 0.0126 | 0.31 | 0.042 | **2.20x10<sup>-13</sup>** |
| Autism | Psychiatric | 32747698 | 0.17 | 0.044 | **1.00x10<sup>-4</sup>** | 0.29 | 0.057 | **3.96x10<sup>-7</sup>** |
| Bipolar disorder | Psychiatric | 34002096 | 0.25 | 0.027 | **3.80x10<sup>-21</sup>** | 0.23 | 0.035 | **4.22x10<sup>-11</sup>** |
| Insomnia | Psychiatric | 30804565 | 0.10 | 0.033 | 2.90x10<sup>-3</sup> | 0.21 | 0.044 | **2.02x10<sup>-6</sup>** |
| MDD | Psychiatric | 39814019 | 0.30 | 0.025 | **1.04x10<sup>-33</sup>** | 0.39 | 0.031 | **1.35x10<sup>-36</sup>** |
| OCD | Psychiatric | 28761083 | 0.48 | 0.069 | **2.85x10<sup>-12</sup>** | 0.08 | 0.081 | 0.322 |
| Probable anxiety diagnosis | Psychiatric | 31748690 | 0.28 | 0.039 | **1.42x10<sup>-12</sup>** | 0.32 | 0.060 | **1.03x10<sup>-7</sup>** |
| PTSD | Psychiatric | 33510476 | 0.11 | 0.049 | 0.0274 | 0.24 | 0.062 | **1.00x10<sup>-4</sup>** |
| Schizophrenia | Psychiatric | 35396580 | 0.24 | 0.021 | **4.85x10<sup>-30</sup>** | 0.23 | 0.033 | **7.04x10<sup>-12</sup>** |
| General risk tolerance | Behavioural | 30643258 | 0.00 | 0.028 | 0.991 | 0.21 | 0.040 | **6.76x10<sup>-8</sup>** |
| Loneliness | Behavioural | 31518406 | 0.13 | 0.032 | **4.96x10<sup>-5</sup>** | 0.37 | 0.044 | **4.14x10<sup>-17</sup>** |
| Physical activity | Behavioural | 36071172 | 0.19 | 0.034 | **5.87x10<sup>-8</sup>** | 0.09 | 0.046 | 0.0633 |
| Educational attainment (years) | Socio-demographic | 30038396 | 0.25 | 0.022 | **1.25x10<sup>-29</sup>** | 0.04 | 0.030 | 0.1417 |
| AUDIT-P | Substance use | 30336701 | 0.01 | 0.040 | 0.870 | 0.37 | 0.057 | **7.00x10<sup>-11</sup>** |
| Cannabis use disorder | Substance use | 33096046 | 0.02 | 0.046 | 0.610 | 0.30 | 0.063 | **2.78x10<sup>-6</sup>** |
| Heavy smoker | Substance use | 28166213 | -0.04 | 0.039 | 0.255 | 0.30 | 0.048 | **3.82x10<sup>-10</sup>** |
| Lifetime cannabis use | Substance use | 30150663 | 0.22 | 0.038 | **8.48x10<sup>-9</sup>** | 0.32 | 0.051 | **6.36x10<sup>-10</sup>** |

Next, we tested for significant differences between the SNP-*r*_*g*_ of BE-BROAD and SNP-*r*_*g*_ of AN with other traits (Figure 3, Supplementary Figure 16, Supplementary Table 10). Most psychiatric and behavioural traits and disorders showed similar SNP-*r*_*g*_ with BE-BROAD and AN. However, attention deficit/hyperactivity disorder (ADHD) showed greater SNP-*r*_*g*_ (difference *P=*1.06x10^-7^) with BE-BROAD than with AN, and obsessive-compulsive disorder showed greater SNP-*r*_*g*_ (*P=*4.79x10^-5^) with AN than with BE-BROAD. BE-BROAD was positively correlated with Alcohol Use Disorder Identification Test problem items, four smoking-related phenotypes, and general risk tolerance, while AN was not (*P*≤4.83x10^-7^). AN was negatively correlated with automobile speeding propensity, while BE-BROAD was 16 not (*P*=1.21x10^-4^).

**Figure 3:**
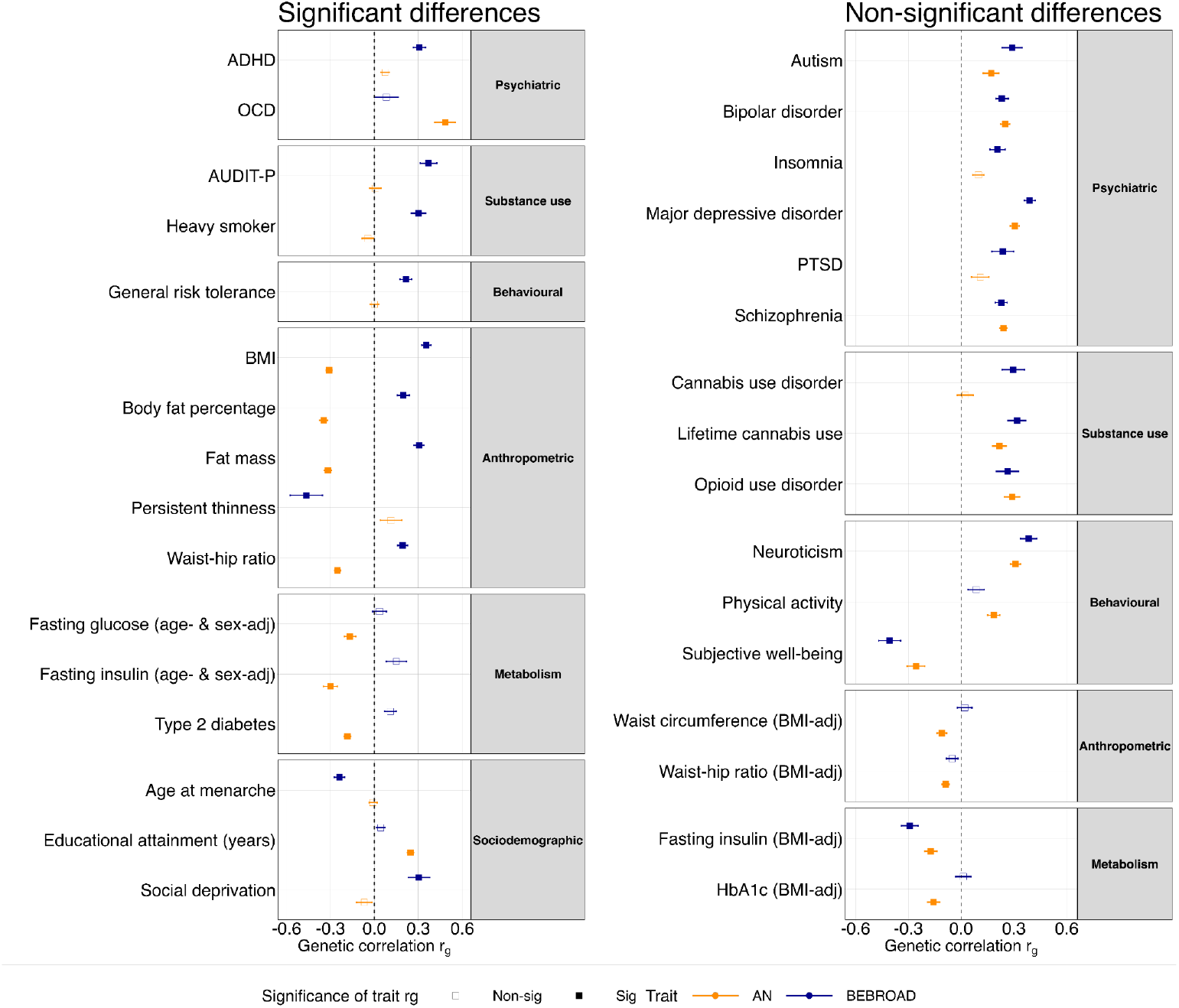
Genetic correlations (rg) of selected external traits with BE-BROAD and AN, split by those that differ significantly between the eating phenotypes (left) and those that do not (right). The rgs were computed by Linkage Disequilibrium Score Regression (LDSC). The rg estimates are indicated by the dots and standard errors are indicated by the lines on either side of each dot. rg estimates have been corrected for multiple testing via the Bonferroni method. Information about the summary statistics used in our analysis can be found in Supplementary Table 9. BE-BROAD = binge eating broad definition; AN = anorexia nervosa; MDD = major depressive disorder; PGC = Psychiatric Genomics Consortium; BMI = Body mass index; F = female; M = male; FFM = fat-free mass; AUDIT-P = Alcohol Use Disorder Identification Test problem items

BE-BROAD and AN also diverged in their associations with anthropometric and metabolic traits. For example, BE-BROAD was positively correlated with waist-to-hip ratio while AN was negatively correlated (*P=*2.05x10^-31^). BE-BROAD showed a stronger pattern of SNP-*r*_*g*_ with certain socio-demographic traits than AN (*P*<2x10^-4^), displaying negative SNP-*r*_*g*_s with age at menarche and age at first birth in females, and positive SNP-*r*_*g*_s with social deprivation and loneliness. In contrast, AN showed no significant SNP-*r*_*g*_ with these traits but was more strongly positively associated with educational traits such as college/university completion than was BE-BROAD.

The genetic signal in our BE-BROAD GWAS may partly be influenced by AN, given that 18% of our BE-BROAD cases had (known) AN (Supplementary Table 11). As a sensitivity analysis, we conducted an additional BE-BROAD GWAS, excluding cohorts that specifically focused on AN recruitment (Supplementary Results). The SNP-*r*_*g*_ between BE-BROAD and the reduced GWAS did not differ from unity (0.96, SE=0.07), but SNP-*r*_*g*_s with anthropometric traits were stronger in the reduced GWAS, suggesting that AN cases with BE 7 may mask BE-anthropometric genetic associations (Supplementary Figure 17, 8 Supplementary Table 12).

### Role of BMI genetics

The role of BMI in eating disorders is complex, with low BMI being pathognomonic of AN and individuals with binge eating disorder often being overweight ^1^. If the genetics of BMI were the only genetic cause of BE and AN (in opposite directions), then our GWASs would just be proxy GWASs for BMI. To assess this, we applied GWAS-by-subtraction to remove the genetic variance of BMI from BE-BROAD and from AN separately ^20^. We modelled a factor shared between each eating phenotype and BMI, and a *non-BMI* factor only loaded on by the eating phenotype. The shared factor explained 12% (SE=2.7%) of genetic variance in BE-BROAD, leaving 88% (SE=7.9%) accounted for by the *non-BMI* factor. In AN, the shared factor accounted for 10% (SE=1.4%) of genetic variance, leaving 90% (SE=5.5%) accounted for by the *non-BMI* factor. GWASs of the *non-BMI* factor for BE-BROAD and for AN generally resulted in larger p-values for lead SNPs, but larger effect sizes in the same direction as the original GWAS (Figure 1, Supplementary Table 13). In pairwise SNP-*r*_*g*_ analyses of the *non-BMI* component of BE-BROAD and AN, SNP-*r*_*g*_ with psychiatric disorders typically remained stable or slightly increased relative to the respective full GWAS, whereas SNP-*r*_*g*_ with anthropometric and metabolic traits were typically attenuated (Supplementary Figure 18, Supplementary Table 14).

We also conducted exploratory two-sample Mendelian randomisation (MR) analyses of each eating phenotype with BMI, testing causal effects in both directions. We used SNPs in linkage equilibrium as genetic instruments, with *P*<5x10^-6^ for BE-BROAD and AN, and *P*<5x10^-9^ for BMI ^21^. For each analysis, we used the full BE-BROAD or AN GWAS, and the GWAS of the respective *non-BMI* component (Supplementary Results, Supplementary Table 15). Significant results were found in both directions between increased BE-BROAD risk and increased risk for higher BMI. BE-BROAD was still associated with increased risk for higher BMI when using just the *non-BMI* component. When examining the effect of BMI on the *non-BMI* component of BE-BROAD, odds ratios (ORs) were consistently >1, but the different methods were not consistently significant. Significant results were found in both directions between increased risk of AN and decreased BMI. However, the *non-BMI* component of AN risk was not associated with BMI, and results were inconsistent across MR methods when examining the effect of BMI on the *non-BMI* component of AN.

### Genetically-regulated gene expression

We used S-PrediXcan ^22^ to identify predicted genetically-regulated gene expression associated with our phenotypes (Supplementary Figure 19; Supplementary Table 16). For BE-BROAD, two gene-tissue associations were significant at the experiment-wide threshold (*PRKAR2A*-Sigmoid colon and *KLHDC8B*-Heart, atrial appendage; *P*<8.32x10^-8^). In AN, we observed 300 experiment-wide significant gene-tissue associations (*P*<8.32x10^-8^) with 29 unique genes, predominantly from the gene-dense locus on chromosome 3:47-52Mb. Among the experiment-wide significant associations, 94 were in central nervous system tissues and 41 in gastrointestinal tissues. Within-tissue significant results for both phenotypes are described in the Supplementary Results.

We calculated cross-tissue predicted genetically-regulated gene expression using S-MultiXcan ^23^ to identify apparent tissue-specific associations that are better interpreted as cross-tissue (Supplementary Results, Supplementary Figure 20, Supplementary Table 17). Ten genes had significant (*P<*2.25x10^-6^) cross-tissue expression in BE-BROAD, four of which were identified as tissue-level associations (including *KLHDC8B* but not *PRKAR2A*). Similarly, 43 genes showed significant cross-tissue expression in AN, of which 23 were identified as tissue-level associations.

### Gene-level associations

We used MAGMA v1.10 ^24^ to conduct gene-wise analyses of the aggregate effect of SNPs mapped to protein-coding genes (Supplementary Table 18); gene-set analyses of groups of genes with shared functional, biological, or other characteristics (Supplementary Table 19); and gene-set analyses restricted to genes targeted by medications (Supplementary Table 20; Supplementary Results). We also examined enrichment of signal within drug-sets belonging to the same class of drugs (Supplementary Table 21; Supplementary Results). For BE-BROAD, MAGMA identified 22 genes significant after Bonferroni correction (*P<*2.59x10^-6^). *FTO* had the strongest association (*P=*2.8x10^-22^), while 9/22 (43%) of the Bonferroni-significant genes were linked to the gene-dense locus on chromosome 3:47-52Mb. The MAGMA gene set, drug set, and drug-class analysis yielded no significant results for BE-BROAD. For AN, MAGMA identified 76 significant genes (*P<*2.58x10^-6^). Most (54/76, 71%) were again mapped to chromosome 3:47-52Mb. Gene-set analysis identified enrichment in three biological pathways, related to the binding targets of RBFOX1-3 (RNA binding proteins that regulate neuronal alternative splicing ^25^), and to mutation-constrained genes with pLI>0.9. Drug-set analysis revealed no significant drug sets, but antimigraine preparations as a class were significantly associated with AN, consistent with previous associations between AN and migraine polygenic risk scores 24 (PRS) ^26^.

Genes implicated both through proximity (MAGMA) and through effects on gene expression (S-PrediXcan) are more likely to be functionally relevant than those implicated through proximity alone ^27^. Further restricting our MAGMA gene-wise results to genes that were at least tissue-level significant in S-PrediXcan resulted in seven prioritised genes across four loci in BE-BROAD, and 38 prioritised genes across eight loci in AN (Supplementary Table 22; Supplementary Results).

### Tissue and cell-type analyses

We used stratified LDSC ^28^ to estimate the enrichment of SNP-based heritability for BE-BROAD and AN among genes specifically expressed in GTEx human tissues (Supplementary Figure 21, Supplementary Table 23; Supplementary Results) and in cell types from the Human Brain Atlas (Supplementary Figure 22, Supplementary Table 24) ^29,30^. After accounting for multiple testing, no associations were significant.

### Polygenic prediction

We tested whether higher BE-BROAD and AN PRS were associated with a higher risk of BE-BROAD and AN, using a leave-one-cohort-out design in well-powered cohorts representative of the ascertainment methods employed in the study (Methods; Supplementary Table 25). Individuals with 1 SD higher BE-BROAD PRS had an average OR of 1.11 for BE-BROAD (average 95% confidence interval [CI] across cohorts: 1.08–1.14; *P* range: 2.38x10^-15^–0.022; Supplementary Figure 23). The average liability-scale variance explained was 0.32%. Individuals with 1 SD higher AN PRS had an average OR of 1.50 for AN (average 95% CI 1.42–1.59, all *P*<2x10^-16^; Supplementary Figure 23) and AN PRS explained 2.32% liability-scale variance on average.

We additionally tested the cross-ancestry prediction of AN in two East Asian ancestry cohorts from Korea and Japan. The European AN PRS was positively associated with AN in the combined Korean and Japanese cohort, with an OR of 1.36 (95% CI 1.09 – 1.70, *P*=0.0066), explaining 1.3% of the variance (assuming population prevalence of AN at 26 1.5%).

Next, we tested whether genetic risk of BE-BROAD and AN were shared across males and females, using a similar leave-one-cohort-out design. BE-BROAD PRS based on female-only GWAS were positively associated with BE-BROAD risk in males (OR 1.06– 1.20), but not all results were significant, possibly due to low case numbers in some cohorts. Similarly, AN PRS calculated based on female-only GWAS were positively associated with AN risk in males (OR 1.07–1.41), but the results were not consistently significant (Supplementary Figure 24, Supplementary Table 26; Supplementary Results).

We further assessed if BE-BROAD and AN PRS differed across BE and AN subgroups, comparing control individuals to (a) those with BE-BROAD only; (b) with both BE-BROAD and AN (BE+AN); and (c) with AN only. Overall, we found both BE-BROAD and AN PRS to be elevated in all subgroups compared to controls (*P≤*0.019). Among the subgroups, the BE+AN and BE-BROAD-only groups did not significantly differ on BE-BROAD PRS, and typically had a significantly higher BE-BROAD PRS than the AN-only group (Supplementary Figure 25). The BE+AN and AN-only groups did not significantly differ from each other on AN PRS, while the BE-BROAD-only group had a significantly lower AN PRS than both other groups in one cohort, but not in another (Supplementary Figure 25, Supplementary Table 27; Supplementary Results).

## DISCUSSION

In this first genome-wide analysis of BE, we implicate six genomic loci. These have been previously associated with smoking ^31^, risk-taking behaviour ^32^, and age at menarche ^33^. Overlap between BE and impulse-control behaviours was further observed in positive SNP-*r*_*g*_ with smoking, general risk tolerance, and problematic alcohol use. Loss of control is a key component of BE, and impulse-control behaviours have been associated with binge-type eating disorders clinically ^34–36^. Alongside significant polygenic overlap with a range of psychiatric disorders, our findings imply that BE shares genetic underpinnings with psychiatric disorders and impulse-control behaviours.

Loci associated with BE-BROAD have also been implicated in anthropometric traits, including a BMI-related signal near *FTO* ^21,37^ that was not associated with AN or its subtypes. The *FTO* locus has been studied extensively (summarised in Loos and Yeo ^38^), but it has been challenging to determine the causal mechanism that contributes to a high BMI ^38^. One study found that *FTO* was related to BE independent of BMI ^39^, and suggested that BE could mediate the pathway between *FTO* and a high BMI. Together with our results, this implies that the relationship between *FTO* and high BMI could result partly from binge-eating behaviours. Further research should examine whether broader disordered eating behaviours mediate the relationship between *FTO* and high BMI. Genetic correlations of BE with anthropometric traits and impulse-control behaviours mirror clinical observations in individuals with BE, and suggest the relationship of BE to these traits and behaviours is partly due to pleiotropy rather than being purely environmental or a consequence of BE itself.

For AN, we validated six previously identified loci, identified two new loci, and identified one locus for AN-R. The four single-gene loci identified in Watson et al. ^6^ remained genome-wide significant, suggesting that genes located in these regions—*CADM1, MGMT, FOXP1, PTBP2—*may warrant further investigation in the aetiology of AN ^40^. The AN-R-identified locus narrowly missed genome-wide significance in AN (*P*=5.89x10^-8^) and has previously been implicated in schizophrenia ^41^. The locus contains several genes, but only *DLX1* was indicated by both proximity-based and expression-based gene mapping. *DLX1* is differentially expressed in the brain and may be involved in several processes of neural development ^42^. Further studies are needed to confirm that *DLX1* is implicated in AN-R aetiology, given the multigenic nature of the locus. Our gene-level results should generally be viewed cautiously, as greater power is needed to effectively fine-map associated loci and link causal variants to genes.

Despite increasing our effective sample size for AN by 64% since our previous freeze ^6^, we identified only two new loci, and two previously implicated loci were no longer genome-wide significant. We speculate that this is because our new cohorts were primarily population-based and used more lenient case criteria compared to the clinical diagnoses and targeted AN-specific recruitment previously used ^43^. Consistent with this, AN PRS typically captured less variance in AN in new cohorts (Supplementary Results, Supplementary Table 25). Genetic signal also tends to become more heterogeneous as GWAS sample sizes increase ^44,45^. This is supported by our data. Although none of the lead SNPs associated with AN showed a heterogeneous effect (Supplementary Figures 7-14), variant heterogeneity statistics (I^2^) were higher in our AN GWAS than in the AN GWAS published by Watson et al (Supplementary Results, Supplementary Table 28) ^6^. Nonetheless, our AN GWAS measured variants with greater precision than that of Watson et al (average variant SE=0.0199 vs SE=0.0255 in Watson et al), indicating that power bincreased despite the increase in heterogeneity (Supplementary Results).

We have previously hypothesised that AN is a metabo-psychiatric disorder ^6^—this study yields for the first time the ability to investigate shared and distinct metabolic and psychiatric components across multiple eating phenotypes. BE-BROAD has typical genetic features of a psychiatric disorder, including significant SNP-*r*_*g*_ with psychiatric traits akin to previous findings ^6,7,26,46^. The SNP-*r*_*g*_ pattern for BE-BROAD is similar to that of AN, with notable exceptions. AN had a positive SNP-*r*_*g*_ with obsessive-compulsive disorder, whereas BE-BROAD showed no significant association. Conversely, BE-BROAD was positively genetically correlated with ADHD, whilst AN showed no significant association. However, there was a significant positive SNP-*r*_*g*_ between ADHD and *non-BMI* AN, suggesting that opposing SNP-*r*_*g*_s of AN and ADHD with BMI were previously masking this association.

We observed some key differences for SNP-*r*_*g*_s with non-psychiatric traits. BE-BROAD displayed a significantly stronger, negative SNP-*r*_*g*_ with age at menarche whilst AN showed no genetic overlap, consistent with previous research that showed early-onset AN was negatively associated with age at menarche, but typical-onset AN was not ^47^. Earlier age at menarche has been associated with increased impulse-associated traits like substance use and risky behaviour ^48^, and with BMI ^49^. Both impulsivity and BMI are often higher in those who binge eat ^1,34,35^. Observational studies find that later age of menarche is associated with AN ^48,50^; however, disentangling this from the effects of starvation and low body weight is difficult. Significant SNP-*r*_*g*_s between BE-BROAD and anthropometric traits were positive compared with the negative SNP-*r*_*g*_s observed in AN, consistent with previous research ^26,46^. We also validated our previous finding that significant SNP-*r*_*g*_s with AN are concentrated in metabolic-related traits ^6^, in contrast to BE-BROAD.

Eating disorders and their component features share genetic factors with anthropometric traits, but these effects act in opposite directions depending on the presentation. To investigate this further, we assessed BE-BROAD and AN after subtracting the genetic component each shares with BMI. The BMI component accounted for 12% of the genetic variance of BE-BROAD and 10% of AN, despite low BMI being a diagnostic requirement for AN. The low variance explained by the BMI component argues that neither our AN nor BE-BROAD GWAS are BMI GWAS by proxy. This is further supported by the *FTO* locus association with BE-BROAD, which is observed in AN-ascertained cohorts where affected individuals are likely to have lower BMI than unaffected individuals. Consistent with previous literature ^51^, we also found no evidence for a genetic overlap between persistent thinness and AN, indicating that the cognitive-behavioural component of AN distinguishes these low-BMI phenotypes on a genomic level.

However, BMI is a blunt measure of body composition ^52^, and its relationship with eating disorders is complicated, with evidence that the negative SNP-*r*_*g*_ between AN and BMI is driven by genetic enrichment for AN risk in females with lower BMI, rather than a uniform linear relationship across BMI ^53^. Both BE and AN are heterogeneous conditions, and subtypes may have differing relationships with BMI. Subtracting the BMI component from our AN and BE-BROAD GWASs affords tentative insights into the physiological aspects of these illnesses, but BMI itself has a partly behavioural aetiology ^28^. More sophisticated analyses with a wider range of body composition measurements and eating disorder presentations (including atypical AN in the normal or high BMI range) will provide deeper understanding.

We present the first GWAS meta-analysis of BE, accompanied by the largest investigation of AN and AN subtypes to date, with chromosome X analysed for all phenotypes. We have extended eating disorder genetic research beyond AN alone and demonstrated that BE is a psychiatric phenotype with distinctive genetic relationships with external traits. Nonetheless, readers should consider the following limitations. Our analyses only considered individuals of European genetically inferred ancestry, limiting their generalisability. We analysed PRSs in two small East Asian cohorts, but used European prevalence estimates to convert risk to the liability scale, potentially introducing bias. Differences in population structure, genetic architecture, and environmental factors, such as lower average BMI in Korean and Japanese populations ^54^, could influence the cross-ancestry prediction of AN. More GWAS and PRS studies in East Asian populations are required to improve the accuracy of genetic risk predictions and our understanding of AN genetics in these populations.

This limitation extends to other global ancestries and should be considered from both phenotypic and genotypic perspectives ^14^. The historic focus of eating disorder studies on females of European descent has influenced disease characterization, potentially leading to certain diagnostic criteria being over- or under-represented. Prioritizing variants common in European ancestry populations may prevent the genetic architecture of eating disorders being fully characterised. To address this, ongoing research efforts aim to enhance the generalisability and applicability of findings across a wider range of individuals through global data collection across all populations experiencing eating disorders.

Given the known diagnostic crossover between eating disorders, cohorts that only contributed cross-sectional diagnoses or symptoms are unable to account for later development of a disorder or symptom; for example, an individual with AN-R might go on to develop AN-BP ^2^. It is not possible to fully mitigate this limitation. However: (1) many of our cohorts include individuals beyond the typical age of diagnosis, making new diagnoses or diagnostic crossover less likely; (2) hidden diagnostic crossover likely contributes to false negatives and under-estimation of differences between GWAS, rather than introducing false positives ^55^; and (3) our previous work has shown that rates of diagnostic contamination (a similar effect to hidden diagnostic crossover) would need to occur at extremely high levels to affect locus discovery ^56^.

Despite our best efforts at harmonisation, heterogeneity might exist within the phenotypes due to factors such as distinct methods of ascertainment. Ideally, one approach to ascertainment would be used, such as structured diagnostic interviews within speciality clinics. However, using multiple ascertainment approaches can increase sample size and thus power despite the resulting heterogeneity. Fourth, our sample is mostly female, and results may not necessarily generalise to those who are not female. Finally, although strongly associated with BE-BROAD and AN risk, PRS remain very weak predictors of BE-BROAD and AN status. Combining PRS with other risk factors is needed to further improve prediction accuracy of BE-BROAD and AN.

Historically, binge-type eating disorders have been overshadowed by research on AN, despite their higher prevalence. This paper redresses that imbalance. We identified six genetic loci relating to broadly defined BE, validated six loci related to AN, reported two novel loci, and found one locus related to the restricting subtype of AN. We demonstrate that BE is genetically related to several other psychiatric phenotypes, with both shared and distinct patterns compared to AN, providing genetic substantiation of clinically-observed comorbidity patterns. The number of loci we implicate in BE-BROAD and AN is typical of early-phase GWAS studies ^57^, motivating GWAS meta-analyses of all major eating disorders (AN, bulimia nervosa, binge eating disorder, avoidant/restrictive food intake disorder) and transdiagnostic behaviours (e.g., BE, restriction) to drive variant discovery and further refine our understanding of shared and unique genetic features that distinguish presentations and inform a genetically guided nosology of eating disorders ^13^.

## Data availability

Individual level genotype data (except in countries where sharing of individual level data is prohibited by national law) and summary statistics used in this study are available to bona fide researchers working in collaboration with a member of the Eating Disorders Working Group of the PGC via secondary analysis proposals (https://pgc.unc.edu/for-researchers/data-access-committee/data-access-information/).

Summary statistics from this work will be made available via Figshare and the PGC website on publication (https://pgc.unc.edu/for-researchers/download-results/).

## Code availability

Code underlying this work will be made available on GitHub (https://github.com/psychiatric-genomics-consortium/PGC3_EAD) on publication.

## Author contributions

Specific author contributions are listed in Supplementary Table 29. All authors made substantial contributions to the conception or design of the work, or to the acquisition, analysis, or interpretation of data for the work, and critically reviewed the work for important intellectual content, gave final approval of the version to be published, and agreed to be accountable for all aspects of the work in ensuring that questions related to the accuracy or integrity of any part of the work are appropriately investigated and resolved.

## Conflicts of Interest

Susana Jiménez-Murcia and Fernando Fernández-Aranda have received consultancy and speaker honoraria from Novo Nordisk. Ole Andreassen is a consultant to Presicion Health and Cortechs.ai, and has received speaker’s honoraria from Lundbeck, Janssen, Lilly, and Otsuka. James Kennedy is a member of the Scientific Advisory Board for Myriad Neuroscience. Mikael Landén has received lecture honoraria from Lundbeck pharmaceuticals. Qingqin Li was an employee of Janssen Research & Development, LLC when the work was completed and holds company equity. Nadia Micali receives an honorarium as associate editor on the European Eating Disorders Review. Dan Rujescu served as consultant for Janssen, received honoraria from Boehringer-Ingelheim, Gerot Lannacher, Janssen and Pharmagenetix, received travel support from Angelini, Janssen and Schwabe, and served on advisory boards of AC Immune, Boehringer-Ingelheim, Roche and Rovi. Patrick Sullivan is a shareholder in Neumora Therapeutics and serves on the advisory board. Cynthia Bulik receives royalties from Pearson Education, Inc. and has served as a consultant for Orbimed. No other authors report conflicts of interest.

## Funding and ethics statements

The Psychiatric Genomics Consortium is supported by NIH grant R01 MH124851. Author and cohort-specific funding and ethics statements are provided in the Supplementary Materials.

## METHODS

### Ethics

The individual studies that comprise this investigation were conducted with advance approval by the appropriate Institutional Review Boards or equivalents at the individual study sites. We provide ethical statements for each study site in the Supplementary Note. This work represents a secondary analysis with data from these individual studies.

### Summary of cohorts

Detailed descriptions of the ascertainment and definition of cases and controls for each cohort is provided in Supplementary Table 1. Broadly, we identified cases and controls based on clinical diagnoses, diagnostic algorithms, and/or self-report questionnaires ^43^. We defined controls as individuals without a history of BE and without a history of an eating disorder, if possible. If this information was unavailable, unscreened controls were included assuming that the large control numbers would outweigh the impact of misclassified individuals in the control groups, given the collective lifetime prevalence of eating disorders 16 is ∼5% ^17^.

We included data from the Eating Disorders Working Group of the Psychiatric Genomics Consortium (PGC-ED; Supplementary Table 1). These data were restricted to individuals of European ancestry due to the limited availability of non-European ancestry samples at the time of analysis—we included two cohorts with individuals of East Asian ancestry for follow-up cross-ancestry polygenic risk score analyses. In total, we combined 27 European ancestry datasets totalling 14 previously analysed ^6^ and 13 new cohorts. Data from cohorts providing individual-level data (*n=*11) were combined with cohorts that contributed summary statistics (*n*=16; Supplementary Table 1). Detailed descriptions of each of the cohorts are provided in the Supplementary Note. We included data if the total number of cases for any phenotype prior to quality control was >100. If cases for an individual phenotype were <50, we excluded that phenotype from analyses.

We included five different phenotypes: broadly-defined BE (BE-BROAD, 39,279 cases), narrowly-defined BE (BE-NARROW, 15,175 cases), AN (24,145 cases), AN restricting subtype (AN-R, 2524 cases), and AN binge eating/purging subtype (AN-BP, 5245 cases). Detailed phenotypic definitions are provided in Table 1.

### Genotype quality control and imputation

Approaches to quality control and imputation differed between cohorts, and are described in detail in the Supplementary Material.

### Association analyses

The statistical model used to conduct the GWAS for each cohort (described in Supplementary Table 4) depended on the design of the specific cohort. For unrelated case/control cohorts, we used PLINK2 to conduct logistic regression using an appropriate number of PCs to account for ancestry as necessary within each cohort (Supplementary Table 4). For related or imbalanced case/control cohorts we used SAIGE ^58^ or REGENIE ^59^. Note that for whole genome regression in REGENIE step 1, we used a set of pruned SNPs with MAF>0.01, excluding high LD regions and only including autosomal chromosomes (PLINK command: --indep-pairwise 1500 150 0.2). Further details on cohort-specific aspects of association analysis are provided in the Supplementary Methods. All analyses were two-tailed.

### Post-GWAS processing and quality control

We aligned the summary statistics of each GWAS to the TOPMed reference panel in Genome Reference Consortium Build 37 (GRCh37/hg19), using variant positions from ENSEMBL (see URLs). For cohorts that were in GRCh38, we first linked the datasets with the GRCh38 TOPMed reference panel (see URLs) by chromosome/base pair, then selected the SNP rsID labels and linked these labels to the GRCh37 TOPMed reference panel, and finally extracted the GRCh37 chromosome/base pair information for each SNP. Variants without a rsID label in the GRCh38 TOPMed reference panel were lifted over to GRCh37 using the liftOver tool (see URLs). After alignment, we applied an INFO and MAF filter to include SNPs with an INFO score of>0.3 and MAF>0.01 in cases and controls. We then used DENTIST ^60^ to remove variants with effects inconsistent with their linkage disequilibrium pattern with other assessed variants, estimating linkage disequilibrium from European ancestry individuals in Phase 3 of the 1000 Genomes project.

### Meta-analysis and quality control

We used the post-imputation module of Ricopili ^61^ to perform meta-analyses in METAL ^62^ using an inverse-variance weighted fixed-effect model, including assessment of variant heterogeneity as I^2^ values (Comparison of variant heterogeneity between our AN GWAS and that of Watson et al is included in the Supplementary Methods ^6^.) We defined independent significant SNPs with a genome-wide significant p-value (*P*<5x10^-8^) that were independent (r^2^<0.6) from each other. We then defined significant genomic loci by merging LD blocks of these independent significant SNPs if they were close to each other (<250 kb). Furthermore, we defined independent lead SNPs if independent significant SNPs were independent of each other at r^2^<0.1.

We ran a stepwise conditional analysis on our GWAS results to select independently-associated SNPs (conditional on lead SNPs) at each loci using GCTA-COJO ^63^. For these analyses we used one of our largest cohorts (*usa2*) as our reference for linkage disequilibrium.

### Female-only analyses and female-to-male polygenic risk scoring

We conducted a supplementary female-only GWAS for BE-BROAD and AN and generated female-only PRS using PRS-CS which we applied on male-only datasets with sufficient data (i.e., *n* case and *n* control >100) available. For the BE-BROAD female-only meta-analysis, all cohorts except *usa1* and *biov* were included, and *alsp, moba*, and *ukd2* were included as male-only target cohorts. For the AN female-only meta-analysis, all cohorts except *itgr, spa1, ukd1*, and *net2* were included, and *ipsy, fngn*, and *ukb2* were included as male-only target cohorts.

### SNP-based heritability and distinguishing polygenicity from other sources of inflation

We used linkage disequilibrium score regression (LDSC ^64^) to estimate SNP-based heritability (*h*^*2*^_SNP_). These estimates were transformed to the liability scale, assuming population prevalences as follows: BE-BROAD 4.5% ^15^, BE-NARROW 3.5% ^15^, AN 1.5% ^17^, AN-R 0.8% ^17,65^, AN-BP 0.7% ^17,65^. For all analyses using LDSC, we applied an LD reference panel based on the European subset of the 1000 Genomes Project (1kGP), restricted to SNPs present in the HapMap3 panel ^66^. For *N*, we calculated the sum of effective *N* across all cohorts and specified 0.5 for sample prevalence ^67^. All LDSC analyses, including tests of the of *h*^*2*^_SNP_ and SNP-*r*_*g*_ from 0 and from 1, were two-tailed tests of deviation from a chi-square distribution with jack-knifed standard errors.

Test statistics from GWAS of a polygenic trait are expected to be inflated, but inflation may also be due to spurious SNP associations caused by population stratification and cryptic relatedness of study participants. We used statistics from LDSC ^64^ to determine the source of inflation. Although the LDSC intercept is commonly used to distinguish polygenicity from spuriously inflated statistics, we calculated the attenuation ratio statistic, defined as (LDSC intercept–1)/(mean of association chi-square statistics–1), which may be a less biassed metric compared to the LDSC intercept ^16^. We included variants with MAF≥0.01 and INFO≥0.6.

### Comparison of the two binge-eating behaviour phenotypes

The two BE phenotype definitions balanced phenotypic certainty with sample size— BE-BROAD had a larger sample size and was potentially a better-powered GWAS than BE-NARROW but was likely to be more heterogeneous and could lack specificity to BE. To determine which phenotype to carry forward to follow-up analyses, we considered the LDSC intercept and attenuation ratio statistics and calculated the genetic correlation (SNP-*r*_*g*_) between the two BE phenotypes. Inflation was a more sizable component of the signal in BE-NARROW (attenuation ratio 0.21 ± 0.06) than in BE-BROAD (0.14 ± 0.04). Furthermore, the SNP-*r*_*g*_ between BE-NARROW and BE-BROAD did not differ from unity (1.00 ± 0.03).

We therefore concluded that BE-BROAD appropriately captured the common genetic component of BE with greater statistical power than BE-NARROW.

### Genetic relationship between traits

We used LDSC to calculate SNP-*r*_*g*_ with several aims. First, we estimated SNP-*r*_*g*_s between both BE phenotypes, AN, and AN subtypes to assess the genetic relationships among these eating phenotypes. Second, we calculated SNP-*r*_*g*_ between BE-BROAD/AN and 225 traits covering eight categories: 1) psychiatric trait or disorder, 2) substance use, 3) psychological/personality/behavioural, 4) anthropometric, 5) metabolism, 6) blood, 7) sociodemographic, and 8) somatic trait or disease. We selected these traits from an internal catalogue based on their power (*h*^*2*^_SNP_ Z-score>4) ^68^. To assess statistical significance, we tested whether SNP-*r*_*g*_ differed from zero and applied a Bonferroni-corrected p-value threshold of 2.20x10^-4^ based on 225 traits. If, according to this threshold, BE-BROAD and/or AN was significantly genetically correlated with another trait, we used the LDSC block-jackknife procedure (described in more detail in Appendix S1 of Hübel et al ^69^) to statistically compare the SNP-*r*_*g*_ between BE-BROAD and AN (Bonferroni-corrected p-value threshold of 23 2.20x10^-4^).

We used MiXeR to conduct univariate and bivariate casual mixture modelling of BE-BROAD and AN ^19^. MiXeR is an extension of LDSC that uses GWAS summary statistics to model the number of causal variants underlying a trait (polygenicity), as well as inferring the average contribution of each causal variant to heritability (discoverability). Bivariate MiXeR extends cross-trait LDSC to refine the interpretation of genetic correlation. It enables the shared number of causal variants between traits to be estimated, as well as the extent to which shared causal variants have the same direction of effect. We conducted MiXeR analyses using BE-BROAD and AN summary statistics, limited to autosomal variants with INFO≥0.6 and MAF≥0.01, and with the MHC region (chr6, 26-34Mb) excluded. We followed protocols and used LD reference files provided by the authors of MiXeR (see URLs).

To further investigate the genetic difference between BE-BROAD and AN we used CC-GWAS ^18^. CC-GWAS uses GWAS summary statistics to test for allele frequency differences between cases of two phenotypes, as opposed to a traditional GWAS that tests for differences between cases and controls. It generates Z-scores and p-values for allele frequency differences between cases of two phenotypes as a two-tailed test, using both ordinary least squares (OLS) and exact weights. We used BE-BROAD and AN summary statistics including non-ambiguous SNPs from HapMap 3 with INFO≥0.6 and MAF≥0.01. We used the liability-scale *h*^*2*^_SNP_, the SNP-*r*_*g*_ and associated covariate intercept between both traits as input. We furthermore set the BE-BROAD population prevalence to 4.5% (range 0.1 - 10%) and AN to 1.5% (range 0.1 - 4.3%) and approximated the number of effective loci to be 10,000–consistent with psychiatric disorder polygenicity ^18^. We additionally defined the number of independent CC-GWAS loci with PLINK 1.9 (--clump-p1 5e-8 --clump-p2 5e-8 -- clump-r2 0.1 --clump-kb 3000) and defined a genome-wide significant SNP if *P*<5x10^-8^ in the CC-GWAS OLS test and *P*<10^-4^ in the CC-GWAS exact test. Further, CC-GWAS calculates genetic distances between cases and controls of the two traits using 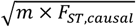 based on liability-scale *h*^*2*^_SNP_, SNP-*r*_*g*_, population prevalence and the number of independent causal variants of the two traits.

### Influence of BMI

We used GWAS-by-subtraction ^20^, an application of genomicSEM ^70^, in R v4.3.1 ^71^ to estimate the proportion of variance in BE-BROAD and AN independent of BMI (Supplementary Methods). Shared genomic covariance across traits is expected due to pleiotropy. Latent Genomic SEM factors explicitly model this covariance, making results less influenced by spurious biases than would conditioning on phenotypic traits in a GWAS ^72^. We used GWAS summary statistics from BE-BROAD, AN, and BMI ^21^.

We specified a structural equation model (SEM) that regressed both sets of summary statistics on a shared variable (“*BMI*”) and a non-BMI variable (“*non-BMI*”) for each eating phenotypes, respectively (Supplementary Figure 26). Specifically, we specified the two latent variables as a function of BMI and (e.g.) BE-BROAD: “*BMI* =∼ *NA* * BEBROAD + start(0.4) * BMI” and *“non-BMI* =∼ *NA* * BE-BROAD”. In line with the GWAS-by-subtraction specification as it has previously been applied ^20^, we additionally set the variance of the latent variables to 1 and a covariance of 0, and constrained the model so all (co)variance in BMI and BE-BROAD was captured by *BMI* and *non-BMI*. We used the diagonally-weighted least squares estimator, which is the default setting in Genomic SEM ^70^. Additional computational settings are shown in Supplementary Table 30. We then regressed the two latent factors on individual SNPs yielding a GWAS of the latent variables *BMI* and *non-BMI*. We subsequently used LDSC ^64^ to calculate SNP-*r*_*g*_ of the *non-BMI* factor with all traits identified in initial SNP-*r*_g_s. As a sensitivity analysis, we restricted our BE-BROAD sample in our GWAS to cohorts that were not ascertained for AN, reasoning that this might better capture BE behaviour outside of AN. We applied the same GWAS-by-subtraction model on that selection of cohorts (listed in Supplementary Table 11).

We also conducted exploratory two-sample Mendelian randomisation analyses of BE-BROAD with BMI and AN with BMI, testing causal effects in both directions with two-tailed tests. We used SNPs in linkage equilibrium as genetic instruments, with *P*<5x10^-6^ for BE-BROAD and AN, and *P*<5x10^-9^ for BMI. The instrument used for BMI was that suggested by the authors of the BMI GWAS ^21^. We repeated analyses using the *non-BMI* factor GWAS of BE-BROAD and of AN. We view these Mendelian randomisation analyses as exploratory because SNPs not passing genome-wide significance were included in the instruments for the eating phenotypes. To ensure our analyses were robust to potential violations of the assumptions of Mendelian randomisation, we conducted analyses using multiple methods in R 4.3.2, including the packages *TwoSampleMR, MendelianRandomisation*, and *MR-PRESSO* (Supplementary Methods) ^71,73,74^. These were inverse-variance weighted analysis, MR-Egger, mode-based estimation and median-based estimation. We determined the strength of association of our genetic instruments using the F-statistics ^75^. We used Cochran’s Q-statistic to test for instrument heterogeneity, with *P*<0.05 indicating heterogeneity ^76^. To investigate potential confounding via horizontal pleiotropy, we assessed the deviation of the MR-Egger intercept from 0, and performed a global bias test in MR-PRESSO. We excluded from the analysis SNPs identified by MR PRESSO as pleiotropic. We ran further methods robust to heterogeneity, including the penalised weighted median estimator, the contamination mixture method, and MR-Lasso ^77^. We assessed results visually (Supplementary Methods).

### Identification of gene-tissue associations with eating phenotypes

We used S-PrediXcan ^22^ to identify genetically regulated gene expression associated with our phenotypes. We tested the association of gene expression with our eating phenotypes using available GTEx v8 MASHR ^22,78,79^ and CommonMind DLPFC ^80,81^ tissue models. MASHR-based PredictDB models use fine-mapping methods for selection of eQTLs included in the predictor models, improving prediction ^22,78,79^. We included 45 GTEx v8 MASHR models, removing non-natural tissues (cell lines), tissues with N<100 individuals (kidney cortex), and testis ^82^. We performed liftover of our GWAS summary statistics to hg38, harmonisation, and imputation based on recommended preprocessing by Barbeira et. al ^78^ using *GWAS tools* (see URLs). S-PrediXcan uses GWAS summary statistics to impute the difference in genetically regulated gene expression between levels of the GWAS phenotype (in this instance, cases and controls). We tested whether these imputed differences differed from 0 as a two-tailed Z test. We used two different Bonferroni significance thresholds: an experiment-wide threshold, where we corrected for 600,382– 602,744 tests performed across all tissues (*P<*0.05/Tests_Total_ = *P<*8.32x10^-8^), and a tissue-specific threshold, where we corrected for varying numbers of tests performed within each tissue (*P<*0.05/Tests_Tissue X_, Supplementary Table 16). We performed two-tailed exact binomial tests for tissue enrichment using *binom*.*test()* in R for associations at three different significance thresholds: experiment-wide significant (*P<*0.05/Tests_Total_), tissue-specific significant (*P<*0.05/Tests_TissueX_), and nominally significant (*P<*0.05). We reported results with a focus on central nervous system tissues (brain and cervical spinal cord) and gastrointestinal tissues (oesophagus, colon, stomach, and small intestine).

S-MultiXcan is a summary-level method for measuring the joint association of genetically regulated gene expression across tissues with a phenotype of interest, leveraging shared eQTLs across tissues ^23^. Using our GTEx v8 MASHR S-PrediXcan results as input, along with MASHR models, and harmonised, imputed GWAS summary statistics, we ran S-MultiXcan on each of our eating phenotypes for all genes (N=22,241). S-MultiXcan gives as output the p-value for association of multi-tissue gene expression with the trait of interest (S-MultiXcan P), along with best single-tissue p-value, both from two-tailed tests. In order to account for potential false positive associations, we removed any significant S-MultiXcan associations where the single best tissue p-value was greater than 1x10^-4 23^. We set a Bonferroni significance threshold for our results, correcting for the number of genes tested in our S-MultiXcan analysis (*P*<0.05/22,241 = 2.25x10^-6^).

### Gene-wise and gene set analysis, including drug target and drug class analyses

Following a previously published approach ^83^, we used MAGMA v1.10 ^24^ to test the association between each phenotype and 1) the aggregate effect of SNPs mapped to protein-coding genes (gene analysis); 2) groups of genes with shared functional, biological, or other characteristics (gene-set analysis); 3) a gene-set analysis restricted to genes targeted by drugs (drug-set analysis) and 4) the enrichment of signal within classes of drug. We restricted SNPs to those with MAF≥0.01, INFO≥0.6, and which are present in 80% of the total sample and 50% of the cohorts. We mapped SNPs to protein-coding genes, applying a 35 kb upstream and 10 kb downstream window around hg19 gene positions from Ensembl release 75 ^84^. We obtained gene-wise p-values with the multi snp-wise model, which combines the lowest and mean p-values of all SNPs mapped to the gene. We tested 19,332-19,418 ENSEMBL genes across the five phenotypes and applied a Bonferroni correction of *P<*2.60x10^-6^. We used the 1kGP reference panel for estimating between-SNP LD.

For gene-set analyses, we applied a competitive analysis, which regresses the phenotype on the mean effect of genes within the gene set, with the mean effect of genes outside the gene set as a covariate, in a one-tailed test that the mean effect of genes within the gene set is greater than 0. We defined biological pathways based on gene ontology and canonical pathways from MSigDB v6.1 and psychiatric pathways identified from the literature. We tested 7324-7325 pathways across the five phenotypes and applied a Bonferroni correction of *P<*6.83x10^-6^.

For drug-set analyses, we defined drug sets based on drug targets from the Drug-Gene Interaction database DGIdb v4.2.0 ^85^; the Psychoactive Drug Screening Database Ki DB ^86^; CheMBL v27 ^87^; the Target Central Resource Database v6.7.0 ^88^; and DSigDB v1.0 ^89^, all downloaded in October 2020. We applied a competitive analysis and subsequently grouped the results based on the Anatomical Therapeutic Chemical class of the respective drugs ^90^. For drug-class analysis, we first ranked all drug-gene sets according to their association in the drug-set analysis. We then generated enrichment curves for specific drug classes, assigning a ‘hit’ if the drug-gene set belonged to the class or a “miss” if it was outside the class. We calculated the area under the curve and determined statistical significance with the Wilcoxon Mann-Whitney test, comparing drug-gene sets within the class to those outside the class as a one-tailed test of whether in-class drugs showed greater association with the disorder than out-of-class drugs. We applied a Bonferroni correction of *P<*3.23x10^-5^ (based on 1546-1547 drug sets) for the drug-set analysis and *P<*3.08x10^-4^ (based on 162 drug classes) for the drug-class analysis to account for multiple testing.

### Tissue and cell-type specific analyses

To identify relevant tissues and cell-types related to the common genetic risk of BE-BROAD and AN, we performed tissue and cell-type heritability (*h*^*2*^_SNP_) enrichment analyses. First, we analysed the enrichment of *h*^*2*^_SNP_ in 27 tissues from the GTEx gene expression data (v8) after excluding tissues with less than 100 donors, non-natural tissues (such as cell lines), and testis tissues (since it was an expression outlier) ^91^. Second, we analysed enrichment of *h*^*2*^_SNP_ in 31 superclusters and 461 cell clusters based on the single-nucleus RNA sequencing data including over three million nuclei from around 100 dissections across the adult human brain ^29^. Within each expression dataset, we calculated the specificity of gene expression per tissue or cell type (superclusters and clusters separately), defined as the expression of each gene (counts per million, CPM) in a tissue or cell type (i.e., the superclusters and clusters respectively) divided by the total expression of this gene across all tissues or cell types in the dataset ^92^. We then used the genes with the top 10% specificity in each tissue or cell type to perform the heritability enrichment analysis using stratified LDSC ^92,93^. Specifically, we compared the per-SNP heritability of SNPs within 100kb flankings of the top 10% specific genes and the per-SNP heritability of other SNPs, using the baseline model that adjusted for 53 baseline annotations ^93^. We then used the coefficient *z*-scores to calculate the one-sided p-values. Finally, we accounted for multiple comparisons by calculating FDR per trait for the GTEx dataset (27 tests), the human brain superclusters (31 tests) and clusters (461 tests) respectively.

### Polygenic prediction

We used PRS-CS ^94^ to generate polygenic risk scores (PRS) for BE-BROAD and AN. First, we performed leave-one-cohort-out (LOO) analyses to generate LOO GWAS summary statistics from all cohorts except the target cohort and used this as the base data to calculate individual-level PRS in each target cohort. We included non-ambiguous SNPs with INFO≥0.6 and MAF≥0.01 in the PRS calculation. We used the 1000 Genomes Project Phase 3 EUR reference as the LD reference panel and provided median sample size per LOO meta-analysis as input for PRS-CS. Posterior SNP effect size estimates from PRS-CS were then combined across chromosomes to calculate individual PRS via PLINK (--score 2 4 6 sum) ^95^. We standardised the individual PRS by applying the scale function in R (version 4.3.2) ^71^. Using the standardised PRS scores, we first assessed the proportion of variance explained by PRS for each phenotype through calculating the nested Nagelkerke’s pseudo-*R*^*2*^ (that is,the pseudo-R^2^ of the full model minus that of the model excluding the PRS) ^96^. The inclusion of target cohorts for each phenotype was based on the effective sample size (N_eff_ half>1000), study characteristics, and availability of individual-level data (more detailed information in the Supplementary Methods). Two-tailed logistic regression was performed for BE-BROAD and AN PRS on BE-BROAD and AN, adjusting for cohort-specific PCs. We then converted the variance explained by each PRS (*R*^*2*^) to the liability scale using populationprevalences of BE-BROAD and AN as used for LDSC ^97^. In addition, we divided individuals into PRS decile groups and assessed their relative risk of BE-BROAD and AN compared to the group of individuals in the lowest PRS decile. We also examined predictive performance of each PRS regarding their sensitivity, specificity, and precision to predict BE-BROAD and AN status using the *pROC* ^98^ and *pracma* ^99^ packages in R by comparing the area under the curve (AUC) of the receiver operating characteristic (ROC) curve and the precision recall (PR) curve of the full model including PRS and PCs as predictors against the null model including PCs only as predictors.

We used female-only GWAS summary statistics as base data in PRS-CS ^94^ to assess whether female BE-BROAD PRS could be applied to male individuals to assess their BE-BROAD risk in three cohorts (*alsp, moba, ukd2*, N_case_male_total_=1055, N_control_male_total_=38,046). The same analysis was performed to examine the association of female AN PRS with male AN risk in three cohorts (*ukb2, fngn, ipsy*, N_case_male_total_=1524, N_control_male_total_=388,891).

We assessed whether the levels of BE-BROAD and AN PRS differed across subgroups, including a) those with BE-BROAD only, b) those with both BE-BROAD and AN, and c) those with AN only. Target cohorts for this analysis included cohorts with sufficient data available on some/all subgroups (Supplementary Methods, Supplementary Table 1). Specifically, we selected *aunz* and *sedk* to assess individuals with both BE-BROAD and AN and to assess the AN only group. We selected *ukb2* and *ukd2* for assessing BE-BROAD and AN PRS levels in all subgroups. We assessed differences between subgroups and controls by performing linear regression on each PRS for each subgroup compared to controls, adjusting for cohort-specific genetic PCs. In addition to setting controls as the reference group, we also set different subgroups as the reference group to compare differences in BE-BROAD PRS and AN PRS across different subgroups.

Last, we generated AN PRS from the main meta-analysis – including solely individuals of European genetic ancestry – and applied this to two East Asian genetic ancestry cohorts (N_case_=77, N_control_=117 in a Japanese cohort, N_case_=75, N_control_=109 in a Korean cohort). Korean data were merged with Japanese data from GCAN cohorts. The same pre-imputation quality control and imputation method used for the European ancestry cohorts was conducted. We used the 1000 Genomes Project Phase 3 EUR reference as the LD reference panel for PRS, as the reference panel should align with the ancestry in the base GWAS ^94^. We used the European AN prevalence estimate for liability scale conversion, as estimates of AN prevalence in East Asia are sparse and what estimates exist are approximately consistent with estimates in countries with primarily European genetic ancestries ^100^.

### Sensitivity analyses using down-sampled BE-BROAD data

A considerable portion (13%) of the cases for the BE-BROAD GWAS includes individuals who were recruited through studies that focused on AN ascertainment. Even though the BE-BROAD phenotype is expected to be heterogeneous given its transdiagnostic nature, we sought to understand the influence of AN on this BE-BROAD phenotype. We therefore conducted an additional BE-BROAD meta-analysis where we excluded individuals with BE-BROAD who were identified in cohorts that were specifically ascertained for AN (e.g., the ANGI cohorts). This resulted in an additional analysis including 87% of the BE-BROAD GWAS, consisting of the following cohorts: *sebe, agds, alsp, biov, esbb, fngn, jans, moba, ukb2, ukd2*. We then calculated SNP-*r*_*g*_ to compare the genetic relationship of the “not-ascertained-for-AN” BE-BROAD GWAS and the original BE-BROAD GWAS with selected traits significantly correlated with the original BE-BROAD GWAS or with the AN GWAS (hypothesising that AN-related effects may mask or drive SNP-*r*_*g*_s between such traits and the original BE-BROAD GWAS).

### URLS

TopMED Hg37 VCF: https://ftp.ensembl.org/pub/grch37/release-113/data_files/homo_sapiens/GRCh37/variation_genotype/TOPMED_GRCh37.vcf.gz

TopMED Hg38 VCF: https://ftp.ensembl.org/pub/grch37/release-113/data_files/homo_sapiens/GRCh38/variation_genotype/TOPMED_GRCh38.vcf.gz

LiftOver: https://ftp.ensembl.org/pub/grch37/release-113/data_files/homo_sapiens/GRCh38/variation_genotype/TOPMED_GRCh38.vcf.gz

MiXeR: https://github.com/precimed/mixer

SPrediXcan best practice: https://github.com/hakyimlab/MetaXcan/wiki/Best-practices-for-integrating-GWAS-and-GTEX-v8-transcriptome-prediction-models

*GWAS tools*: https://github.com/hakyimlab/summary-gwas-imputation/wiki/GWAS-Harmonization-And-Imputation

## Notes

### Funding Statement

This overall study was funded by NIH grant R01MH124851.
Katherine Schaumberg is supported by NIH K01MH123914. Karanvir Singh is supported by the Canadian Institutes of Health Research. Jiayi Xu is supported by NIMH R01MH136149, R01MH124839, R01ES033630. Jerry Guintivano reports funding from NIMH K01 MH116413.
GCAN was funded by the Wellcome Trust Case Control Consortium 3 (WTCCC3) WT088827/Z/09 (Collier/Bulik/Sullivan) A Genome-wide Association Study of Anorexia Nervosa. Spa1 was partially funded by the European Union's Horizon Europe innovation programme, under grant agreement No. 101080219 (eprObes, Early Prevention of Obesity). Spa1 was also funded by Instituto de Salud Carlos III (ISCIII) (FORT23/00032), co-funded by FEDER funds/European Regional Development Fund (ERDF), a way to build Europe. CIBERobn is an initiative of ISCIII. Additional funding for spa1 was received by AGAUR-Generalitat de Catalunya (2021-SGR-00824). Gns2 was supported by the German Research Foundation collaborative research center grant (DFG, SFB 940/3) bzw. neuerdings "SFB940 TP C03" (laut Mail von Solveig Otto an Julius vom 06.01.2022). DFG research grant: "Hormonal modulation of neural networks in anorexia nervosa" EH 367/5-1 (PI S. Ehrlich). DFG research grant: EH 367/7-1 "Dynamische Veränderungen des strukturellen und funktionellen Hirn-Konnektoms bei Patientinnen mit Anorexia Nervosa" (PI S. Ehrlich), the Schweizer Anorexia Nervosa Stiftung and the B. Braun-Stiftung; Marga und Walter Boll-Stiftung. Anke Hinney reports support from Deutsche Forschungsgemeinschaft (DFG, HI 865/2-1), BMBF (01GS0820). Lars Alfredsson reports funding from the Swedish Research Council, Swedish Brain Foundation, Swedish Council for Health, Working Life and Welfare. Stéphanie Le Hellard reports support from the Trond Mohn Fondation. Fre1 was supported by EC framework V 'Factors in healthy eating'. James Kennedy reports support from the Larry and Judith Tanenbaum Family Foundation. Stephen Scherer is the Northbridge Chair in Paediatric Research, and reports support from the SickKids Foundation. Japanese samples were funded by Grants-in-Aid for Scientific Research (20390201 and 23390201) awarded to G. Komaki by the Ministry of Education, Culture, Sports, Science, and Technology of Japan.
CHOP was funded by the Price Foundation. Andrew Bergen reports funding from Price Foundation Collaborative Group and NIH R01DA044014 and R01DA050495.
The Anorexia Nervosa Genetics Initiative was an initiative of the Klarman Family Foundation. The QSkin Study is supported by a Clinical Trials and Cohort Grant [APP1185416] from the National Health and Medical Research Council of Australia (NHMRC) and was previously supported by grants APP1073898, APP552429, APP1063061. Jenny Jordan reports support from the Klarman Foundation, the University of Otago Research Fund, and the NIH. Tracey Wade reports support from the National Health and Medical Research Council Project Grant 480420.
ANGI-DK was part of the Anorexia Nervosa Genetics Initiative, which was an initiative of the Klarman Family Foundation (Mortensen, local PI; Bulik, PI). The iPSYCH was supported by grants from the Lundbeck Foundation (R102-A9118; R155-2014-1724; R248-2017-2003) and the Universities and University Hospitals of Aarhus and Copenhagen. Zeynep Yilmaz is supported by Independent Research Fund Denmark (DFF; Sapere Aude no. 1052-00029B); DFF (grant no. 3166-00063B, 4309-00050B); Lundbeck Foundation Ascending Investigator (R434-2023-269). Liselotte Petersen and Janne Larsen report funding from the Lundbeck Foundation (grant no. R276-2018-4581, recipient Petersen) and NIMH (R01MH120170).
ANGI-Sweden was part of the Anorexia Nervosa Genetics Initiative, which was an initiative of the Klarman Family Foundation (Landén local PI; Bulik, PI). Cynthia Bulik is supported by the U.S. National Institute of Mental Health (R01MH136149;R01MH134039,R56MH129437; R01MH120170; R01MH124871; R01MH119084; R01MH118278; R01MH124871) and the Swedish Research Council (Vetenskapsrådet, award: 538-2013-8864). Patrick Sullivan reports support from the Swedish Research Council (Vetenskapsrådet, award D0886501), and NIMH R01 MH124871. Ruyue Zhang is supported by the Swedish Research Council (Vetenskapsrådet) grant no. 2022-00242. Andreas Birgegård reports funding from the Swedish Research Council (Vetenskapsrådet) grant no. 538-2013-8864, PI Bulik.
Genotyping of Danish clinical cases was supported by the Anorexia Nervosa Genetics Initiative, which was an initiative of the Klarman Family Foundation (Werge, local PI; Bulik, PI).
Funding for BEGIN-Sweden was received from the Swedish Research Council Swedish Research Council (Vetenskapsrådet, award: 538-2013-8864) (Bulik, PI).
The AGDS was primarily funded by National Health and Medical Research Council (NHMRC) of Australia grant 1086683. Sarah Medland is supported by NHMRC Leadership Grant APP1172917.
This study represents independent research partly funded by the UK National Institute for Health Research (NIHR) Biomedical Research Centre at South London and Maudsley NHS Foundation Trust and King's College London. High-performance computing facilities were funded with capital equipment grants from the GSTT Charity (TR130505) and Maudsley Charity (980). This work was supported by the UK Medical Research Council and the Medical Research Foundation (MR/R004803/1). The UK Medical Research Council and Wellcome (102215/2/13/2 and 217065/Z/19/Z) and the University of Bristol provide core support for ALSPAC. A comprehensive list of grants is available on the ALSPAC website. This research was specifically funded by the NIHR (CS/01/2008/014) and the NIH (MH087786-01). GWAS data were generated by sample logistics and genotyping facilities at Wellcome Sanger Institute and LabCorp (Laboratory Corporation of America) using support from 23andMe. Mohamed Abdulkadir acknowledges grant support from the National Institute of Mental Health (R01MH120170).
Research using BioVU was funded by R01MH118233 to Lea K Davis.
This research in the Estonian Biobank was supported by the European Union through the European Regional Development Fund (Project No. 2014-2020.4.01.15-0012), and the Estonian Research Council's grant No. PSG615. Krista Fischer reports support from grant PRG1197 by the Estonian Research Council. The funding agencies had no role in the design and conduct of the study; collection, analysis, and interpretation of data; or the writing of the manuscript or the decision to submit it for publication.
The FinnGen project is funded by two grants from Business Finland (HUS 4685/31/2016 and UH 4386/31/2016) and nine industry partners (AbbVie, AstraZeneca, Biogen, Celgene, Genentech, GSK, MSD, Pfizer, and Sanofi). Samuli Ripatti and Jaakko Kaprio report funding from the Academy of Finland Center of Excellence in Complex Disease Genetics (grant # 352792).
The original studies included in gcfm were funded by three different grants from the Canadian Institutes of Health Research (MOP-79490, MOP-57929, MOP-142717) or by industry support from Cogir Immobilier ("COGIR"). Funders had no role in study design, data collection, analysis and interpretation, report writing, or in decisions surrounding submission of papers for publication.
The Janssen study was funded by Johnson & Johnson Pharmaceutical Research & Development, L.L.C.
MoBA thanks the Norwegian Institute of Public Health (NIPH) for generating high-quality genomic data. This research is part of the HARVEST collaboration, supported by the Research Council of Norway (#229624) and the South-Eastern Norway Regional Health Authority (#2023031). We also thank the NORMENT Centre, in collaboration with deCODE Genetics, for providing genotype data, funded by the Research Council of Norway (#223273), South-Eastern Norway Health Authorities and Stiftelsen Kristian Gerhard Jebsen. We also acknowledge funding from the U.S. National Institute of Child Health and Development (R01-HD047186, Bulik, PI). We further thank the Center for Diabetes Research, the University of Bergen for providing genotype data and performing quality control and imputation of the data funded by the ERC AdG project SELECTionPREDISPOSED, Stiftelsen Kristian Gerhard Jebsen, Trond Mohn Foundation, the Research Council of Norway, the Novo Nordisk Foundation, the University of Bergen, and the Western Norway Health Authorities. Ole Andreassen was funded by the Research Council of Norway (#324499, #324252), EU's H2020 RIA grant (#964874), NordForsk project (#164218) and KG Jebsen Stiftelsen, Alexandra Havdahl was funded by the South-Eastern Norway Health Authorities (#2019097, #2020022), Helga Ask was funded by the Research Council of Norway (#324620) and NordForsk (#156298). Elizabeth Corfield was supported by the Research Council of Norway (#274611) and South-Eastern Norway Health Authorities fellowship (#2021045).
This work was performed on the Tjeneste for Sensitive Data facilities, owned by the University of Oslo, operated and developed by the Tjeneste for Sensitive Data service group at the University of Oslo, IT Department (USIT). The computations were performed on resources provided by Sigma2 - the National Infrastructure for High-Performance Computing and Data Storage in Norway.
Research in URGE was supported by grants of Netherlands Organisation for Scientific Research (NWO: 024.004.012, ALWOP.137 and OCENW.KLEIN.071) to Roger Adan.
Research in GLAD and the NIHR Bioresource was supported by the National Institute of Health Research (NIHR) BioResource, NIHR Biomedical Research Centre [IS-BRC-1215-20018], HSC R&D Division, Public Health Agency [COM/5516/18], MRC Mental Health Data Pathfinder Award (MC_PC_17,217), and the National Centre for Mental Health funding through Health and Care Research Wales.
Charlotte's Helix was partly funded by the UK National Institute for Health Research (NIHR) Biomedical Research Centre at South London and Maudsley NHS Foundation Trust and King's College London. Ulrike Schmidt receives salary support from the NIHR BRC, and from the Medical Research Council/Arts and Humanities Research Council/Economic and Social Research Council Adolescence, Mental Health and the Developing Mind initiative as part of the EDIFY programme (grant number MR/W002418/1).
Research in the Korean Study of Anorexia Nervosa was supported by the Korea Centers for Disease Control and Prevention Research Fund [grant number: 2016-ER6310-00, 2016].
No authors nor their institutions at any time received payment or services from a third party for any aspect of the submitted work (including but not limited to grants, data monitoring board, study design, manuscript preparation, statistical analysis, etc.)

### Author Declarations

Price Foundation Collaborative Group and Children's Hospital of Philadelphia (CHOP): Ethical approval was obtained by each of the individual study sites (Cornell University, University of California at Los Angeles, University of Pittsburgh, University of Toronto, University of London, and University of Munich.). For the CHOP controls, CHOP's Research Ethics Board approved the study, and informed consent was obtained from all participants or their parents. Anorexia Nervosa Genetics Initiative (ANGI): ANGI-Australia and New Zealand (ANGI-ANZ) and Queensland Skin Study (QSkin): For ANGI-ANZ, all participants provided informed consent. Ethical approval for the Australian arm of the study was obtained by the QIMR Berfhoger Human Research Ethics Committee; for New Zealand, this was provided by the Health and Disability Ethics Committee of the New Zealand Ministry of Health. For QSkin, ethical approval was obtained by the QIMR Berghofer Human Research Ethics Committee (QIMR-HEC approval P1309, P2034). ANGI-Denmark (ANGI-DK) and the The Lundbeck Foundation Initiative for Integrative Psychiatric Research (iPSYCH): Participants from ANGI-DK and the iPSYCH study did not provide written informed consent, as an exemption from consent was provided by the Danish Scientific Ethics Committee (Videnskabsetisk Komité). This exemption was most recently approved in 2018. Additional Danish cases: All participants provided written informed consent before participating in the study. The study was approved by the Capital Region of Denmark's Committees on Health Research Ethics (De Videnskabsetiske Komiteer for Region Hovedstaden) (approval number H-KF-01-024/01). ANGI-United States of America (ANGI-US): ANGI-US was approved by the Institutional Review Board of the University of North Carolina at Chapel Hill (IRB: 13-0081). All participants were broadly consented for research. ANGI-Sweden, LifeGENE and Binge-Eating Genetics Initiative (BEGIN): This project has been approved by the ethical review board in Sweden. The fundament of the ethical permit comes from ANGI Sweden (dnr 2013/112-31/2) which has later been amended to also include individuals with another diagnosis than AN (dnr 2014/1563) and to change the study name to BEGIN (dnr 2016/1852-32). For LifeGene, the Regional Ethical Review Board of Stockholm provided ethical approval, and all participants provided online consent. Australian Genetics of Depression Study: Ethics approval for all aspects of the project was obtained from the QIMR Berghofer Human Research Ethics Committee (P2118 & P3434). Avon Longitudinal Study of Parents and Children (ALSPAC): Ethical approval for the study was obtained from the ALSPAC Ethics and Law Committee and the Local Research Ethics Committees. Informed consent for the use of data collected via questionnaires and clinics was obtained from participants following the recommendations of the ALSPAC Ethics and Law Committee at the time. The main caregiver initially provided consent for child participation, and from the age of 16 years, the offspring themselves have provided informed written consent. The Vanderbilt University biorepository (BioVU): The study was approved by the Vanderbilt University Medical Center Institutional Review Board (IRB #201609). The Estonian Biobank: The research project has obtained approval from the Estonian Council on Bioethics and Human Research (1.1-12/624). The Finnish Study of Genetics (FinnGEN): Written informed consent was obtained from all the study participants. For the Finnish Institute of Health and Welfare (THL)-driven FinnGen preparatory project and FinnGen project, all patients and control subjects had provided informed consent for biobank research, based on the Finnish Biobank Act. Alternatively, the North Karelia Project (FINRISK) and Health 2000 cohorts were based on study specific consents and later transferred to the THL Biobank after approval by Valvira, the National Supervisory Authority for Welfare and Health. Recruitment protocols followed the biobank protocols approved by Valvira. Data from the Eating Disorders Continuum (EDC) at the Douglas Mental Health University Institute in Montreal: In all cases, there was consent to participation in one or more of the original studies, as well as to sharing of DNA with other research groups for additional genetic studies. All aspects were approved by the Research Ethics Boards of the Douglas Mental Health University Institute or Montreal West Island Integrated University Health and Social Services Centre. Jannsen: The CAPSS220BED clinical study (ClinicalTrials.gov ID: NCT00210808) was conducted by Johnson & Johnson Pharmaceutical Research & Development, L.L.C., and was approved by the appropriate ethical review boards and followed the principles outlined in the Declaration of Helsinki for all human investigations. In addition, informed consent has been obtained from the study participants. The Norwegian Mother, Father and Child Cohort Study (MoBa): The establishment of MoBa and initial data collection was based on a licence from the Norwegian Data Protection Agency and approval from The Regional Committees for Medical and Health Research Ethics. The MoBa cohort is currently regulated by the Norwegian Health Registry Act. The current study was approved by The Regional Committees for Medical and Health Research Ethics (20311). Utrecht Research Group Eating Disorders (URGE) cohort: Informed consent was obtained from all patients who provided DNA. The Biobank Ethics Committee of the University of Utrecht gave ethical approval for this work. UK Biobank: UK Biobank is registered as a Research Tissue Bank (North West - Haydock Research Ethics Committee, 21/NW/0157). All data and sample applications may use this ethical clearance to conduct their research. Separate Research Ethics Committee or other ethical clearance is not required. This research was conducted under application 82087 (PI: Jonathan Coleman). Charlotte's Helix: Charlotte's Helix participants were recruited using ethics approval provided by the Research Ethics Committee South Central Oxford C (15/SC/0388). The Genetic Links to Anxiety and Depression (GLAD) Study, and the National Institute for Health Research (NIHR) Bioresource: The GLAD Study was approved by the London - Fulham Research Ethics Committee on 21st August 2018 (REC reference: 18/LO/1218) following a full review by the committee. The NIHR BioResource has been approved as a Research Tissue Bank by the East of England - Cambridge Central Committee (REC reference: 17/EE/0025). Korean Study of Anorexia Nervosa: The study was approved by the Institutional Review Board of Inje University (INJE 2016-01-003-002). Genetics Consortium for Anorexia Nervosa: Each study site (Medical University of Vienna, Toronto General Hospital, Charles University, University of Helsinki, Institut National de la Santé et de la Recherche Médicale, University of Pittsburgh, University of North Carolina at Chapel Hill, Roseneck Hospital for Behavioral Medicine, University of Munich, Kings College London, Laureate Psychiatric Hospital, Weill Cornell Medical College, Sheppard Pratt Health System, Neuropsychiatric Research Institute Fargo, University of California at Los Angeles, University of Pennsylvania, University of Birmingham, University of Duisburg-Essen, GlaxoSmithKline Leeds, Athens University, Medical School, Padua/Verona/Treviso/Vicenza/Portogruaro hospitals, University Medical Center Utrecht, Altrecht in Zeist, Poznan University of Medical Sciences, Center for Genomic Regulation Barcelona, Department of Psychiatry University Hospital of Bellvitge, Karolinska Institutet, McLean Hospital/Harvard Medical School, Vanderbilt University School of Medicine) obtained informed consent from all participants, as well as permission from local ethical committees.

### Summary of Updates

Additional analyses added: per-variant heterogeneity in AN; BE Broad and AN univariate and bivariate mixture modelling, with SBayesRC validation; additional power analysis for Watson et al AN GWAS. Minor edits to text. Additional authors and affiliations added.

