## Supplementary Note for "Genome-wide association studies of binge-eating behaviour and anorexia nervosa yield insights into the unique and shared biology of eating disorder phenotypes"

### Cohort descriptions

#### Summary

The current analysis includes data that have been previously published in combination with new datasets. Previously published datasets include:

- Case-only data from the Genetics Consortium for Anorexia Nervosa/Wellcome Trust Case Control Consortium-3 (GCAN/WTCCC-3) as described in Watson et al [^1^](https://sciwheel.com/work/citation?ids=7192844&pre=&suf=&sa=0). These data were first included in Boraska et al [^2^](https://sciwheel.com/work/citation?ids=3303352&pre=&suf=&sa=0), and thereafter in Duncan et al [^3^](https://sciwheel.com/work/citation?ids=3611133&pre=&suf=&sa=0). Furthermore, we included the control data for these datasets as described in Watson et al [^1^](https://sciwheel.com/work/citation?ids=7192844&pre=&suf=&sa=0). We refer to these (10) datasets as follows: *fin1, fre1, gns2, itgr, net1, poco, spa1, ukd1, usa1.* The dataset *gcfm* is part of GCAN/WTCCC-3 but has not been previously published.
- Case-control data from the Children’s Hospital of Philadelphia/Price Foundation Collaborative Group (CHOP/PFCG) (dataset *chop*), first published in Wang et al [^4^](https://sciwheel.com/work/citation?ids=1337768&pre=&suf=&sa=0), and thereafter included in Duncan et al [^3^](https://sciwheel.com/work/citation?ids=3611133&pre=&suf=&sa=0) and Watson et al [^1^](https://sciwheel.com/work/citation?ids=7192844&pre=&suf=&sa=0).
- Datasets including case-control data from the Anorexia Nervosa Genetics Initiative (ANGI) studies from different sites have partly been previously published in Watson et al [^1^](https://sciwheel.com/work/citation?ids=7192844&pre=&suf=&sa=0), and partly updated. Here, we included ANGI-Australia and New Zealand (dataset *aunz*), ANGI-Denmark (dataset *ipsy*), ANGI-Sweden (part of dataset *sedk*), and ANGI-United States (dataset *usa2*) as described in Watson et al [^1^](https://sciwheel.com/work/citation?ids=7192844&pre=&suf=&sa=0).
- Part of the UK Biobank dataset (*ukb2*) has been previously published in Watson et
  al [^1^](https://sciwheel.com/work/citation?ids=7192844&pre=&suf=&sa=0).

The remaining datasets (*agds*, *alsp, biov, esbb, fngn, gcfm, jans, moba, net2, sebe, ukch, ukd2*) include data that have not been previously published as part of a GWAS.

***Non-cohort funding statements:*** Katherine Schaumberg is supported by NIH K01MH123914. Karanvir Singh is supported by the Canadian Institutes of Health Research. Jiayi Xu is supported by NIMH R01MH136149, R01MH124839, R01ES033630. Jerry Guintivano reports funding from NIMH K01 MH116413.

#### GCAN/WTCCC-3

GCAN/WTCCC-3 is short for “Genetics Consortium for Anorexia Nervosa / Wellcome Trust Case Control Consortium-3” [^2^](https://sciwheel.com/work/citation?ids=3303352&pre=&suf=&sa=0). In 2007, the Genetic Consortium for Anorexia Nervosa (GCAN) was established to combine existing genetic samples from individuals with AN into a single resource. GCAN contains data from different countries, and study participants were recruited through clinical sites or existing studies.

In the current publication, the following datasets contain GCAN cases: *fin1, fre1, gcfm, gns2, itgr, net1, poco, spa1, ukd1, and usa1*. These case-only data were first included in Boraska et al [^2^](https://sciwheel.com/work/citation?ids=3303352&pre=&suf=&sa=0). As the GCAN consists of cases only, these were combined with a separate set of controls which were sourced as described in Duncan et al [^3^](https://sciwheel.com/work/citation?ids=3611133&pre=&suf=&sa=0) (cohorts *fin1, fre1, itgr, net1, spa1, ukd1, usa1*). The Swedish cases have been merged with German and Norwegian cases, forming the dataset *gns2*.

To summarise, a set of PGC controls was selected based on Illumina genotyping and geographical location. Then, we performed quality control to identify suitably matched controls of European genetic ancestry (following Supplementary Methods above). It is important to note that some of the controls were not screened for AN which, although ideal, was not feasible. This could potentially result in an attenuation of the GWAS signal because of misclassification; however, we do not anticipate false results because of the low prevalence of AN.

Some GCAN cohorts were not included in Duncan et al. [^3^](https://sciwheel.com/work/citation?ids=3611133&pre=&suf=&sa=0) (*gcfm*, *gns2*, *poco*) due to a small number of cases (Sweden, N <100) or a lack of controls (Poland) but were included in Watson et al [^1^](https://sciwheel.com/work/citation?ids=7192844&pre=&suf=&sa=0) and are also part of the current study. A detailed description is given in Watson et al [^1^](https://sciwheel.com/work/citation?ids=7192844&pre=&suf=&sa=0). Briefly, study participants consisted of females only and were diagnosed with AN via a semi-structured or structured interview based on the DSM or via DSM-based population assessments. Cases consisted of individuals with a lifetime DSM-IV diagnosis of AN (restricting or binge-purge subtype) or lifetime DSM-IV EDs ‘not otherwise specified’ AN subtype (i.e., all criteria of AN without amenorrhea OR all criteria for AN except that, despite significant weight loss, the weight is in the normal range). Individuals were excluded if medical or psychiatric conditions were present that could have confounded a diagnosis of AN (i.e., conditions causing weight loss, schizophrenia, or intellectual disability). Controls for the Polish sample were gathered from a hospital-based case-control sample and a population-based case-control sample in Łódź and Warsaw for a study of upper aerodigestive tract cancers [^5^](https://sciwheel.com/work/citation?ids=5610920&pre=&suf=&sa=0). Controls were unscreened for psychiatric phenotypes and were included in the dataset *poco*.

***Acknowledgements:*** Fernando Fernández-Aranda thanks CERCA for its institutional support.

***Ethics statement:*** Each study site obtained informed consent from all participants, as well as permission from local ethical committees.

***Funding statement:*** This study was funded by the Wellcome Trust Case Control Consortium 3 (WTCCC3) WT088827/Z/09 (Collier/Bulik/Sullivan) A Genome-wide Association Study of Anorexia Nervosa. *Spa1* was partially funded by the European Union’s Horizon Europe innovation programme, under grant agreement No. 101080219 (eprObes, Early Prevention of Obesity). *Spa1* was also funded by Instituto de Salud Carlos III (ISCIII) (FORT23/00032), co-funded by FEDER funds/European Regional Development Fund (ERDF), a way to build Europe. CIBERobn is an initiative of ISCIII. Additional funding for *spa1* was received by AGAUR-Generalitat de Catalunya (2021-SGR-00824). *Gns2* was supported by the German Research Foundation collaborative research center grant (DFG, SFB 940/3) bzw. neuerdings “SFB940 TP C03” (laut Mail von Solveig Otto an Julius vom 06.01.2022). DFG research grant: "Hormonal modulation of neural networks in anorexia nervosa" EH 367/5-1 (PI S. Ehrlich). DFG research grant: EH 367/7-1 "Dynamische Veränderungen des strukturellen und funktionellen Hirn-Konnektoms bei Patientinnen mit Anorexia Nervosa" (PI S. Ehrlich), the Schweizer Anorexia Nervosa Stiftung and the B. Braun-Stiftung; Marga und Walter Boll-Stiftung. Anke Hinney reports support from Deutsche Forschungsgemeinschaft (DFG, HI 865/2-1), BMBF (01GS0820). Lars Alfredsson reports funding from the Swedish Research Council, Swedish Brain Foundation, Swedish Council for Health, Working Life and Welfare. Stéphanie Le Hellard reports support from the Trond Mohn Fondation. *Fre1* was supported by EC framework V 'Factors in healthy eating'. James Kennedy reports support from the Larry and Judith Tanenbaum Family Foundation. Stephen Scherer is the Northbridge Chair in Paediatric Research, and reports support from the SickKids Foundation.

#### *chop*

This dataset combines two different cohorts: (1) Cases obtained from the Price Foundation Collaborative Group (PFCG) [^6,7^](https://sciwheel.com/work/citation?ids=932320,930334&pre=&pre=&suf=&suf=&sa=0,0), and (2) controls obtained from the Children’s Hospital of Philadelphia (CHOP) [^4^](https://sciwheel.com/work/citation?ids=1337768&pre=&suf=&sa=0). The PFCG was established in 2000 through collaboration between The Price Foundation, the University of Pittsburgh, and other universities in North America and Europe [^6^](https://sciwheel.com/work/citation?ids=932320&pre=&suf=&sa=0). Cases were recruited from different sites based on ED assessment experience, number of records of individuals treated for AN, and the geographical distribution (rationale described in more detail in Kaye et al [^6^](https://sciwheel.com/work/citation?ids=932320&pre=&suf=&sa=0)). Included study sites were Cornell University, University of California at Los Angeles, University of Pittsburgh, University of Toronto, University of London, and University of Munich.

Cases included individuals with a DSM-IV diagnosis of AN with or without amenorrhea via the Structured Interview of Anorexia nervosa and Bulimic Syndromes
(SIAB) [^8^](https://sciwheel.com/work/citation?ids=17424695&pre=&suf=&sa=0). Furthermore, cases had to be aged between 13 and 65 years and have fulfilled the AN criteria for at least three years prior to ascertainment. Individuals were excluded if the onset of AN was more than 25 years previously, or if they had a lifetime history of any of the following: organic brain syndrome; binge ED; “regular” binge eating (i.e., binge eating at least once weekly for three or more consecutive months); a medical illness that could affect appetite, eating behaviour, or body weight; IQ <70; dementia; schizophrenia; bipolar illness; obesity.

Unscreened controls were recruited by CHOP clinicians and medical staff. The recruiters were part of the CHOP Health Care Network, which includes multiple outpatient practices and primary care clinics. The controls were paediatric; nonetheless, we assume that the loss of power related to the few potential cases that might develop AN will not significantly influence the current analysis because of the low prevalence of AN. The controls had a mean (SD) age of 12.75 (4.2) years. Multidimensional scaling was used to confirm a proper match of genetic backgrounds.

***Ethics statement:*** Ethical approval was obtained by each of the individual study sites (PFCG). For the CHOP controls, CHOP’s Research Ethics Board approved the study, and informed consent was obtained from all participants or their parents.

***Funding statement:*** Funded by the Price Foundation. Andrew Bergen reports funding from Price Foundation Collaborative Group and NIH R01DA044014 and R01DA050495.

#### ANGI & BEGIN

The Anorexia Nervosa Genetics Initiative (ANGI) was a multi-site effort to collect biological samples and clinical information from individuals with AN and healthy controls [^9^](https://sciwheel.com/work/citation?ids=6345652&pre=&suf=&sa=0). Prof. Cynthia M. Bulik was the lead investigator, based at the University of North Carolina at Chapel Hill (dataset *usa2*), and collaborated with sites in Sweden (dataset *sedk*, PI: Dr. Mikael Landén), Denmark (dataset *ipsy*, PI: Prof. Preben Bo Mortensen), and Australia combined with New Zealand (dataset *aunz*, PIs: Prof. Nick Martin, Prof. Martin Kennedy, Dr. Jenny Jordan). Generally, across all sites (except *ipsy*), potential cases were recruited via professional organisations such as the Academy for Eating Disorders, conventional media, social media, (press) conferences and support groups, and word-of-mouth. The studies were designed as such to attract individuals who are most likely to fulfil the case criteria.

The Binge Eating Genetics INitiative (BEGIN) is a similarly designed follow-up study of ANGI, performed in the US and Sweden, and is focused on recruiting individuals with EDs other than AN, particularly BN and BED (dataset *sebe* [^10^](https://sciwheel.com/work/citation?ids=9336604&pre=&suf=&sa=0)). This is an ongoing project—initial data from Sweden only are included in this paper.

##### aunz

A detailed description of the Australia and New Zealand arm of ANGI is provided by Kirk et al., 2017 [^11^](https://sciwheel.com/work/citation?ids=9336853&pre=&suf=&sa=0). Residents from Australia (≥13 years) and New Zealand (≥14 years) were recruited between May 2013 and July 2016 and presented with an online questionnaire that was adapted from the Structured Clinical Interview for DSM-IV, section H (SCID-H). Amenorrhea was not required for being included as an AN case in the study. Controls were recruited via the QSkin Sun and Health Study [^12^](https://sciwheel.com/work/citation?ids=17418774&pre=&suf=&sa=0). The QSkin Sun & Health Study was established in 2010 to assess risk factors for skin cancers and is a randomly sampled cohort of individuals between 40 and 69 years residing in Queensland. The study participants were asked at the time of saliva collection whether they had ever been diagnosed with or treated for bipolar disorder, schizophrenia/psychosis, depression, obsessive-compulsive disorder, bulimia, anorexia nervosa, attention deficit hyperactivity disorder, or autism. Those who indicated no ED history were included as controls.

***Ethics statement:*** For ANGI-ANZ, all participants provided informed consent. Ethical approval for the Australian arm of the study was obtained by the QIMR Berghofer Human Research Ethics Committee; for New Zealand, this was provided by the Health and Disability Ethics Committee of the New Zealand Ministry of Health. For QSkin, ethical approval was obtained by the QIMR Berghofer Human Research Ethics Committee (QIMR-HEC approval P1309, P2034).

***Funding statement:*** The Anorexia Nervosa Genetics Initiative was an initiative of the Klarman Family Foundation. The QSkin Study is supported by a Clinical Trials and Cohort Grant [APP1185416] from the National Health and Medical Research Council of Australia (NHMRC) and was previously supported by grants APP1073898, APP552429, APP1063061. Jenny Jordan reports support from the Klarman Foundation, the University of Otago Research Fund, and the NIH. Tracey Wade reports support from the National Health and Medical Research Council Project Grant 480420.

##### ipsy

This dataset consists of cases and controls recruited via ANGI Denmark (ANGI-DK) plus additional cases and controls from the Lundbeck Foundation Initiative for Integrative Psychiatric Research (iPSYCH, <https://ipsych.dk/>) [^13^](https://sciwheel.com/work/citation?ids=4994648&pre=&suf=&sa=0). The recruitment approach for ANGI-DK was through the Danish national registers. The Danish Civil Registration System [^14^](https://sciwheel.com/work/citation?ids=2063634&pre=&suf=&sa=0) was established in 1968 and allows for the linkage of the national population registers. Everyone born between 1981 and 2005, who was a resident of Denmark on their first birthday, and who had a known mother was considered. Individuals in the Danish Psychiatric Central Research Register [^15^](https://sciwheel.com/work/citation?ids=5005585&pre=&suf=&sa=0) with a lifetime AN diagnosis (ICD-10 F50.0 or F50.1) were included as a case, and controls—matched on sex, date of birth, and without any major psychiatric disorder—were randomly selected from the same nationwide birth cohort. After identifying eligible cases and controls, their biological samples (phenylketonuria cards) were extracted from the Danish Neonatal Screening Biobank (DNSB) through their national identification number. Specifically, these biological samples consist of blood samples obtained by a heel prick postpartum, which were dried and subsequently stored at the DNSB. After extraction, the samples were genotyped on the Illumina PsychArray at the Broad Institute. Additional cases and controls were recruited through iPSYCH and genotyped similarly to the individuals recruited through the national registers.

***Ethics statement****:* Participants from ANGI-DK and the iPSYCH study did not provide written informed consent, as an exemption from consent was provided by the Danish Scientific Ethics Committee (Videnskabsetisk Komité). This exemption was most recently approved in 2018.

***Funding statement:*** ANGI-DK was part of the Anorexia Nervosa Genetics Initiative, which was an initiative of the Klarman Family Foundation (Mortensen, local PI; Bulik, PI). The iPSYCH was supported by grants from the Lundbeck Foundation (R102-A9118; R155-2014-1724; R248-2017-2003) and the Universities and University Hospitals of Aarhus and Copenhagen. Zeynep Yilmaz is supported by Independent Research Fund Denmark (DFF; Sapere Aude no. 1052-00029B); DFF (grant no. 3166-00063B, 4309-00050B); Lundbeck Foundation Ascending Investigator (R434-2023-269). Liselotte Petersen and Janne Larsen report funding from the Lundbeck Foundation (grant no. R276-2018-4581, recipient Petersen) and NIMH (R01MH120170).

##### sedk

The *sedk* cohort merged case-control data from ANGI recruitment efforts in Sweden with additional case individuals from clinical sites in Denmark.

###### ANGI-Sweden

Individuals were recruited from four different sources: (1) the Swedish ED quality registers RIKSÄT and Stepwise [^16^](https://sciwheel.com/work/citation?ids=17421662&pre=&suf=&sa=0); (2) the Stockholm Centre for Eating Disorders (SCÄ); (3) the community via conventional media and social media including the ANGI website (<https://angi.se/>); (4) LifeGene [^17^](https://sciwheel.com/work/citation?ids=5457213&pre=&suf=&sa=0), a study that was running from 2009 to 2019 to study how genes, environment, and lifestyle affect health. Most cases were recruited via the ED quality registers and were approached with a letter asking them to complete a questionnaire including the ED100K (described below). Cases approached through SCÄ were identified by research nurses who discussed the study and reviewed the consent. After providing consent, the study participant received an online questionnaire.

The ED100K questionnaire—used to assess case status—is a self-report assessment based on the Structured Clinical Interview for the DSM-5 Eating Disorder chapter and assesses criteria for AN, BN, BED, and OSFED. Individuals who met the criteria for AN were included as AN cases and if information was available, these individuals were subsequently assessed for AN subtypes (AN-R and AN-BP). Individuals who met the criteria for BN or BED were included as BE-NARROW and BE-BROAD cases. Individuals who have or have ever had regular episodes of binge eating with loss of control were included as BE-BROAD cases. Participants from LifeGene completed an online questionnaire that was similar to the ED100K algorithm and were included as cases if they met self-report criteria as defined above, or as controls if they screened negative for a history of EDs.

***Ethics statement:*** ANGI-Sweden has been approved by the ethical review board in Sweden (dnr 2013/112-31/2). For LifeGene, the Regional Ethical Review Board of Stockholm provided ethical approval, and all participants provided online consent.

***Funding statement:*** ANGI-Sweden was part of the Anorexia Nervosa Genetics Initiative, which was an initiative of the Klarman Family Foundation (Landén local PI; Bulik, PI). Cynthia Bulik is supported by the U.S. National Institute of Mental Health (R01MH136149;R01MH134039,R56MH129437; R01MH120170; R01MH124871; R01MH119084; R01MH118278; R01MH124871) and the Swedish Research Council (Vetenskapsrådet, award: 538-2013-8864). Patrick Sullivan reports support from the Swedish Research Council (Vetenskapsrådet, award D0886501), and NIMH R01 MH124871. Ruyue Zhang is supported by the Swedish Research Council (Vetenskapsrådet) grant no. 2022-00242. Andreas Birgegård reports funding from the Swedish Research Council (Vetenskapsrådet) grant no. 538-2013-8864, PI Bulik.

###### Danish clinical cases

Women born between 1947 and 1980 with at least one recorded hospital admission with an ICD-10 code of F50.0 or F50.1 identified through the Danish Psychiatric Biobank were included as AN cases.

***Ethics statement:*** All participants provided written informed consent before participating in the study. The study was approved by the Capital Region of Denmark’s Committees on Health Research Ethics (De Videnskabsetiske Komiteer for Region Hovedstaden) (approval number H-KF-01-024/01).

***Funding statement:*** Genotyping of these cases was supported by the Anorexia Nervosa Genetics Initiative, which was an initiative of the Klarman Family Foundation (Werge, local PI; Bulik, PI).

##### usa2

Cases and controls were recruited as described under ANGI, and additional controls were sourced from the Price Foundation AN Trios Study [^18,19^](https://sciwheel.com/work/citation?ids=929770,927936&pre=&pre=&suf=&suf=&sa=0,0). Case recruitment focused on reaching not just those in treatment for AN but also those who never received treatment. Potential participants (aged ≥12 years) either self-identified or were referred to take part in the study and received a brief screener to determine their eligibility for the study. Those interested and eligible progressed to sign an informed consent and online questionnaire (ED100K.v1). Upon completion of the questionnaire, participants provided blood samples in EDTA tubes which were drawn at a local laboratory, UNC, a physician’s office, or through a mobile phlebotomy company. DNA was extracted at the Rutgers University Cell and DNA repository of the National Institute of Mental Health, and samples were genotyped with the Illumina GSA chip. Furthermore, additional controls were recruited as part of the AN Trios study by advertisements in local communities of the various participating sites in the US and Europe. Individuals were included as controls if they were female, between the ages of 18 and 65 years, with a lifetime adult minimum BMI>19 and maximum BMI<27 kg/m^2^, no history of an ED, and no first-degree relative with an ED. Controls were matched to cases by study site, age, ancestry, and education. Samples were genotyped on the Illumina GSA array at the Broad Institute.

***Ethics statement:*** ANGI-US was approved by the Institutional Review Board of the University of North Carolina at Chapel Hill (IRB: 13-0081). All participants were broadly consented for research.

***Funding statement:*** The Anorexia Nervosa Genetics Initiative was an initiative of the Klarman Family Foundation (Bulik, PI).

##### sebe

The Binge Eating Genetics INitiative (BEGIN) Sweden study includes individuals with a diagnosis of BN or BED in the Swedish ED quality register Riksät, or individuals who answered the ANGI questionnaire but were not eligible for ANGI [^9^](https://sciwheel.com/work/citation?ids=6345652&pre=&suf=&sa=0). Individuals who consented to partake in the study received a saliva sampling kit. Furthermore, additional controls who were matched for age, sex, and geographical region were recruited via Ipsos, a telemarketing company. All individuals were aged ≥18 years. Cases were ascertained similarly to ANGI Sweden (see *sedk* - ANGI-Sweden).

Controls received screening questions before participating in BEGIN. Individuals were not included as controls if: they had ever had or had ever been treated for an ED; if they had a close family member with an ED; if they had received weight loss surgery; or if they received hormonal treatment (excl. anticonception).

***Ethics statement:*** This project has been approved by the ethical review board in Sweden. The fundament of the ethical permit comes from ANGI Sweden (dnr 2013/112-31/2) which has later been amended to also include individuals with another diagnosis than AN (dnr 2014/1563) and to change the study name to BEGIN (dnr 2016/1852-32).

***Funding statement:*** Funding was from the Swedish Research Council Swedish Research Council (Vetenskapsrådet, award: 538-2013-8864) (Bulik, PI).

#### Other cohorts

##### agds

Cases were recruited via the Australian Genetics of Depression Study (AGDS) [^20^](https://sciwheel.com/work/citation?ids=9143241&pre=&suf=&sa=0). AGDS was established in Australia to recruit a cohort of individuals with lifetime depression and investigate genetic and environmental risk factors, as well as response to antidepressants. Individuals were recruited based on (1) a media publicity campaign and (2) a pharmaceutical prescription history of common antidepressant medications in the last 4.5 years. Recruitment was initiated in September 2016. Controls were recruited via the QSkin Sun & Health Study (see summary under *aunz*) [^12^](https://sciwheel.com/work/citation?ids=17418774&pre=&suf=&sa=0).

AGDS study participants were asked to submit an online questionnaire. The question ‘*Have you ever been diagnosed with any of the following? Please select all that apply.*’ was used to assign individuals a case status. This was one of the first questions in the extensive online questionnaire. Individuals who answered ‘Yes’ to ‘Anorexia nervosa’ were coded as cases for AN, and individuals who answered ‘Yes’ to ‘Bulimia nervosa’ as cases for BE-BROAD. These individuals were not included in the BE-NARROW definition because of the single-item self-report nature of this questionnaire.

***Ethics statement***: Ethics approval for all aspects of the project was obtained from the QIMR Berghofer Human Research Ethics Committee (P2118 & P3434).

***Funding statement:*** The AGDS was primarily funded by National Health and Medical Research Council (NHMRC) of Australia grant 1086683. Sarah Medland is supported by NHMRC Leadership Grant APP1172917.

##### alsp

The Avon Longitudinal Study of Parents And Children (ALSPAC, <https://www.bristol.ac.uk/alspac/>) study is an ongoing population-based birth-cohort study of 14,541 mothers and their children (who were born between April 1, 1991 and December 31, 1992) residing in the southwest of England (UK). Criteria for defining cases (children and mothers) is detailed in Micali et al., 2015 [^21^](https://sciwheel.com/work/citation?ids=6298139&pre=&suf=&sa=0). Specifically, individuals with a diagnosis of AN at waves 14+ and 16+ were included as AN cases, and individuals who reported episodes of binge eating via the Youth Risk Behavior Surveillance System questionnaire [^22^](https://sciwheel.com/work/citation?ids=17421679&pre=&suf=&sa=0) (“eating a very large amount of food ≥ once a week and feeling out of control”) were included as cases for BE-BROAD (not BE-NARROW). Controls were individuals with no history of binge eating or any ED. Note that the number of controls between the AN and binge eating phenotype differ (AN 525 controls and binge eating 4204 controls). This is due to differential missingness on the ALSPAC questions.

***Acknowledgements:*** This publication is the work of the authors and Nadia Micali will serve as guarantor for the contents of this paper.

***Ethics statement***: The authors assert that all procedures contributing to this work comply with the ethical standards of the relevant national and institutional committees on human experimentation and with the Helsinki Declaration of 1975, as revised in 2008. Ethical approval for the study was obtained from the ALSPAC Ethics and Law Committee and the Local Research Ethics Committees. Informed consent for the use of data collected via questionnaires and clinics was obtained from participants following the recommendations of the ALSPAC Ethics and Law Committee at the time. The main caregiver initially provided consent for child participation, and from the age of 16 years, the offspring themselves have provided informed written consent.

***Funding statement:*** This study represents independent research partly funded by the UK National Institute for Health Research (NIHR) Biomedical Research Centre at South London and Maudsley NHS Foundation Trust and King’s College London. High-performance computing facilities were funded with capital equipment grants from the GSTT Charity (TR130505) and Maudsley Charity (980). This work was supported by the UK Medical Research Council and the Medical Research Foundation (MR/R004803/1). The UK Medical Research Council and Wellcome (102215/2/13/2 and 217065/Z/19/Z) and the University of Bristol provide core support for ALSPAC. A comprehensive list of grants is available on the ALSPAC website. This research was specifically funded by the NIHR (CS/01/2008/014) and the NIH (MH087786-01). GWAS data were generated by sample logistics and genotyping facilities at Wellcome Sanger Institute and LabCorp (Laboratory Corporation of America) using support from 23andMe. Mohamed Abdulkadir acknowledges grant support from the National Institute of Mental Health (R01MH120170).

##### biov

The study was conducted in the Synthetic Derivative (SD) database at the Vanderbilt University Medical Center (VUMC, Nashville, Tenessee, USA). The SD is a de-identified mirror image of the VUMC Electronic Health Records which was established in 1994. DNA was obtained from approximately 275,000 patients who consented to allow leftover clinical blood draws to be banked and linked to de-identified clinical data (BioVU). Further information on BioVU can be found at <https://victr.vumc.org/biovu-description/>.

AN cases were defined as individuals who have more than one of the following ICD codes: 307.1, F50.0, F50.00, F50.01, F50.02, F50.1 in the electronic health record. Individuals were defined as having BE-NARROW (and BE-BROAD) if they had more than one of the following ICD codes: 307.51, F50.81, F50.2. Controls consisted of all individuals without any ED ICD codes (i.e., 307.1, 307.5, 307.50, 307.51, 307.52, 307.53, 307.54, 307.59, F50, F50.0, F50.00, F50.01, F50.02, F50.2, F50.8, F50.81, F50.82, F50.89, F50.9, F50.3, F50.1, 307.81).

***Ethics statement:*** The study was approved by the VUMC Institutional Review Board (IRB #201609).

***Funding statement:*** This study was funded by R01MH118233 to Lea K Davis.

##### esbb

The Estonian Biobank is a large data-rich population-based biobank, covering approximately 20% of the adult population in Estonia (N=~210,000) [^23^](https://sciwheel.com/work/citation?ids=7020644&pre=&suf=&sa=0). All participants have signed an informed consent form and provided blood samples for genotyping. Electronic health records are regularly retrieved by linking to the national health databases and registries, such as the National Health Insurance Funds (NHIF) database, cause of death register, and hospital records.

Individuals with an ICD-10 diagnosis of F50.0 or F50.1 were defined as AN cases. Individuals with a F50.2 diagnosis were included as BE-NARROW cases, and individuals with a F50.2 or F50.3 diagnosis were included as BE-BROAD cases. Controls were defined as individuals without a diagnosis in the ICD-10 F50 chapter.

***Ethics statement:*** The research project has obtained approval from the Estonian Council on Bioethics and Human Research (1.1-12/624).

***Funding statement:*** This research in the Estonian Biobank was supported by the European Union through the European Regional Development Fund (Project No. 2014-2020.4.01.15-0012), and the Estonian Research Council’s grant No. PSG615. Krista Fischer reports support from grant PRG1197 by the Estonian Research Council.

##### fngn

The FinnGen research project ([www.finngen.fi](http://www.finngen.fi/)) was launched in 2017 with an aim to improve human health through genetic research [^24^](https://sciwheel.com/work/citation?ids=14248322&pre=&suf=&sa=0). The project combines genome information with digital health care data from national registries: the genotype data are linked to national hospital discharge, death, cancer, and medication reimbursement registries using the national personal identification numbers. The FinnGen study has combined approximately 200,000 existing samples from Finnish biobanks with approximately 300,000 samples from ongoing collections, primarily of hospitalised patients. The final data resource covers roughly 10% of the Finnish population.

Phenotypic information was obtained from Finnish hospital discharge registers. Individuals were defined as AN cases by the following criteria: those with an ICD-10 F50.0, F50.1 code; ICD-9 7830A, 3071A code; and/or ICD-8 784 code. Individuals were defined as BE-NARROW (and BE-BROAD) cases by the following: ICD-10 F50.2, F50.3; and/or ICD-9 3075B. Individuals who did not have any of the diagnoses mentioned in the above case definition were included as controls; individuals with any behavioural syndromes associated with psychological disturbances and physical factors were excluded.

***Ethics statement:*** The study was conducted in accordance with the principles of the Helsinki declaration. Written informed consent was obtained from all the study participants. For the Finnish Institute of Health and Welfare (THL)-driven FinnGen preparatory project and FinnGen project, all patients and control subjects had provided informed consent for biobank research, based on the Finnish Biobank Act. Alternatively, FINRISK and Health 2000 cohorts were based on study specific consents and later transferred to the THL Biobank after approval by Valvira, the National Supervisory Authority for Welfare and Health. Recruitment protocols followed the biobank protocols approved by Valvira.

***Funding statement:*** The funding agencies had no role in the design and conduct of the study; collection, analysis, and interpretation of data; or the writing of the manuscript or the decision to submit it for publication. The FinnGen project is funded by two grants from Business Finland (HUS 4685/31/2016 and UH 4386/31/2016) and nine industry partners (AbbVie, AstraZeneca, Biogen, Celgene, Genentech, GSK, MSD, Pfizer, and Sanofi). Samuli Ripatti and Jaakko Kaprio report funding from the Academy of Finland Center of Excellence in Complex Disease Genetics (grant # 352792).

##### gcfm

French GCAN samples (described under “GCAN/WTCCC-3”) that were not included in previous AN GWAS efforts were combined with new data from the Eating Disorders Continuum (EDC) at the Douglas Mental Health University Institute in Montreal. The EDC is the only large-scale, supraregional (province-wide) clinical-research-teaching program specialised in eating disorders (EDs) in the Province of Quebec, Canada. Having a supraregional mandate, the EDC serves as a main hub in the effort to care for Quebecers with EDs. The EDC offers inpatient, day program and outpatient services to people living in various regions of the province.

DNA samples provided by the Douglas EDC were sourced from blood obtained from people who participated in various genetic and epigenetic studies conducted in the program between 2001 and 2016. The studies implicated people with anorexia nervosa, bulimia nervosa, Eating Disorder Not Otherwise Specified, Other Specified Eating or Feeding Disorder, or no lifetime history of eating disorder. Individuals with EDs were recruited mainly from the EDC patient population. Controls were solicited mainly through public announcements. DSM-IV or DSM-5 diagnostic determinations were drawn using data from the Eating Disorder Examination Interview and/or the Eating Disorder Examination Questionnaire. Normal eaters had a body mass index greater than 18, lifetime absence of eating disorder, and no current major psychiatric disorder. Findings involving specific subgroups of the individuals described have been included in several previous publications from the EDC group, addressing gene-environment interactions acting in the EDs, or alterations in DNA methylation linked to the EDs and their comorbid features.

***Ethics statement:*** In all cases, there was consent to participation in one or more of the original studies, as well as to sharing of DNA with other research groups for additional genetic studies. All aspects were approved by the Research Ethics Boards of the Douglas Mental Health University Institute or Montreal West Island Integrated University Health and Social Services Centre.

***Funding statement:*** The original studies were funded by three different grants from the Canadian Institutes of Health Research (MOP-79490, MOP-57929, MOP-142717) or by industry support from Cogir Immobilier ("COGIR"). Funders had no role in study design, data collection, analysis and interpretation, report writing, or in decisions surrounding submission of papers for publication.

##### jans

Study participants were from the CAPSS220BED (ClinicalTrials.gov ID: NCT00210808) study, which is a multicentre, randomised, double-blind, placebo-controlled, flexible-dose study to assess the safety and efficacy of Topiramate in the treatment of moderate to severe BED associated with obesity.

All cases had a diagnosis of BED as defined by the DSM-IV and supported by the Structured Clinical Interview for DSM-IV Axis I Disorder Patient Edition (SCID-I/P). In addition, they reported binge-eating episodes >3 days per week in the week prior to baseline. Control samples were drawn from NINDS Human Genetics Resource center (neurologically normal White control panel NDPT020, NDPT079, NDPT084, NDPT090, NDPT093, NDPT094, NDPT095, NDPT096, NDPT098, and NDPT099) with known medical and family history managed by the Coriell Institute for Medical Research (Camden, NJ).

***Ethics statement:*** The clinical study was approved by the appropriate ethical review boards and followed the principles outlined in the Declaration of Helsinki for all human investigations. In addition, informed consent has been obtained from the study participants.

***Funding statement:*** The study was funded by Johnson & Johnson Pharmaceutical Research & Development, L.L.C.

##### moba

The Norwegian Mother, Father and Child Cohort Study (*MoBa*, <https://www.fhi.no/en/ch/studies/moba/>) is a population-based pregnancy cohort study conducted by the Norwegian Institute of Public Health [^25,26^](https://sciwheel.com/work/citation?ids=5934574,877759&pre=&pre=&suf=&suf=&sa=0,0). Participants were recruited from all over Norway from 1999-2008. The women consented to participation in 41% of the pregnancies. The cohort includes approximately 114,500 children, 95,200 mothers and 75,200 fathers. The current study is based on version 12 of the quality-assured data files released for research in January 2019. Blood samples were obtained from both parents during pregnancy and from mothers and children (umbilical cord) at birth [^27^](https://sciwheel.com/work/citation?ids=5949479&pre=&suf=&sa=0). The Medical Birth Registry, a national health registry containing information about all births in Norway from 1967 onwards, was used to identify sex registered at birth for the offspring generation. Full details of genotyping, quality control, phasing, imputation, and post-imputation quality control have been previously described [^28^](https://sciwheel.com/work/citation?ids=13218721&pre=&suf=&sa=0). After post-imputation quality control, 207,569 individuals with 6,981,748 autosomal, 174,462 chromosome X and 3,200 pseudoautosomal SNPs were available for analysis.

Parental cases were selected based on a combination of questionnaire data (collected from mothers and fathers during pregnancy and when their children were 8 years old) and diagnostic codes from specialist health care (reported in the Norwegian Patient Registry between 2008 and 2020). Offspring cases were based on maternal reports when children were 8 years old and diagnostic codes from specialist health care as above. Individuals were excluded as controls if they were (1) an AN or BE case or (2) were offspring of a parent who was an AN or BE case.

***Acknowledgements:*** The Norwegian Mother, Father and Child Cohort Study is supported by the Norwegian Ministry of Health and Care Services and the Ministry of Education and Research. We are grateful to all the participating families in Norway who take part in this on-going cohort study. Data from the Norwegian Patient Registry has been used in this publication. The interpretation and reporting of these data are the sole responsibility of the authors, and no endorsement by the Norwegian Patient Registry is intended nor should be inferred.

***Ethics statement:*** The establishment of MoBa and initial data collection was based on a licence from the Norwegian Data Protection Agency and approval from The Regional Committees for Medical and Health Research Ethics. The MoBa cohort is currently regulated by the Norwegian Health Registry Act. The current study was approved by The Regional Committees for Medical and Health Research Ethics (20311).

***Funding statement:*** We thank the Norwegian Institute of Public Health (NIPH) for generating high-quality genomic data. This research is part of the HARVEST collaboration, supported by the Research Council of Norway (#229624) and the South-Eastern Norway Regional Health Authority (#2023031). We also thank the NORMENT Centre, in collaboration with deCODE Genetics, for providing genotype data, funded by the Research Council of Norway (#223273), South-Eastern Norway Health Authorities and Stiftelsen Kristian Gerhard Jebsen. We also acknowledge funding from the U.S. National Institute of Child Health and Development (R01-HD047186, Bulik, PI). We further thank the Center for Diabetes Research, the University of Bergen for providing genotype data and performing quality control and imputation of the data funded by the ERC AdG project SELECTionPREDISPOSED, Stiftelsen Kristian Gerhard Jebsen, Trond Mohn Foundation, the Research Council of Norway, the Novo Nordisk Foundation, the University of Bergen, and the Western Norway Health Authorities. Ole Andreassen was funded by the Research Council of Norway (#324499, #324252), EU’s H2020 RIA grant (#964874), NordForsk project (#164218) and KG Jebsen Stiftelsen, Alexandra Havdahl was funded by the South-Eastern Norway Health Authorities (#2019097, #2020022), Helga Ask was funded by the Research Council of Norway (#324620) and NordForsk (#156298). Elizabeth Corfield was supported by the Research Council of Norway (#274611) and South-Eastern Norway Health Authorities fellowship (#2021045).

This work was performed on the Tjeneste for Sensitive Data facilities, owned by the University of Oslo, operated and developed by the Tjeneste for Sensitive Data service group at the University of Oslo, IT Department (USIT). The computations were performed on resources provided by Sigma2 - the National Infrastructure for High-Performance Computing and Data Storage in Norway.

##### net2

Dutch GCAN samples (described under “GCAN/WTCCC-3”) that were not included in previous AN GWAS efforts were combined with new data from the Utrecht Research Group Eating Disorders (URGE, <https://urge-eatingdisorders.nl/>). URGE is a case-only cohort and patients were recruited from the Eating Disorder clinic Rintveld (Altrecht, Zeist, the Netherlands) and provided informed consent and a blood sample to extract DNA.

ED diagnoses were determined according to the DSM-IV or DSM-5 criteria (whichever was appropriate at the time of data collection) by psychiatrists or (clinical) psychologists with ample experience in the treatment and classification of people with EDs. Each diagnosis was verified using questions from the Eating Disorders Examination.

***Ethics statement:*** Informed consent was obtained from all patients who provided DNA. The Biobank Ethics Committee of the University of Utrecht gave ethical approval for this work.

***Funding statement:*** Roger Adan was supported by grants of Netherlands Organisation for Scientific Research (NWO: 024.004.012, ALWOP.137 and OCENW.KLEIN.071).

##### ukb2

The UK Biobank represents a large population-based prospective study designed for in-depth exploration of both genetic and non-genetic factors that influence diseases prevalent in middle and old age [^29^](https://sciwheel.com/work/citation?ids=311441&pre=&suf=&sa=0). By 2010, 500,000 participants were part of the UK Biobank. Cases for the current study were identified via:

1. Mental health questionnaire (MHQ data) [^30^](https://sciwheel.com/work/citation?ids=8188505&pre=&suf=&sa=0): A questionnaire to examine lifetime and current mental health problems.
2. Hospital Episode Statistics (HES data): This category contains summary fields relating to diagnoses made during hospital inpatient admissions. The diagnosis data-fields lists, for each participant, all the distinct values for that data-field in the inpatient data, including main and secondary diagnoses, coded according to the International Classification of Diseases (ICD-9 & ICD-10).
3. Death register (DR data): This category contains coded data on the cause of death (International Classification of Diseases [ICD10]), obtained through linkage to national death registries.
4. Primary care linked data (GP data): UK Biobank has been liaising with various data suppliers and other intermediaries (including the main primary care computer system suppliers in England) to obtain primary care data for UK Biobank participants, all of whom have provided written consent for linkage to their health-related records.

Controls were defined as UK Biobank participants without a reported ED or binge eating (i.e., answered and indicated NOT having an ED or binge eating) or missing for that information.

***Ethics statement:*** UK Biobank is registered as a Research Tissue Bank (North West – Haydock Research Ethics Committee, 21/NW/0157). All data and sample applications may use this ethical clearance to conduct their research. Separate Research Ethics Committee (“REC”) or other ethical clearance is not required. This research was conducted under application 82087 (PI: Jonathan Coleman).

***Funding statement:*** This study represents independent research partly funded by the UK National Institute for Health Research (NIHR) Biomedical Research Centre at South London and Maudsley NHS Foundation Trust and King’s College London. High-performance computing facilities were supported by the King's College London Computational Research, Engineering and Technology Environment [^31^](https://sciwheel.com/work/citation?ids=17464684&pre=&suf=&sa=0).

##### ukch

This dataset consists of a combination of 1) cases recruited via the Charlotte’s Helix Project and 2) controls recruited via the National Institute for Health and Care Research BioResource (NBR, <https://bioresource.nihr.ac.uk>). Charlotte's Helix was supported by the Charlotte's Helix charity, in memory of the activist Charlotte Bevan, and consists of volunteers who had a diagnosis of anorexia nervosa or bulimia nervosa. Cases were ascertained through algorithms based on answers to the ED100K, reported diagnoses from clinicians, self-reported diagnoses, or registration notes. The genotyping data for cases were generated using Illumina Global Screening Array, while the genotyping data for controls were generated using the Affymetrix Axiom microarray (Supplementary Table 4).

***Ethics statement***: Charlotte's Helix participants were recruited using ethics approval provided by the Research Ethics Committee South Central Oxford C (15/SC/0388).

***Funding statement:*** This study represents independent research partly funded by the UK National Institute for Health Research (NIHR) Biomedical Research Centre at South London and Maudsley NHS Foundation Trust and King’s College London. Ulrike Schmidt receives salary support from the NIHR BRC, and from the Medical Research Council/Arts and Humanities Research Council/Economic and Social Research Council Adolescence, Mental Health and the Developing Mind initiative as part of the EDIFY programme (grant number MR/W002418/1).

##### ukd2

This dataset consists of the Genetic Links to Anxiety and Depression (GLAD)
Study [^32^](https://sciwheel.com/work/citation?ids=7749381&pre=&suf=&sa=0), Charlotte’s Helix (described under *ukch*), and the COVID19 Psychiatry and Neurological Genetics study (COPING) drawn from participants in the UK National Bioresource (NBR). The GLAD Study is a recontactable data resource of ~40,000 individuals based in the UK with lifetime anxiety and/or depression and forms part of the National Institute for Health and Care Research (NIHR) Mental Health BioResource. Participants are recruited online and as part of this answer multiple questionnaires, including the optional ED100K which provides information about their ED status. Participants have to be over 16 years old and live in the UK to participate in the GLAD Study. The Charlotte’s Helix data included in *ukd2* are independent and non-overlapping with regards to *ukch.* These data were separated because they were genotyped on different platforms – the genotyping data of the Charlotte’s Helix participants in *ukd2* were generated using the Affymetrix Axiom array.

The NBR is a databank and recontactable resource of volunteers who have provided medical, clinical, and biological data. Volunteers were recruited through a variety of approaches, including National Health Service (NHS) blood transfusion services and various disease/disorder-focused research efforts. Throughout the pandemic, NBR participants were given the opportunity to join the COPING study, which launched in April 2020. The COPING study contained questionnaires from the sign-up surveys of the GLAD study [^32^](https://sciwheel.com/work/citation?ids=7749381&pre=&suf=&sa=0) and the Eating Disorders Genetics Initiative (EDGI UK), as well as additional questionnaires to assess COVID-related variables, that is, experiences related to the COVID-19 pandemic. Notably, the ED100K questionnaire was mandatory in COPING. Participants first completed a baseline survey and then follow-up surveys, initially every two weeks but then monthly from August 2020. The last round of invites was sent on 19th January 2021.

COPING study participants come from multiple sub-cohorts of the NBR, including: the GLAD Study (n=14,948); EDGI UK (n=1010); the Inflammatory Bowel Disease BioResource (IBD; n=3203); NHS blood and transplant studies, including INTERVAL (n=4656), COMPARE (n=1928), and STRategies to Improve Donor Experiences (STRIDES; n=2808); and the Research Tissue Bank—Generic (n=4343). We restricted the number of controls from IBD via random selection to reflect the population prevalence of inflammatory bowel disease.

Both for GLAD and NBR, we ascertained AN or BE cases via self-reported diagnoses with (AN, BE-NARROW, and BE-BROAD) and without (AN and BE-BROAD) specification that this has been confirmed by a healthcare professional and/or DSM-5 algorithm-derived diagnoses (AN, AN-R, and AN-BP) via answers to the ED100K, or via self-reported binge eating plus frequency and duration via answers to the ED100K (BE-BROAD and BE-NARROW). Controls were defined as those without algorithm-derived or self-reported AN or BE and have indicated that they “have never suspected an eating disorder”.

***Ethics statement:*** The GLAD Study was approved by the London - Fulham Research Ethics Committee on 21st August 2018 (REC reference: 18/LO/1218) following a full review by the committee. The NIHR BioResource has been approved as a Research Tissue Bank by the East of England - Cambridge Central Committee (REC reference: 17/EE/0025). Prior to submission for ethical approval, this research was reviewed by a team with experience of mental health problems and their carers who have been specially trained to advise on research proposals and documentation through the Feasibility and Acceptability Support Team for Researchers (FAST-R): a free, confidential service in England provided by the NIHR Maudsley Biomedical Research Centre via King's College London and South London and Maudsley NHS Foundation Trust.

***Funding statement:*** This work was supported by the National Institute of Health Research (NIHR) BioResource, NIHR Biomedical Research Centre [IS-BRC-1215-20018], HSC R&D Division, Public Health Agency [COM/5516/18], MRC Mental Health Data Pathfinder Award (MC_PC_17,217), and the National Centre for Mental Health funding through Health and Care Research Wales.

##### jpan

Some DNA samples from Japanese participants collected as part of the GCAN effort (referenced under “GCAN/WTCCC-3”) were not included in previous AN GWAS efforts. These samples were newly genotyped for inclusion in this paper. As described in a previous publication [^2^](https://sciwheel.com/work/citation?ids=3303352&pre=&suf=&sa=0), cases met DSM-IV criteria for AN, with AN-free controls recruited in parallel.

***Ethics statement:*** Please refer to “GCAN/WTCCC-3”.

***Funding statement:*** The data and sample collection were supported by Grants-in-Aid for Scientific Research (20390201 and 23390201) awarded to G. Komaki by the Ministry of Education, Culture, Sports, Science, and Technology of Japan.

##### kran

Study participants were from HD16A1353 (cris.nih.go.kr ID: KCT 00210808) study, which was a multicentre study to investigate eating behaviours and the physical and psychological status of women with abnormal weight or patients with EDs.

Cases had a diagnosis of anorexia nervosa (restricting type or binge-purging type) as defined by the DSM-IV and supported by the Structured Clinical Interview for DSM-IV Axis I Disorder Patient Edition (SCID-I/P). Controls were university students recruited for the study and confirmed to have no ED after screening.

**Ethics statement:** The study was conducted according to the guidelines of the Declaration of Helsinki and approved by the Institutional Review Board of Inje University (INJE 2016-01-003-002).

**Funding statement:** This work was supported by the Korea Centers for Disease Control and Prevention Research Fund [grant number: 2016-ER6310-00, 2016].

### Supplementary Methods

#### Genotype quality control

Information about genotyping platforms for each cohort, and availability of chromosome X data, is documented in Supplementary Table 4. For cohorts included in our previous publication [^1^](https://sciwheel.com/work/citation?ids=7192844&pre=&suf=&sa=0), quality-controlled data were used for analyses of anorexia nervosa (AN). For analyses of AN subtypes and for the broad and narrow binge-eating phenotypes (BE-BROAD and BE-NARROW), additional exclusions of single-nucleotide polymorphisms (SNPs) and individuals were made where necessary to align these data with the quality control of newly included data, as described below. Summary-level data were available from contributing cohorts (Supplementary Table 1). Quality control of these datasets followed a basic shared protocol, with deviations as necessary for each cohort as described below.

In general, newly included data underwent the following process of quality control. Duplicate SNPs were removed, excluding the variant with the lower call rate. We also removed monomorphic variants, and variants with a call rate <95%. We then included samples with a call rate of ≥98%, and then applied a second SNP filter, excluding variants with a call rate of ≤98%, variants with >2% difference in call rates between cases and controls, and variants with significant Hardy-Weinberg Equilibrium (HWE) deviations of *P*<10^-10^ (cases) and *P*<10^-6^ (controls).

For the following quality control steps, we performed pruning of the variants based on linkage disequilibrium (LD) to only retain SNPs in relative linkage equilibrium with each other (PLINK --indep-pairwise 1500 150 0.2; window size was 1500 SNPs, 150 step size, excludes SNP pairs with *r^2^*>0.2). We also excluded high LD-regions (4 Mb inversion on chromosome 8 and an 8 Mb region covering the extended MHC region on chromosome 6) and only retained chromosomes 1-22. We compared sex assignments in the dataset with those imputed from chromosome X inbreeding coefficients (F^het^), excluding samples with a sex mismatch. We also excluded individuals with a genome-wide F^het^ deviation of >0.2. We calculated the proportion of identity-by-descent (IBD) or pi-hat for each pair within a cohort. The cohorts that used mixed models included relatives, but for the cohorts that did not use mixed models, we removed one member of each related pair (pi-hat>0.1875).

We then calculated principal components (PCs) on the set of pruned SNPs using reference data from the 1000 Genomes Project [^33^](https://sciwheel.com/work/citation?ids=790619&pre=&suf=&sa=0), which we limited to variants with a genotyping call rate of >99%, minor allele frequency (MAF) >0.01, and HWE deviations of *P*>10^-3^. For each cohort, we then plotted the PCs to visualise the largest homogenous cluster, which colocalised with reference individuals of European ethnicity. Retaining only these individuals with European ancestry, we then repeated the entire quality control process and calculated PCs for each cohort separately–without reference to the 1kGP–to use at a later stage in the association analyses (Supplementary Table 4). We restricted chromosome X analyses to individuals included in the autosomal analysis and conducted quality control as for autosomal SNPs.

#### Imputation

For all individual-level data cohorts where patient consent and national data privacy laws allowed, autosomal and chromosome X genotypes were imputed to TOPMed
freeze 8 [^34^](https://sciwheel.com/work/citation?ids=10461553&pre=&suf=&sa=0) using Minimac4, and phasing was performed with Eagle 2.4 [^35^](https://sciwheel.com/work/citation?ids=3224851&pre=&suf=&sa=0) via the TOPMed Imputation Server [^36^](https://sciwheel.com/work/citation?ids=2094306&pre=&suf=&sa=0). For individual-level data cohorts that could not be imputed to TOPMed, we imputed to the Haplotype Reference Consortium (HRC) [^37^](https://sciwheel.com/work/citation?ids=2311631&pre=&suf=&sa=0) using Minimac3, with phasing conducted via Eagle v2.3.5, implemented in the Ricopili pipeline [^38^](https://sciwheel.com/work/citation?ids=9702644&pre=&suf=&sa=0). Cohorts supplying summary statistics used TOPMed, HRC, or in some cases used bespoke imputation reference panels (Supplementary Table 4). We plotted imputation INFO scores against various MAF bins to check imputation quality, and we assessed missingness for each MAF bin.

#### Cohort by cohort deviations from established quality control and GWAS protocol

The following cohorts are fully described by the established quality control and GWAS protocol: *chop, fin1, fre1, itgr, sedk, gcfm, jans, net2, sebe, biov, alsp, moba*. Other cohorts, and their important deviations from these protocols, are listed below. For cohorts included in our previous publication (*gns2*, *net1*, *poco*, *spa1*, *ukd1*, *usa1*, *aunz*, *usa2*), these deviations were carried over from that publication [^1^](https://sciwheel.com/work/citation?ids=7192844&pre=&suf=&sa=0).

##### gns2

Variants failing the Hardy-Weinberg test at *P*<10^-6^ in AN cases were excluded.

##### net1

Variants failing the Hardy-Weinberg test at *P*<10^-6^ in AN cases were excluded, as were variants with a call rate difference >1% between AN cases and controls. The minor allele frequency of rs4492741 differed notably from reference data and this variant was removed.

##### poco

Variants with a call rate difference >1% between AN cases and controls were excluded, as were variants with a call rate in AN cases <99%, and variants with a minor allele frequency <1% in controls.

##### spa1

Variants with a call rate in AN cases <99% were excluded, as were variants failing the Hardy-Weinberg test at *P*<1.26 x 10^-6^ in AN cases.

##### ukd1

Variants with a call rate difference >1% between AN cases and controls were excluded, as were variants with a call rate in AN cases <99%, variants with a minor allele frequency <1% in controls, and variants failing the Hardy-Weinberg test at *P*<10^-6^ in AN cases.

##### usa1

Variants with a call rate in AN cases and controls combined <99% were excluded, as were variants failing the Hardy-Weinberg test at *P*<10^-6^ in AN cases.

##### aunz

Quality control for this cohort was conducted at the Queensland Institute of Medical Research. Prior to principal component analysis, variants were excluded for missing strand or position information, if they were duplicates of other genotyped variants, or if they showed inadequate cluster separation during genotyping (Illumina GenCall GenTran score <0.6). Variants were then excluded if they had an overall call rate across AN cases and controls combined <95%, or if they failed the Hardy-Weinberg test at *P*<10^-6^ in AN cases and controls combined. Participants were excluded if they were duplicates of existing individuals. Following principal component analysis, quality control proceeded as per protocol, except that variants were excluded if they had a call rate across AN cases <99%, and that one of each pair of related participants were excluded (pi-hat >0.2).

##### ipsy

iPSYCH analyses used the same participants included in our previous analyses [^1^](https://sciwheel.com/work/citation?ids=7192844&pre=&suf=&sa=0), with additional control filtering criteria. Specifically, controls were filtered for evidence of anorexia nervosa, bulimia nervosa, or eating disorders not otherwise specified. Quality control and imputation was performed on the entire iPSYCH cohort (N≈86,000), including psychiatric diagnoses beyond eating disorders [^39^](https://sciwheel.com/work/citation?ids=13438446&pre=&suf=&sa=0). Unlike our previous analyses [^1^](https://sciwheel.com/work/citation?ids=7192844&pre=&suf=&sa=0), in which 24 separate GWAS analyses were conducted within genotyping batches, all genotyping batches were included in a single mega-analysis with a covariate for genotyping batch.

##### usa2

Variants with a call rate <99% in AN cases were excluded, as were variants with minor allele frequency <1% in AN cases and in controls (considered separately), and variants failing the Hardy-Weinberg test at *P*<2.24x10^-3^ in controls. In addition, the minor allele frequency of rs183114681 differed notably from reference data, and the association of rs6917603 did not propagate to variants in linkage disequilibrium – both of these variants were removed.

##### agds

Imputed variants were limited to an INFO threshold of 0.6.

##### esbb

Imputation in the Estonian Biobank was conducted to a bespoke panel comprising whole genome sequence data from 2244 Estonians [^40^](https://sciwheel.com/work/citation?ids=4426917&pre=&suf=&sa=0).

##### fngn

Data from FinnGen underwent central quality control and imputation as described here [^24^](https://sciwheel.com/work/citation?ids=14248322&pre=&suf=&sa=0). Data release R7 was used for these analyses. In brief, quality control was similar to our established protocol, but some parameter settings differed, such as higher cut-offs for individual call rate (99.5%), heterozygosity outliers being determined based on standard deviations (+/- 4SD), and allele frequencies being compared to the bespoke imputation panel. Imputation was conducted to a bespoke panel constructed from high-coverage whole genome sequencing data from 3775 Finns (SISu v3).

##### ukb2

The imputed data were restricted to variants with R^2^≥0.4, MAF≥0.01. GWAS analysis was conducted using REGENIE’s mixed model with 10 PCs, genotyping batch, and UK Biobank recruitment centre as covariates.

##### ukch

Charlotte’s Helix (Illumina) and NBR (Affymetrix) datasets were imputed separately, and then merged [^41^](https://sciwheel.com/work/citation?ids=652493&pre=&suf=&sa=0). Each set of imputed data was limited to variants with R^2^≥0.3, MAF≥0.01. Following merging, we excluded variants with differences in MAF and R^2^ greater than 0.05 between the two datasets. GWAS analysis followed the established protocol.

##### ukd2

GLAD, Charlotte’s Helix, and the NBR dataset underwent quality control and imputation separately and were restricted to variants with R^2^≥0.3, MAF≥0.001. Imputed data were merged, and we excluded variants with MAF<0.01, with differences in MAF and R^2^>0.05 between any pairing of the three datasets, and/or with significant Hardy-Weinberg Equilibrium deviations of *P*<10^-10^ (cases) and *P*<10^-6^ (controls). GWAS analysis was conducted using REGENIE’s mixed model with 10 principal components and array version as covariates.

#### Power analysis

We estimated what magnitude of relative risk we were powered to detect using Genetic Power Calculator [^42^](https://sciwheel.com/work/citation?ids=3289200&pre=&suf=&sa=0). Assuming perfect linkage disequilibrium between the marker and the risk SNP (D'=1), we set the number of cases to ½ N_eff_ (Supplementary Table 1) and the control:case ratio to 1. We specified lifetime risk as described for SNP-based heritability in the main text. We assumed an additive model, set the p-value significance threshold to 5x10^-8^, and assessed MAF ranging from 0.01 to 0.5. For AN, we also conducted analyses based on the previous GWAS of Watson et al for comparison [^1^](https://sciwheel.com/work/citation?ids=7192844&pre=&suf=&sa=0&dbf=0), calculating number of cases as the sum of ½ Neff across cohorts, which totalled 23,161 (compared to 36,753 in our analyses).

#### Polygenic prediction

##### Cohort selection

We selected the target cohorts based on effective sample size (N_eff_ half >1000). The predictive performance of BE-BROAD, BE-NARROW and AN PRS on each available outcome were tested in these cohorts. The target cohorts for BE-BROAD, BE-NARROW, and AN are as follows:

- BE-BROAD:
  - *aunz, sedk, ukb2, ukd2* (data available to PGC-ED analysts);
  - *alsp, moba, esbb* (data from cohorts that contributed summary statistics).
- BE-NARROW:
  - *aunz, sedk, ukb2, ukd2* (data available to PGC-ED analysts);
  - *esbb, fngn,* and *moba* (data from cohorts that contributed summary statistics).
- AN:
  - *gns2, chop, aunz, sedk, ukb2, ukd2* (data available to PGC-ED analysts);
  - *usa2, esbb, fngn, moba* and *ipsy* (data from cohorts that contributed summary statistics).

We also sought to explore the difference of BE-BROAD and AN PRS in three subgroups of individuals with eating disorders compared to controls (i.e., individuals without BE-BROAD and AN): (a) those with BE-BROAD only; (b) individuals with both BE-BROAD and AN (BE+AN), and (c) those with AN only.

We selected cohorts with sufficient sample size for ≥2 subgroups to assess differences between subgroups. Specifically, we selected *aunz* and *sedk* to assess how the PRS levels differ from controls in BE+AN individuals and in those with AN only. In these cohorts, we assessed binge-eating behaviour (and thus fulfilling criteria for BE-BROAD) in individuals with AN. Consequently, the AN-only group reflects a mix of individuals with both AN-R and AN-BP as some individuals with AN-BP may never have had a binge-eating episode but are classified for AN-BP due to the presence of purging behaviour. BE+AN individuals will always have a diagnosis of AN-BP. Both *aunz* and *sedk* comprise individuals recruited through the Anorexia Nervosa Genetics Initiative (ANGI) and thus consist only of individuals with AN and do not have any individuals with BE-BROAD-only.

We also performed subgroup analyses in cohorts not specifically recruited for AN. We selected *ukb2* and *ukd2* as additional target cohorts to assess differences between all subgroups. In *ukb2,* no symptom or AN subtype-level data were available, so individuals with lifetime diagnoses of both AN and BN/BED are considered to be in the BE+AN group. *ukd2* mostly comprises data on eating disorders in the context of depression. Therefore, it may not be representative of the general clinical population of individuals with AN and binge-eating behaviour.

#### Sex-specific analyses

##### Cohort selection

As the participants from most cohorts are predominantly female, the combined sample sizes were not sufficient for a male-only GWAS meta-analysis. We therefore opted to perform female-only meta-analyses for BE-BROAD and AN and use this as base data for generating a PRS. We tested this PRS in males in selected cohorts to examine whether BE-BROAD and AN PRS built in females could be applied to predict BE-BROAD and AN risk in males.

Both for the female-only GWAS and the male-only target, we included only cohorts where sex-specific case and control n were both >100. For female-only BE-BROAD GWAS (37,580 cases, 691,808 controls), we included all cohorts except *fin1, fre1, gcfm, gns2, itgr, net1, poco, spa1, ukd1,* and *net2*. For female-only AN GWAS (21,898 cases, 694,722 controls), we included all cohorts except *itgr, spa1, ukd1, jans,* and *net2.* For the male-only BE-BROAD target (1055 cases, 38,046 controls), we included *alsp, ukd2,* and *moba* (we opted to include *moba* despite N_case_=98 after excluding relatives). For the male-only AN (1524 cases, 388,891 controls) target, we included *fngn, ipsy,* and *ukb2*.

#### Casual mixture modelling

To validate estimates of polygenicity and discoverability from MiXeR, we additionally conducted analyses using SBayesRC [^43^](https://sciwheel.com/work/citation?ids=16432573&pre=&suf=&sa=0&dbf=0). SBayesRC is a method for enhancing polygenic risk scoring through identifying likely causal variants from GWAS summary statistics, leveraging functional annotations. As part of this, it outputs estimates of polygenicity, as well as grouping the proposed causal variants into sets of zero, very small (per SNP heritability=0.001%), small (0.01%), medium (0.1%), and large (1%) effect. In this instance, we were interested only in these, rather than the broader functions of SBayesRC. SBayesRC and MiXeR are distinct methods, reliant on different assumptions about genetic architecture, and as such results are not expected to align perfectly but should be broadly consistent when applied to the same data. We conducted analyses on our BE-BROAD and AN summary statistics, restricted to variants with MAF≥0.01 and INFO≥0.6, using per-variant Neff (calculated by Ricopili as Neff_half, and then doubled) in place of N, and following the protocols provided on the SBayesRC GitHub (https://github.com/zhilizheng/SBayesRC).

#### Mendelian randomisation

Three main assumptions are made for Mendelian randomisation (MR): (1) the genetic instrument must have a strong association with the exposure, (2) the genetic instrument is independent of confounding factors in the relationship between the exposure and the outcome, and (3) the outcome is associated with the genetic instrument only through the effect of the exposure [^44,45^](https://sciwheel.com/work/citation?ids=435648,3802982&pre=&pre=&suf=&suf=&sa=0,0).

Of these three assumptions, only the first can be tested directly by calculating the F-statistic [^45^](https://sciwheel.com/work/citation?ids=3802982&pre=&suf=&sa=0). An F<10 suggests potential weak instrument bias [^46^](https://sciwheel.com/work/citation?ids=12085635&pre=&suf=&sa=0). In addition, for BMI as exposure independent genome-wide significant SNPs (the threshold of *P*<5x10^-9^ was used in GWAS on BMI due to denser imputation data) are used as instrument variables (IV). To create IV for BE and AN, we used a threshold of *P*<5x10^-6^ to include more SNPs. Heterogeneity of IV and pleiotropy lead to a violation of the second and third assumption and to biased effect estimates. We used Cochran's Q-statistic to test for IV heterogeneity. Cochran's Q-statistic assesses whether causal estimates of SNPs are comparable, with a significant finding (*P*<0.05) indicating heterogeneity [^47^](https://sciwheel.com/work/citation?ids=3128075&pre=&suf=&sa=0). Heterogeneity may have several causes, of which horizontal pleiotropy is the most likely [^48^](https://sciwheel.com/work/citation?ids=5935431&pre=&suf=&sa=0). To specifically investigate horizontal pleiotropy, we calculated MR–Egger regression and performed an MR-PRESSO analysis. If Egger's intercept is not significantly different from zero, horizontal pleiotropy is unlikely [^49^](https://sciwheel.com/work/citation?ids=3932894&pre=&suf=&sa=0). MR PRESSO uses a global bias test to evaluate whether the removal of potentially pleiotropic instruments results in a significant difference in the overall causal estimate and provides a corrected causal estimate after the removal of pleiotropic instruments [^50^](https://sciwheel.com/work/citation?ids=5149171&pre=&suf=&sa=0). Simulation studies have shown that MR PRESSO and Cochran’s Q-statistic are substantially more sensitive and powerful to horizontal (global) pleiotropy than Egger's intercept [^48,50^](https://sciwheel.com/work/citation?ids=5149171,5935431&pre=&pre=&suf=&suf=&sa=0,0). We excluded from the analysis SNPs identified by MR PRESSO as pleiotropic.

Since it is unlikely that all IVs fulfil the assumptions for instrumental variables, and since it is not possible to test the fulfilment of these assumptions, many different MR methods have been developed. These methods differ in their robustness to various violations of assumptions. Because no method alone can provide infallible proof of causality, the use of different methods was recommended to assess whether a causal effect detected by MR is robust [^51,52^](https://sciwheel.com/work/citation?ids=6057311,7851037&pre=&pre=&suf=&suf=&sa=0,0). Accordingly, we applied the following MR methods. The inverse-variance-weighted (IVW) method assumes that all ratio estimates provide independent evidence of the causal effect and that all genetic variants are valid instruments [^49,53^](https://sciwheel.com/work/citation?ids=3932894,686003&pre=&pre=&suf=&suf=&sa=0,0). MR–Egger considers an intercept term interpreted as the average pleiotropic effect of the genetic variants included in the analyses. If the pleiotropic effects are distributed independently of the genetic associations with the risk factor (InSIDE assumption: INstrument Strength Independent of Direct Effect), then the MR–Egger estimate is a consistent estimate of the causal effect as both the sample size and the number of genetic variants increase [^49,54^](https://sciwheel.com/work/citation?ids=3932894,14422501&pre=&pre=&suf=&suf=&sa=0,0). Mode-based estimation (MBE; simple mode, weighted mode) consistently estimates the true causal effect under the assumption that across all instruments, the most frequent value of bias due to pleiotropy is zero (ZEro-modal pleiotropy assumption (ZEMPA)) [^54^](https://sciwheel.com/work/citation?ids=14422501&pre=&suf=&sa=0). Simple MBE is less accurate than weighted MBE, but simple MBE is less prone to bias due to violations of the InSIDE assumption. Median-based estimators are consistent even when up to 50% of the instruments are invalid. The weighted median estimator has similar efficiency to the IVW method, but the simple median estimator is less efficient than either the IVW or the weighted median method [^55^](https://sciwheel.com/work/citation?ids=3128078&pre=&suf=&sa=0). The penalised weighted median estimator is robust in the case of IV heterogeneity [^56^](https://sciwheel.com/work/citation?ids=15226040&pre=&suf=&sa=0).

In the case of IV heterogeneity as indicated by Cochran's Q statistic, two methods robust to heterogeneity were used in addition to the penalised weighted median method [^56^](https://sciwheel.com/work/citation?ids=15226040&pre=&suf=&sa=0). The contamination mixture method has a good overall performance (the lowest mean squared error) with up to 40% invalid instruments compared to other robust methods. The contamination mixture method identifies distinct subgroups of genetic variants with mutually similar causal estimates [^57^](https://sciwheel.com/work/citation?ids=8965662&pre=&suf=&sa=0). MR-Lasso extends the IVW model to include an intercept term for each genetic variant. These intercept terms represent associations between genetic variants and the outcome that bypasses the risk factor. The causal effect is estimated by weighted linear regression, with the intercept terms subjected to lasso penalisation. Lasso penalisation shrinks the intercept of the valid instruments to zero [^56^](https://sciwheel.com/work/citation?ids=15226040&pre=&suf=&sa=0).

Forest and scatter plots were used to visualise the combined results of single and multi-SNP analyses (Supplementary Figures 27 and 28). The scatter plots show single SNP effects on the exposure against single SNP effects on the outcome with corresponding standard deviations and estimated regression lines of the multi-SNP analyses.

In case of unavailability of SNPs in GWAS for the outcome phenotype, we used proxy-SNPs [^58^](https://sciwheel.com/work/citation?ids=13832867&pre=&suf=&sa=0). The SNPs with linkage disequilibrium (LD) of at least r^2^≥0.8 (on the basis of GRCh37, Ensembl version 87, 1000 genomes: phase 3 version 5 for European ancestry) were extracted from the *in silico* tool SNIPA (http://www.snipa.org. Accessed in November 2024) [^59^](https://sciwheel.com/work/citation?ids=5302844&pre=&suf=&sa=0). Selection criteria for proxy-SNPs were defined: 1st highest r2, 2nd smallest distance to the lead SNP [^58^](https://sciwheel.com/work/citation?ids=13832867&pre=&suf=&sa=0).

Analyses were performed using R 4.3.2, and packages *TwoSampleMR* (0.6.8; https://github.com/MRCIEU/TwoSampleMR) [^60^](https://sciwheel.com/work/citation?ids=5380003&pre=&suf=&sa=0), *Mendelian Randomization* (0.10.0; [https://CRAN.R-project.org/package=MendelianRandomization](https://cran.r-project.org/package=MendelianRandomization)), and *MR-PRESSO* (1.0; <https://github.com/rondolab/MR-PRESSO>) [^50^](https://sciwheel.com/work/citation?ids=5149171&pre=&suf=&sa=0). Figures were created with the R package *forestplot* 3.1.5.

#### Variant heterogeneity in AN

Despite an increase in N of 64%, variant discovery in our AN GWAS did not increase markedly compared to the previous AN GWAS [^1^](https://sciwheel.com/work/citation?ids=7192844&pre=&suf=&sa=0&dbf=0). We hypothesise that this is due to the ascertainment strategy of new cohorts, resulting in a more heterogeneous signal in the new analyses. To determine whether this hypothesis is supported by our results, we examined I^2^ heterogeneity statistics in three sets of summary statistics: (1) our main AN GWAS, (2) the AN GWAS from Watson et al [^1^](https://sciwheel.com/work/citation?ids=7192844&pre=&suf=&sa=0&dbf=0), and (3) a repeat of our AN GWAS restricted to only the new AN cohorts (i.e. gcfm, sebe, agds, alsp, biov, esbb, fngn, moba, net2, ukb2, ukch, ukd2). We included updated analyses from the UK Biobank in our new GWAS, so UK Biobank is present in both the Watson GWAS and in the new AN cohorts. All sets of summary statistics were derived from the Ricopili pipeline and had heterogeneity statistics calculated in METAL (Main Methods). For each set of summary statistics, we calculated the mean and standard deviation of I^2^ across all variants, and binned I^2^ across all variants into bins of ≤25%, 25-50%, 50-75%, and >75%.

We then conducted a series of sensitivity analyses. We restricted variants to those shared across the different summary statistics (1+2, and 1+2+3). We also restricted variants to those in linkage equilibrium, in two ways. First, we clumped all variants from the Watson et al GWAS summary statistics without filtering on *P* using PLINK2 [^61^](https://sciwheel.com/work/citation?ids=1158431&pre=&suf=&sa=0&dbf=0), with the European ancestry individuals from 1000 Genomes as an LD reference [^33^](https://sciwheel.com/work/citation?ids=790619&pre=&suf=&sa=0&dbf=0). We removed variants in linkage disequilibrium (r^2^>0.9) with variants with a lower p-value within 1Mb. Second, we pruned the set of variants shared across all three sets of summary statistics using PLINK and the European ancestry individuals from 1000 Genomes as an LD reference, removing variants in linkage disequilibrium (r^2^>0.9) with a window of 1Mb. We then repeated the I^2^ calculations above, and additionally compared the I^2^ values for each shared variant across the summary statistics using linear regression and cross-tabulating I^2^ bins. Finally, we repeated analyses of shared variants limiting only to variants with *P*<5x10^-8^ and *P*<0.05 in Watson et al [^1^](https://sciwheel.com/work/citation?ids=7192844&pre=&suf=&sa=0&dbf=0).

### Supplementary Results

#### Association meta-analyses

LDSC attenuation ratio estimates were significantly >0 across all phenotypes (0.070 [AN] - 0.59 [AN-R]). A larger attenuation ratio can indicate that the observed signal results from genome-wide inflation rather than true polygenic signal. Ratios were larger in the comparatively underpowered BE-NARROW and the AN subtypes, but smaller in BE-BROAD and AN.

##### BE-NARROW

For BE-NARROW, we analysed 15 datasets comprising 15,175 cases and 1,217,725 controls and identified five genomic loci (Supplementary Table 5, Supplementary Figure 15).

##### AN subtypes

For the AN subtypes, we estimated a *h^2^*_SNP_ of 9% (SE=3%) for AN-R and 15%
(SE=2%) for AN-BP (Supplementary Table 3). We identified one genomic locus for AN-R (13 datasets, 2524 cases, 33,374 controls) on chromosome 2 which was not reported in the previous AN GWAS [^1^](https://sciwheel.com/work/citation?ids=7192844&pre=&suf=&sa=0) (Supplementary Table 5, Supplementary Figure 15). We did not identify any genome-wide significant loci for AN-BP (14 datasets, 5209 cases, 33,951 controls; Supplementary Figure 15).

#### Power analyses (AN-R, AN-BP, BE-NARROW)

We had 80% power to detect a genotypic relative risk of 1.05-1.26 for BE-BROAD, 1.09-1.44 for BE-NARROW, 1.07-1.36 for AN, 1.21-2.19 for AN-R, and 1.15-1.81 for AN-BP. The AN GWAS of Watson et al had 80% power to detect a genotypic relative risk of 1.09-1.46, indicating an increase in power with our analyses, although we caution that these analyses assume all cases contribute equally, which is unlikely in this instance (Main Discussion, Heterogeneity analyses in Supplementary Results).

#### Female-only analyses

##### BE-BROAD

The female-only GWAS in BE-BROAD was largely consistent with the full GWAS (Supplementary Table 6). The three top loci remained genome-wide significant, along with the locus on chromosome 3. The two loci on chromosomes 11 and 12 did not pass genome-wide significance in female-only analyses, and two loci on chromosomes 1 and 11 passed genome-wide significance in the female-only analyses but not the full analyses. However, the effect estimates of the top SNPs across all loci (full GWAS and female-only) did not significantly differ between GWAS (z-test *P*>0.05), indicating differences in results were an artefact of sampling error, consistent with the smaller size of the female-only analyses.

### AN

For AN, the three top loci from the main GWAS remained genome-wide significant in female-only GWAS. However, remaining loci were more variable—the chromosome 10 locus remained genome-wide significant, but the remaining four loci (on chromosomes 1, 5, 10, and 12) did not pass significance in the female-only analyses. Six additional loci passed genome-wide significance in the female-only analyses but not the full analyses, including the chromosome 2 locus previously observed in Watson et al [^1^](https://sciwheel.com/work/citation?ids=7192844&pre=&suf=&sa=0) and loci on chromosomes 3, 4, 6, 20 and 22. However, the effect estimates of the top SNPs across all loci (full GWAS and female-only) did not significantly differ between GWAS (z-test *P*>0.05).

#### BE-BROAD in cohorts not ascertained for anorexia nervosa (BE-BROAD_NAAN_)

Genetic signal in our BE-BROAD GWAS may partly be influenced by AN, given that 18% of our BE-BROAD cases have (known) AN (Supplementary Table 11). As a sensitivity analysis, we conducted an additional BE-BROAD GWAS, excluding cohorts that specifically focused on AN recruitment. We will refer to this phenotype as “not-ascertained-for-AN BE-BROAD” (BE-BROAD_NAAN_). We calculated SNP-*r_g_* between BE-BROAD_NAAN_ and a selection of other traits based on their associations with BE-BROAD and AN, and tested for significant differences from the respective SNP-*r_g_* with BE-BROAD (Supplementary Figure 17; Supplementary Table 12). BE-BROAD_NAAN_ had significantly stronger genetic associations with the anthropometric traits: body fat percentage was more strongly associated with BE-BROAD_NAAN_ than with BE-BROAD, as was BMI, suggesting that AN cases in the excluded cohorts may be masking this genetic signal. Further, whilst non-significant, the SNP-*r_g_* between BE-BROAD and OCD changed significantly after removing the AN-focused cohorts, remaining non-significant but becoming negative. We observed a similar significant change for physical activity: BE-BROAD displayed a non-significant but stronger positive SNP-*r_g_* than BE-BROAD_NAAN_. Such findings suggest that some of the originally observed genetic signal may be attributed to AN cases.

#### Casual mixture modelling

Univariate MiXeR analysis of BE-BROAD and of AN fit well, with positive AIC values compared to the LDSC model, suggesting MiXeR modelling is justified (Supplementary Table 8). Univariate analysis for BE-BROAD resulted in comparable discoverability estimates to psychiatric disorders like ADHD and ASD (2.73x10^-5^), but much lower polygenicity than is typically observed in psychiatric disorders (3195 variants explaining 90% of heritability) [^62^](https://sciwheel.com/work/citation?ids=18356583&pre=&suf=&sa=0&dbf=0). Estimates of BE-BROAD heritability on the observed scale were comparable to those from LDSC (h^2^=6%, SE=0.1%). In comparison, univariate MiXeR analyses for AN resulted in a higher discoverability (3.53x10^-5^) and polygenicity (8926 variants), similar to that previously reported for bipolar disorder [^62^](https://sciwheel.com/work/citation?ids=18356583&pre=&suf=&sa=0&dbf=0). Again, estimates of AN heritability on the observed scale were comparable those from LDSC (h^2^=20%, SE=0.4%).

Bivariate MiXeR analyses should be viewed with caution in this instance: although the MiXeR best overlap model fit better than a maximal model of full overlap, it did not fit better than a minimal model of least overlap possible given the observed genetic correlation (Supplementary Table 8). This is in part because the low polygenicity of BE-BROAD restricts the modelling space for the MiXeR model, with the minimal overlap model still requiring >75% of the putative causal variants for BE-BROAD to be shared with AN. This caveat aside, the results from the bivariate analysis are consistent with other results in this study, showing a sizable overlap of causal variants between BE-BROAD and AN with highly concordant directions of effect, as well as non-trivial non-overlapping causal components for both traits, consistent with the differing genetic correlations we observed.

Estimates of observed-scale heritability from SBayesRC were slightly higher than from LDSC for BE-BROAD (h^2^=8%, SE=0.1%) and comparable for AN (h^2^=21%, SE=0.1%). However, unlike univariate MiXeR, polygenicity estimates were similar between BE-BROAD (53,739 non-zero effect variants) and AN (52,054), with very small and small effect components predominant in both traits (Supplementary Table 8). MiXeR and SBayesRC rely on different assumptions and underlying models, and so inconsistent results can occur, but nonetheless, we caution against over-interpreting our MiXeR results given this lack of validation.

#### S-PrediXcan

##### BE-BROAD

In addition to two experiment-wide significant results (Main Text), 41 gene-tissue associations (representing 17 unique genes) were significant at the within-tissue threshold (Supplementary Table 16). Among these, 11 within-tissue associations were in the central nervous system, and five in gastrointestinal tissues.

To test whether tissue association patterns were significant or merely artifactual findings reflecting uneven power across sample sizes, we performed exact binomial tests to assess tissue enrichment in our associations (Supplementary Table 31). None of these tests were significant after accounting for multiple testing.

### AN

In addition to 312 experiment-wide significant gene-tissue associations (Main Text), 280 gene-tissue associations were significant at a within-tissue threshold (Supplementary Table 16).

In exact binomial tests for tissue enrichment in AN, no tests were significant after accounting for multiple testing (Supplementary Table 31).

##### BE-NARROW

For BE-NARROW, we observed nine experiment-significant gene-tissue associations in two loci on chromosome 3 (four genes: *HADHA*, *KLHDC8B*, *PODXL2*, *PRKAR2A*; nine tissues) with the most significant association in *KLHDC8B*-Heart, Atrial appendage
(*P*=2.08 x 10^-12^; Supplementary Figure 19, Supplementary Table 16).

In exact binomial tests for tissue enrichment in BE-NARROW, no tests were significant after accounting for multiple testing (Supplementary Table 31).

##### AN subtypes

For AN-BP subtype S-PrediXcan, we found eight experiment-significant gene-tissue associations in two loci encompassing three genes (*CALML6*, *KLHDC8B*, *PRKAR2A*) and eight tissues, with the most significant association in *PRKAR2A*-Sigmoid colon
(*P*=1.33 x 10^-13^; Supplementary Figure 19, Supplementary Table 16). At a tissue-specific significance threshold, there were 108 gene-tissue associations (14 genes: *C3orf62*, *CALML6*, *CELSR3*, *DALRD3*, *HSPA2*, *KLHDC8B*, *MLKL*, *NEDD4*, *NICN1*, *PPOX*, *PRKAR2A*, *RP11-57206.1*, *ZBTB25*, *ZNF33A*; 44 tissues).

In AN-R, we observed one experiment-significant hit in *DLX1* (chr2) in brain frontal cortex tissue (*P*=5.96 x 10^-8^; Supplementary Figure 19, Supplementary Table 16). An additional 25 gene-tissue associations were significant at a tissue-specific significance level (five genes: *DLX1*, *PRKAR2A*, *LIPT2*, *EPM2A*, *NBPF20*; 23 tissues).

No tests were significant in exact binomial tests for tissue enrichment after accounting for multiple testing (Supplementary Table 31).

#### S-MultiXcan

We ran S-MultiXcan using harmonised GWAS summary statistics and S-PrediXcan MASHR model output for each of our ED traits for all genes (N=22,241).

##### BE-BROAD

S-MultiXcan results of BE-BROAD phenotype found ten significantly associated genes (eight loci, six chromosomes) (*P*<2.22 x 10^-6^), with the most significant association with gene *SAP30* on chromosome 4 (*P*=6.97 x 10^-46^; Supplementary Figure 20, Supplementary Table 17).

##### BE-NARROW

Sixteen genes were significantly associated (13 loci, 10 chromosomes) with the BE-NARROW phenotype (*P*<1.40 x 10^-6^), with gene *SPTBN2* on chromosome 11 most significantly associated (*P*=1.45 x 10^-34^; Supplementary Figure 20, Supplementary Table 17).

### AN

Forty-three genes across 14 loci on 13 chromosomes were significantly associated with AN phenotype (*P*<1.45 x 10^-6^), with the most significant association of AN with the chromosome 3 gene *USP19* (*P*=2.00 x 10^-40^; Supplementary Figure 20, Supplementary Table 17).

##### AN subtypes

For AN subtypes, nine genes were significantly associated with AN-BP (four loci, three chromosomes) (*P*<4.87 x 10^-7^) with the most significant association on chromosome 14 with gene *AKAP5* (*P*=1.31 x 10^-15^). For AN-R, there were eight genes (eight loci, seven chromosomes) significantly associated with the phenotype (*P*<1.83 x 10^-6^), with the top gene *CCNB1IP1* on chromosome 14 (*P*=7.38 x 10^-24^; Supplementary Figure 20, Supplementary Table 17).

#### MAGMA

##### BE-NARROW

In the gene analysis for BE-NARROW, MAGMA identified 13 genes significant after Bonferroni correction (*P*<2.58 x 10^-6^; Supplementary Table 18). As in BE-BROAD, the FTO gene had the strongest association (*P*=2.1 x 10^-17^). The MAGMA gene set and drug set analyses did not yield significant results, but drug-class analyses identified enrichment in corticosteroids (particularly those in group III, potent) as well as enrichment in anxiolytics (Supplementary Table 21).

##### AN subtypes

In the gene analysis for AN-R, MAGMA identified 6 genes significant after Bonferroni correction (*P*<2.59 x 10^-6^), all in the same locus on chromosome 2q31 (Supplementary Table 18). The MAGMA gene set and drug set analyses did not yield significant results, but drug-class analyses identified enrichment in hypnotics and sedatives, and anti-epileptic drugs, as well as in multiple classes of anti-inflammatory drugs (Supplementary Table 21).

In the gene analysis for AN-BP, MAGMA identified 11 genes significant after Bonferroni correction (*P*<2.59 x 10^-6^) across two loci – the gene-dense locus on chromosome 3p21 and a locus on chromosome 14q23 (Supplementary Table 18). The MAGMA gene set and drug-class analyses did not identify Bonferroni-corrected significant gene sets or drug classes, but the targets of inositol (vitamin B8) were enriched for signal (Supplementary Table 20).

#### Cell and tissue gene expression specificity

We used stratified LDSC to estimate the enrichment of SNP-based heritability for BE-BROAD and AN among genes specifically expressed in GTEx human tissues and in human brain cell types from the Human Brain Atlas. After accounting for multiple testing, none of the associations was significant. However, we observed two notable trends. First, both phenotypes had greater enrichment in brain tissues than in other tissues. Second, the strongest relationship was with cortical excitatory pyramidal neurons in BE-BROAD (supercluster “Upper_layer_intratelencephalic”). For AN, we mainly observed enrichment in the cortical and subcortical inhibitory neurons represented in five superclusters.

#### Influence of BMI

##### GWAS-by-subtraction

In cohorts restricted to those not ascertained by AN, BMI accounted for 22% of the genetic variance in BE-BROAD.

We calculated SNP-*r_g_* between the full BE-BROAD and AN phenotypes and their *NonBMI* components to assess how subtracting BMI affects the relationship with other traits (Supplementary Figure 18, Supplementary Table 14). Among psychiatric traits, the BE-BROAD *NonBMI* factor had a higher positive SNP-*r_g_* with schizophrenia, but a lower SNP-*r_g_* with ADHD. The SNP-*r_g_* of BE-BROAD with educational attainment increased and became significant after subtracting BMI, and the association with self-rated health became non-significant. Subtraction of BMI-related genetic risk attenuated positive SNP-*r_g_* between BE-BROAD and most metabolic and anthropometric phenotypes, except childhood obesity and childhood BMI, which still showed significantly positive SNP-*r_g_* with BE-BROAD.

We see a mirrored pattern in the *NonBMI* AN factor: first, subtracting BMI significantly increased correlations between AN and several psychiatric phenotypes, including ADHD, posttraumatic stress disorder symptoms, and cross-disorder risk. Furthermore, subtracting BMI significantly reduced associations between AN and educational attainment. We observed reduced associations between AN and most other metabolic and anthropometric phenotypes, with only waist-hip ratio, body fat percentage, fat mass, fasting insulin, and BMI-adjusted fasting insulin still being significantly negatively correlated with *NonBMI* AN.

Most of the anthropometric genetic correlations with BE-BROAD and with AN were reduced following our subtraction of the genetic variance of BMI. However, some correlations remained significant, such as those between BE-BROAD and childhood BMI, and of AN with fat mass and body fat percentage. Similarly, the metabolic genetic associations with AN were attenuated to non-significance when subtracting BMI, except for the correlation with fasting insulin (age-and-sex adjusted and BMI-adjusted). In contrast, with a few exceptions like the AN-ADHD genetic correlation discussed above, genetic correlations with psychiatric traits were largely unaffected by subtracting the BMI component. Even when BMI is accounted for, BE and AN are genetically distinct psychiatric phenotypes. Similar patterns of SNP-*r_g_* between the EDs and other psychiatric disorders (with the notable exception of OCD) persist, as do differing patterns of SNP-*r_g_* with impulsive traits (positive with BE-BROAD, not correlated with AN), and with education and fasting insulin levels (positive with AN, not correlated with BE-BROAD).

Overall, BMI accounts for a small but meaningful portion of BE-BROAD and AN risk, acting in opposing directions. Genetic correlations between BE-BROAD and AN, and between the EDs and associated traits converge when assessing the non-BMI components. When considering specific SNPs, there were no major changes in patterns of associations after accounting for BMI. Ultimately, this leads to the question of, beyond BMI, what differentiates risk for BE from that of AN? Though patterns of genetic correlation converge in the *NonBMI* analysis, certain traits show notable divergence, including OCD (associated with AN and not with BE-BROAD), and risk tolerance (associated with BE-BROAD and not with AN), highlighting a potential influence of biological predisposition relevant to uncertainty and risk in differentiating BE and AN risk. Accounting for BMI affected the genetic correlation of EDs with diverse traits, not just with anthropometric traits, indicating that shared risk with anthropometric factors may be relevant to many psychological, psychiatric, and behavioural traits.

One major consideration in understanding how BMI, specifically, impacts genetic risk for AN, is that low BMI is definitional to an AN diagnosis in the current samples. However, there is ongoing debate over the degree to which low BMI is a necessary diagnostic criterion. For instance, individuals may lose significant amounts of weight and experience all other hallmark symptoms of AN and yet not receive this diagnosis. Recent research suggests that this presentation (currently termed ‘atypical AN’) may be quite common and, compared to AN with low weight presentations, has similarly debilitating symptoms, medical complications, and non-ED comorbidities [^63^](https://sciwheel.com/work/citation?ids=15199151&pre=&suf=&sa=0). Relevant to the current analysis, genetic associations between AN and (low)BMI and associated traits may be higher in the current sample as compared to an ascertainment and phenotyping strategy in which low BMI is not central to an AN case definition.

With regards to BE-BROAD, a phenotyping and ascertainment strategy which included individuals across the weight spectrum (including a potential oversampling of those with binge eating who also meet AN criteria), and focused on behavioural indicators rather than diagnostics, suggests that the positive associations between BEBROAD and BMI are likely to be robust.

##### Mendelian randomisation

###### Exposure: BE-BROAD, Outcome: BMI

Of a total of 41 SNPs associated with BE-BROAD at *P*<5 x 10^-6^, MR-PRESSO identified 22 as pleiotropic. These were excluded from further analysis, leaving an instrumental variable (IV) of 19 SNPs. All these SNPs are strong instruments (minimum F-value 19.47). Neither heterogeneity nor pleiotropy were indicated by the relevant tests. Multiple methods supported the hypothesis that BE-BROAD is causal for higher BMI (Supplementary Figure 27a, Supplementary Table 15).

###### Exposure: *NonBMI* BE-BROAD, Outcome: BMI

Of the 28 SNPs with *P*<5 x 10^-6^, 27 were present in the summary statistics for BMI. Ten SNPs were classified as pleiotropic by MR PRESSO. Thus, an IV was formed with 17 SNPs. All SNPs can be classified as strong instruments (minimum F-value 21.09). Tests indicated possible heterogeneity and pleiotropy, but multiple methods consistently supported the hypothesis that BE-BROAD is causal for a higher BMI, even after accounting for shared genetics (Supplementary Figure 27c, Supplementary Table 15).

###### Exposure: AN, Outcome: BMI

There were 84 independent SNPs available as an IV (*P*<5 x 10^-6^). MR-PRESSO identified 16 SNPs as pleiotropic and these SNPs were excluded from the analysis, leaving an IV formed from 68 SNPs. All these SNPs can be considered as strong instruments (minimum F-value 17.81). The Q-test indicated that the IV was heterogeneous, therefore attention should be focused on the heterogeneity robust methods when looking at the results. The MR-Egger intercept did not indicate pleiotropy, but the MR-PRESSO global test was significant. However, no pleiotropic SNPs were identified. Multiple methods supported the hypothesis that AN is causal for lower BMI (Supplementary Figure 28a, Supplementary Table 15).

###### Exposure: *NonBMI* AN, Outcome: BMI

A total of 67 independent SNPs (*P*<5 x 10^-6^) associated with *NonBMI* AN were available. 15 SNPs were classified as pleiotropic by MR-PRESSO and were therefore excluded, so the final IV was constructed from 51 SNPs (minimum F-value 20.85). The Q-test showed that IV was significantly heterogeneous. The intercept of MR-Egger was not significant, but the global test of MR-PRESSO indicated horizontal pleiotropy. No methods yielded an estimate significantly different from zero, and so there was no support for the hypothesis that AN has a causal effect on BMI after accounting for shared genetics (Supplementary Figure 28c, Supplementary Table 15).

###### Exposure: BMI, Outcome: BE-BROAD

For BMI acting on BE-BROAD, 670 independent SNPs (*P*<5 x 10^-9^) were available as an IV. Of these, 95 were not present in the summary statistics for BE-BROAD, but 58 proxies were available. Of the 633 SNPs, 6 SNPs were classified as pleiotropic by MR-PRESSO and excluded to leave 627 SNPs (minimum F-value 25.00). The Q-test indicated heterogeneity, and pleiotropy was also probable. Multiple methods were consistent with the hypothesis that there may be a causal effect of a higher BMI on a higher risk of BE (Supplementary Figure 27b, Supplementary Table 15).

###### Exposure: BMI, Outcome: *NonBMI* BE-BROAD

An IV was constructed from 642 SNPs (minimum F-value 25.00): out of the 670 index SNPs for BMI, 76 were not available in the *NonBMI* BE-BROAD summary statistics, 53 proxy SNPs were included, and four SNPs were excluded as pleiotropic. Both pleiotropy and heterogeneity were indicated by statistical tests. Although point estimates were consistently OR>1 (which would be in support of the hypothesis that higher BMI causally increases the risk of BE), the different methods are not consistently significant and therefore no statement can be made as to whether BMI causally affects BE after accounting for shared genetics (Supplementary Figure 27d, Supplementary Table 15).

###### Exposure: BMI, Outcome: AN

For BMI acting on AN, 670 independent SNPs (*P*<5 x 10^-9^) were available as an IV. Of these SNPs, 11 were identified as pleiotropic by MR-PRESSO and therefore excluded, and 71 were not available in summary statistics for AN. Proxies were found for 49 unavailable SNPs, resulting in an IV of 642 SNPs (minimum F-value 25.00). The Q-test indicated heterogeneity. The MR-Egger intercept was not significant, but the MR-PRESSO global test result was consistent with horizontal pleiotropy, although MR-PRESSO did not detect further pleiotropic SNPs. Robust methods supported the hypothesis that low BMI causally increases the risk of AN (Supplementary Figure 28b, Supplementary Table 15).

###### Exposure: BMI, Outcome: *NonBMI* AN

The same 642-SNP IV was available for BMI as in the BMI-to-AN analysis above. Again, tests indicated heterogeneity of the IV and pleiotropy. Although point estimates were consistently OR>1 (which would be in support of the hypothesis that *higher* BMI causally increases the risk of AN), the different methods are not consistently significant and therefore no statement can be made as to whether there is an effect of BMI on AN after accounting for shared genetics (Supplementary Figure 28d, Supplementary Table 15).

#### Polygenic prediction

##### Liability-scale R^2^

###### BE-BROAD

The average liability-scale variance of BE-BROAD explained by BE-BROAD PRS was 0.32% (range 0.08% – 2.02%), compared to 0.29% (range 0.09% – 1.35%) by BE-NARROW PRS and 0.53% (range 0.01% – 4.84%) by AN PRS, respectively (Supplementary Table 25). After excluding cohorts in which all the BE-BROAD cases also have AN diagnosis (*aunz, sedk*), the average liability-scale variance of BE-BROAD explained by BE-BROAD PRS did not change (0.32%, range 0.08% – 2.02%), but that explained by AN PRS dropped to 0.12% (range 0.01% – 1.6%), suggesting a larger variance of BE-BROAD being explained by BE-BROAD PRS than AN PRS in individuals with binge-eating behaviours who do not have AN.

###### BE-NARROW

Likewise, the average liability-scale variance of BE-NARROW explained by BE-NARROW PRS was 0.64% (range 0.19% – 1.81%), compared to 0.73% (range 0.21% – 1.99%) by BE-BROAD PRS and 0.70% (range 0.01% – 2.44%) by AN PRS, respectively (Supplementary Table 25). After excluding cohorts in which all the BE-BROAD cases also have AN diagnosis (*aunz, sedk*), the average liability-scale variance of BE-NARROW explained by BE-NARROW PRS and BE-BROAD PRS changed minimally to 0.57% (range 0.19% – 1.81%) and 0.67% (range 0.21% – 1.99%), respectively, but dropped to 0.46% (range 0.01% – 2.02%) for the liability-scale R^2^ of BE-NARROW explained by AN PRS.

#### AN

The average liability-scale variance of AN explained by AN PRS was 2.32% (range 1.21% – 4.72%), compared to 0.32% (range 0.06% – 0.85%) by BE-BROAD PRS and 0.29% (range 0.14% – 0.73%) by BE-NARROW PRS, respectively (Supplementary Table 25).

##### PRS predictive performance

We evaluated predicted performance of PRS by subtracting the ROC AUC (or PR AUC) of the null model (i.e., a model with only genetic PCs as predictors) from the ROC AUC (or PR AUC) of the full model (i.e., a model with PRS and genetic PCs as predictors). Given a random prediction model has a ROC AUC of 0.5 and a perfect prediction model has a ROC AUC of 1, we observed wide variability in the null model such that ROC AUC varied from 0.519 to 0.828 when only using genetic PCs to predict BE-BROAD and varied from 0.525 to 0.864 when only using genetic PCs to predict AN. Since some of our cohorts are very imbalanced in terms of case:control ratio (i.e., proportion of cases ranges from 0.3% to 52% for AN and 0.1% to 59% for BE-BROAD), we included PR AUC in addition to ROC AUC to evaluate prediction performance as PR AUC provides a more accurate evaluation of prediction performance in imbalanced datasets, in which ROC AUC may provide misleading conclusions on prediction performance due to false interpretation on specificity [^64^](https://sciwheel.com/work/citation?ids=1400780&pre=&suf=&sa=0).

###### BE-BROAD

Individuals with 1SD higher BE-BROAD PRS had an average OR of 1.11 for BE-BROAD (average 95% CI: 1.08 – 1.14, all *P*≤0.022), an average OR of 1.20 for BE-NARROW (average 95% CI: 1.14 – 1.27, all *P*≤7.14$\times$10^-7^), and an average OR of 1.16 for AN (average 95% CI: 1.10 – 1.22, all *P*≤0.023; Supplementary Figure 23, Supplementary Table 25).

BE-BROAD PRS contributed to an average increase of 0.009 (range: 0.005 - 0.027) in the area under the receiver operating characteristic curve (ROC AUC) and 0.004 (range: 0.001 - 0.019) in the precision-recall curve (PR AUC) for BE-BROAD prediction. In addition, it contributed to an average increase of 0.013 (ROC AUC) and 0.004 (PR AUC) for BE-NARROW prediction and an average increase of 0.010 (ROC AUC) and 0.005 (PR AUC) for AN prediction (Supplementary Table 25).

###### BE-NARROW

Individuals with 1 SD higher BE-NARROW PRS had an average OR of 1.18 for BE-NARROW (Supplementary Figure 23; average 95% CI: 1.12 – 1.25, all *P*≤1.72x10^-5^), an average OR of 1.11 for BE-BROAD (average 95% CI: 1.08 – 1.14, all P≤8.40x10^-9^ except *alsp* where *P*=0.058), and an average OR of 1.15 for AN (average 95% CI: 1.09 – 1.21, all *P*≤6.62x10^-4^; Supplementary Figure 23, Supplementary Table 25).

Using BE-NARROW PRS, the average improvement in AUC was 0.012 (ROC) and 0.004 (PR) to predict BE-NARROW, 0.01 (ROC) and 0.004 (PR) to predict BE-BROAD, and 0.009 (ROC) and 0.005 (PR) to predict AN.

#### AN

Individuals with 1 SD higher AN PRS had an average OR of 1.50 for AN (average 95% CI: 1.42 – 1.59, all *P*≤2x10^-16^), an average OR of 1.10 for BE-BROAD (average 95% CI: 1.07 – 1.13, all *P*≤4.54x10^-3^ except *alsp* where *P*=0.11), and an average OR of 1.17 for BE-NARROW (average 95% CI: 1.11 – 1.23, all *P*≤4.23x10^-6^ except *moba* where *P*=0.176; Supplementary Figure 23, Supplementary Table 25). This suggests a diminished association of AN PRS with BE in large population-based cohorts where individuals with BE mostly do not have AN (e.g., *moba, alsp*). Indeed, after excluding the two cohorts where all BE cases are AN-BE cases (*aunz, sedk*), individuals with 1 SD higher AN PRS had a lowered average OR of 1.05 for BE-BROAD (average 95% CI: 1.02 – 1.08), and a lowered average OR of 1.12 for BE-NARROW (average 95% CI: 1.06 – 1.17).

AN PRS contributed to an average increase of 0.046 (range: 0.006 - 0.084) in ROC AUC and 0.034 (range: 0.001 - 0.073) in PR AUC for AN prediction. Among all prediction models tested, the prediction of AN using AN PRS produced the highest prediction improvement (an average increase of 0.046 in ROC AUC and an average increase of 0.034 in PR AUC). The average improvement in AUC to predict BE-BROAD using AN PRS was 0.011 (ROC) and 0.005 (PR), and 0.012 (ROC) and 0.005 (PR) to predict BE-NARROW. When excluding cohorts where all the BE cases also have AN diagnosis (*aunz, sedk*), the average improvement in AUC for AN PRS to predict BE-BROAD dropped substantially to 0.004 (ROC) and 0.0004 (PR) and dropped to 0.004 (ROC) and 0.001 (PR) to predict BE-NARROW, suggesting little predictive capability of AN PRS on binge-eating behaviour outside of the context of AN.

##### Sex-specific PRS

###### BE-BROAD

We examined whether the effect of BE-BROAD PRS on BE-BROAD was different in males versus in all samples across three selected cohorts. We found that the OR of BE-BROAD per 1 SD higher BE-BROAD PRS in males in *ukd2* was 1.20 [95% CI: 1.09, 1.33], compared to OR of 1.23 [95% CI: 1.17, 1.30] in all samples in *ukd2* (*P*_diff_=0.68). In *moba*, the OR was 1.15 [95% CI: 0.94, 1.40] in males, with a wide confidence interval due to small case number (N=98), and 1.06 [95% CI: 1.05, 1.08] in all samples (*P*_diff_=0.47). In *alsp*, the OR was 1.06 [95% CI: 0.91, 1.24] in males and 1.09 [95% CI: 1.01, 1.18] in all samples (*P*_diff_=0.74). Given the PRS result in males was not significant in both *moba* (*P*=0.18, N_case_=98) and *alsp* (*P*=0.43, N_case_=179) due to small samples, future studies with more male BE cases are required to explore if there is any sex-specific genetic difference in binge-eating behaviour (Supplementary Table 26, Supplementary Figure 24).

#### AN

The OR of AN per 1 SD higher AN PRS in males in *fngn* was 1.07 [95% CI: 0.96, 1.20], compared to OR of 1.32 [95% CI:1.26, 1.39] in all samples in *fngn* (*P*_diff_=8.75x10^-4^). In *ipsy*, the OR was 1.41 [95% CI: 1.23, 1.60] in males and 1.49 [95% CI: 1.44, 1.54] in all samples (*P*_diff_=0.40). In *ukb2*, the OR was 1.09 [95% CI: 0.90, 1.31] in males and 1.45 [95% CI: 1.38, 1.54] in all samples (*P*_diff_=0.003). Given the limited data, it is hard to draw a definitive conclusion on the transferability of AN PRS based on female-only GWAS for AN risk prediction in male individuals. Future studies with more samples and data from men with AN are needed to confirm whether there is a sex-specific genetic effect on AN (Supplementary Table 26, Supplementary Figure 24).

##### Subgroup PRS

In addition to the results for BE-BROAD and AN (reported in the main text), we also assessed the difference in BE-NARROW PRS across the three subgroups. Overall, we found BE-NARROW PRS to be elevated in all subgroups compared to controls (9.76x10^-19^ 9.76$\times$10^-19^ ≤*P*≤0.036). Among the subgroups, comorbid and BE-BROAD-only subgroups tended to have the highest BE-NARROW PRS with no significant difference across these two groups (*P*≥0.19). The AN-only group tended to have lower BE-NARROW PRS, although the difference between the AN-only and the comorbid groups was only significant in *sedk* (*P*=1.1x10^-^4) and that between AN-only and BEB-only groups was only significant in *ukb2* (*P*=1.9x10^-4^; Supplementary Table 27, Supplementary Figure 25).

#### Comparison of BE-BROAD with Burstein et al

A previous publication reported a GWAS of a model-derived phenotype aimed at capturing binge eating disorder in the Million Veteran Program [^65^](https://sciwheel.com/work/citation?ids=15256599&pre=&suf=&sa=0). As this study primarily reported an algorithmic phenotype distinct from our BE definitions, and additionally controlled for BMI, we did not include it within our BE cohorts.

Within individuals from European ancestries, Burstein et al implicated two loci at genome-wide significance in BE. Both loci were on chromosome 6, and the authors mapped these to the *HFE* and *MCHR2* genes respectively. The lead variants rs79220007 and rs17789218 had *P=*0.018 and 0.0025 respectively in BE-BROAD with a consistent magnitude and direction of effect. Results were similar in the *non-BMI* BE-BROAD analysis (*P=*0.0089 and 0.0077 respectively). As such, we replicated these findings controlling for two tests, but lacked power to replicate them at a genome-wide level of significance. Notably, rs17789218 is significantly associated with BMI [^66^](https://sciwheel.com/work/citation?ids=7190974&pre=&suf=&sa=0) (and also passes genome-wide significance in the GWAS of the shared component between BMI and BE-BROAD) and so this finding may be at risk of collider bias [^67^](https://sciwheel.com/work/citation?ids=12655476&pre=&suf=&sa=0).

Burstein et al additionally report genetic correlations of their model-derived phenotype with external traits. Burstein et al report modest positive correlations of BE with psychiatric disorders, ranging from 0.21 with AN to 0.52 with depression. The equivalent correlations with BE-BROAD are 0.43 for AN (previous freeze) and 0.39 for depression (Supplementary Table 9), suggesting the pattern of correlations with psychiatric disorders is similar, but correlations with specific disorders differ. Results for *non-BMI* BE-BROAD are similar to those for BE-BROAD (Supplementary Table 14). Burstein et al also report small negative genetic correlations with cognitive phenotypes (-0.11 to -0.13), which we do not observe with BE-BROAD; a modest negative genetic correlation with subjective well-being (-0.21, we observe -0.41 with BE-BROAD), and modest positive genetic correlations with risk taking (0.30) and lifetime smoking (0.19), which mirror our results from BE-BROAD (0.21 and 0.30 respectively). These results are not materially different when considering *non-BMI* BE-BROAD. Overall, we see both agreement and disagreement between genetic correlations with BE-BROAD and those reported by Burstein et al.

Burstein et al report several significant gene-sets associated with BE – we did not find any significant gene-sets associated with BE-BROAD. It is difficult to make strong conclusions about this difference, given methodological differences in the gene-set analyses performed in the two studies.

#### Variant heterogeneity in AN

Mean I^2^ across variants was lower in Watson et al (7.52%, SD=11.13%) than in our main AN analysis (7.96%, SD=11.97%), and was considerably higher in the new AN cohorts (10.5%, SD=15.95%). Bins of I^2^ are shown in Supplementary Table 28. More variants were present in higher bins (>25%) in the main AN analysis and the new AN cohorts than in Watson et al, supporting the assertation that the new AN cohorts introduced greater heterogeneity into the analyses. Analyses of shared variants across the three sets of summary statistics showed good concordance in I^2^ bins between analyses, with a general pattern of higher I^2^ in the main AN analysis and the new AN cohorts than in Watson et al (Supplementary Table 28). Sensitivity analyses limited to variants in linkage equilibrium and variants associated with AN in Watson et al were consistent with the unrestricted analyses (Supplementary Table 28).
