## Supplementary Figures for "Genome-wide association studies of binge-eating behaviour and anorexia nervosa yield insights into the unique and shared biology of eating disorder phenotypes"

### Supplementary Figures 1-6: Forest plots for the six genome-wide significant associations with BE-BROAD in order of position on the genome.

Each row represents one cohort, with the final row showing the meta-analytic result. Initial information gives the variant name, effect and non-effect alleles, and variant position, followed by a summary of directions of effect, heterogeneity p-value, and heterogeneity I statistic. Main data columns include the imputation information metric (info), the association p-value, the frequency of the variant in cases (f_ca) and in controls (f_co), and the log odds ratio and standard error of the association, which are also visualised in the plot. The genotyping indicator (ngt) was set to missing for all cohorts and should be ignored.

#### Supplementary Figure 1

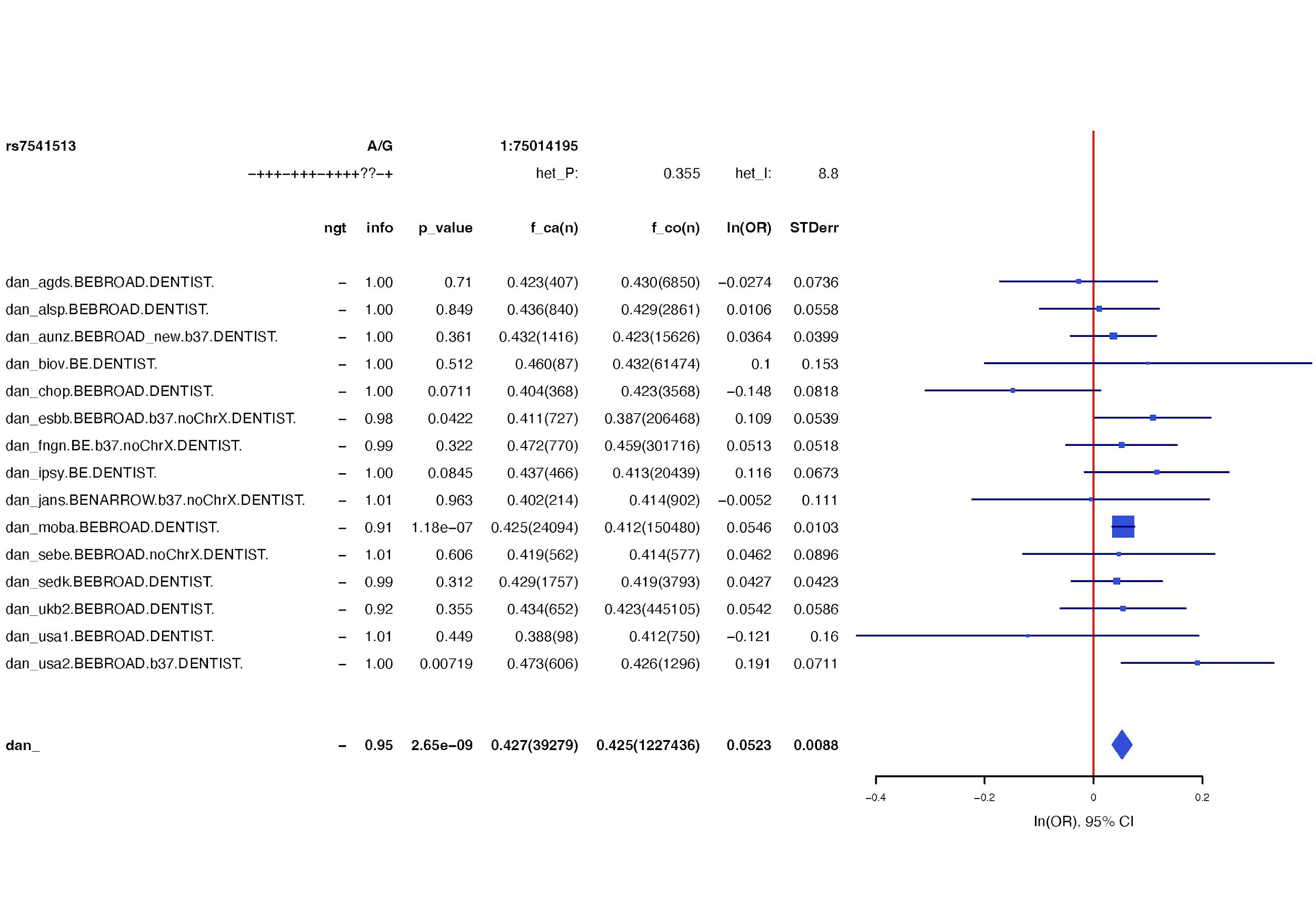

#### Supplementary Figure 2

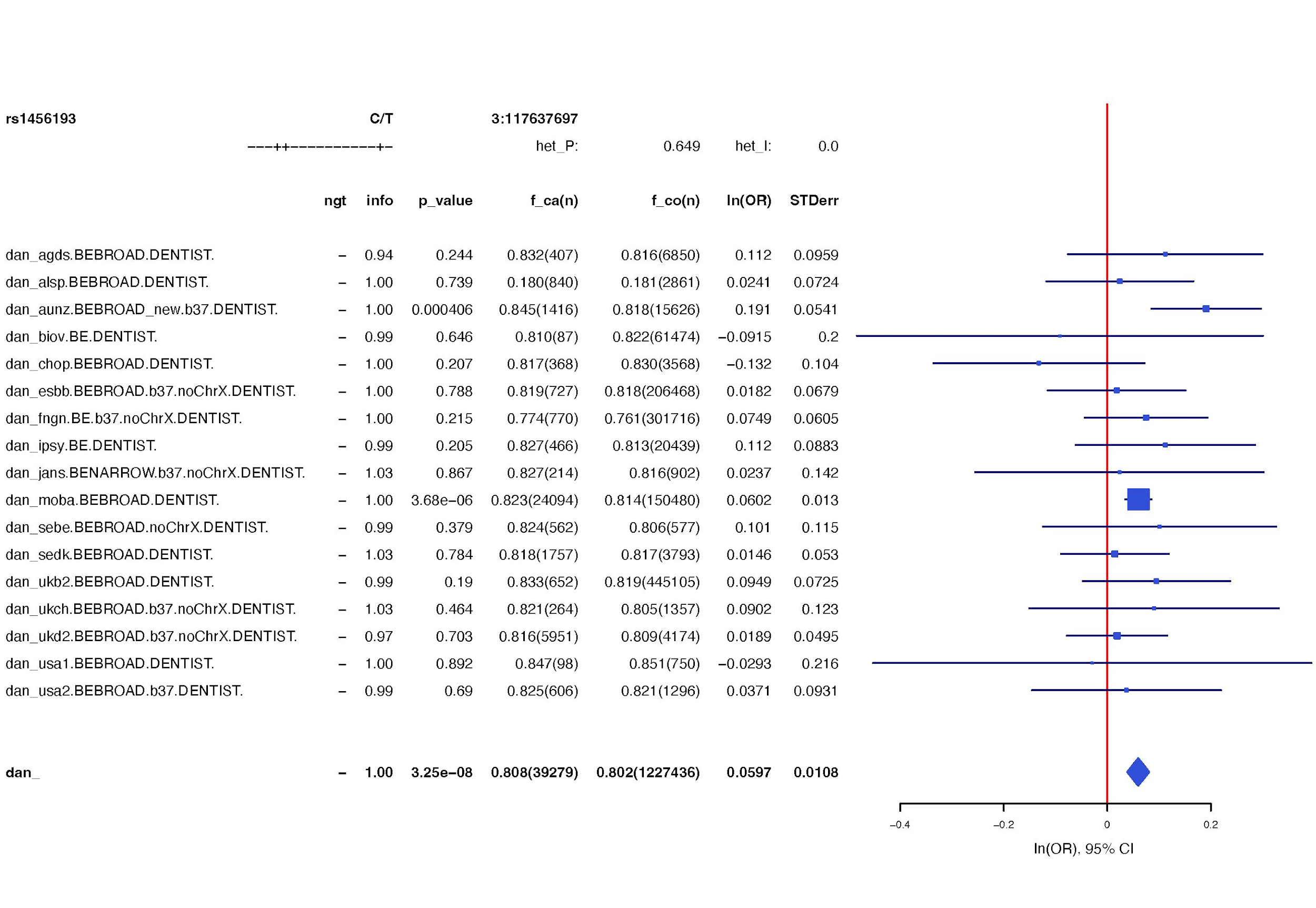

#### Supplementary Figure 3

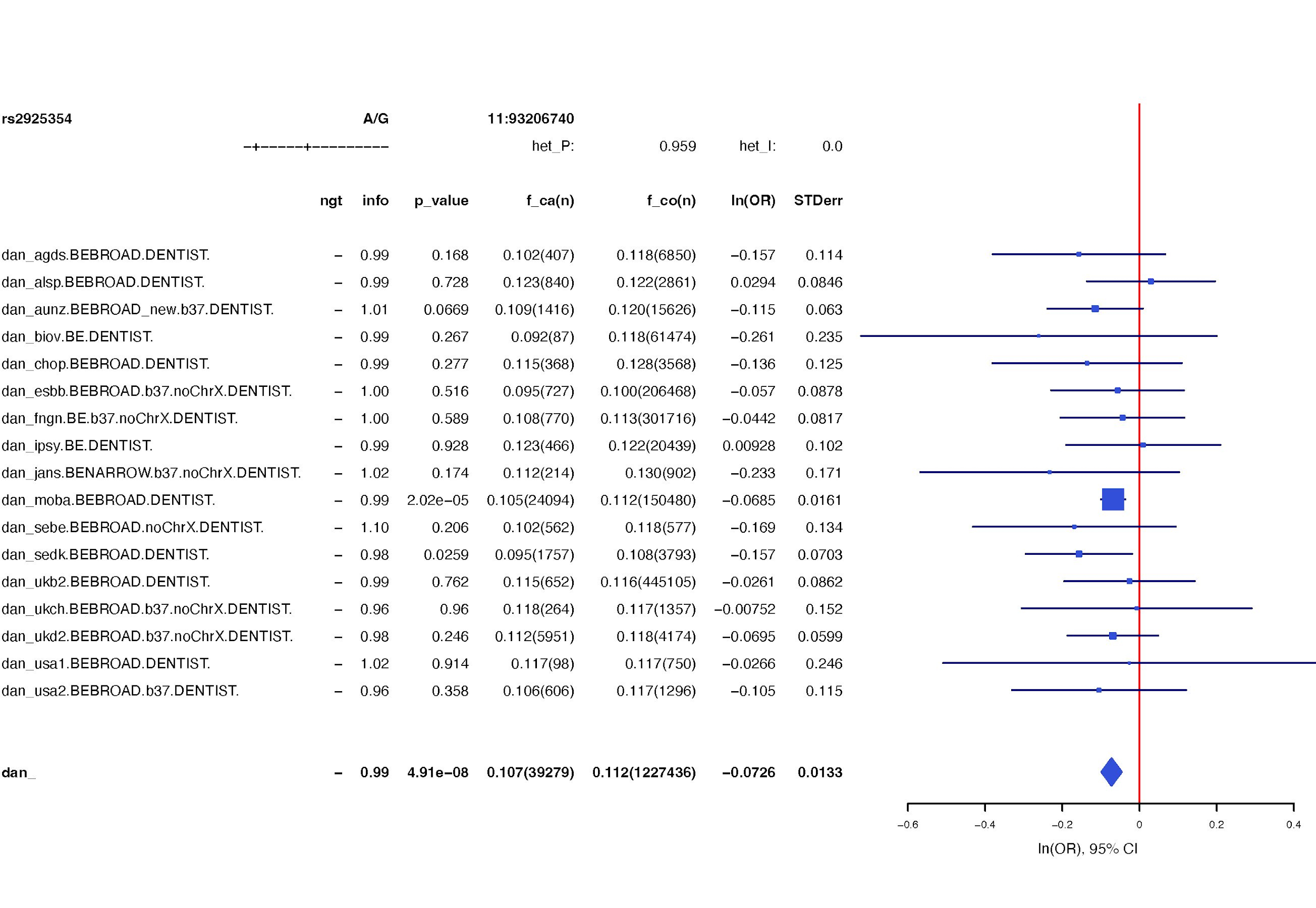

#### Supplementary Figure 4

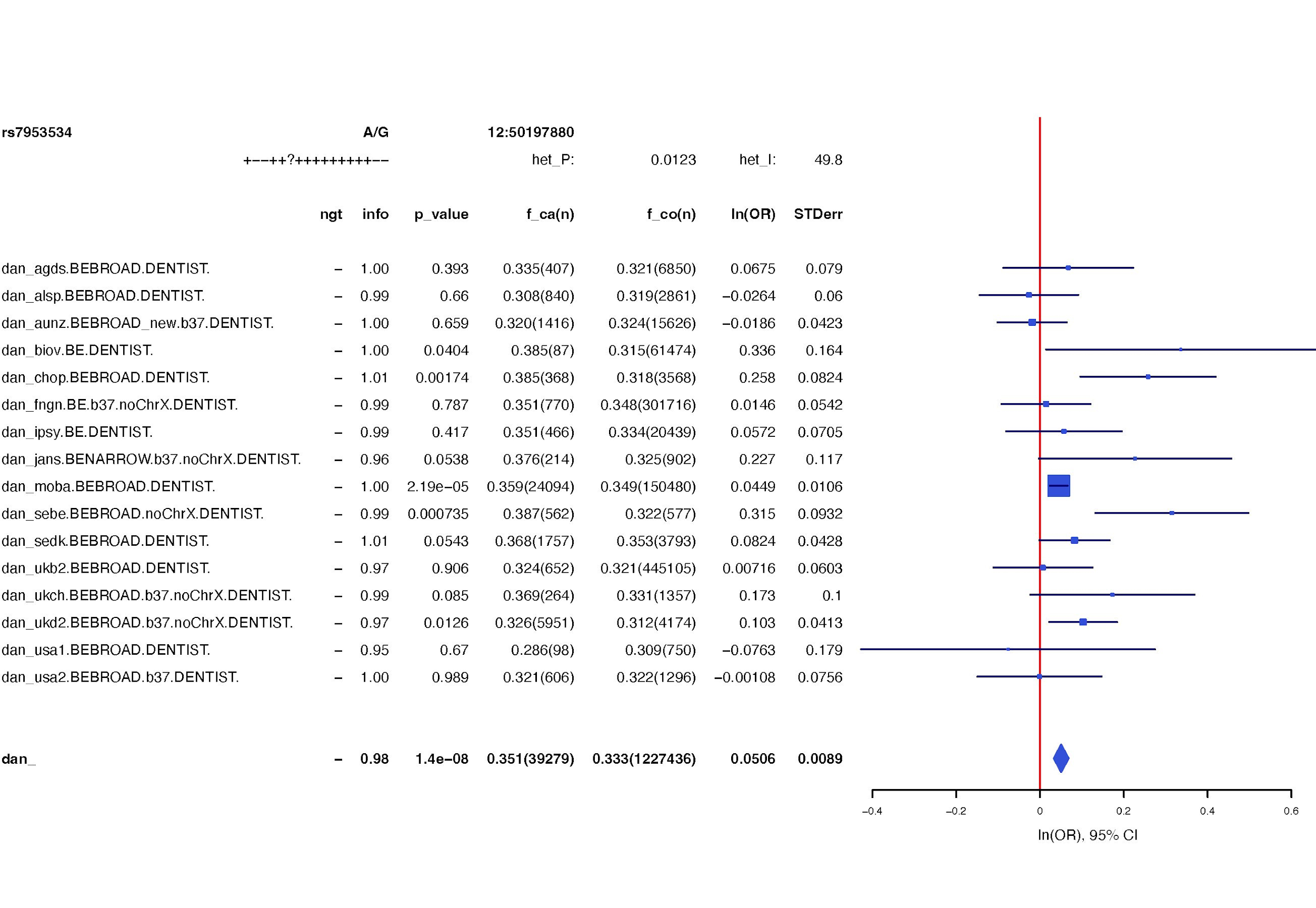

#### Supplementary Figure 5

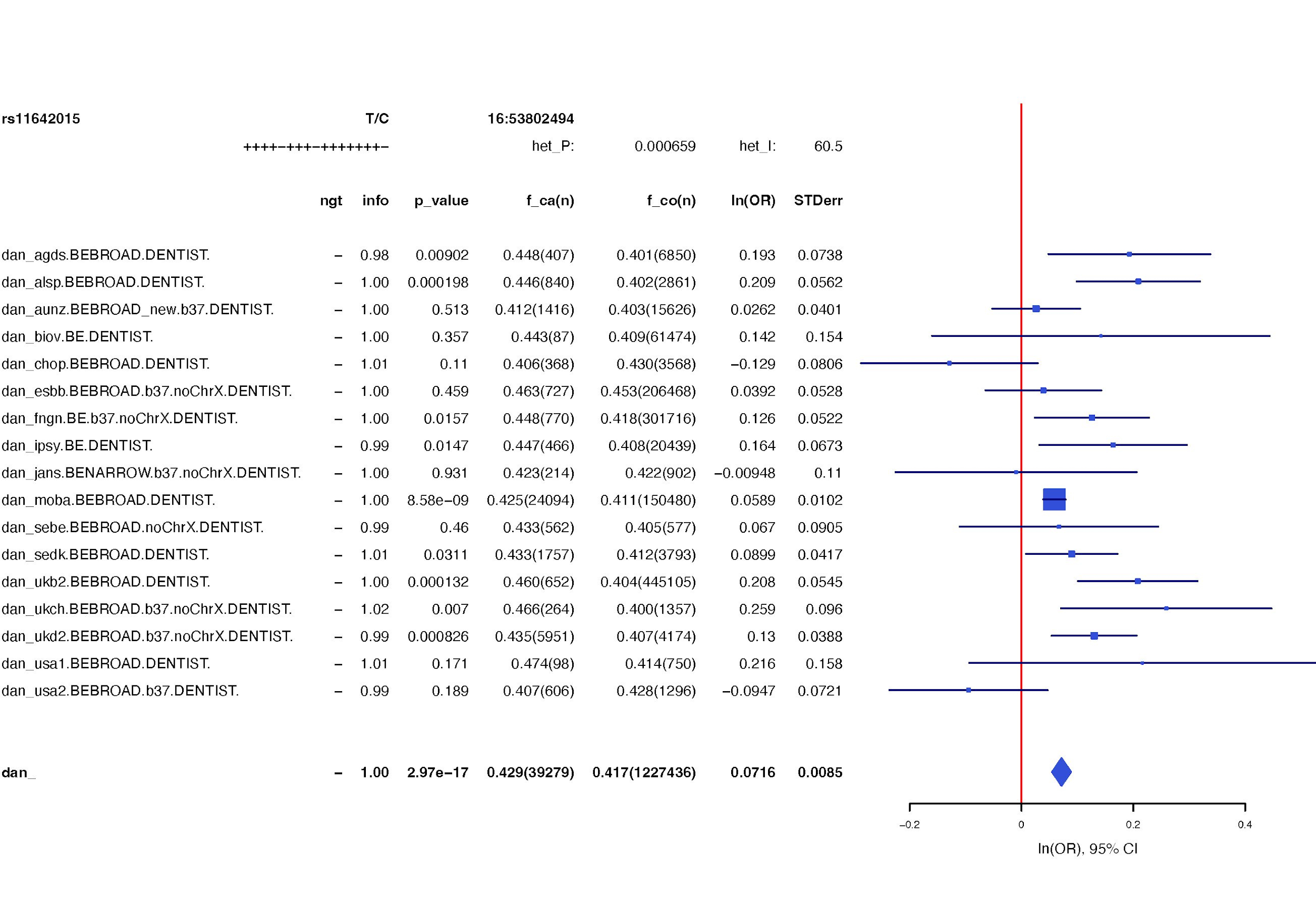

#### Supplementary Figure 6

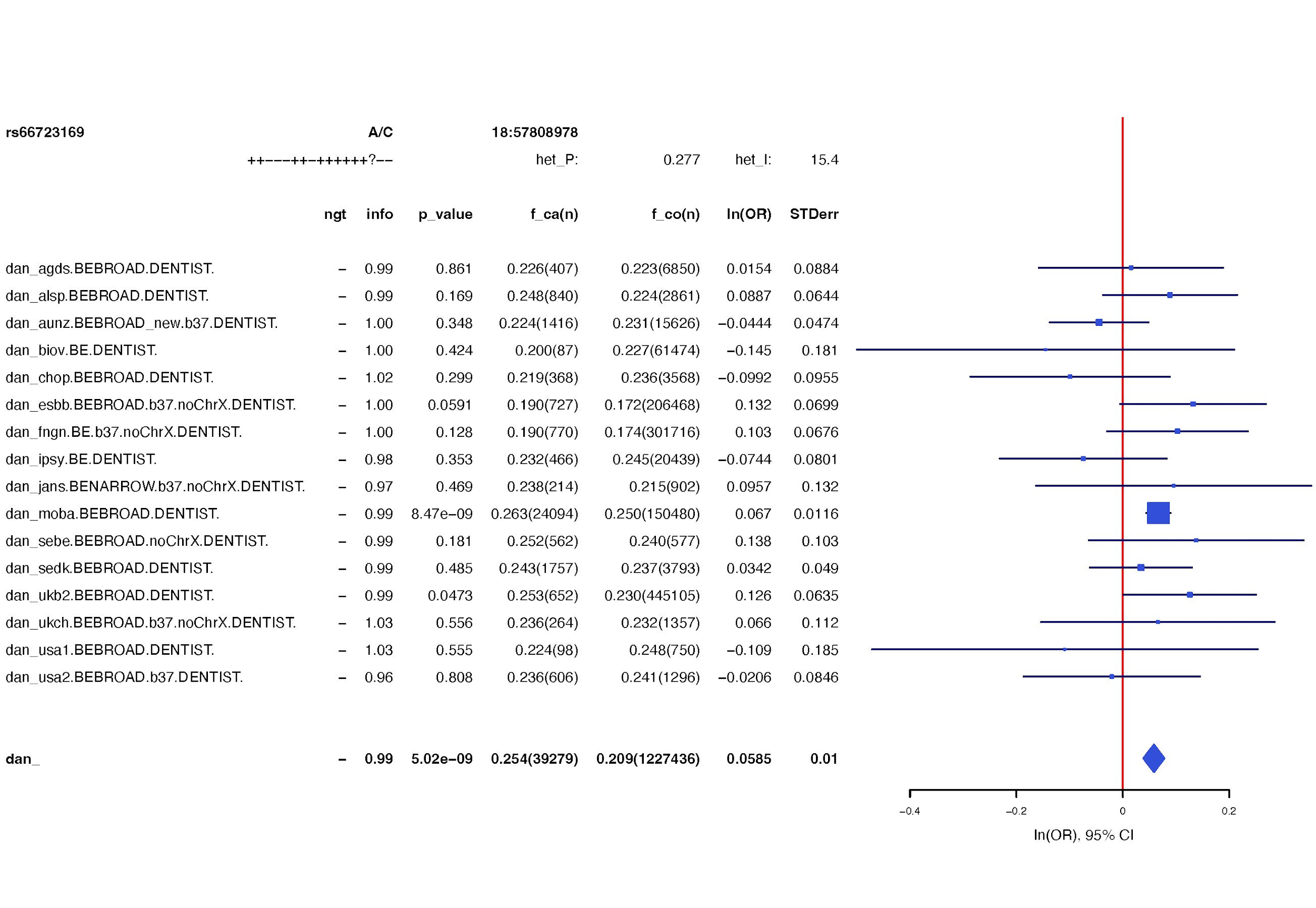

### Supplementary Figures 7-14: Forest plots for the eight genome-wide significant associations with AN in order of position on the genome.

Each row represents one cohort, with the final row showing the meta-analytic result. Initial information gives the variant name, effect and non-effect alleles, and variant position, followed by a summary of directions of effect, heterogeneity p-value, and heterogeneity I statistic. Main data columns include the imputation information metric (info), the association p-value, the frequency of the variant in cases (f_ca) and in controls (f_co), and the log odds ratio and standard error of the association, which are also visualised in the plot. The genotyping indicator (ngt) was set to missing for all cohorts and should be ignored.

#### Supplementary Figure 7

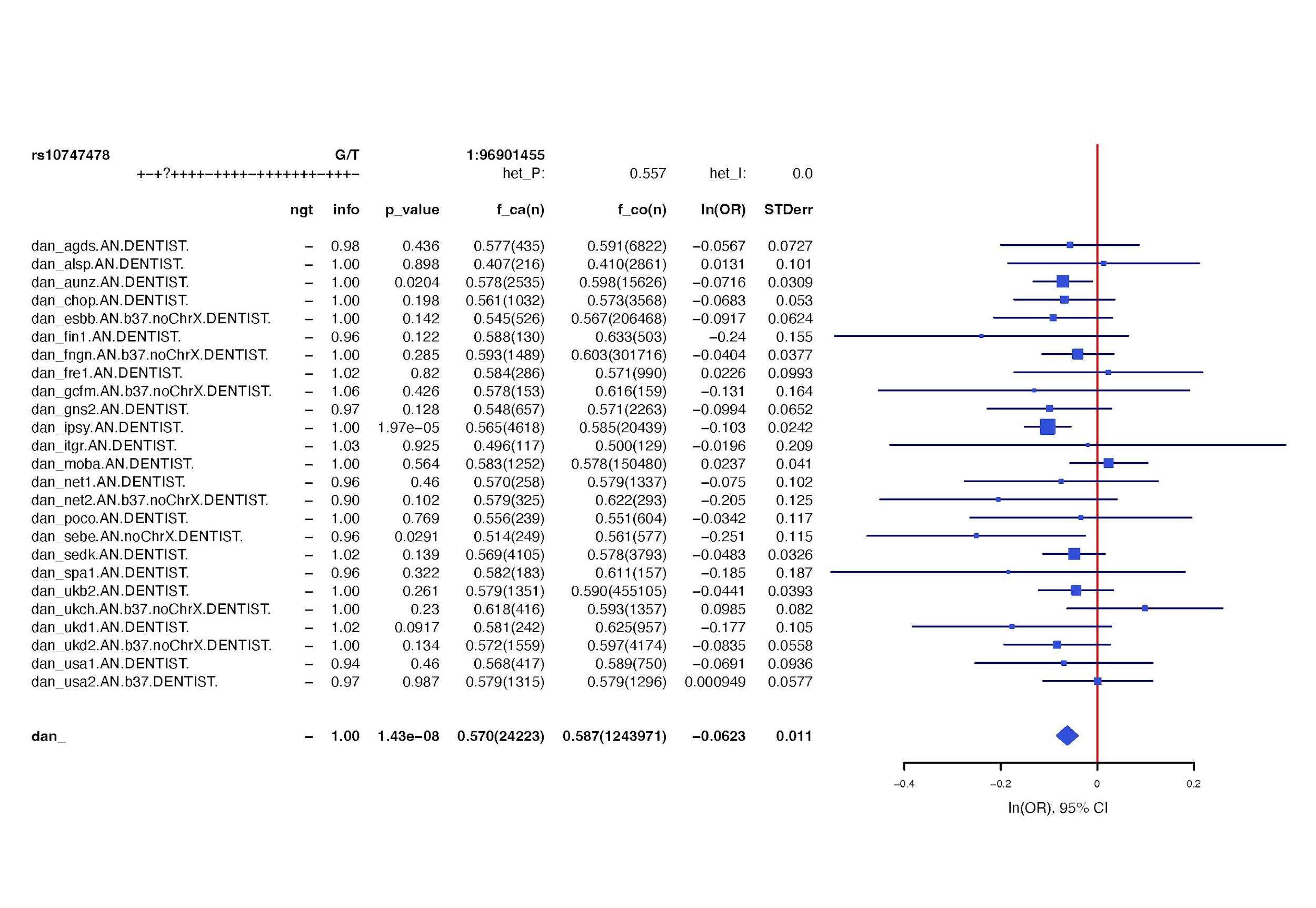

#### Supplementary Figure 8

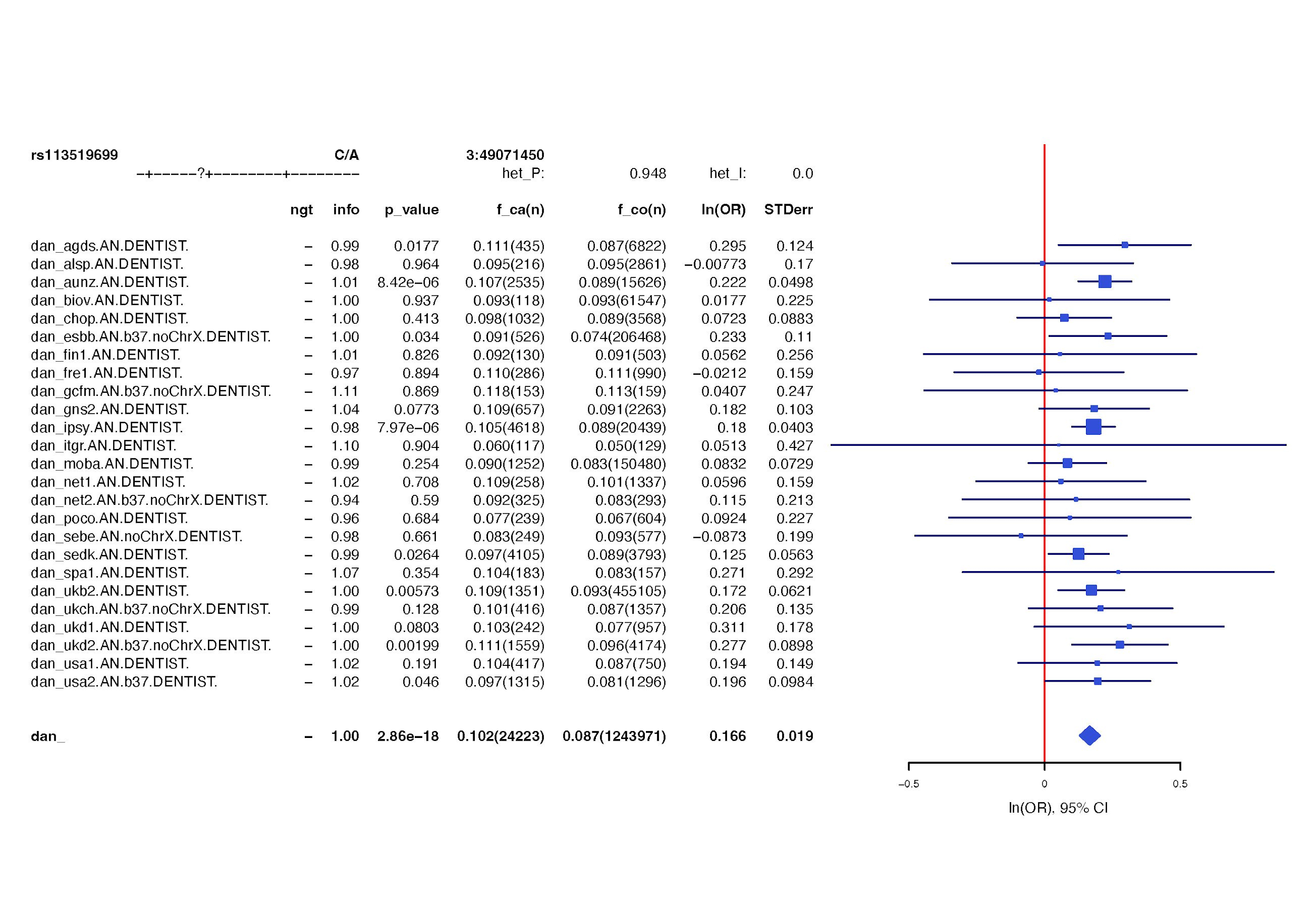

#### Supplementary Figure 9

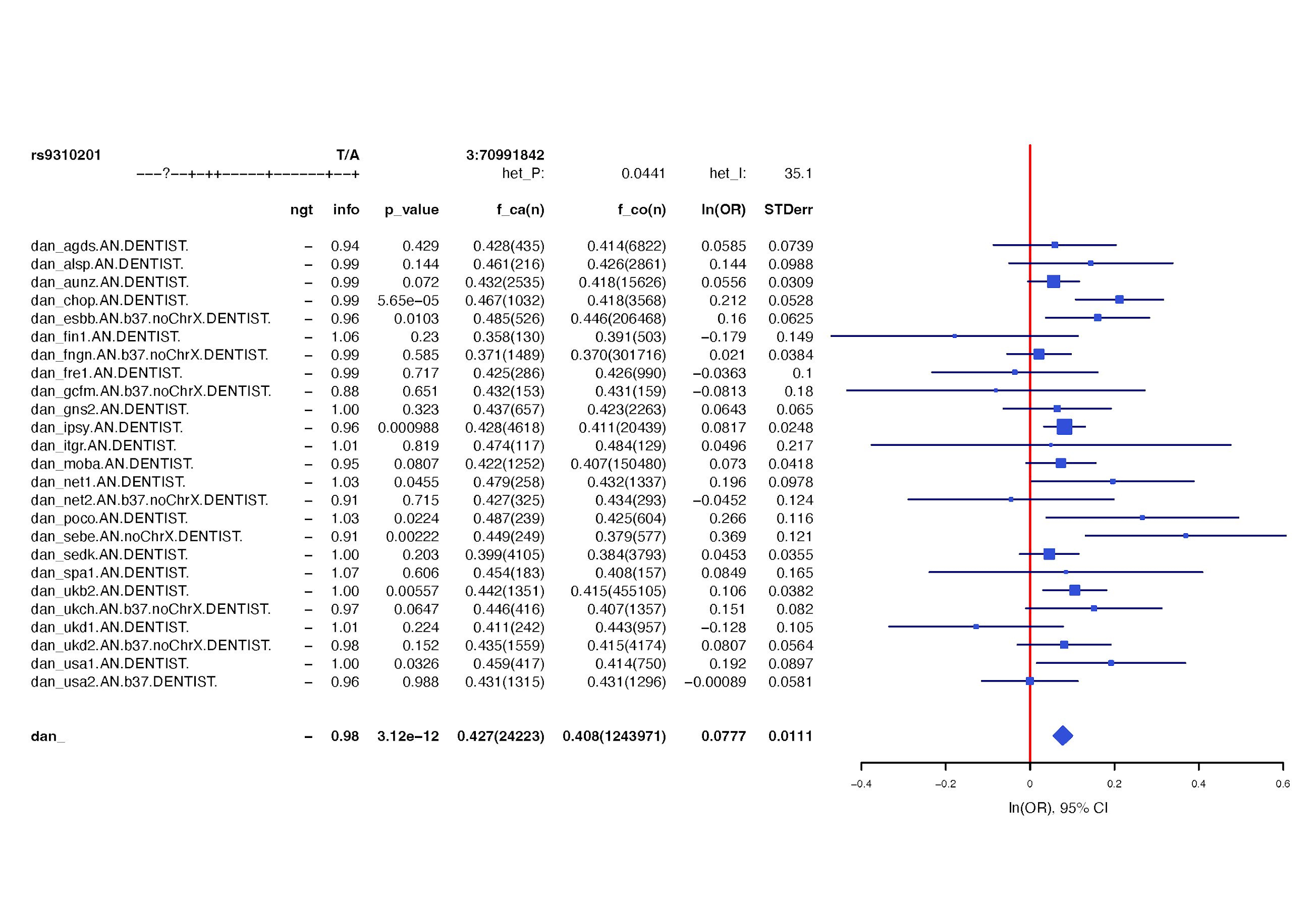

#### Supplementary Figure 10

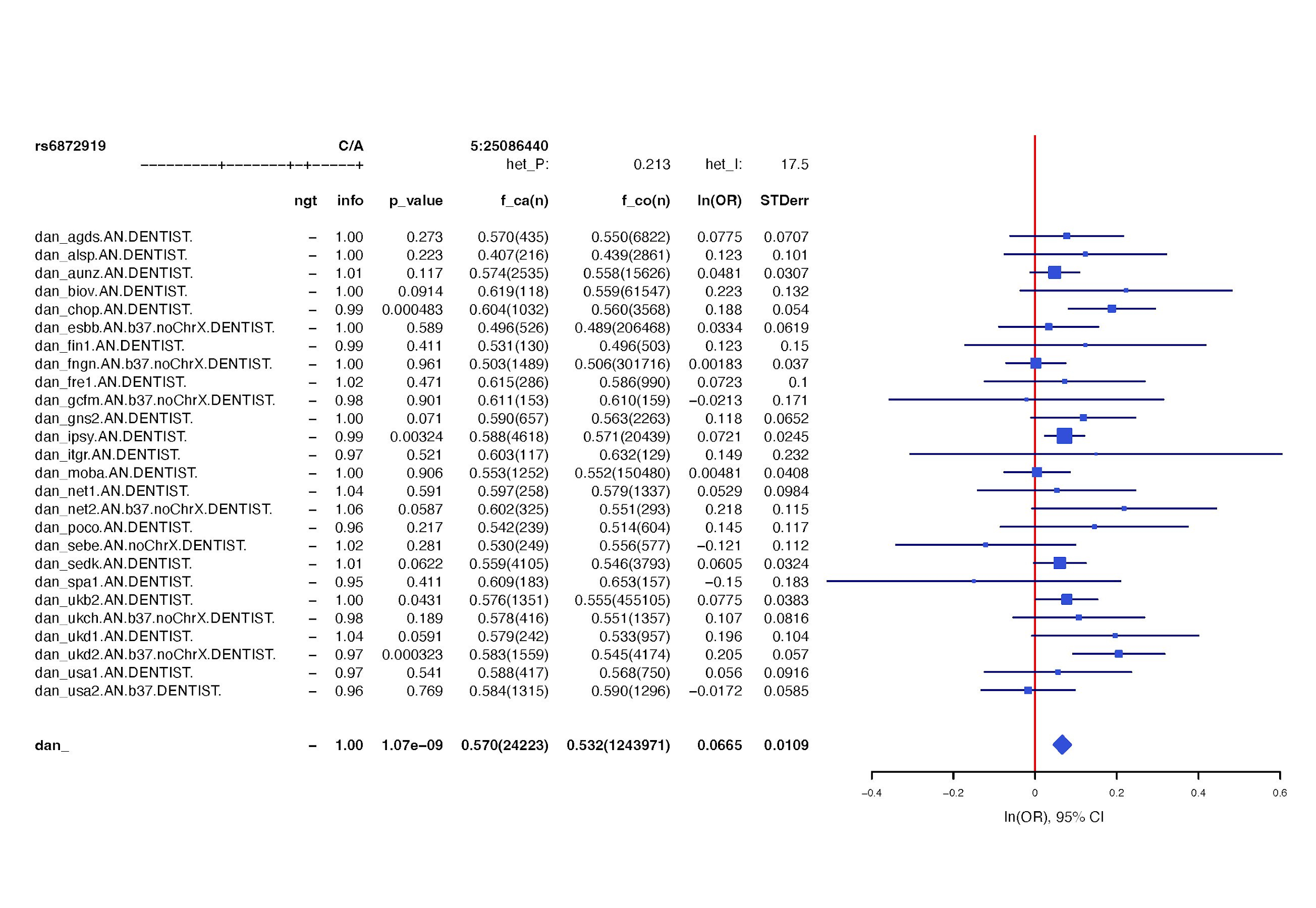

#### Supplementary Figure 11

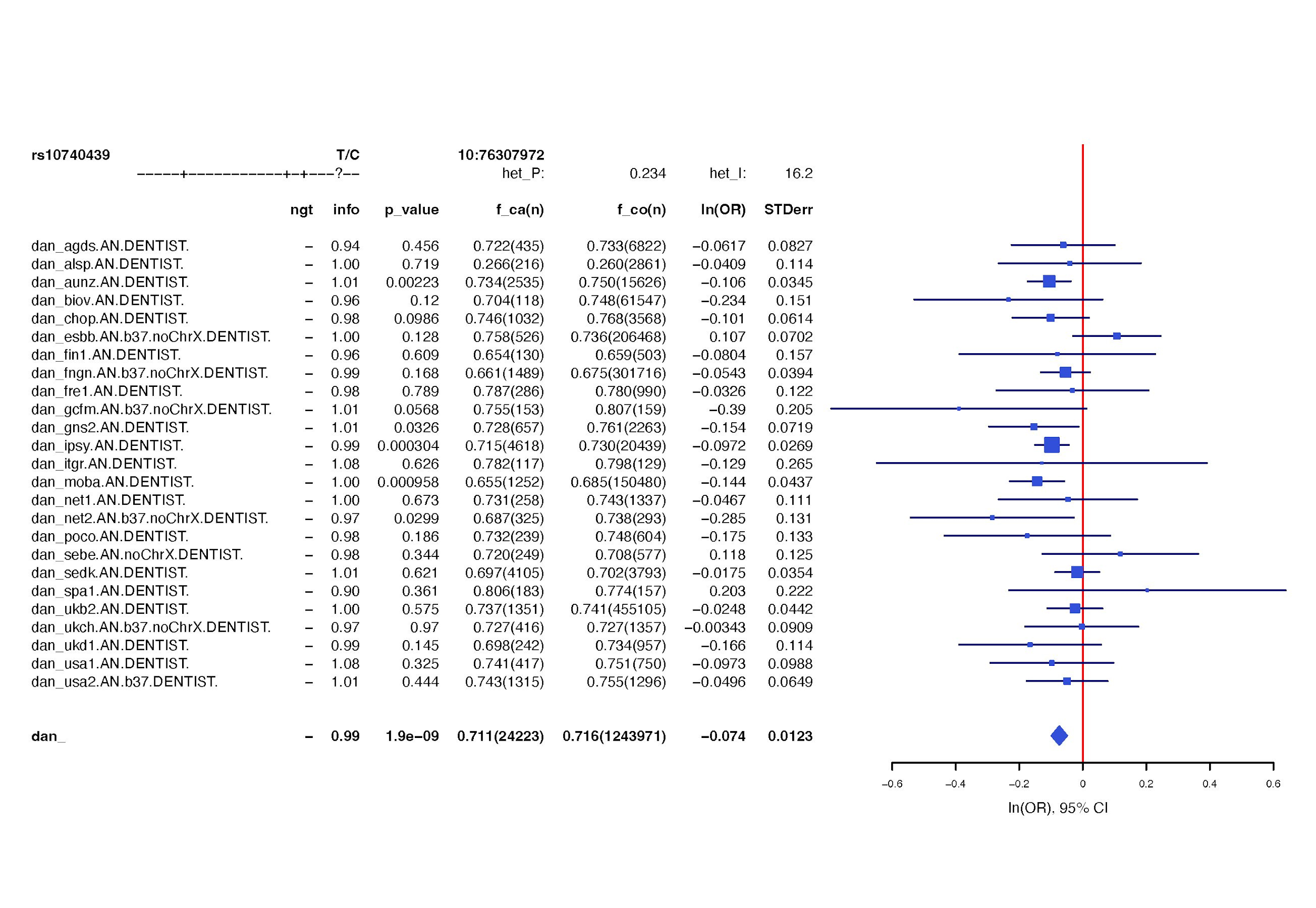

#### Supplementary Figure 12

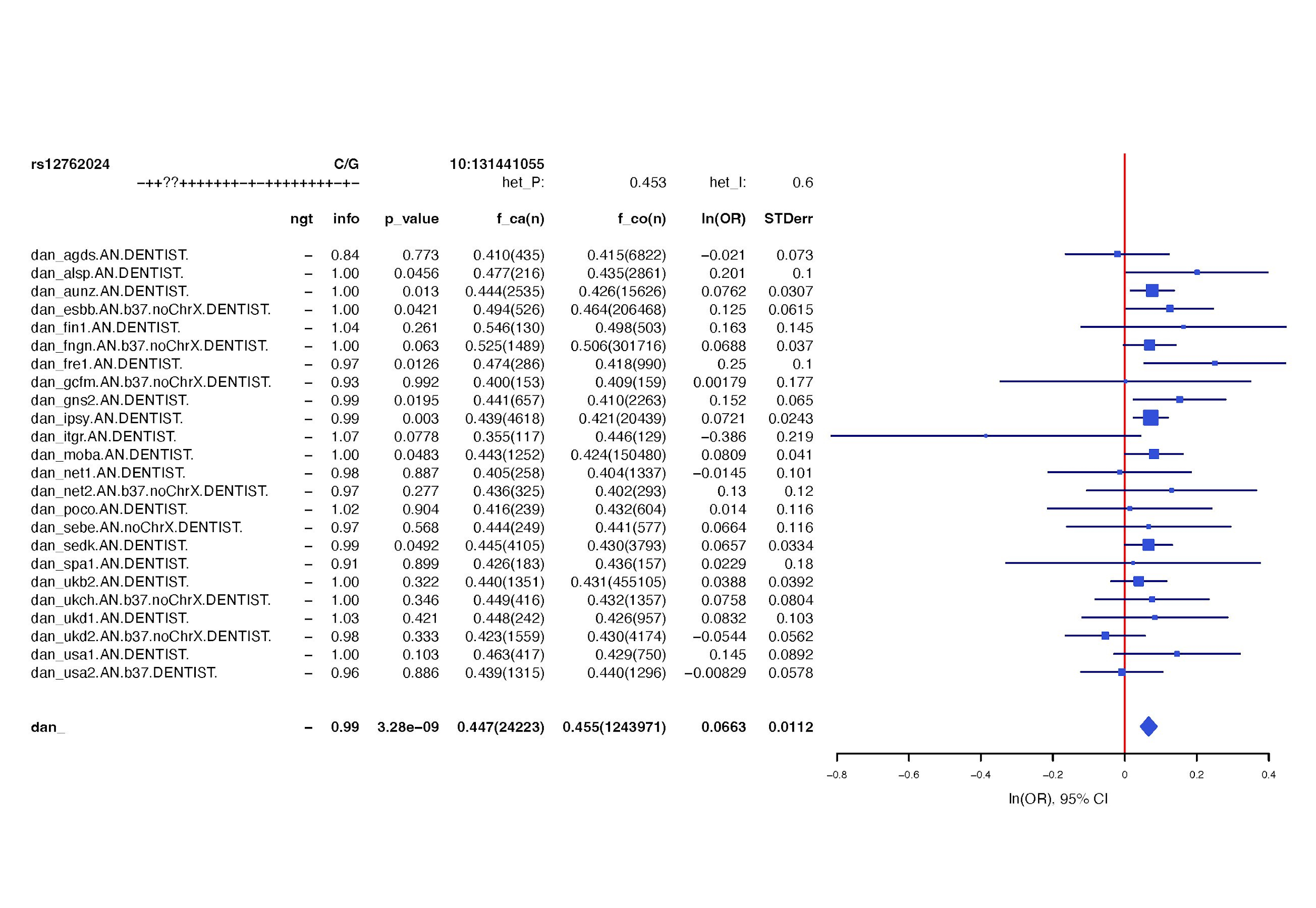

#### Supplementary Figure 13

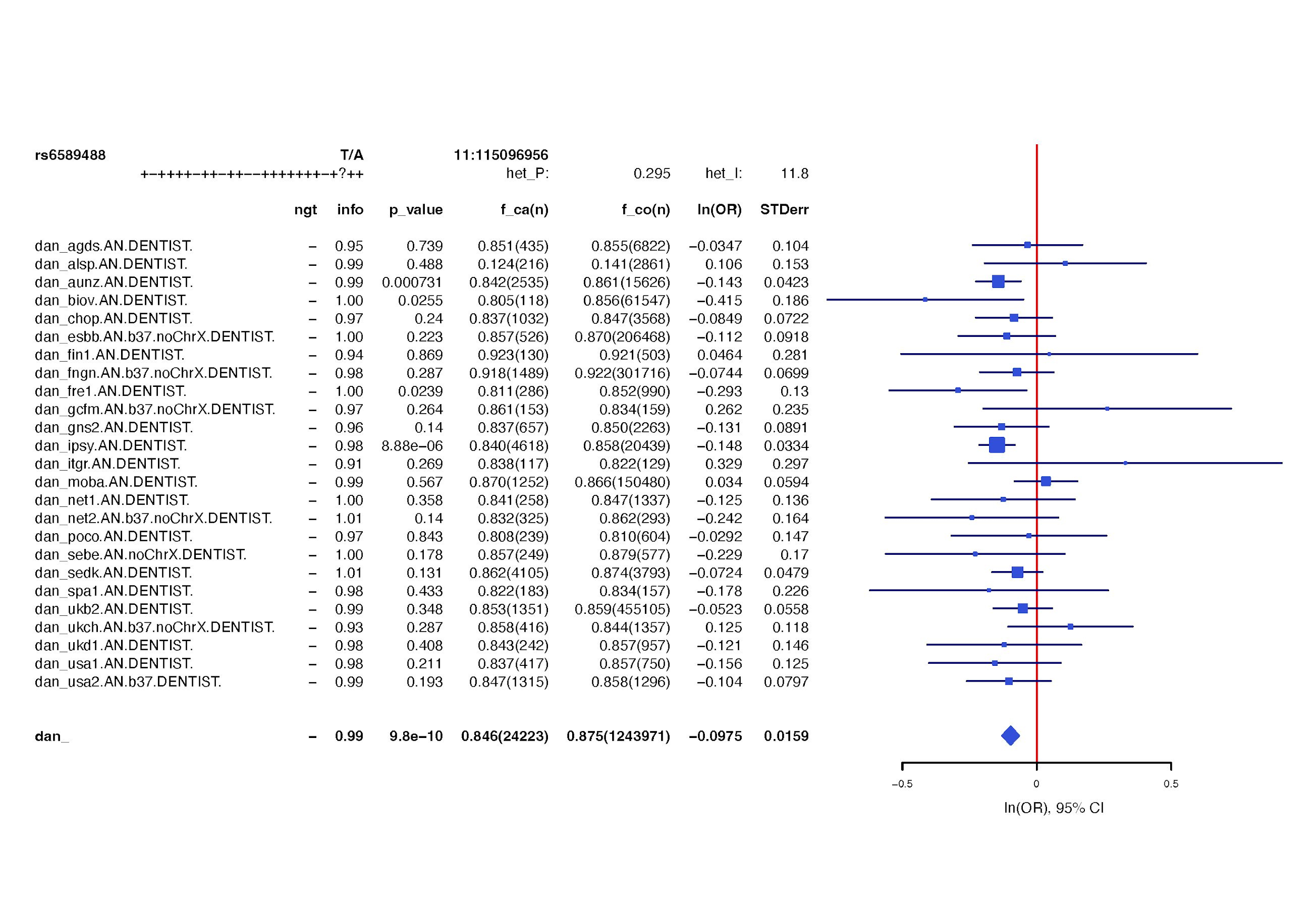

#### Supplementary Figure 14

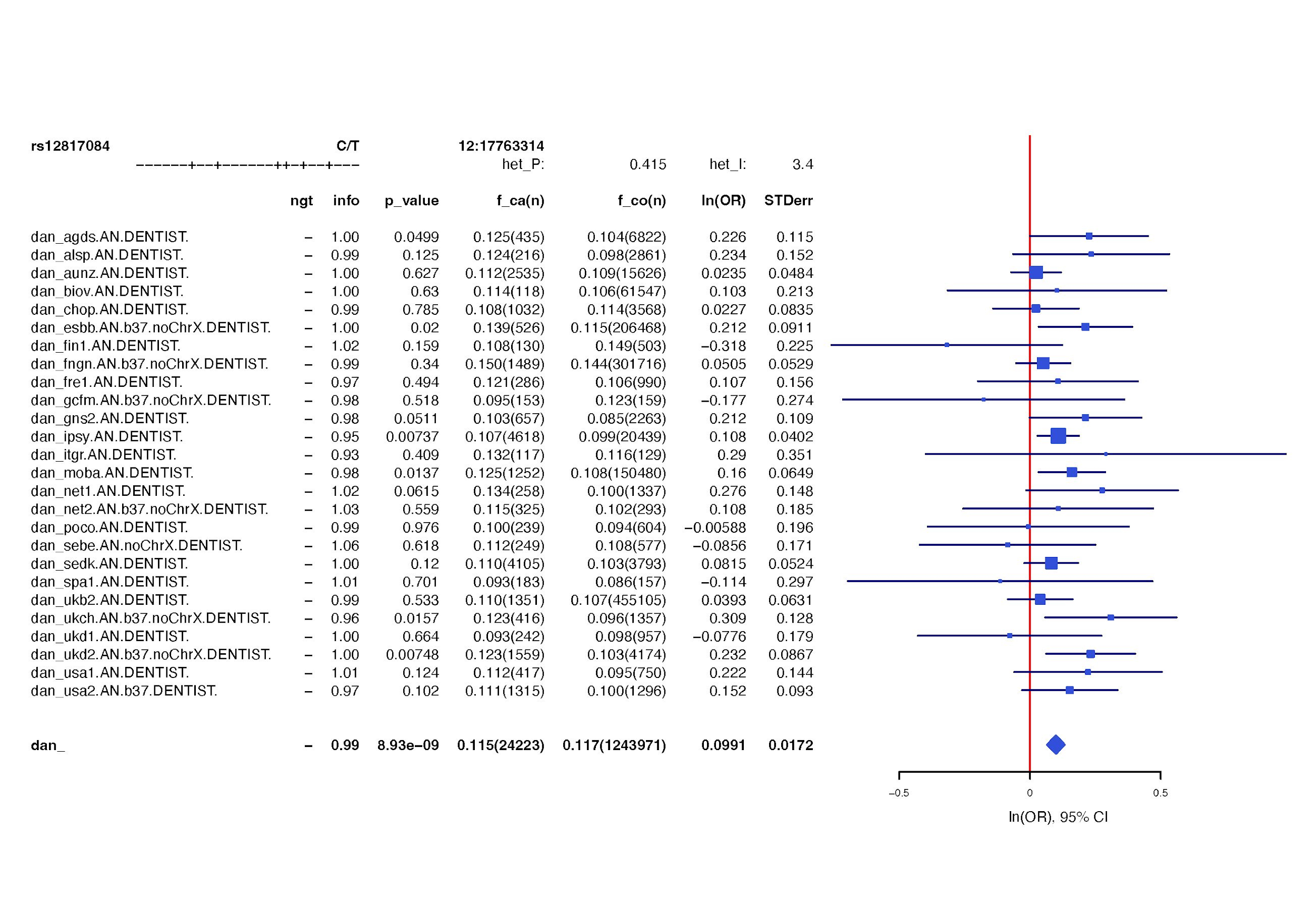

### Supplementary Figure 15: Manhattan plots for (a) BE-BROAD, (b) BE-NARROW, (c) AN, (d) AN-R, (e) AN-BP

The dotted red line is the genome-wide significance threshold (*P*≤5x10^-8^).

#### (a) BE-BROAD

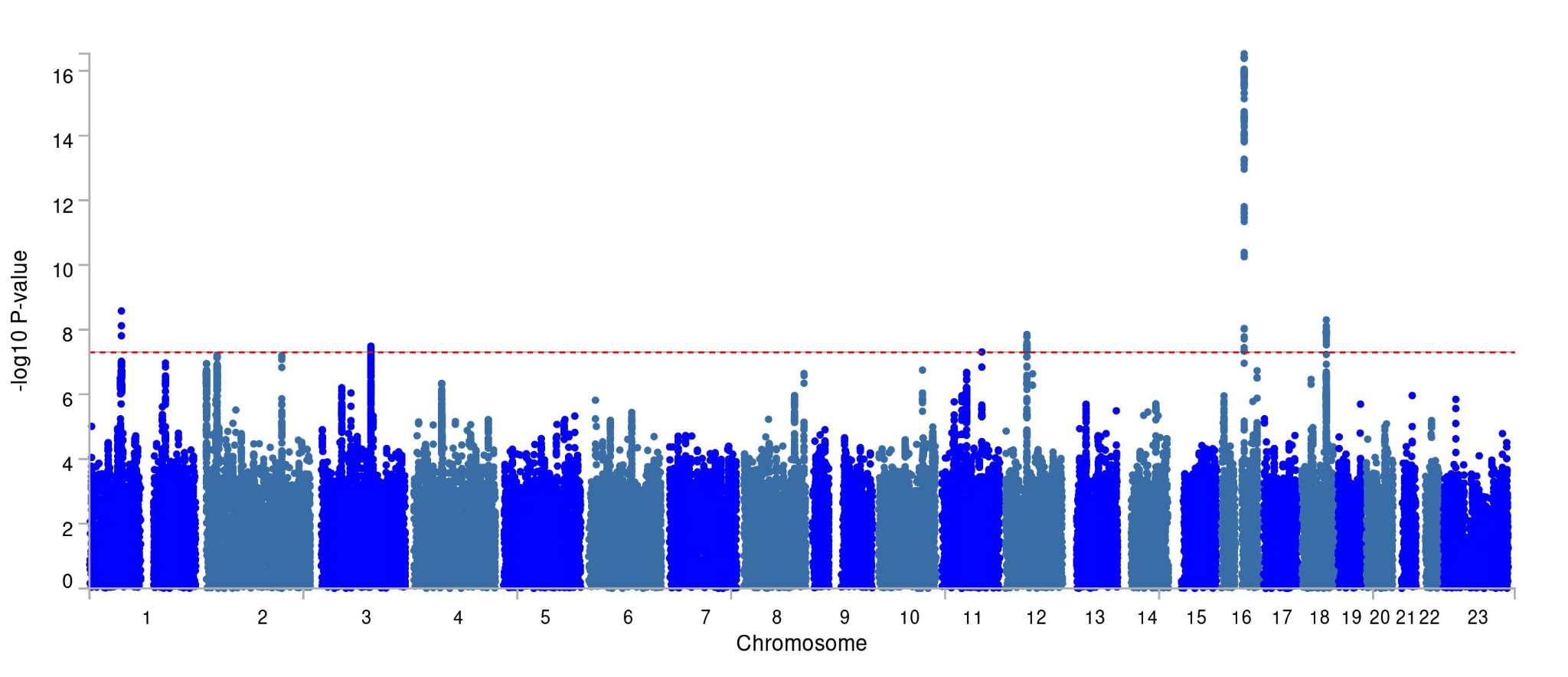

### (b) AN

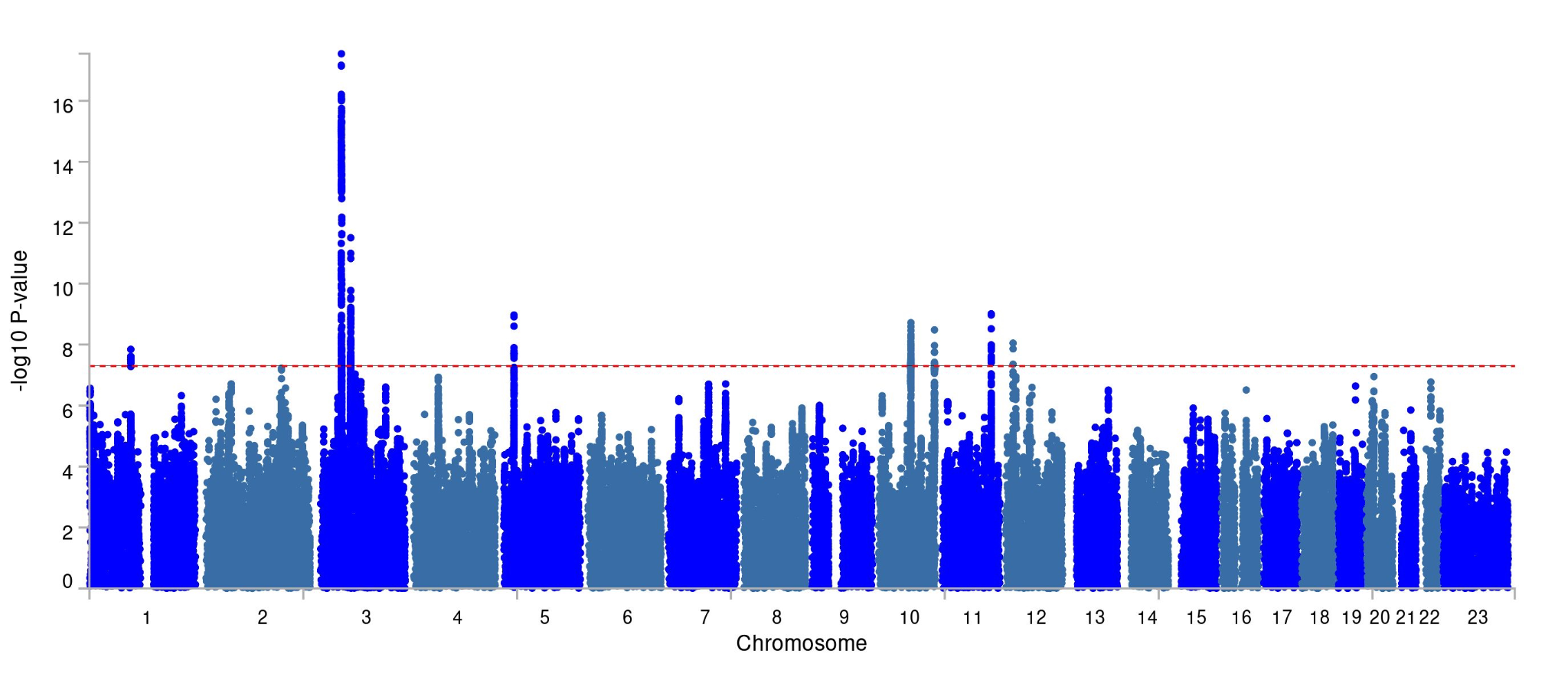

#### (c) BE-NARROW

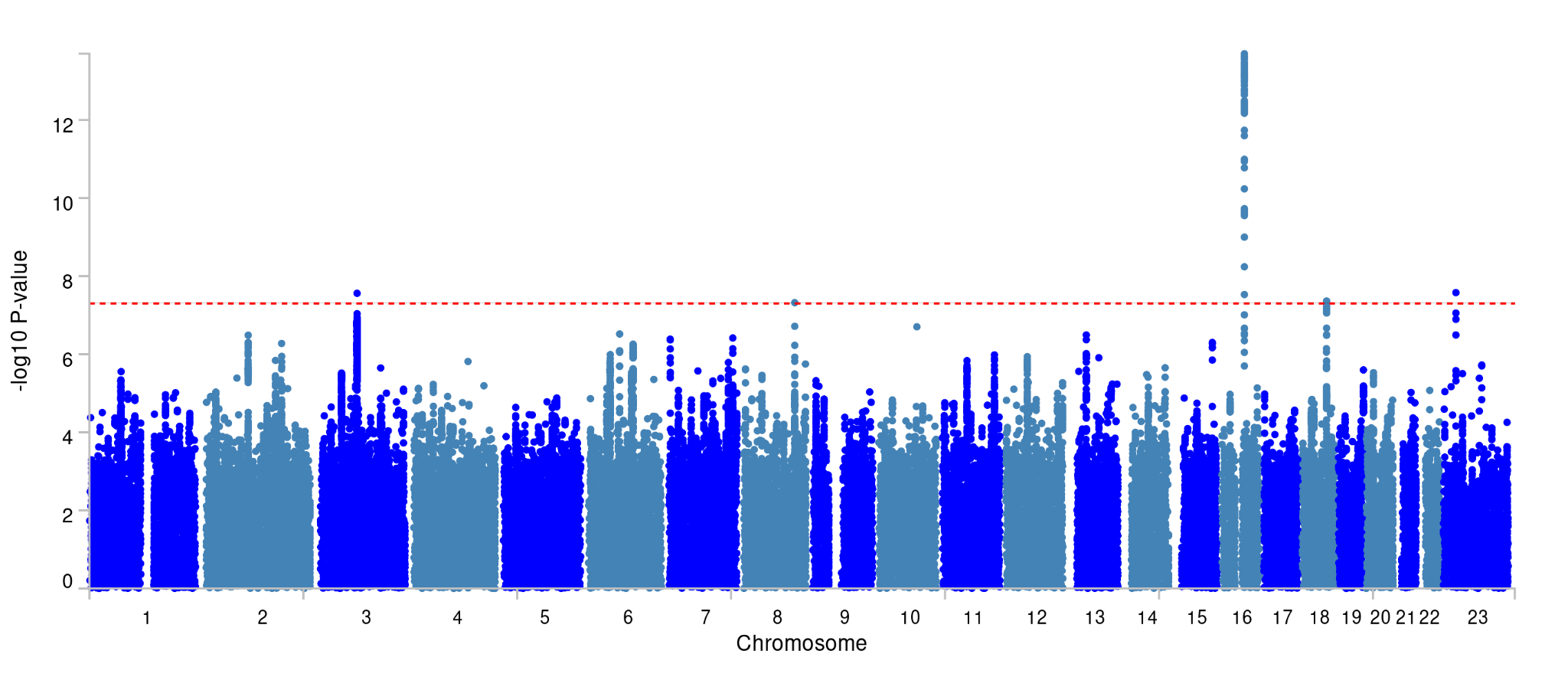

### (d) AN-R

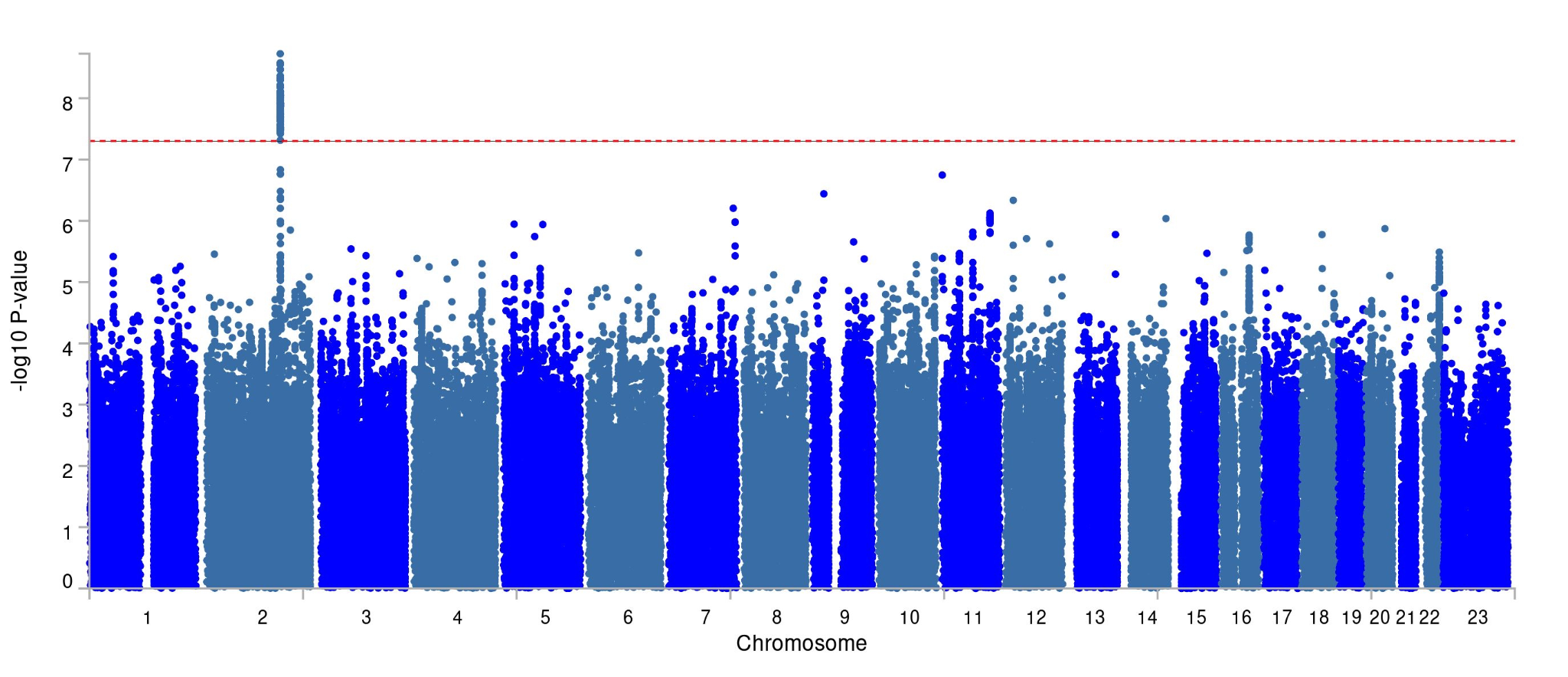

### (e) AN-BP

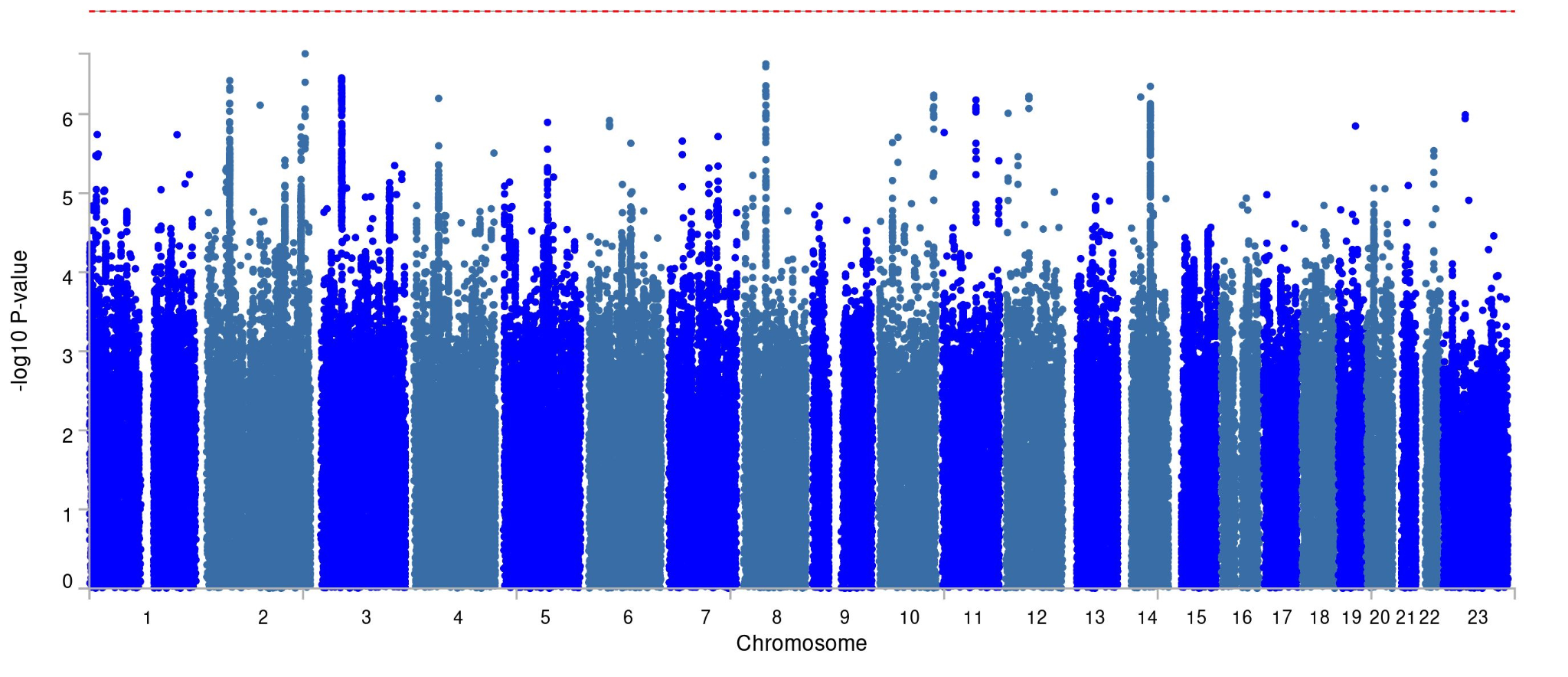

### Supplementary Figure 16: Significant differences between the genetic correlations (r_g_) of external traits with BE-BROAD and AN.

Genetic correlations were computed by Linkage Disequilibrium Score Regression (LDSC). Dot = r_g_ estimate, lines = standard errors. Significance determined via the Bonferroni method. Information about the summary statistics used in our analysis can be found in Supplementary Table 8.

#### (a) Significant differences

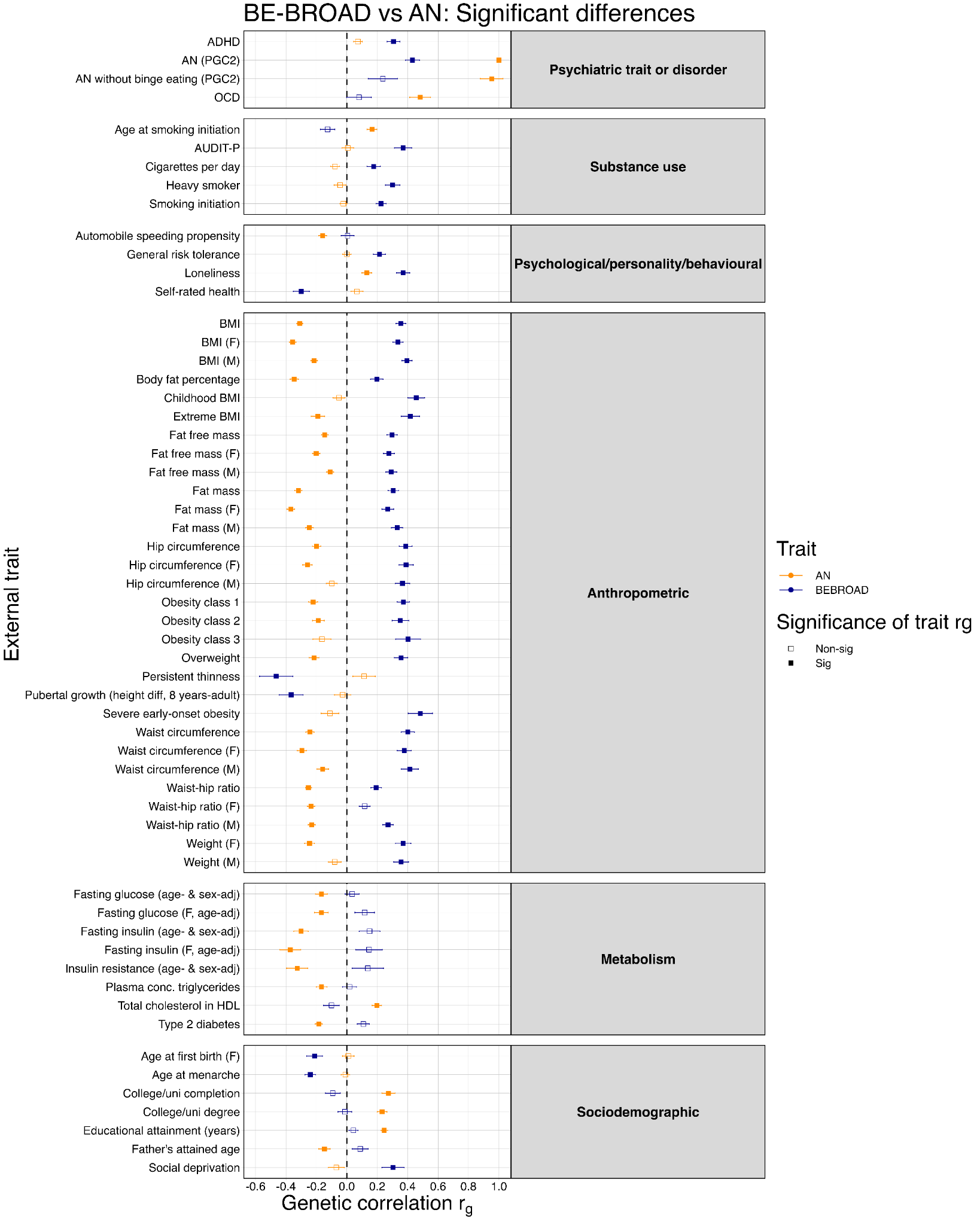

BE-BROAD = binge-eating broad definition; AN = anorexia nervosa; ADHD = attention deficit hyperactivity disorder; PGC2 = Psychiatric Genomics Consortium Freeze 2; OCD = obsessive compulsive disorder; AUDIT-P = Alcohol Use Disorder Identification Test problem items; BMI = body mass index; F = female; M = male; HDL = high-density lipoprotein

#### (b) Non-significant differences

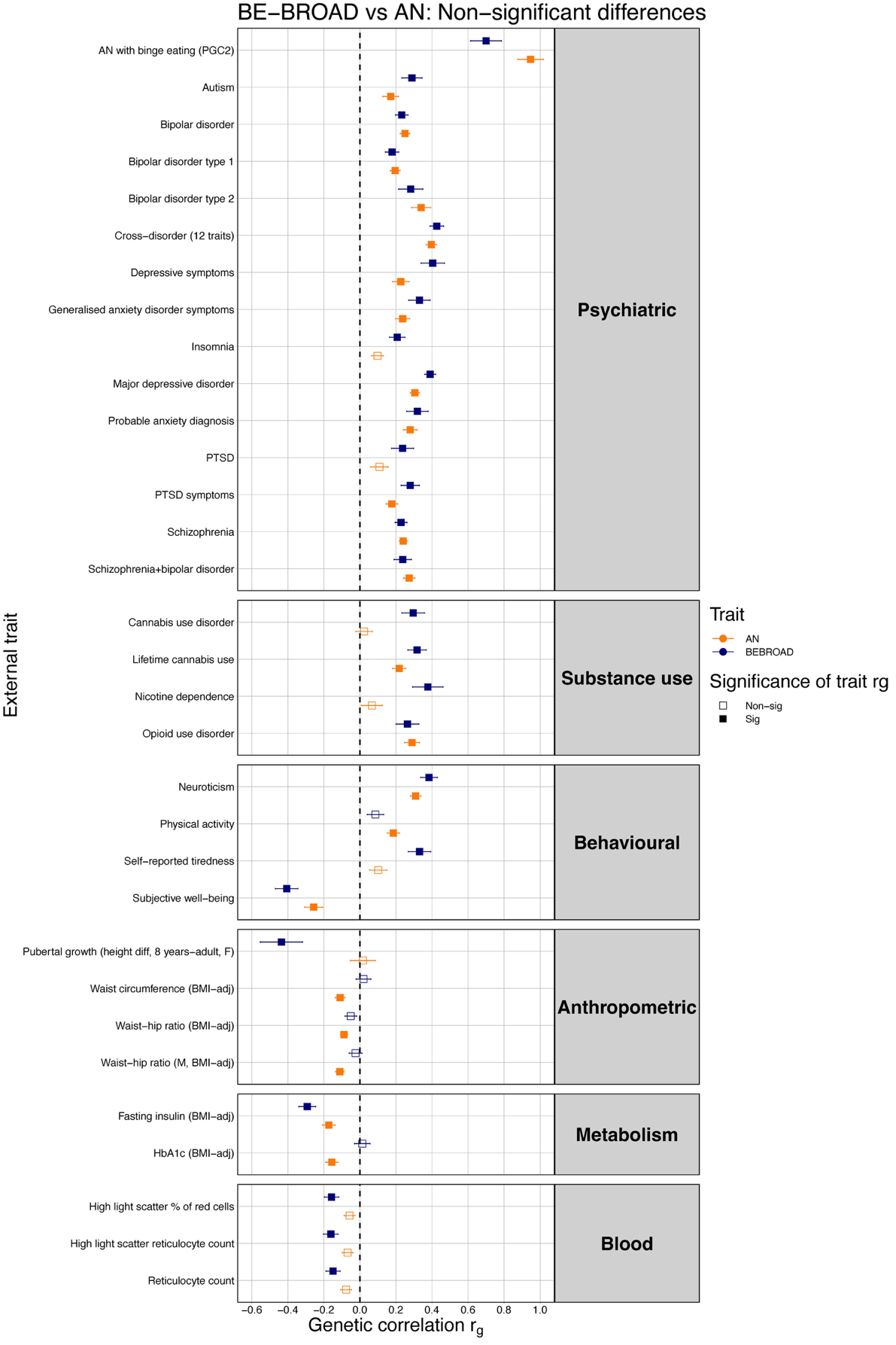

BE-BROAD = binge-eating broad definition; AN = anorexia nervosa; PGC = Psychiatric Genomics Consortium; PTSD = post-traumatic stress disorder; BMI-adj = adjusted for body mass index

### Supplementary Figure 17: Comparison of the genetic correlations (r_g_) of selected external traits with not-ascertained-for-AN BE-BROAD (BE-BROAD_NAAN_) and BE-BROAD.

Genetic correlations were computed by Linkage Disequilibrium Score Regression (LDSC). The rg estimation is indicated by the dot and standard errors are indicated by the lines on either side of each dot. rg estimates have been corrected for multiple testing via the Bonferroni method. Information about the summary statistics used in our analysis can be found in Supplementary Table 8.

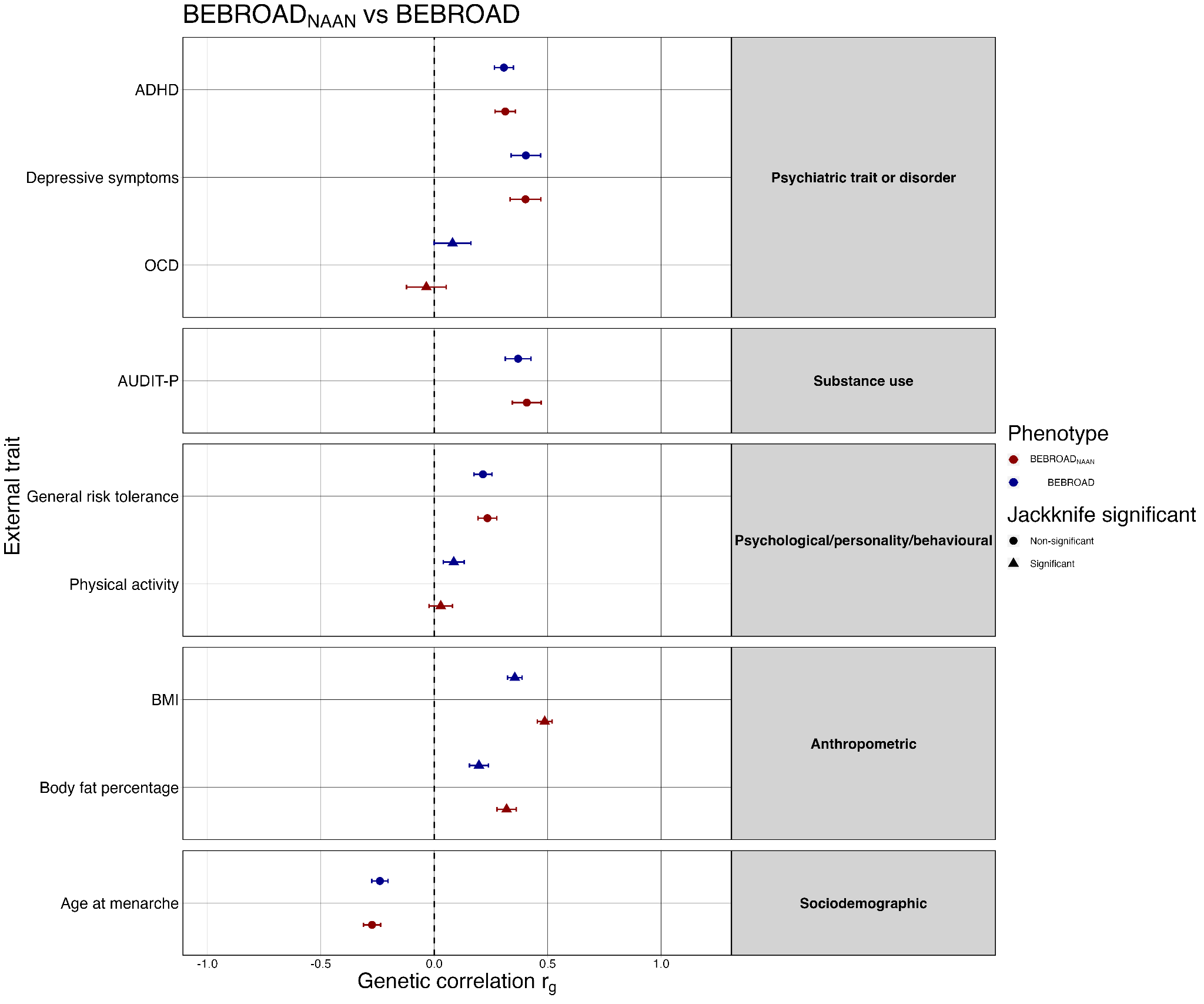

BE-BROAD_NAAN_ = BE-BROAD from cohorts not-ascertained-for-AN; BE-BROAD = binge-eating broad definition; AN = anorexia nervosa; ADHD = attention deficit hyperactivity disorder; OCD = obsessive compulsive disorder; AUDIT-P = Alcohol Use Disorder Identification Test problem items; BMI = body mass index

### Supplementary Figure 18: Comparison of the genetic correlations of BE-BROAD (left) and AN (right) and of their respective NonBMI components with external traits.

Experiment-wide significance set at p < 2.22*10^-4^, consistent with p-value used for genetic correlations throughout the paper. Trait-level significance represented by darkened circles; jackknife significance represented by solid black connection between original and nonBMI trait. Traits tested noted on y-axis, trait categories indicated between BE-BROAD (left) and AN (right).

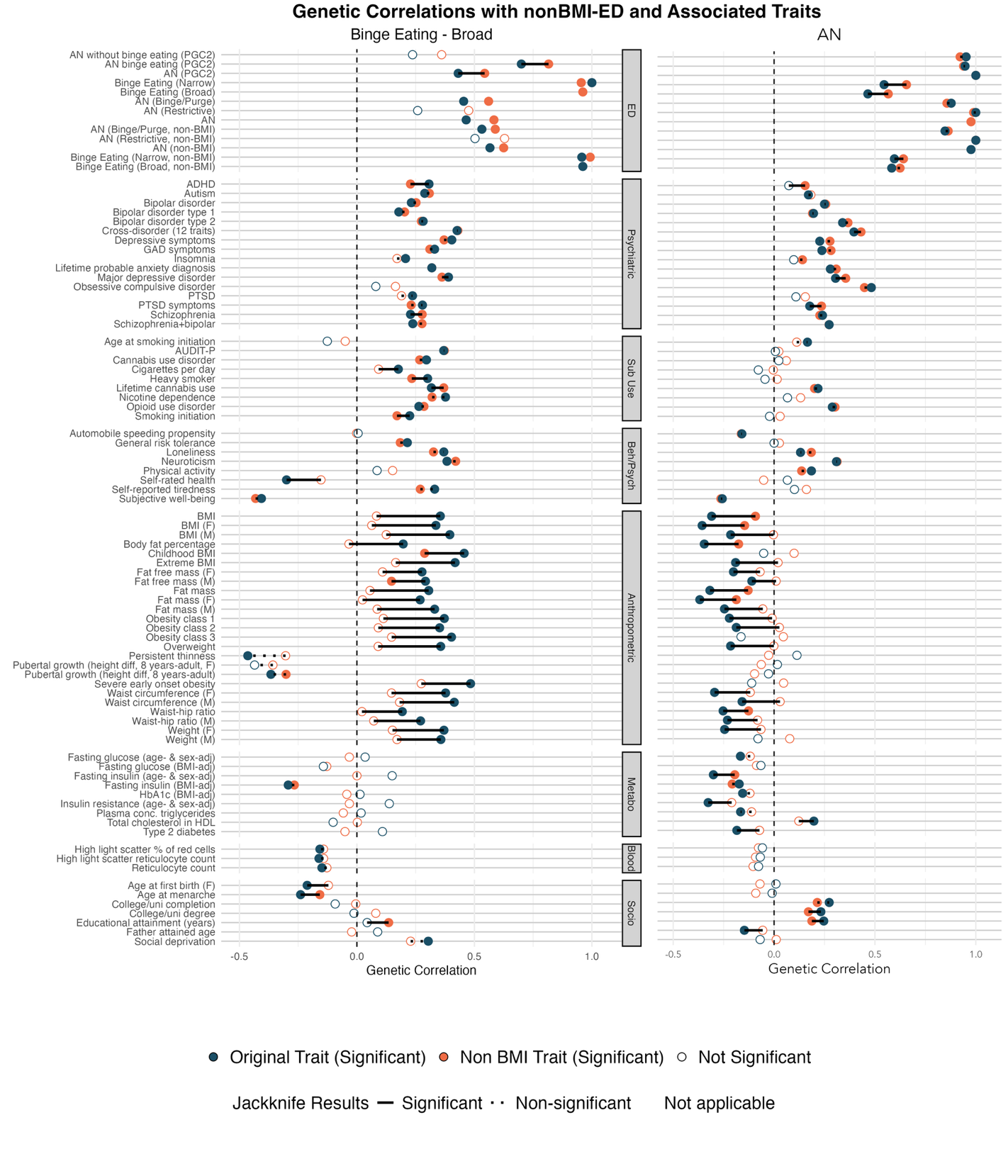

### Supplementary Figure 19: Gene-tissue associations from S-PrediXcan for (a) BE-BROAD (b) AN (c) BE-NARROW (d) ANBP (e) ANR.

Enrichment of gene-tissue associations using two-tailed exact binomial tests, testing for enrichment of associations at (1) nominal significance, p<0.05, (2) tissue-specific significance, p<0.05/Tests_TissueX_, and (3) experiment-wide significance, p<8.32 x 10^-8^. Manhattan plot of S-PrediXcan gene-tissue associations with annotations of top genes. Experiment-wide significance threshold of p<8.32 x 10^-8^ (dotted line); experiment-wide associations are highlighted in red; tissue-specific associations (p<0.05/Tests_TissueX_) are shown in other colours.

#### (a) BE-BROAD

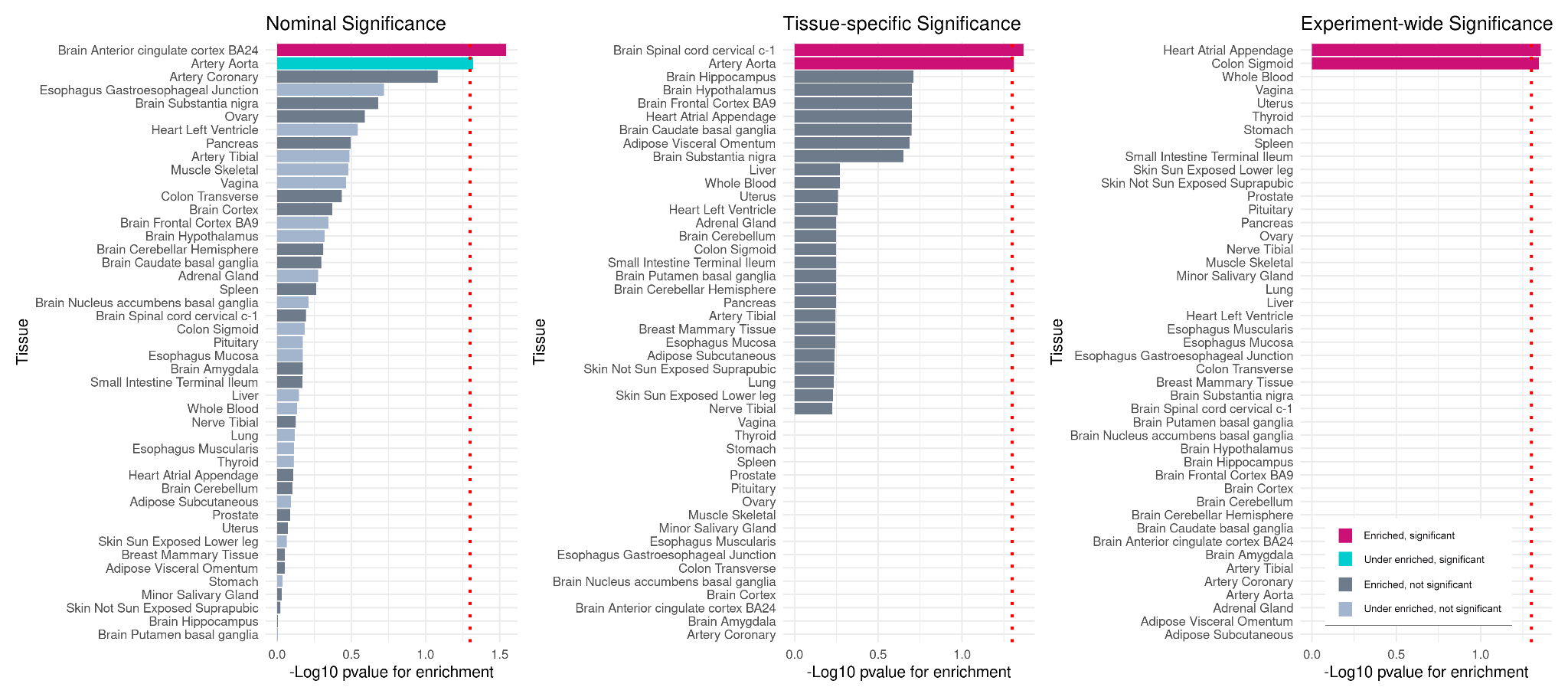

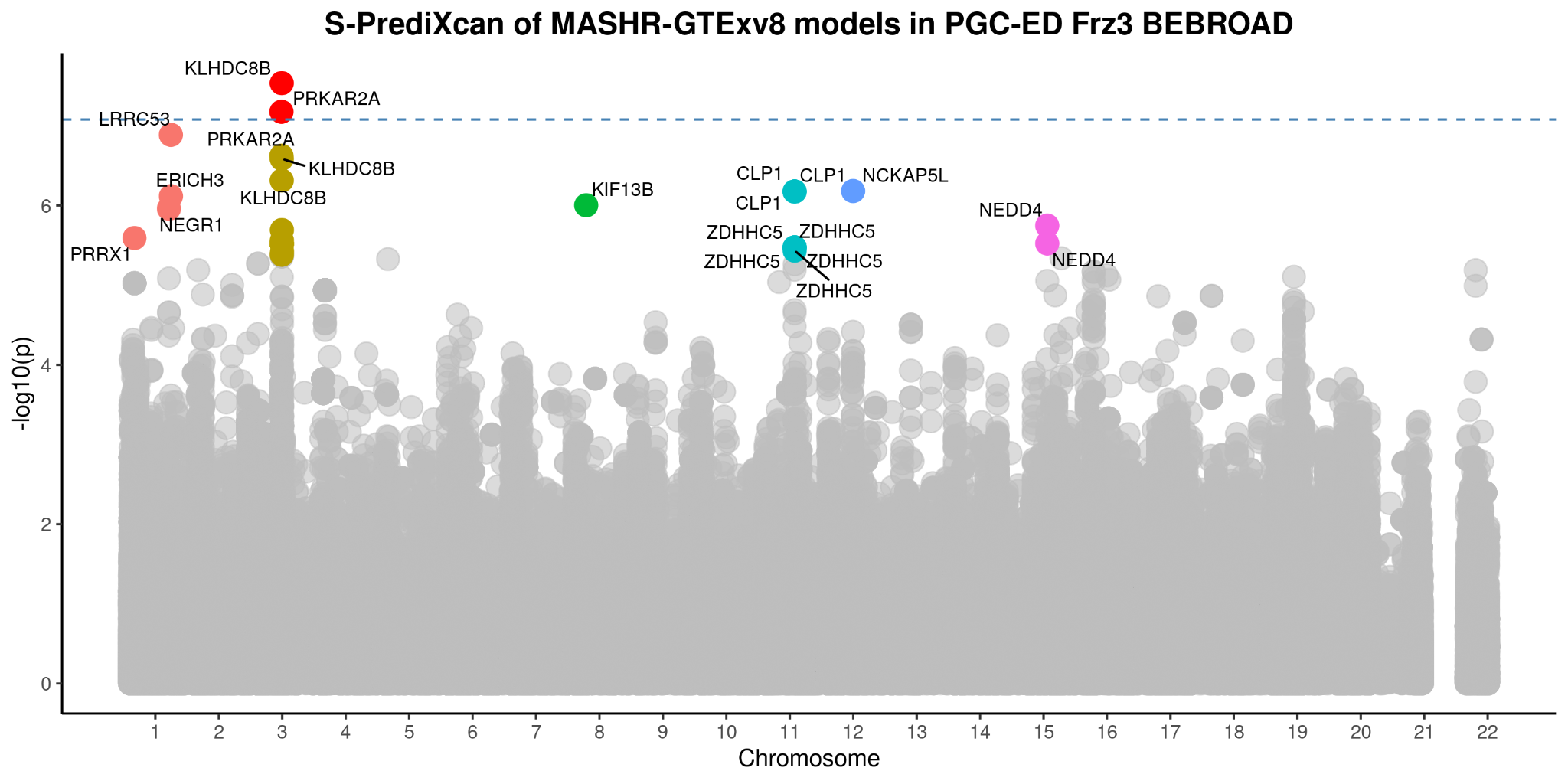

### (b) AN

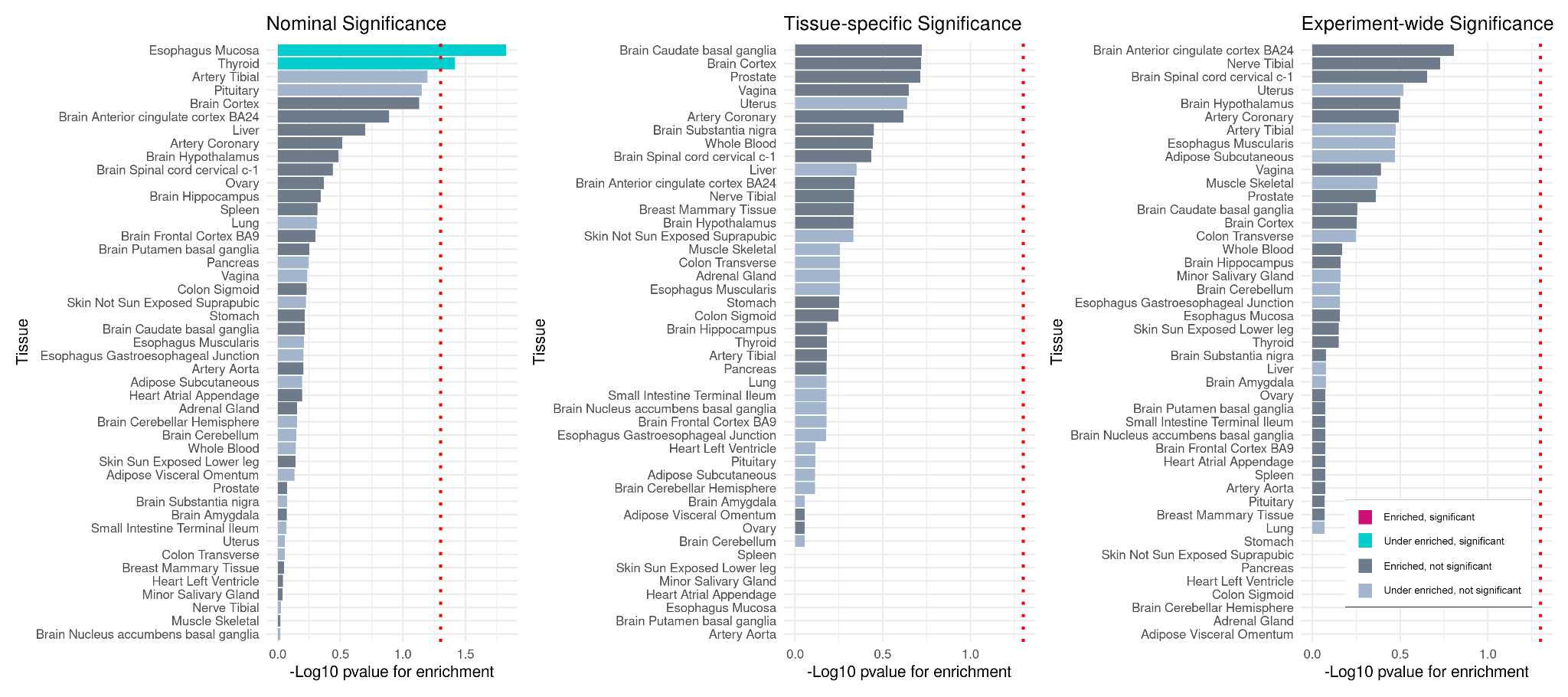

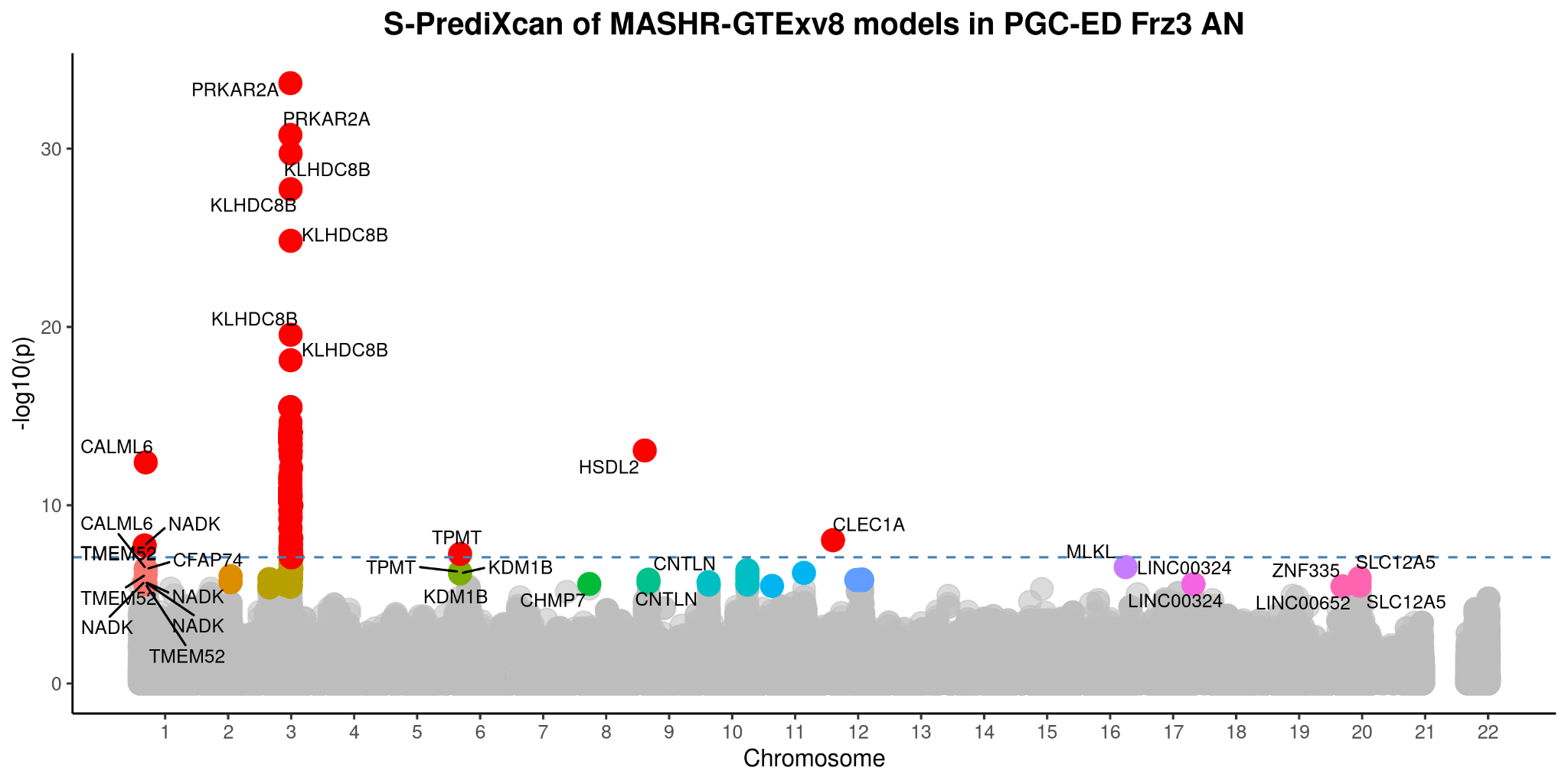

#### (c) BE-NARROW

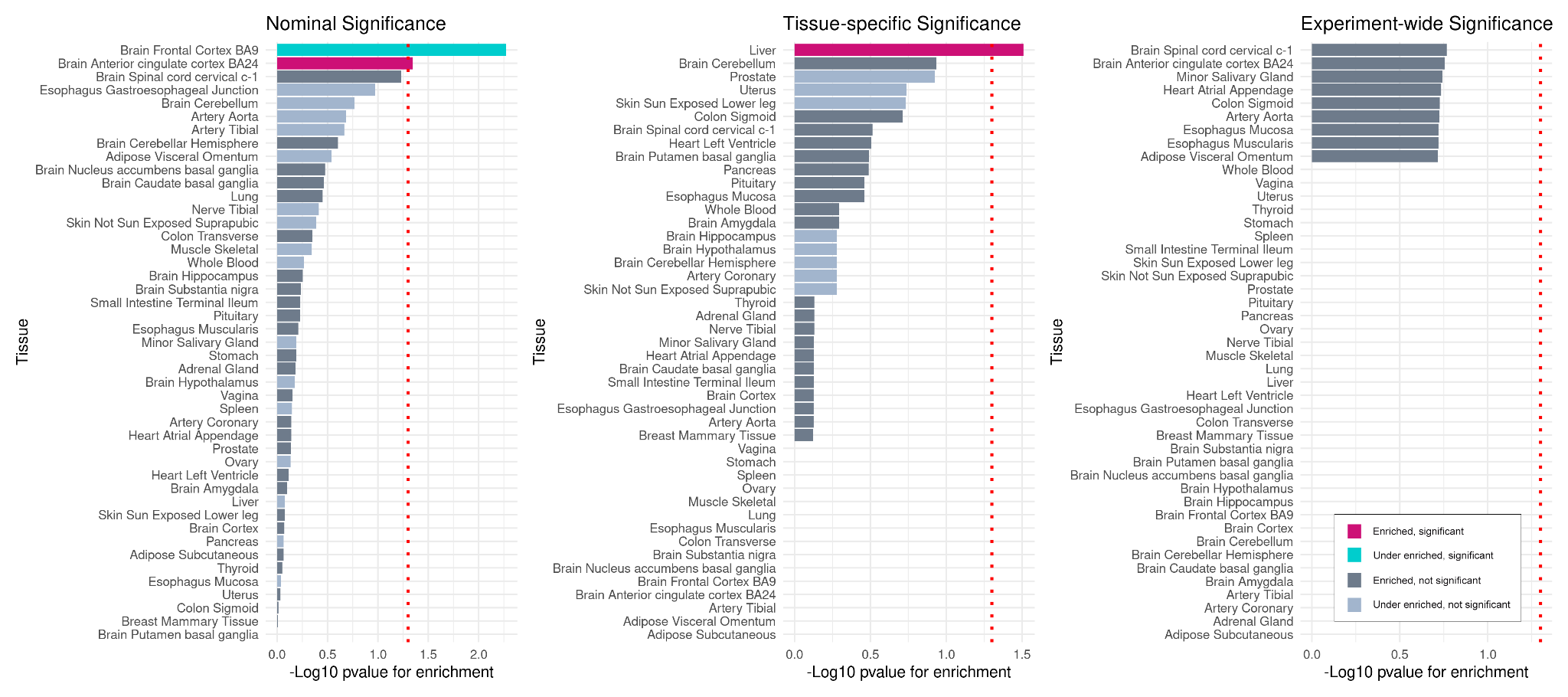

#### (d) ANBP

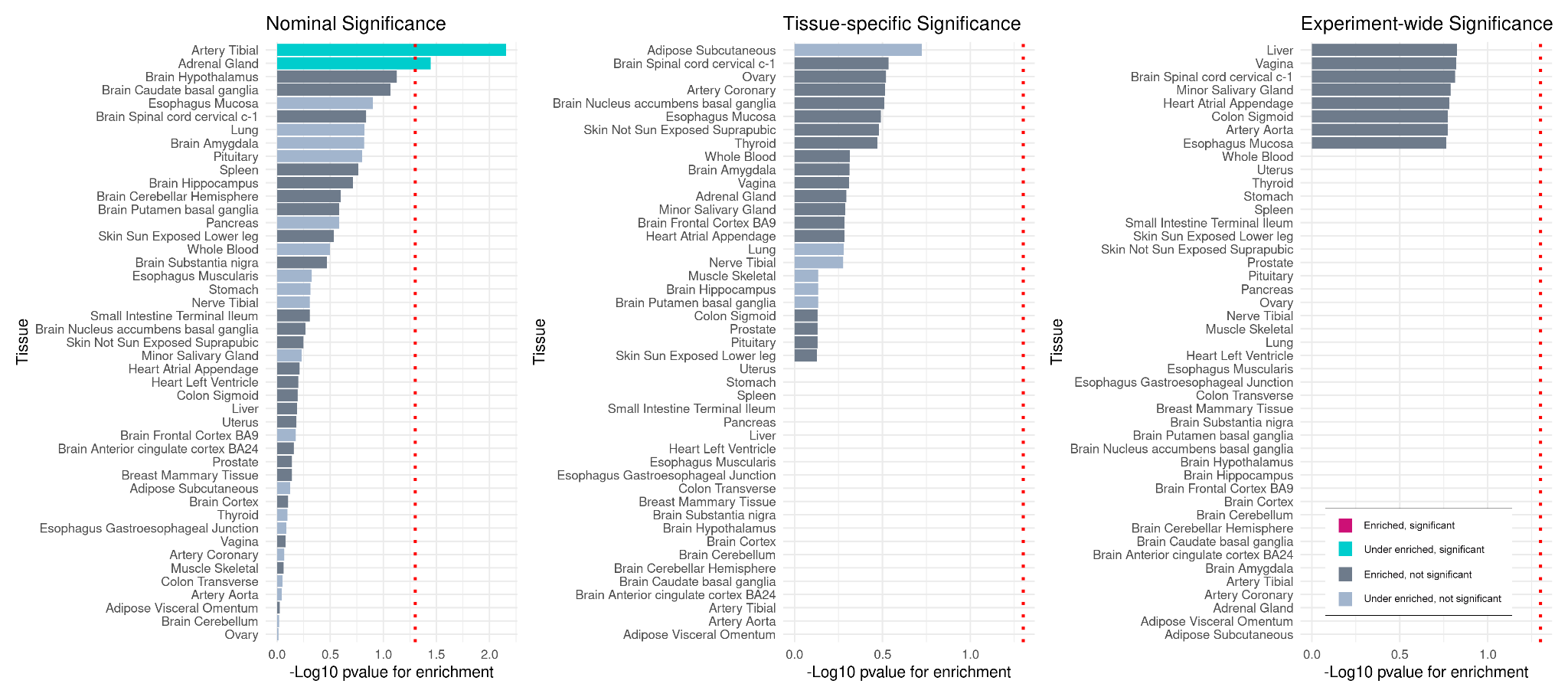

#### (e) ANR

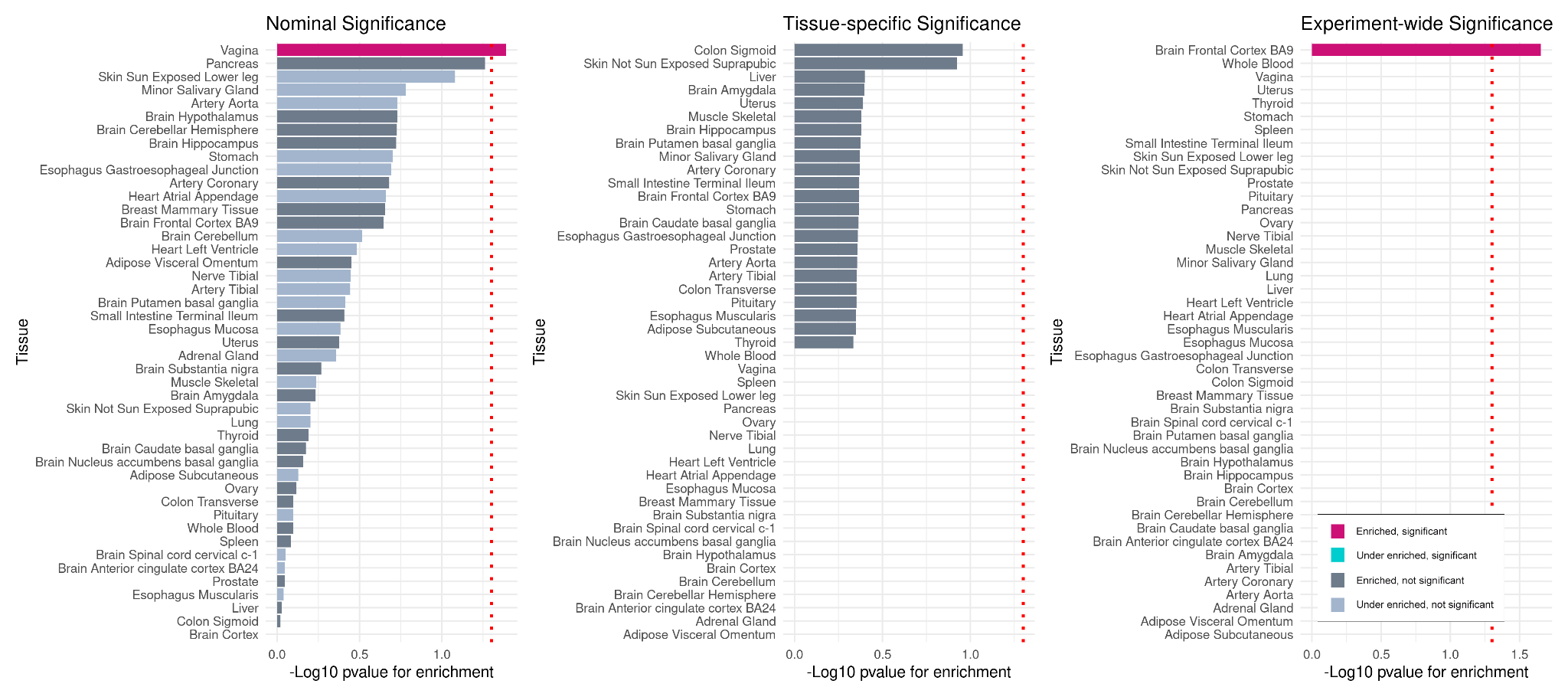

### Supplementary Figure 20: Cross-tissue gene-wise associations from S-MultiXcan for (a) BE-BROAD (b) AN (c) BE-NARROW (d) ANBP (e) ANR.

Manhattan plot of significant gene-tissue associations (coloured points) from S-MultiXcan results, with annotation of top genes. Experiment-wide significance threshold of p<2.25 x 10^-6^ (dotted line).

#### (a) BE-BROAD

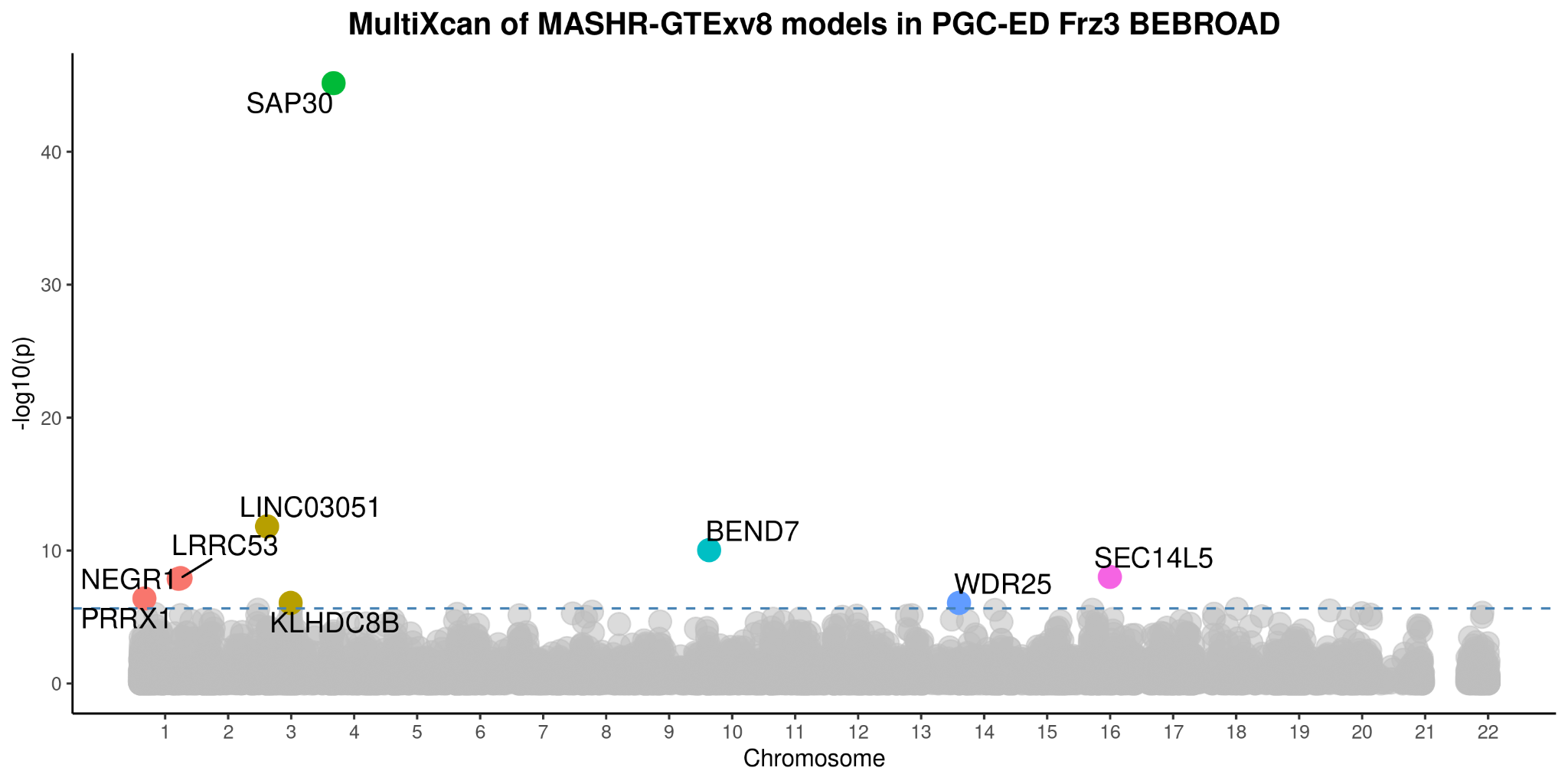

### (b) AN

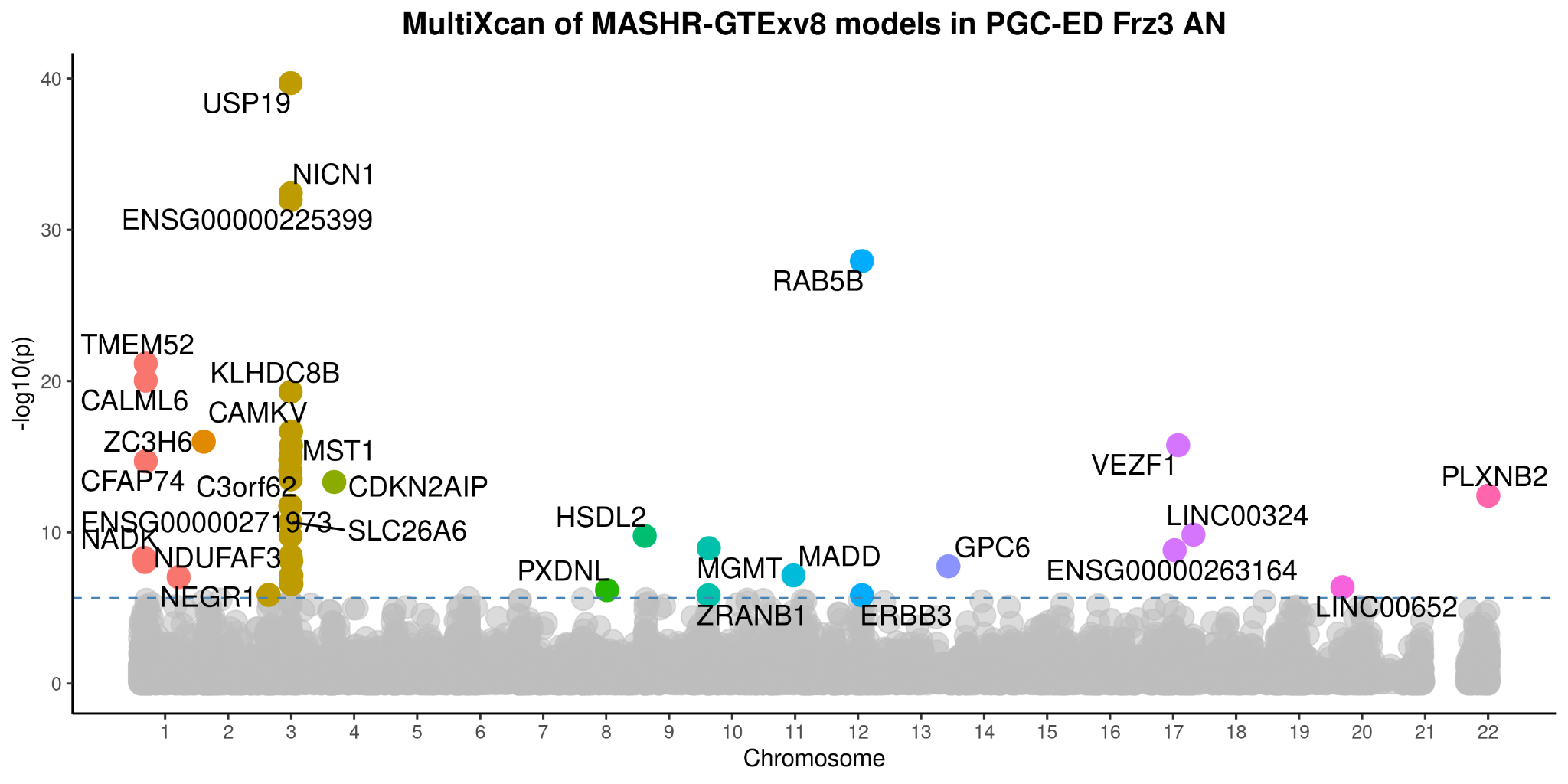

#### (c) BE-NARROW

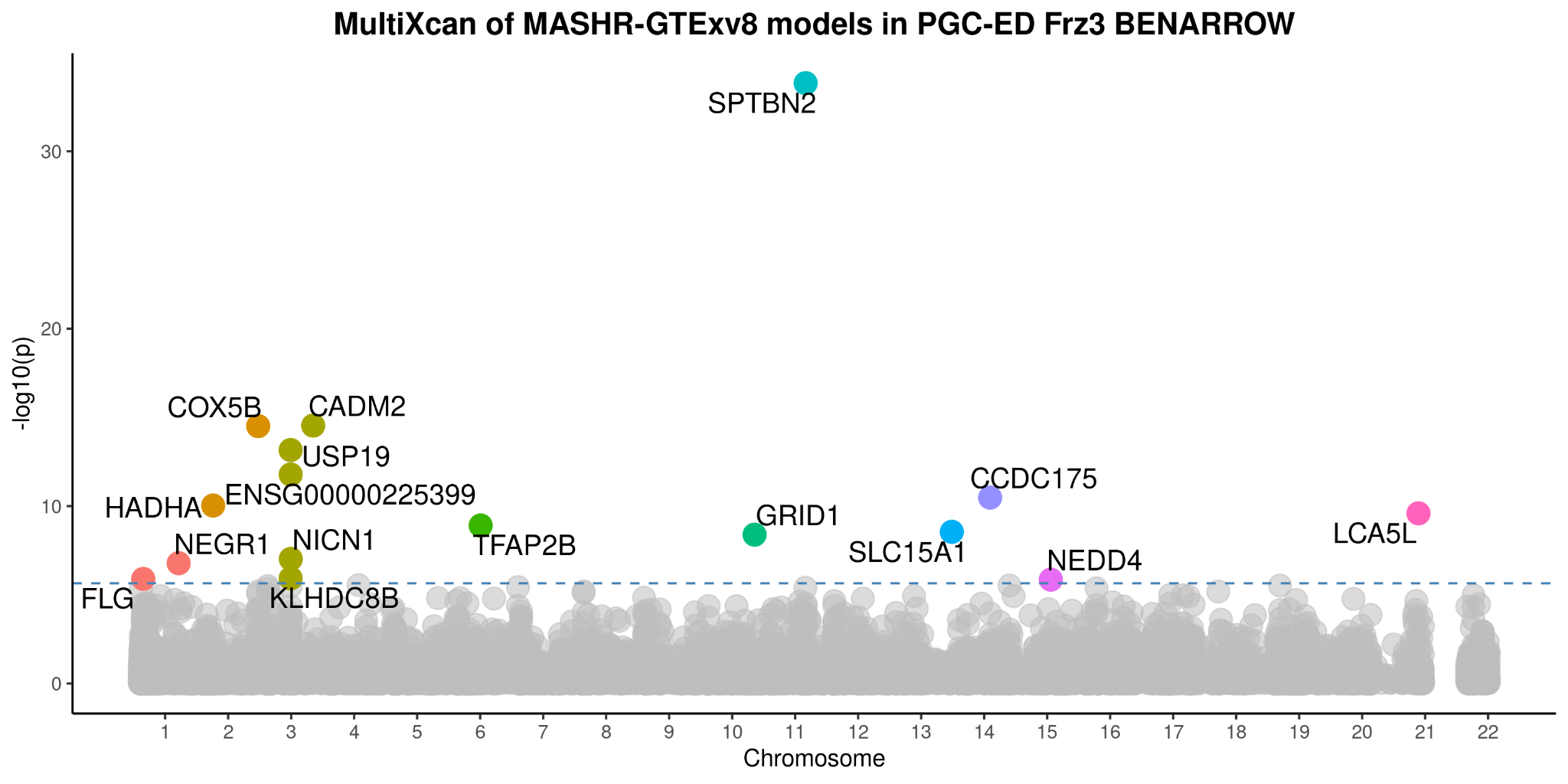

#### (d) ANBP

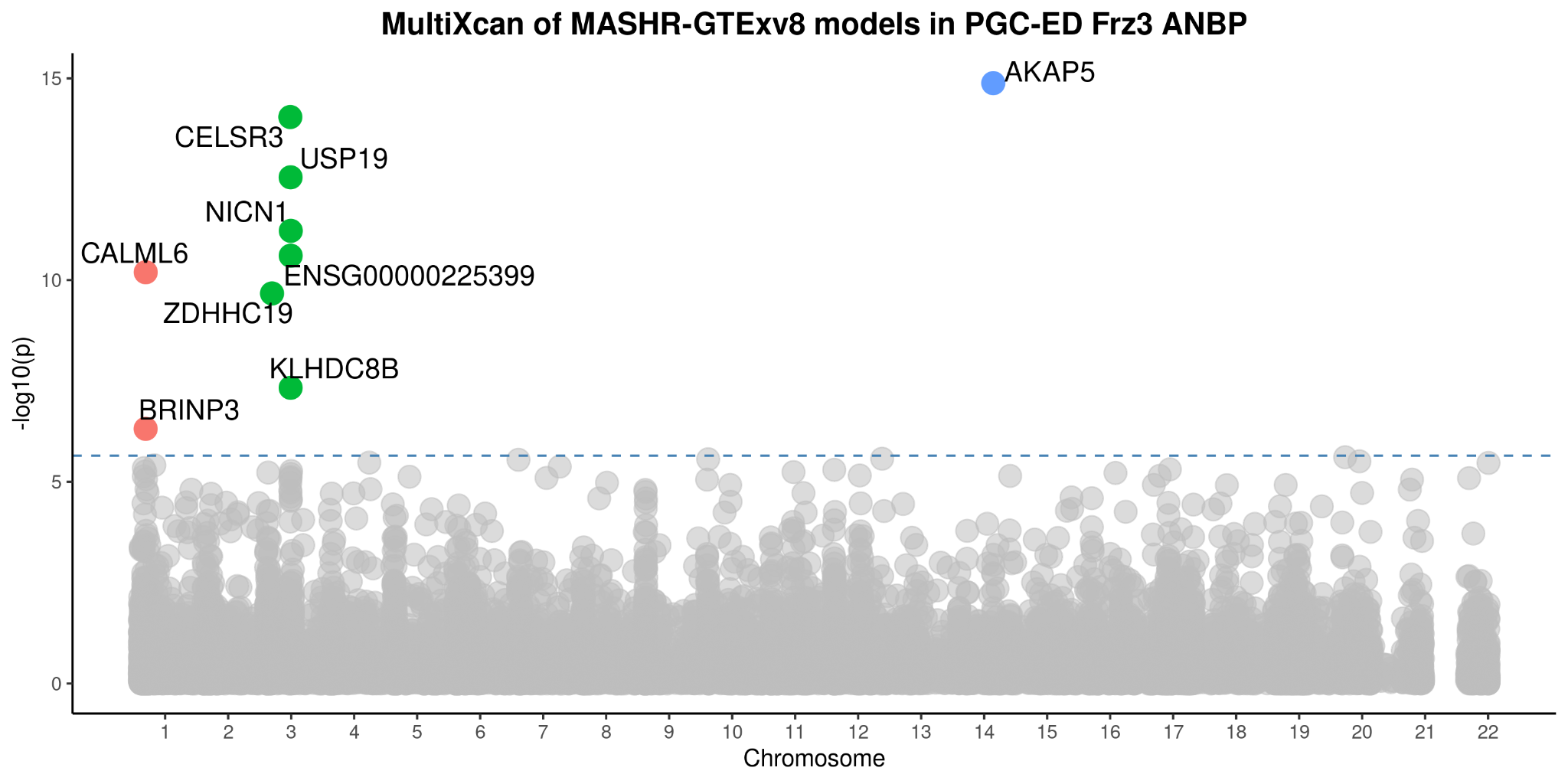

#### (e) ANR

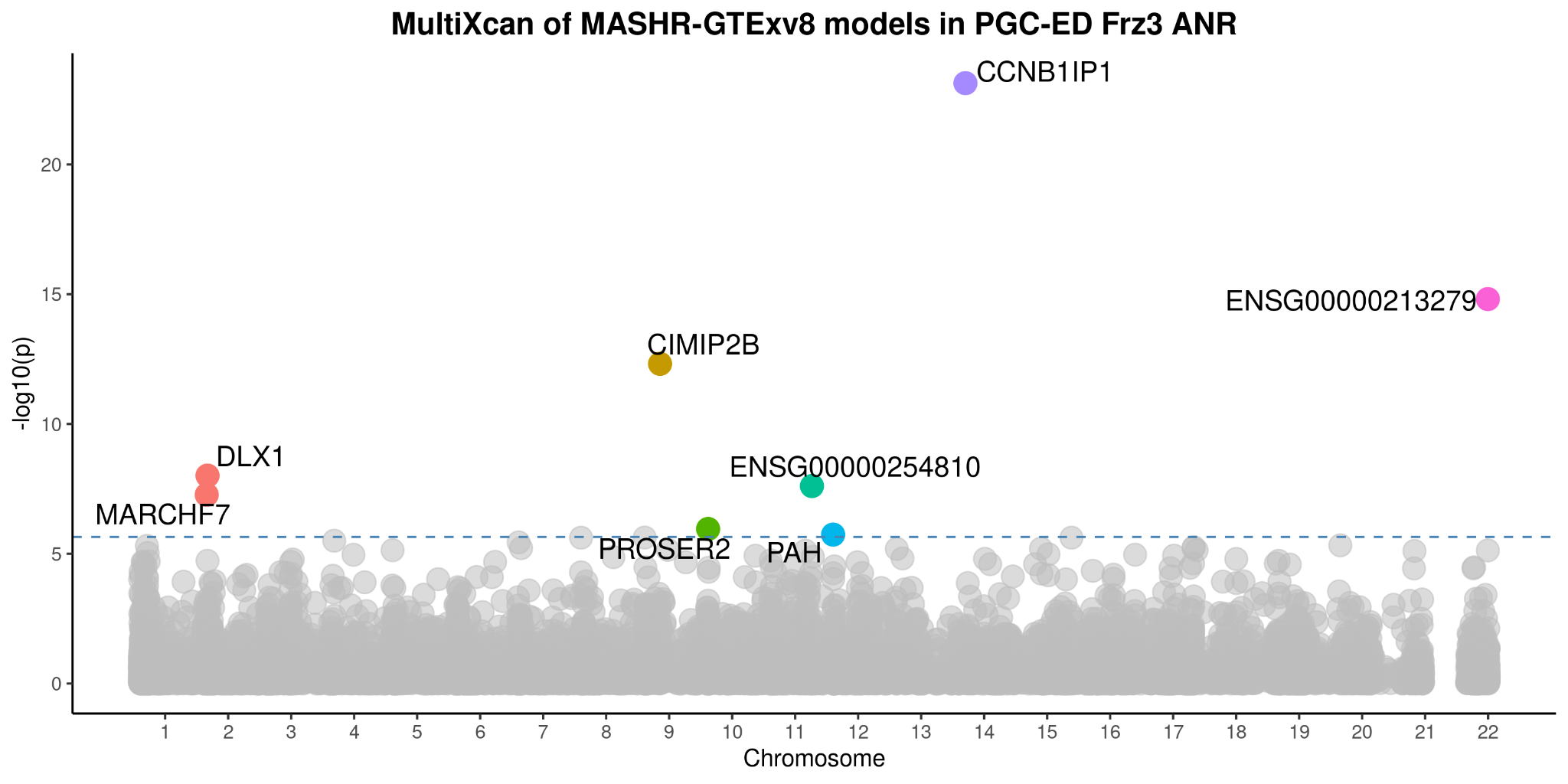

### Supplementary Figure 21 - Tissue and cell-type analyses in GTEx human tissues

This figure presents the significance of the enrichment of eating disorder heritability in human tissues; expression data from GTEx version 8. (a) Barchart for the false discovery rate (FDR) on -log10 scale; dashed line indicates FDR threshold at 0.05. No results survived multiple testing corrections. (b) Heatmap of the unadjusted enrichment P value on -log10 scale. Brain tissues (brain-cortex and brain-other) showed relatively higher levels of significance for all tested eating disorders.

#### Supplementary Figure 21a

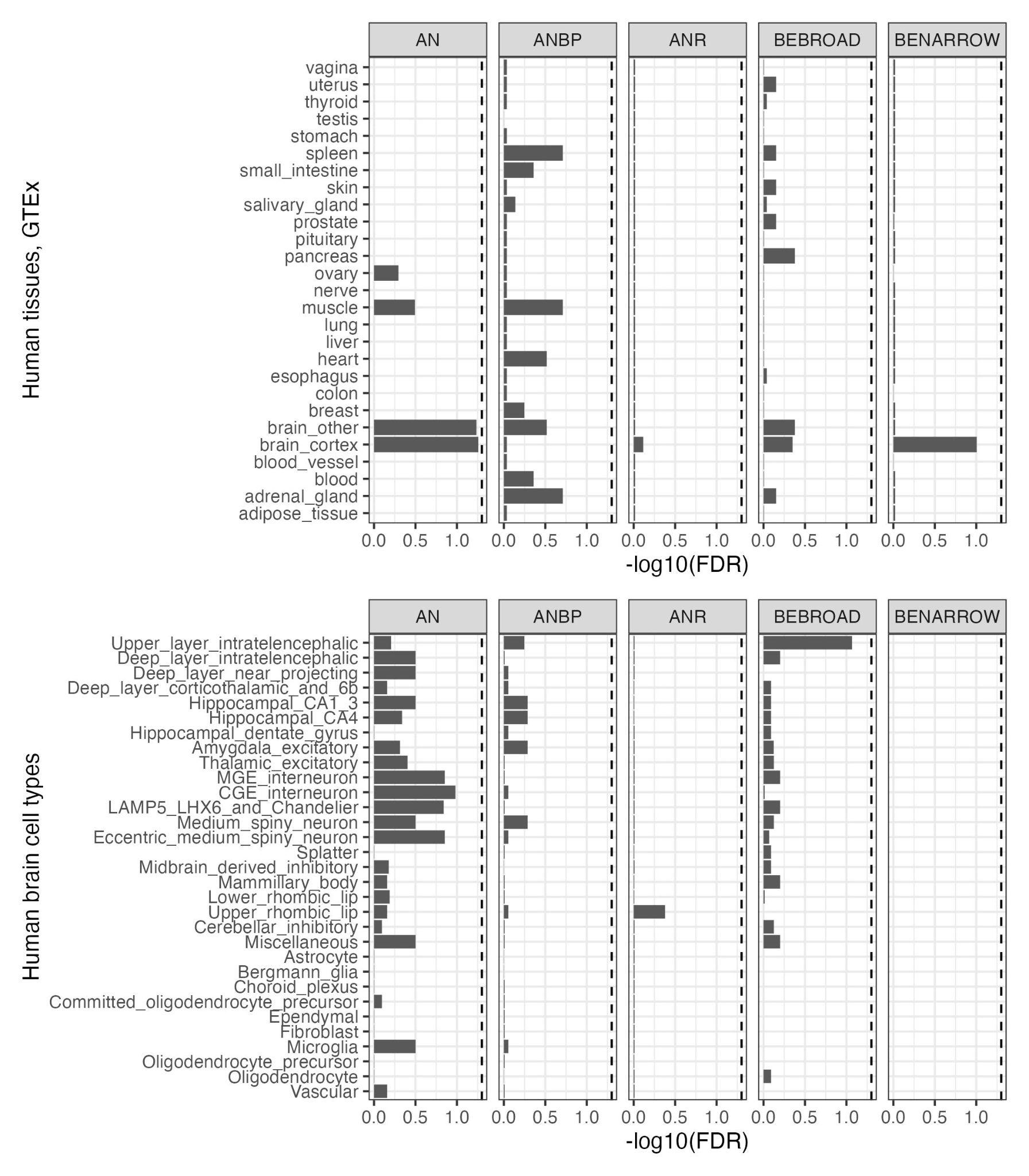

#### Supplementary Figure 21b

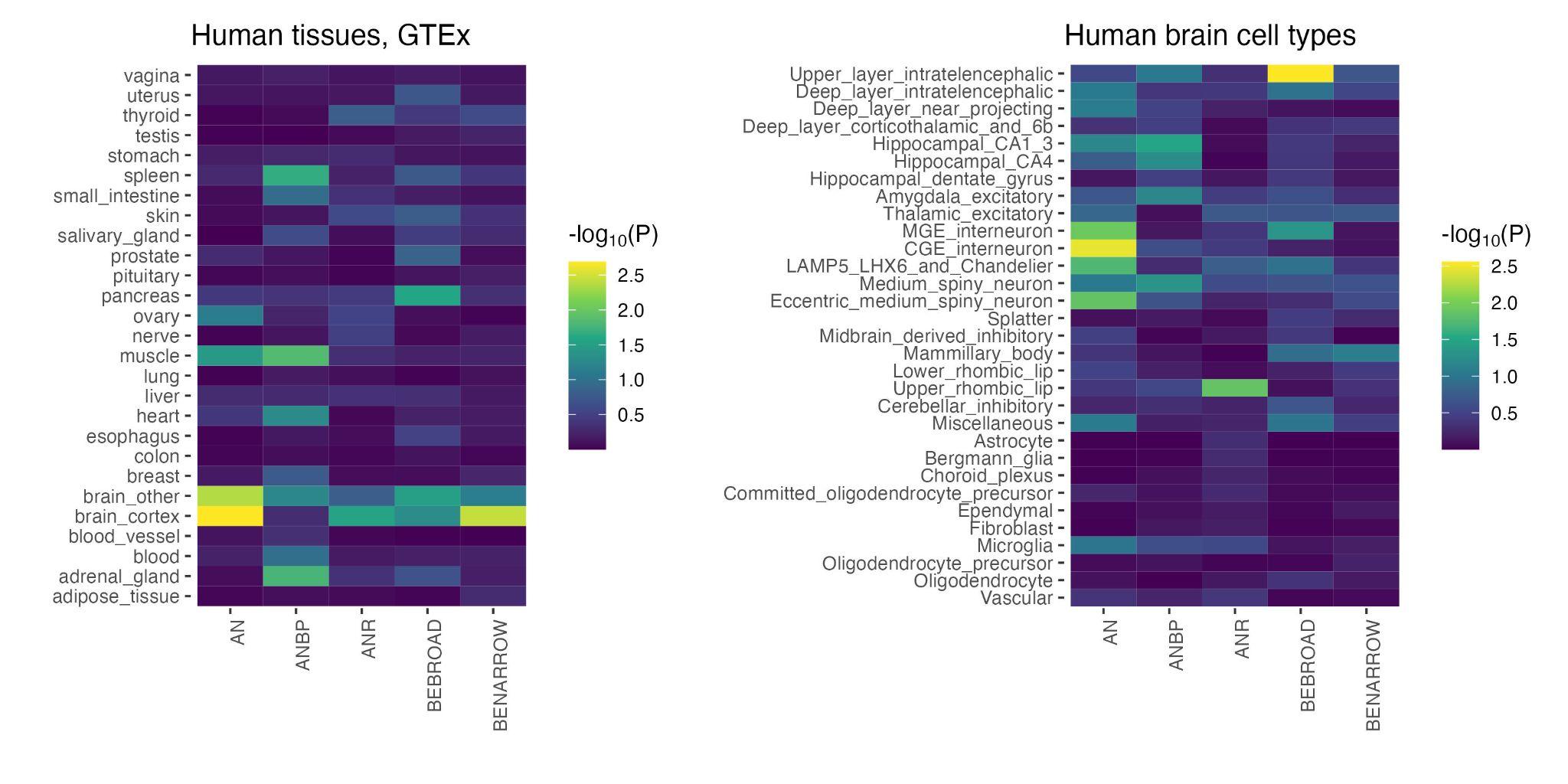

### Supplementary Figure 22 - Tissue and cell-type analyses in human brain cell types

This figure presents the significance of the enrichment of eating disorder heritability in human brain cell type; based on the 31 superclusters from the single-nucleus RNA-sequencing adult brain atlas. (a) Barchart for the false discovery rate (FDR) on -log10 scale; dashed line indicates FDR threshold at 0.05. No results survived multiple testing corrections. (b) Heatmap of the unadjusted enrichment P value on -log10 scale. The highest significance results are observed in the upper-layer intratelencephalic neurons for BEBROAD. For AN, the higher levels of significance were seen in inhibitory neurons, particularly caudal ganglionic eminence (CGE) interneurons, followed by excitatory neurons.

#### Supplementary Figure 22a

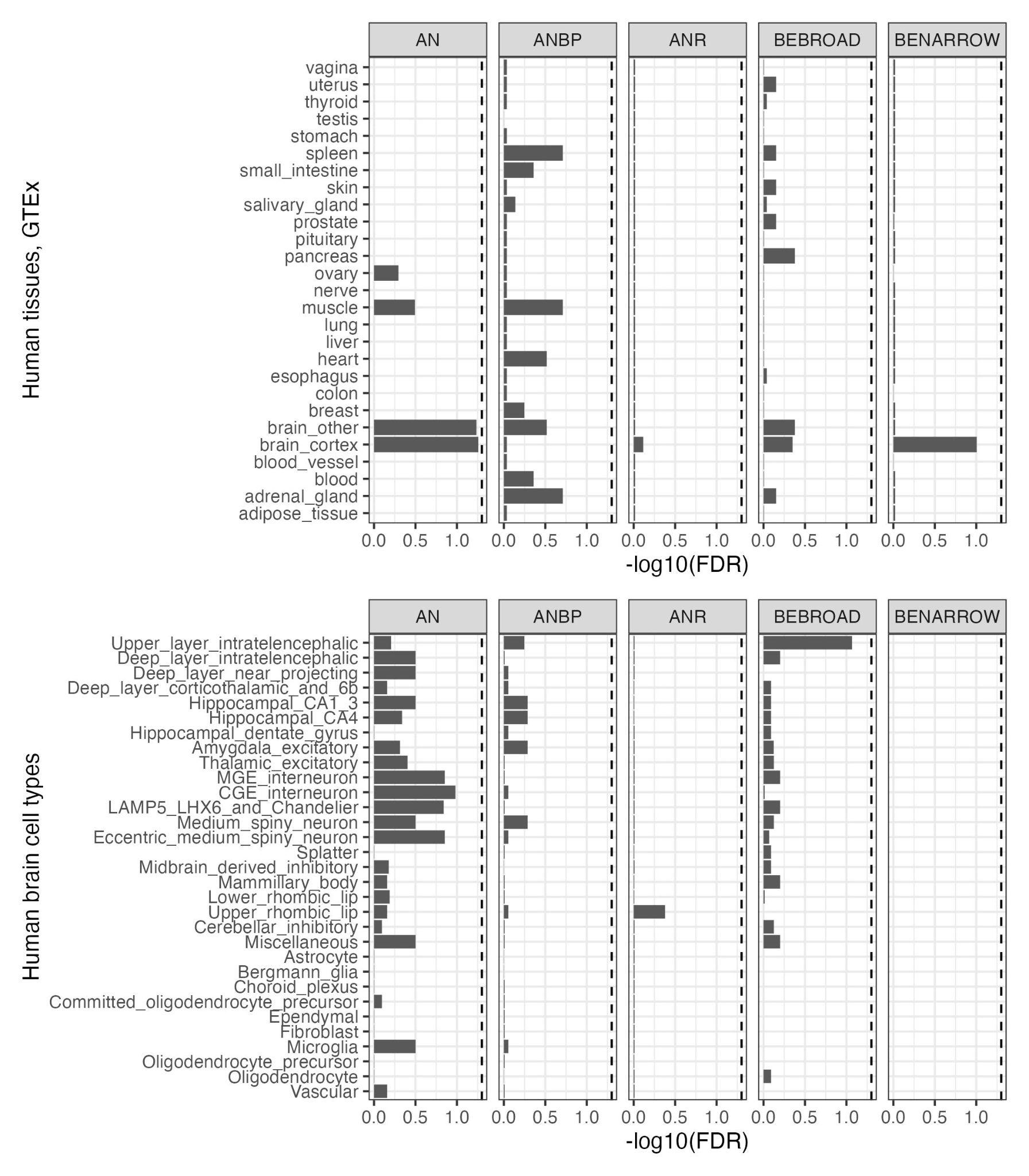

#### Supplementary Figure 22b

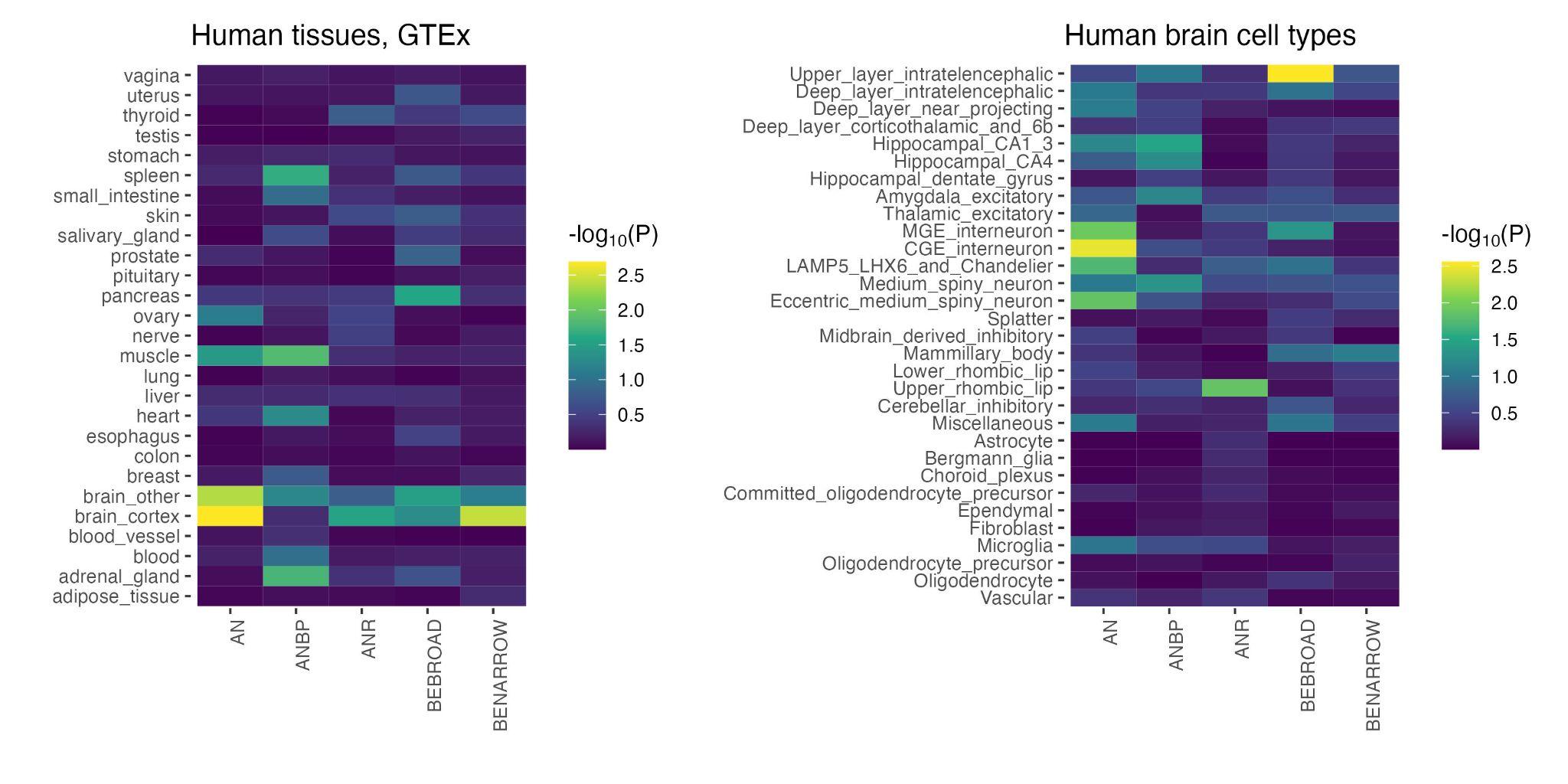

### Supplementary Figure 23: Within-and-cross-trait polygenic scoring.

Decile plots depict the odds ratio (OR) of each decile of polygenic risk scores of BE-BROAD, BE-NARROW, and AN on the BE-BROAD, BE-NARROW, AN outcomes, with the lowest decile serving as the reference group. The receiver operating characteristic (ROC) curve is a graphical illustration of the performance of a regression model to predict the binary outcome (i.e., case/control status of BE-BROAD, BE-NARROW, or AN here). If a regression model has a ROC curve overlapping with the diagonal grey line in the ROC curve plot, it indicates that the regression model is no better than chance to differentiate cases and controls with an area under the curve (AUC) of 0.5. An AUC greater than 0.5 suggests that the prediction performance of a model is better than chance and an AUC of 1 indicates perfect performance. In each ROC curve plot, ROC curves of two models are plotted: one for the baseline model (e.g., genetic principal components (PCs) as predictor variables) and one for the PRS model (i.e., both PRS and baseline predictors are included as predictor variables). Overall, the PRS model improves the predictive performance on BE-BROAD, BE-NARROW, and AN outcomes, but to a small extent.

#### Supplementary Figure 23a: Decile plots and area under the receiver operating characteristic curve for the association of BE-BROAD PRS with BE-BROAD

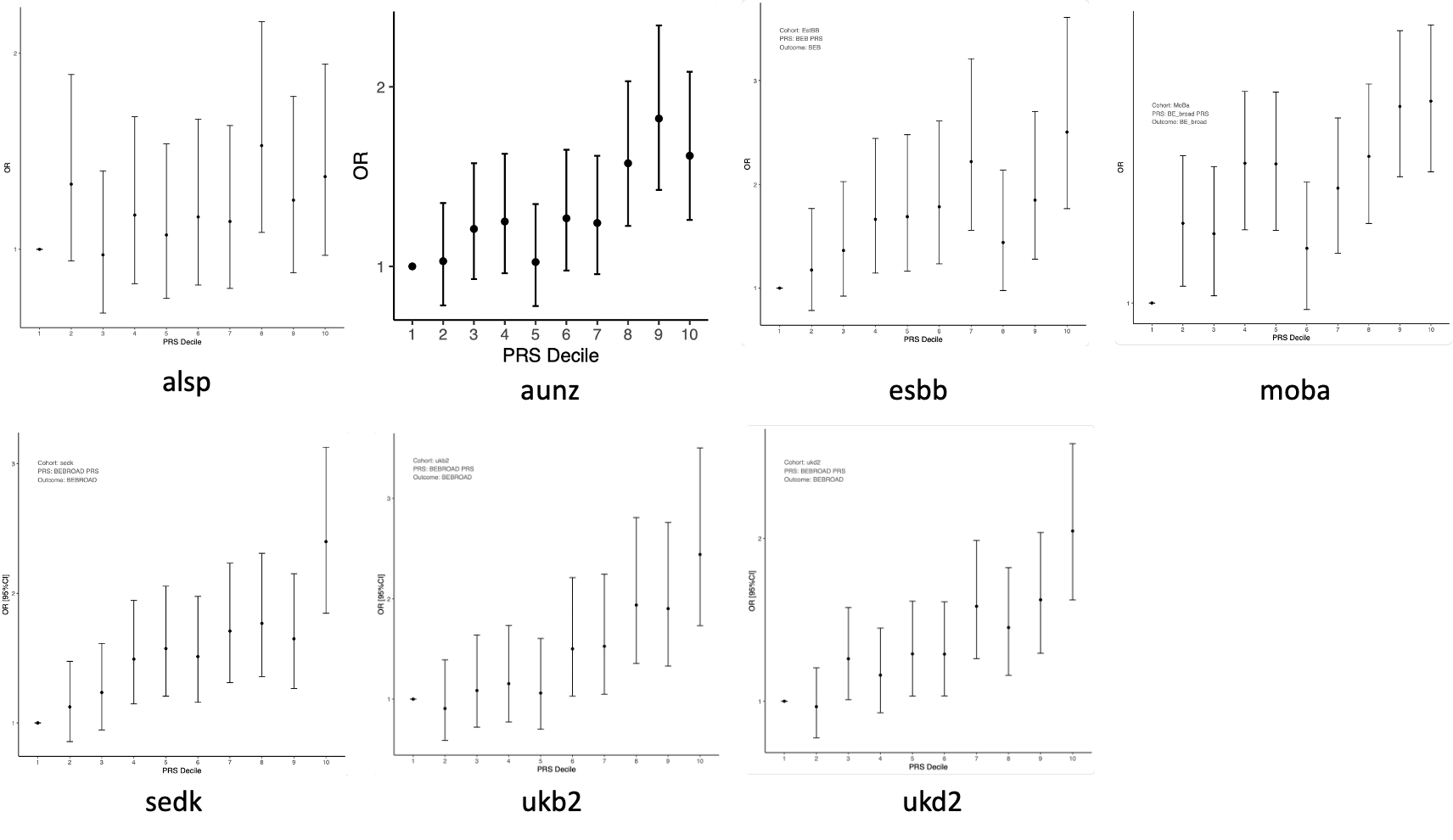

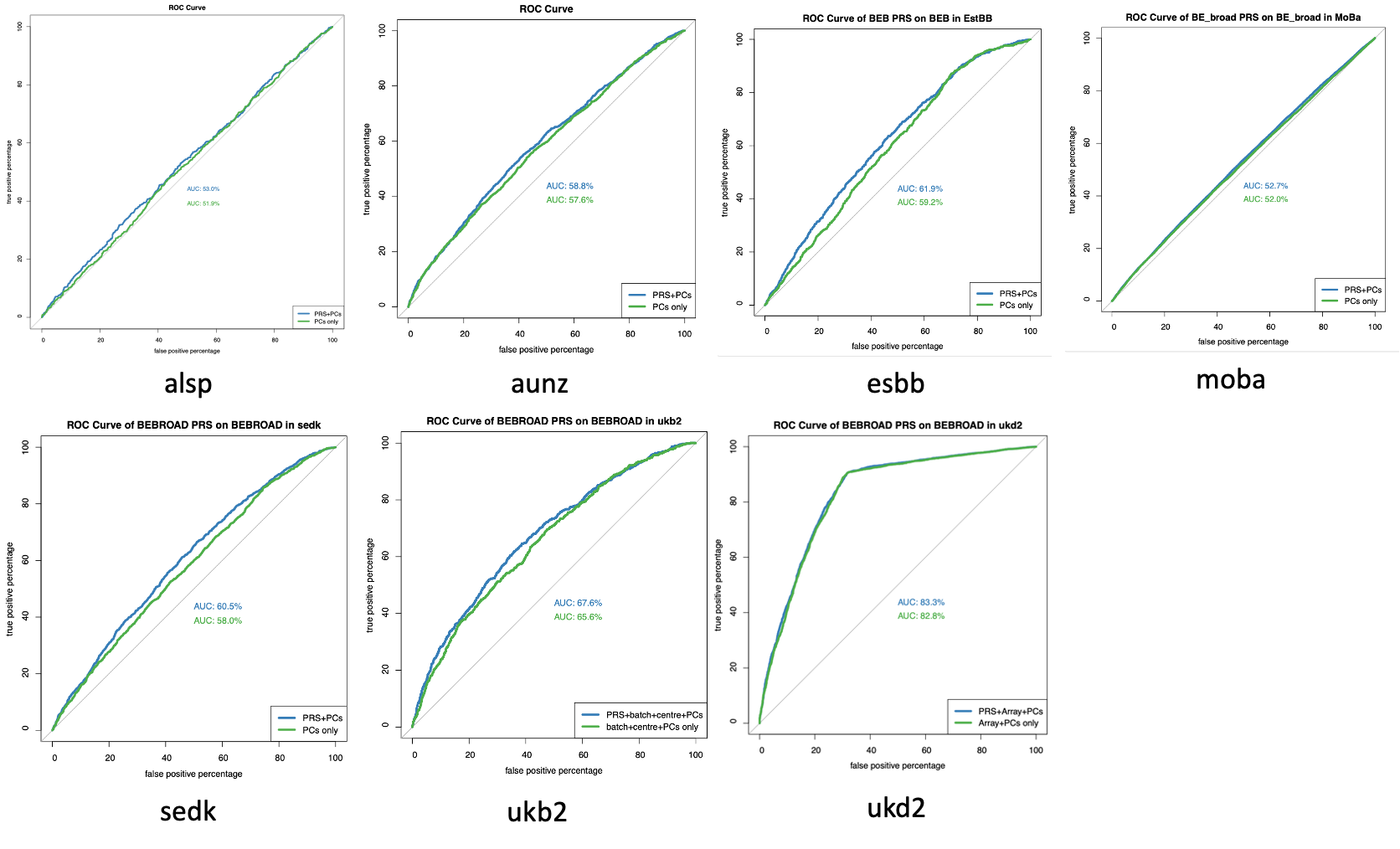

#### Supplementary Figure 23b: Decile plots and area under the receiver operating characteristic curve for the association of AN PRS with AN

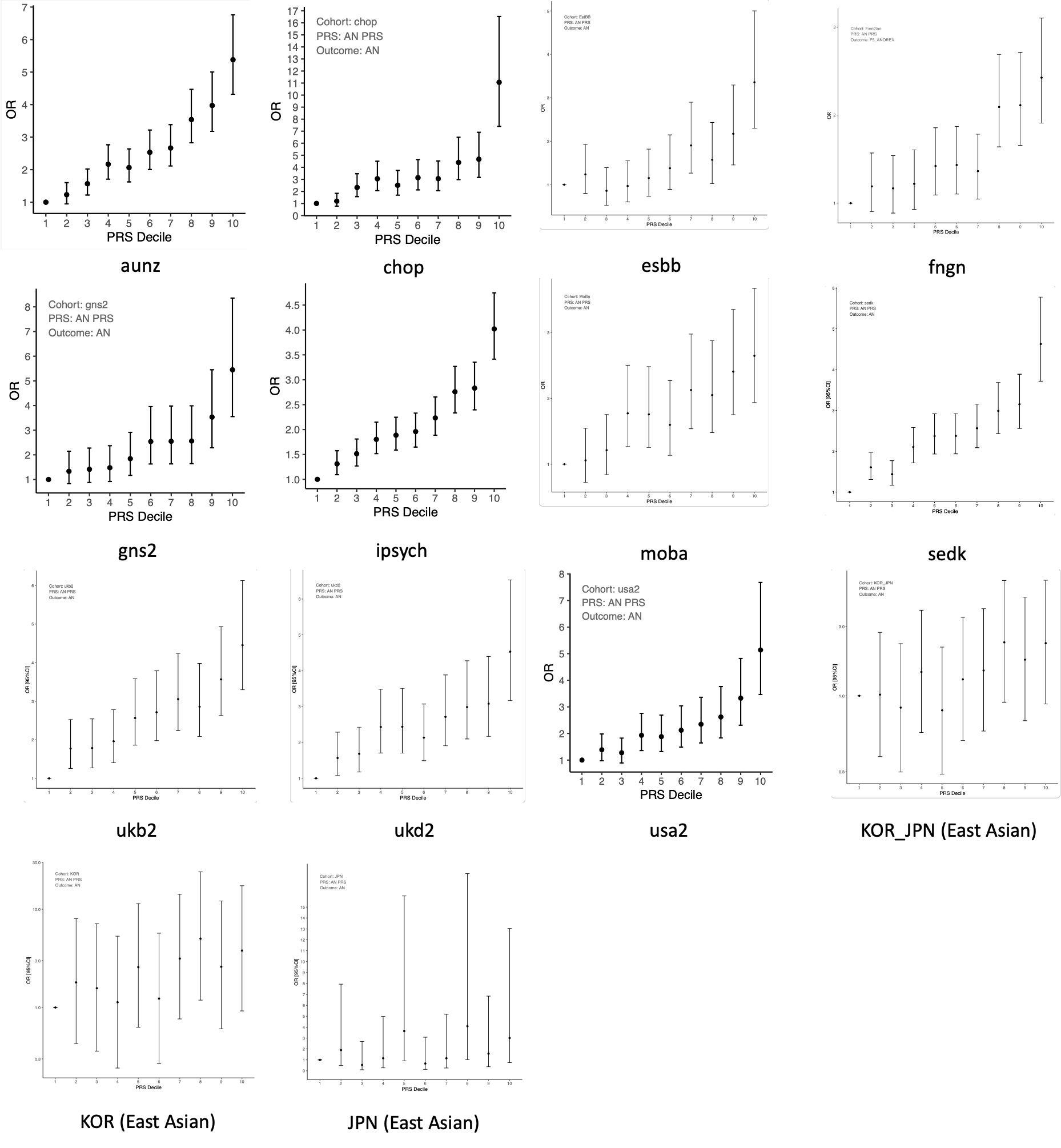

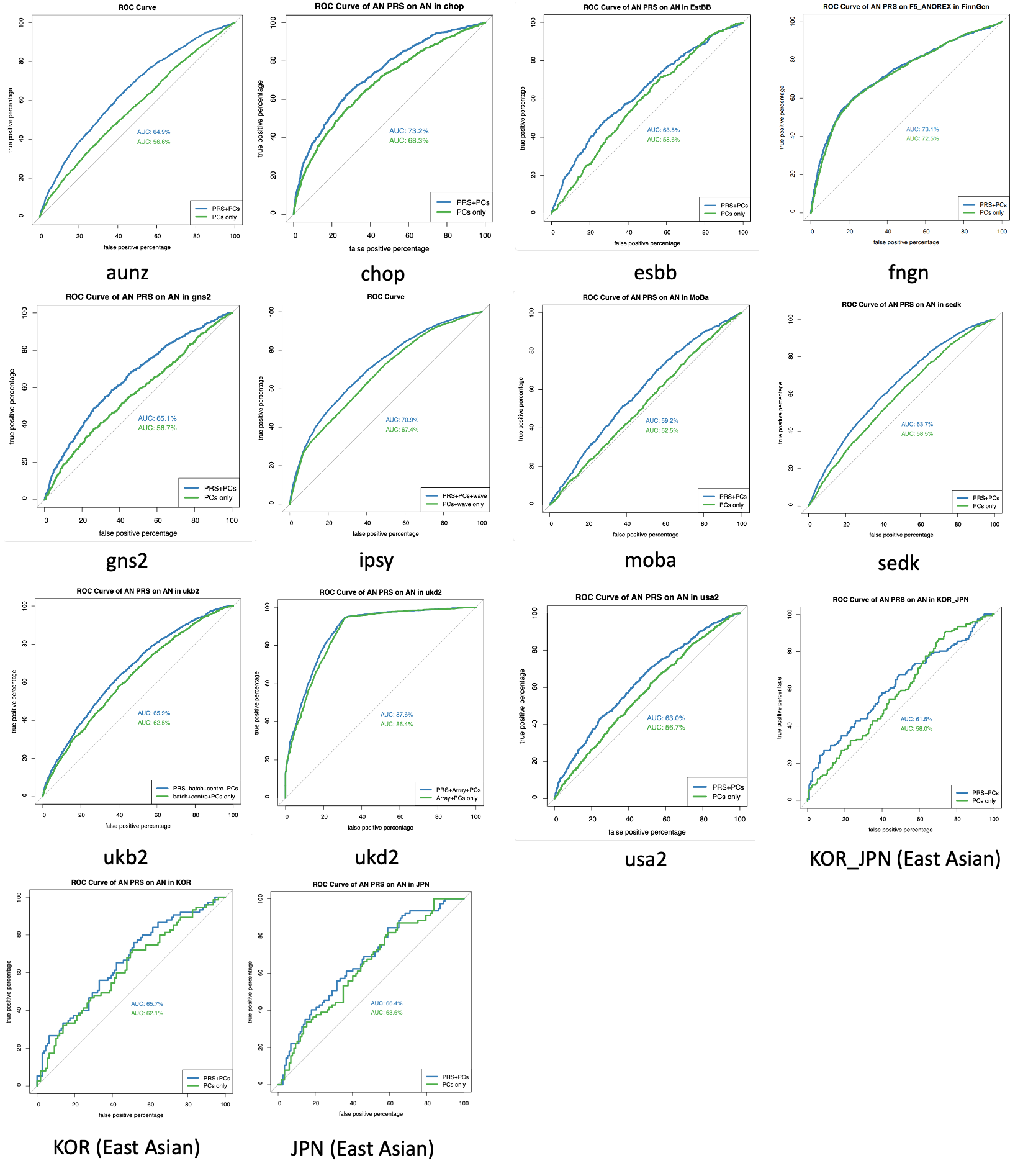

#### Supplementary Figure 23c: Decile plots and area under the receiver operating characteristic curve for the association of BE-BROAD PRS with BE-NARROW

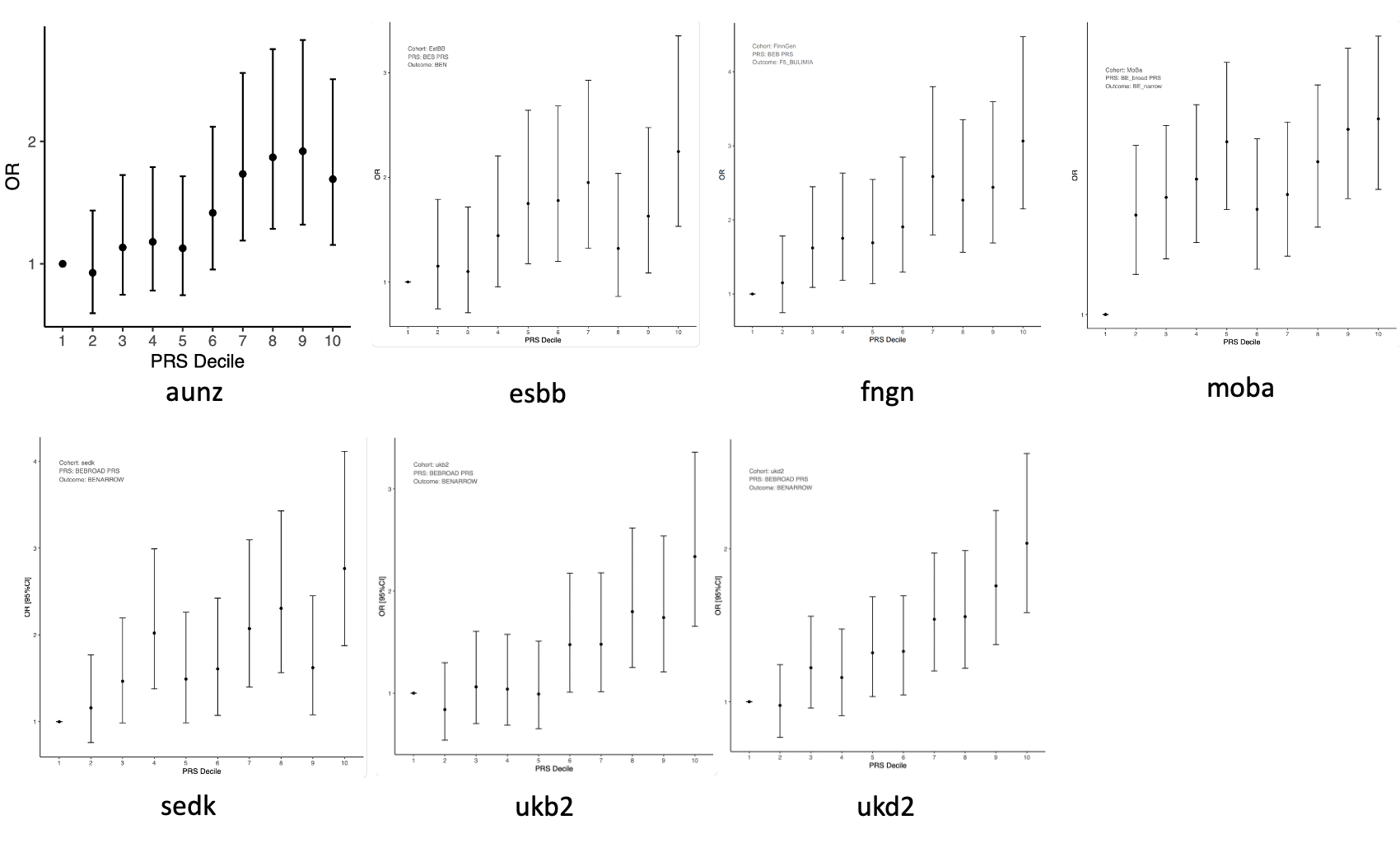

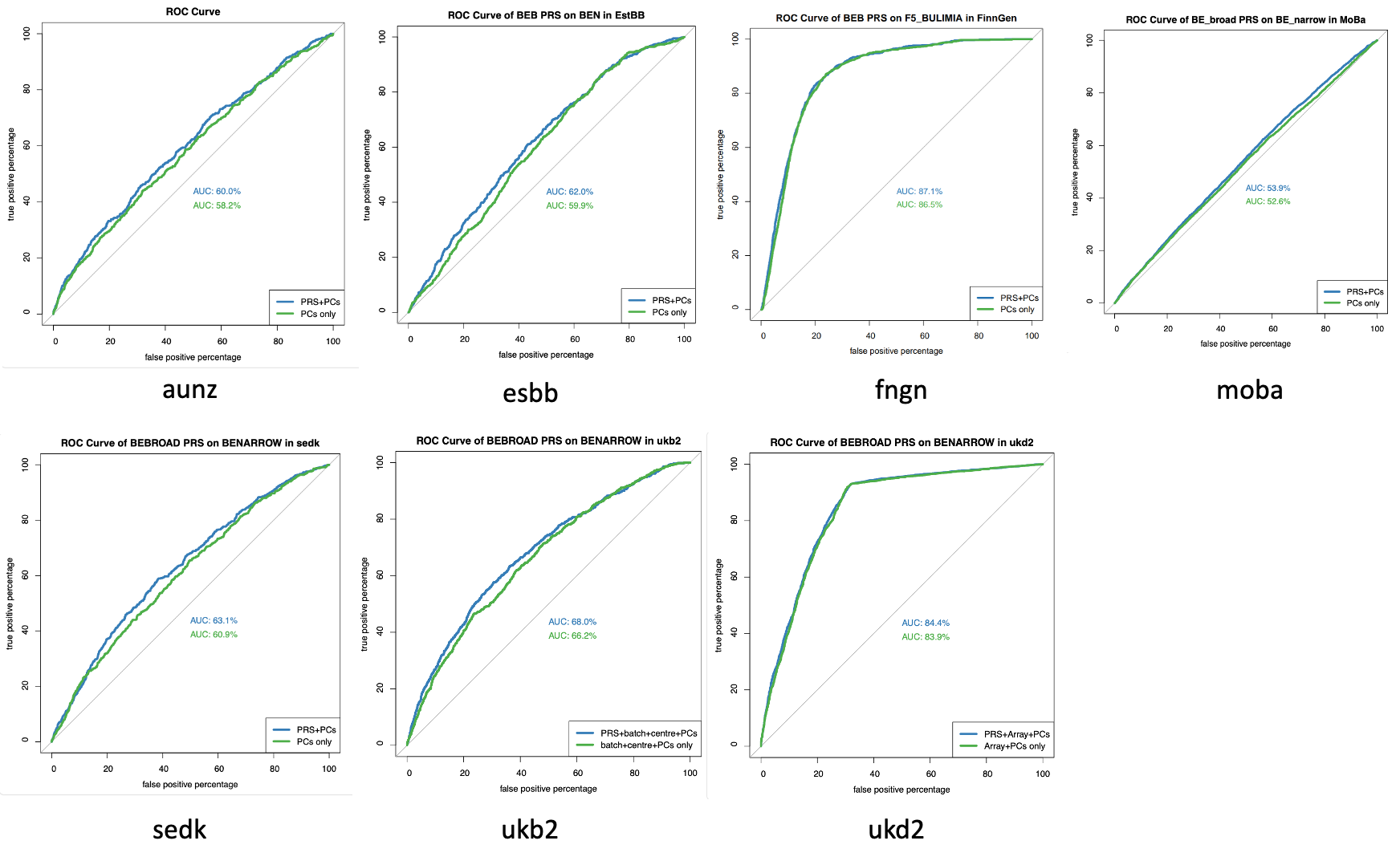

#### Supplementary Figure 23d: Decile plots and area under the receiver operating characteristic curve for the association of BE-BROAD PRS with AN

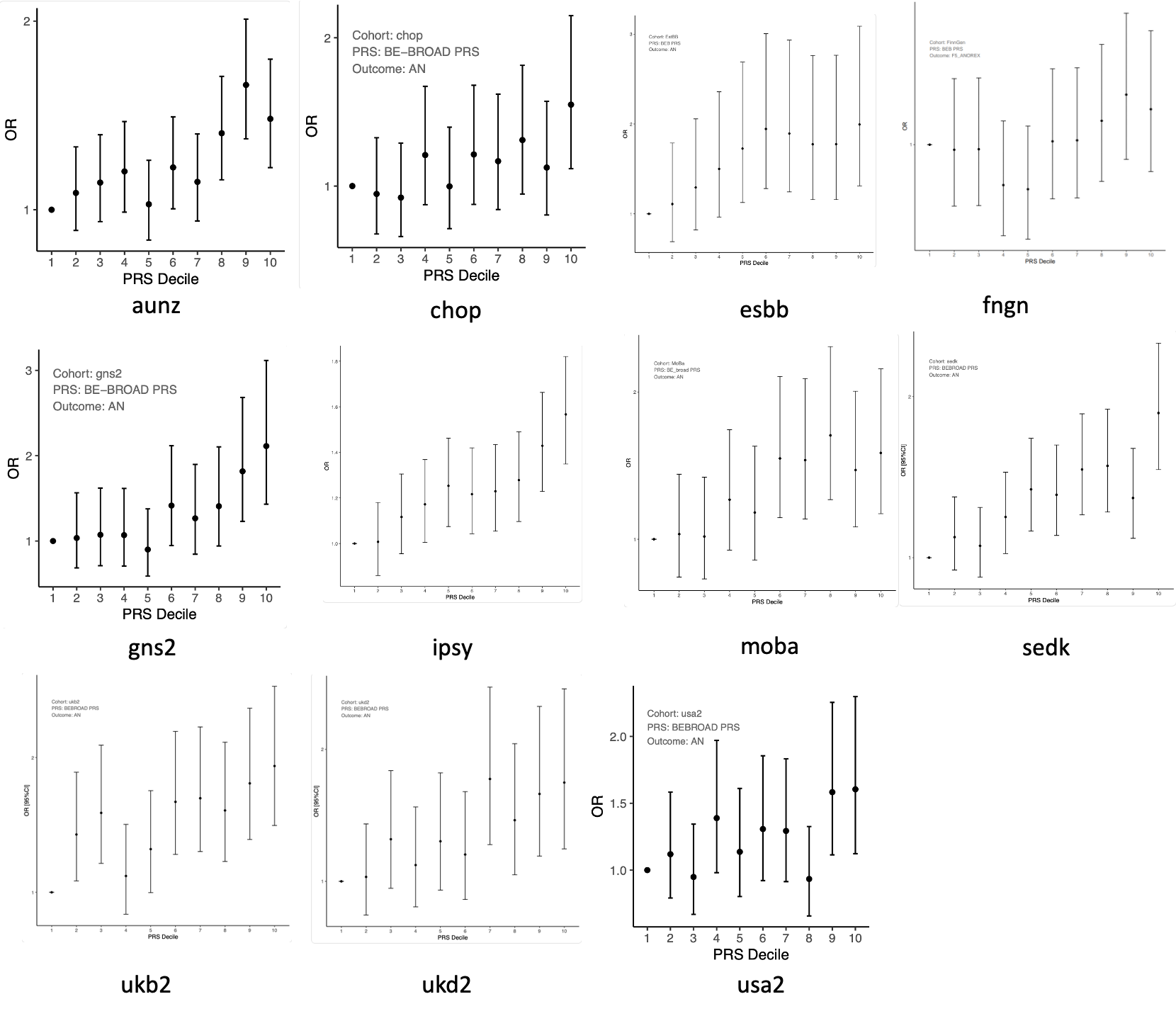

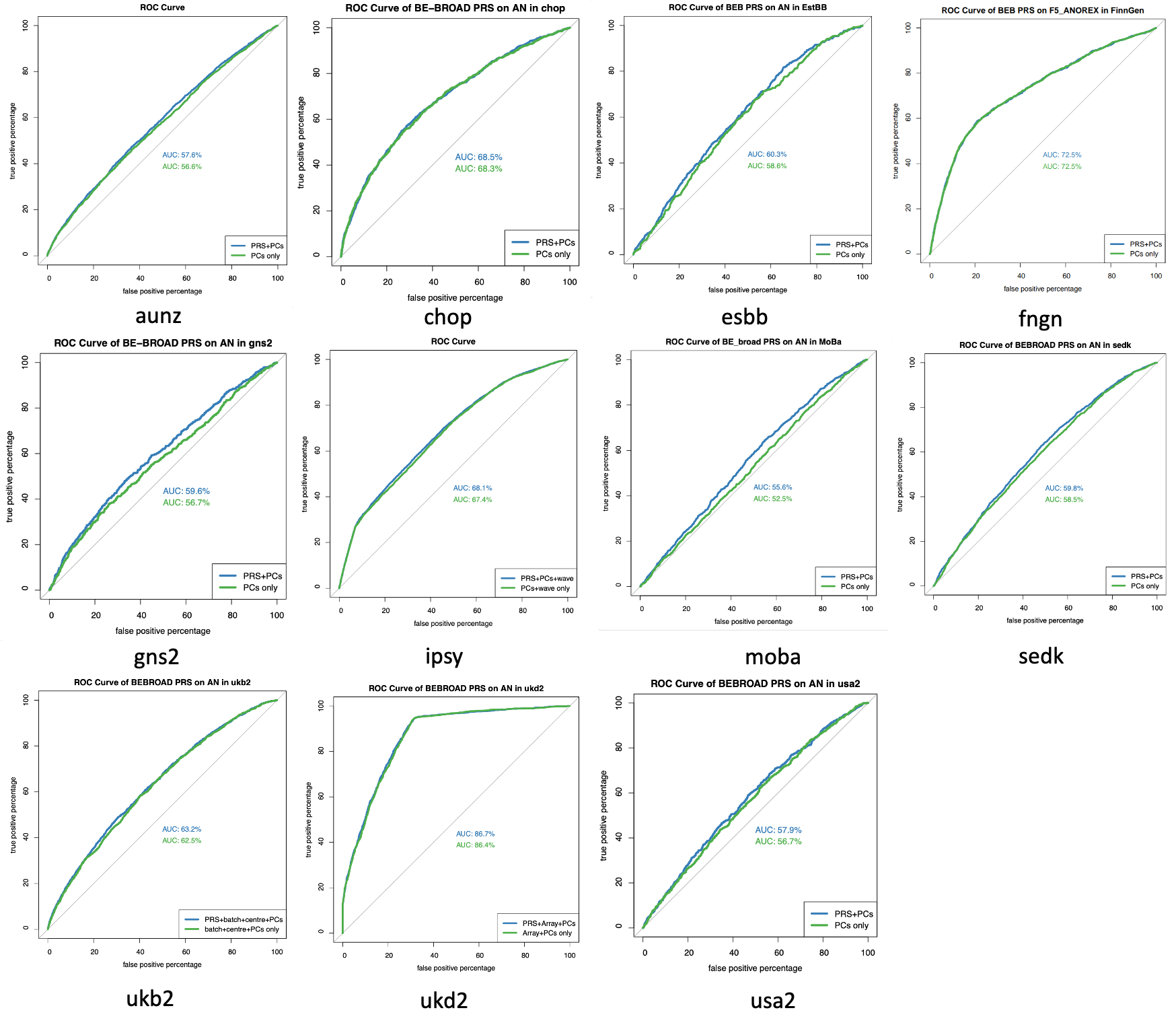

#### Supplementary Figure 23e: Decile plots and area under the receiver operating characteristic curve for the association of BE-NARROW PRS with BE-NARROW

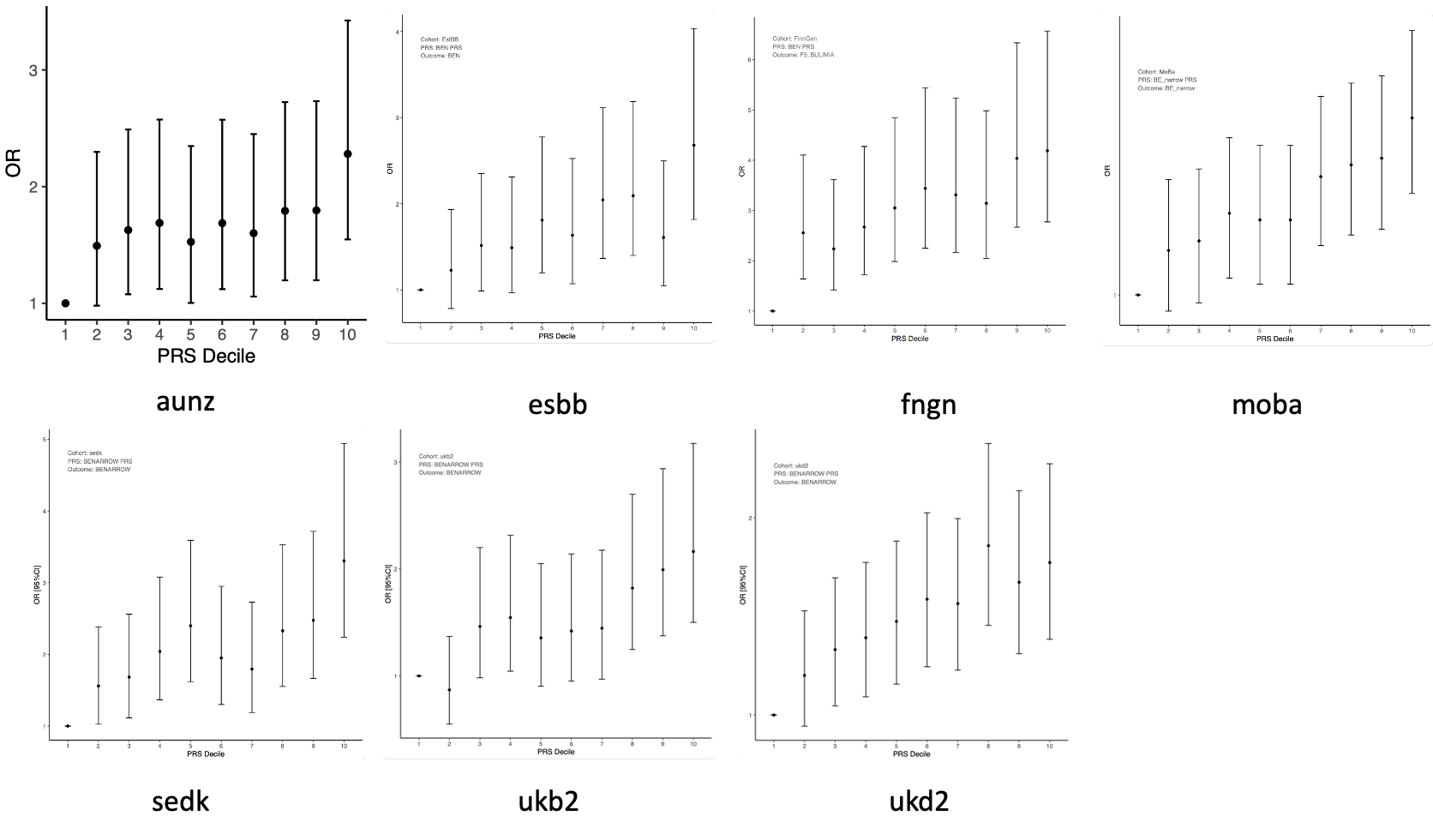

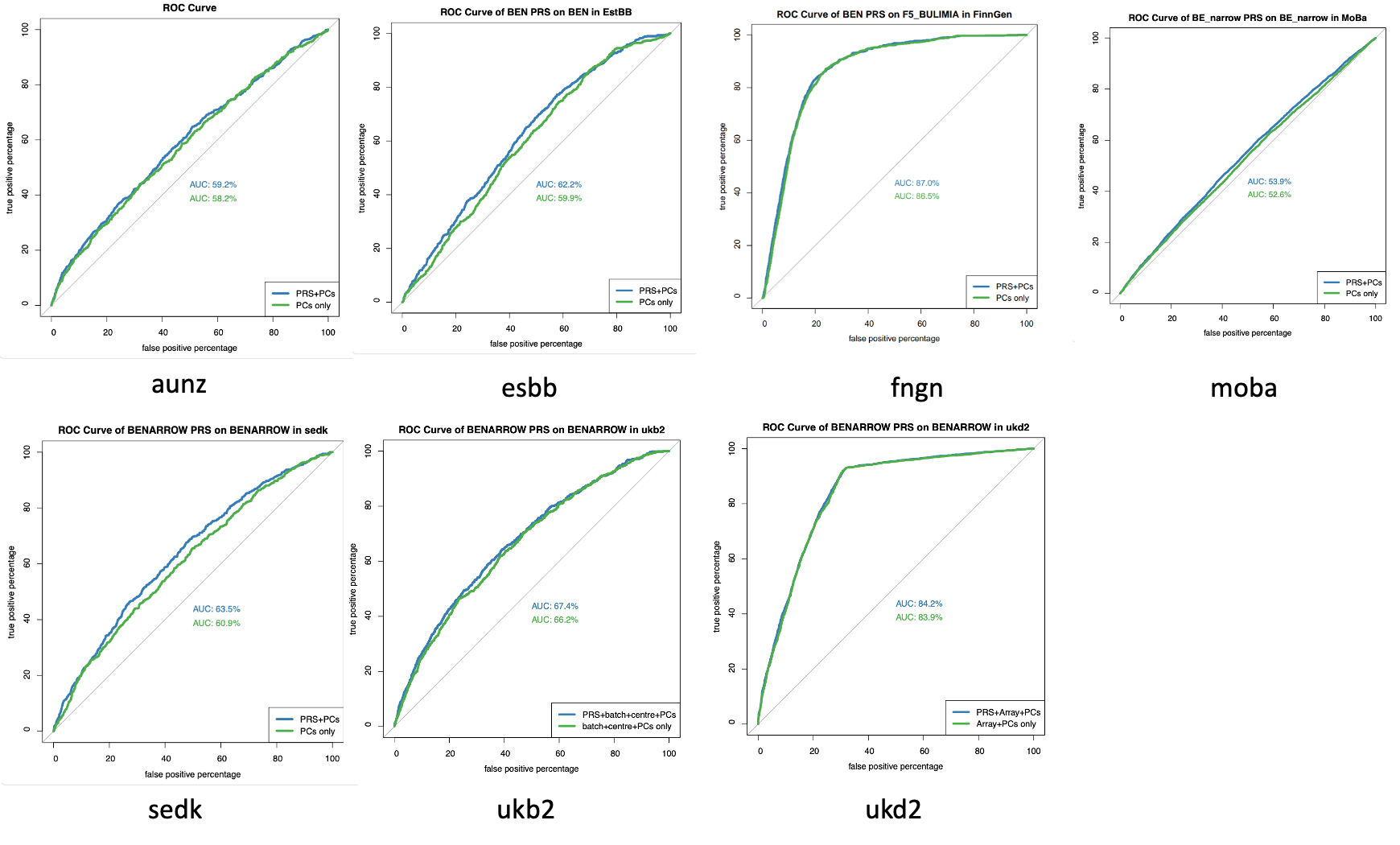

#### Supplementary Figure 23f: Decile plots and area under the receiver operating characteristic curve for the association of BE-NARROW PRS with BE-BROAD

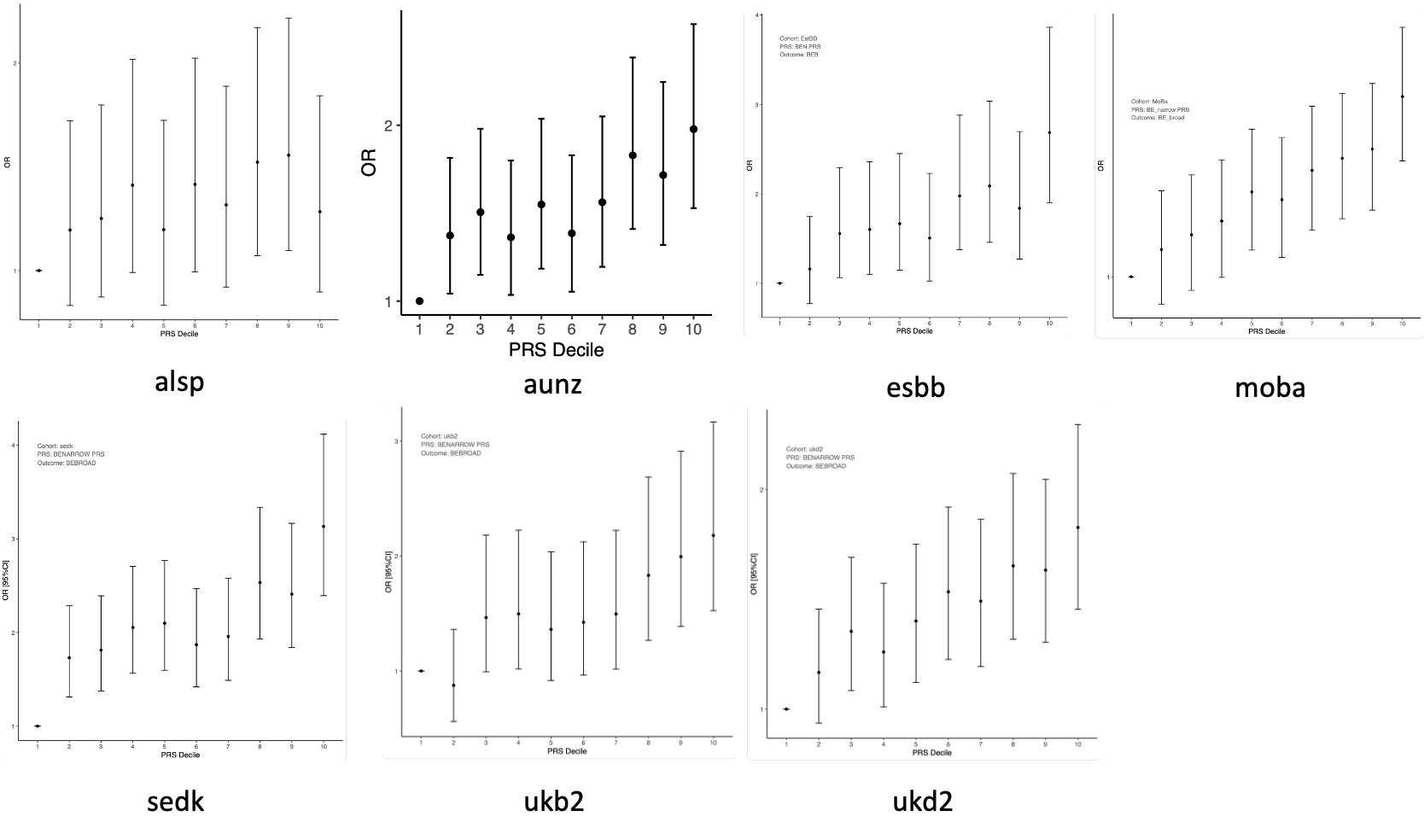

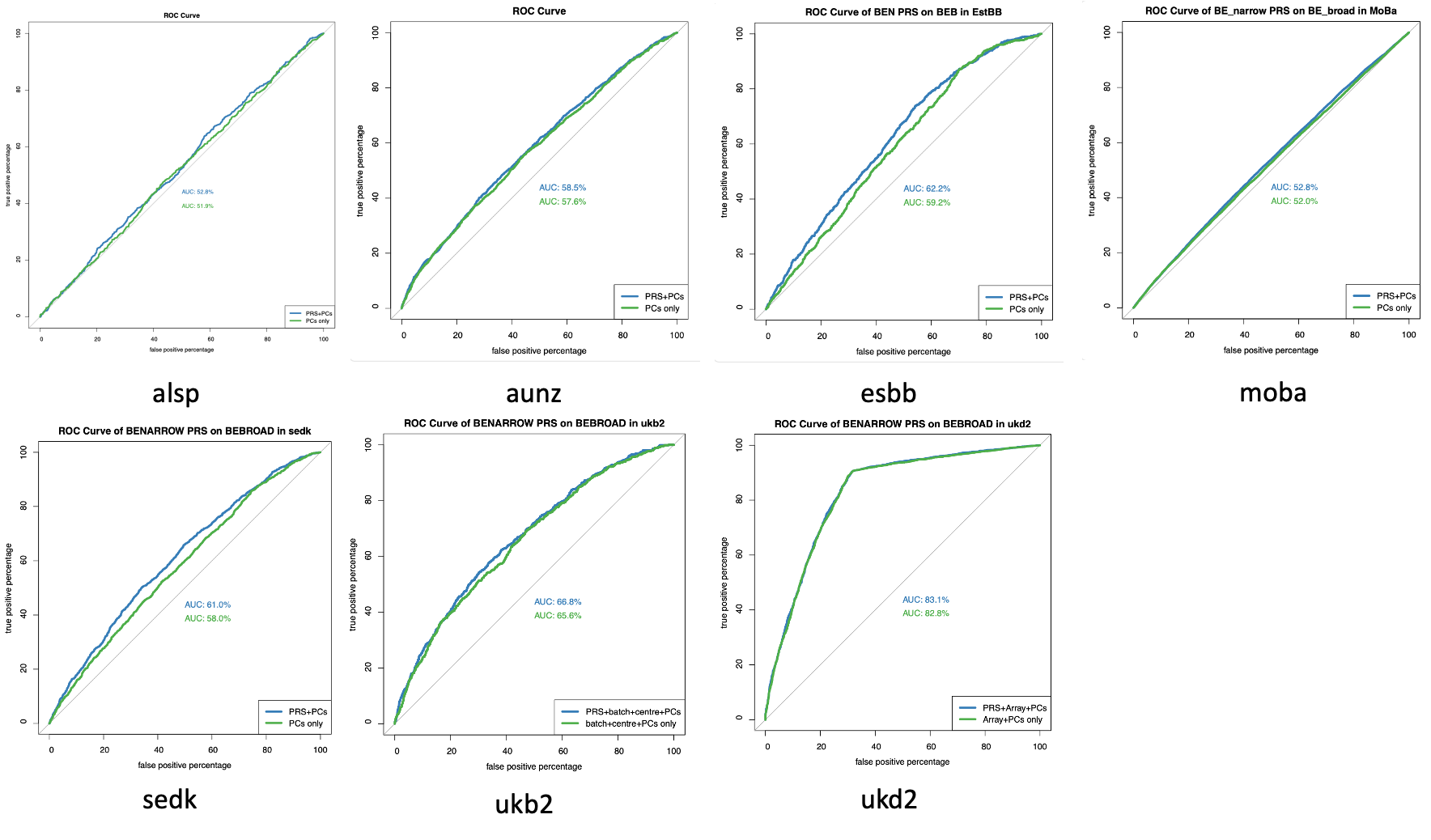

#### Supplementary Figure 23g: Decile plots and area under the receiver operating characteristic curve for the association of BE-NARROW PRS with AN

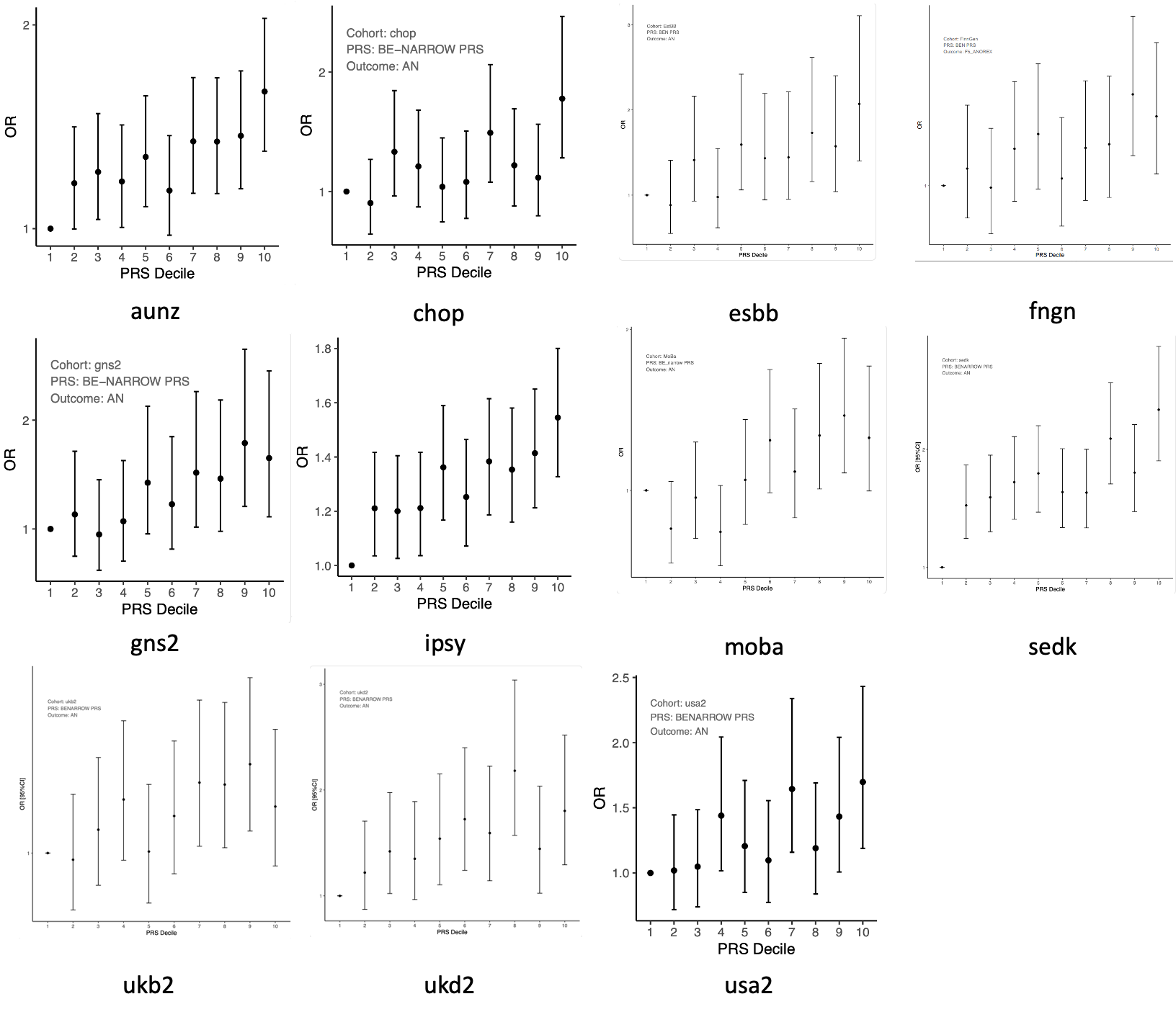

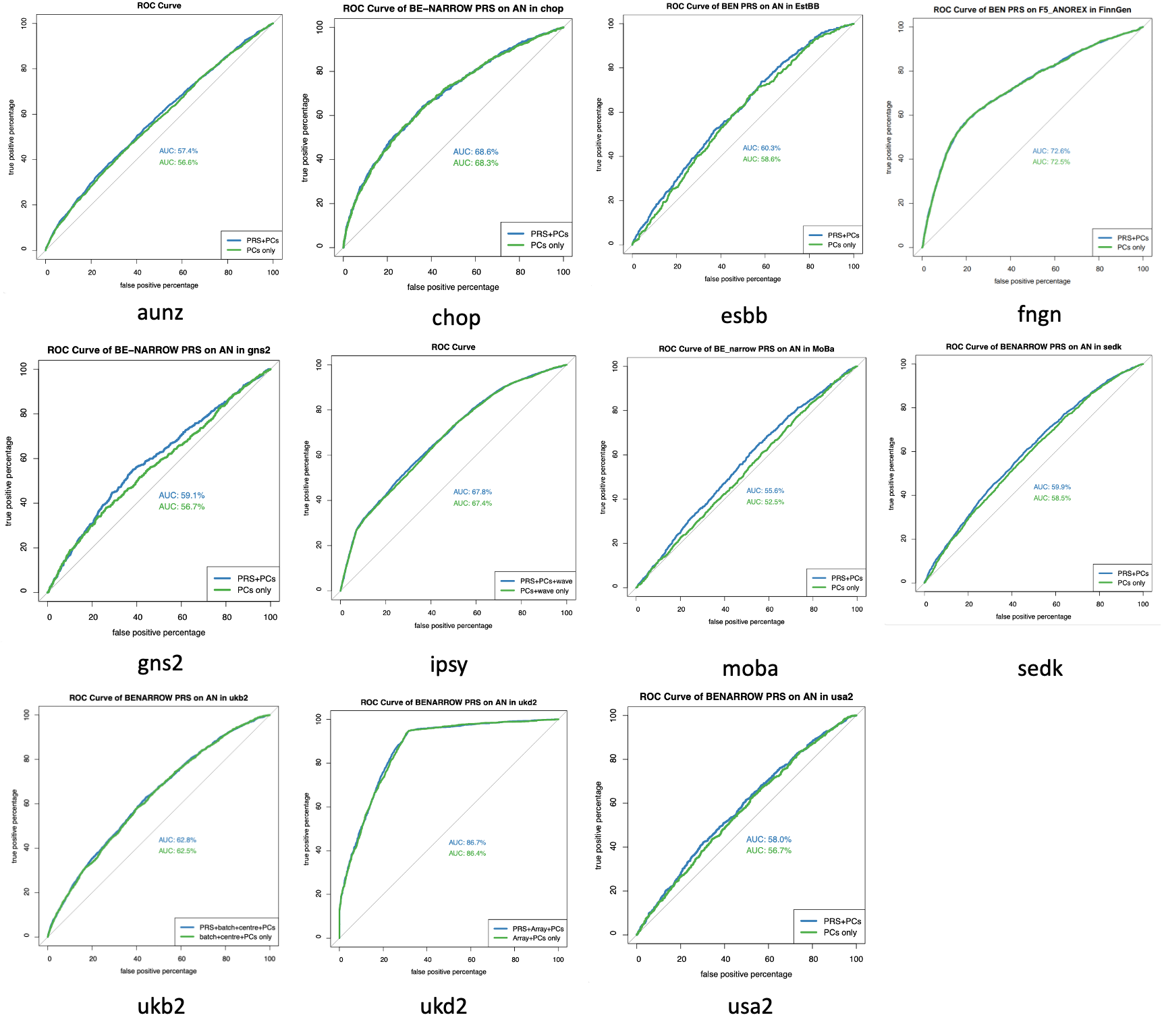

#### Supplementary Figure 23h: Decile plots and area under the receiver operating characteristic curve for the association of AN PRS with BE-BROAD

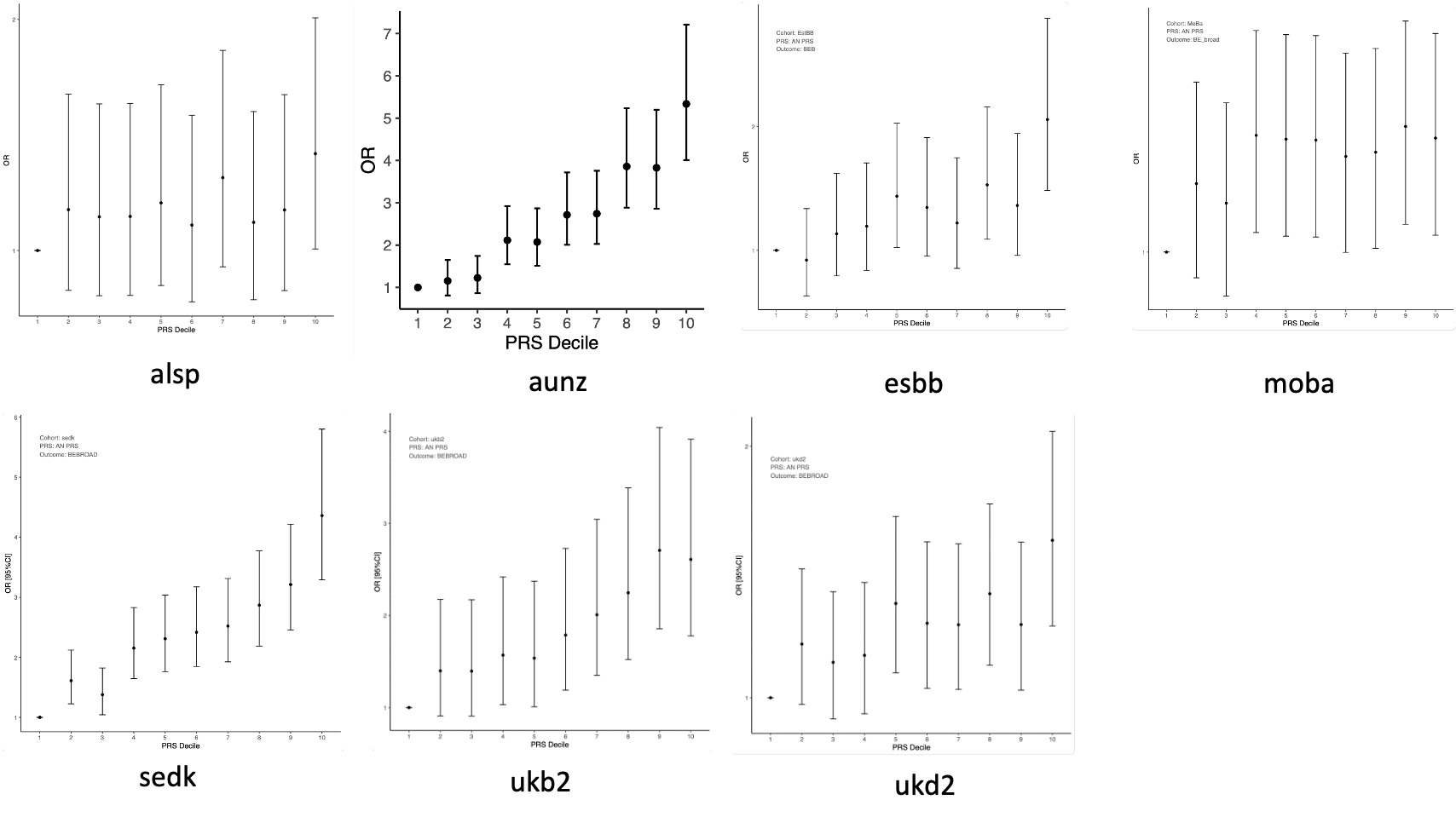

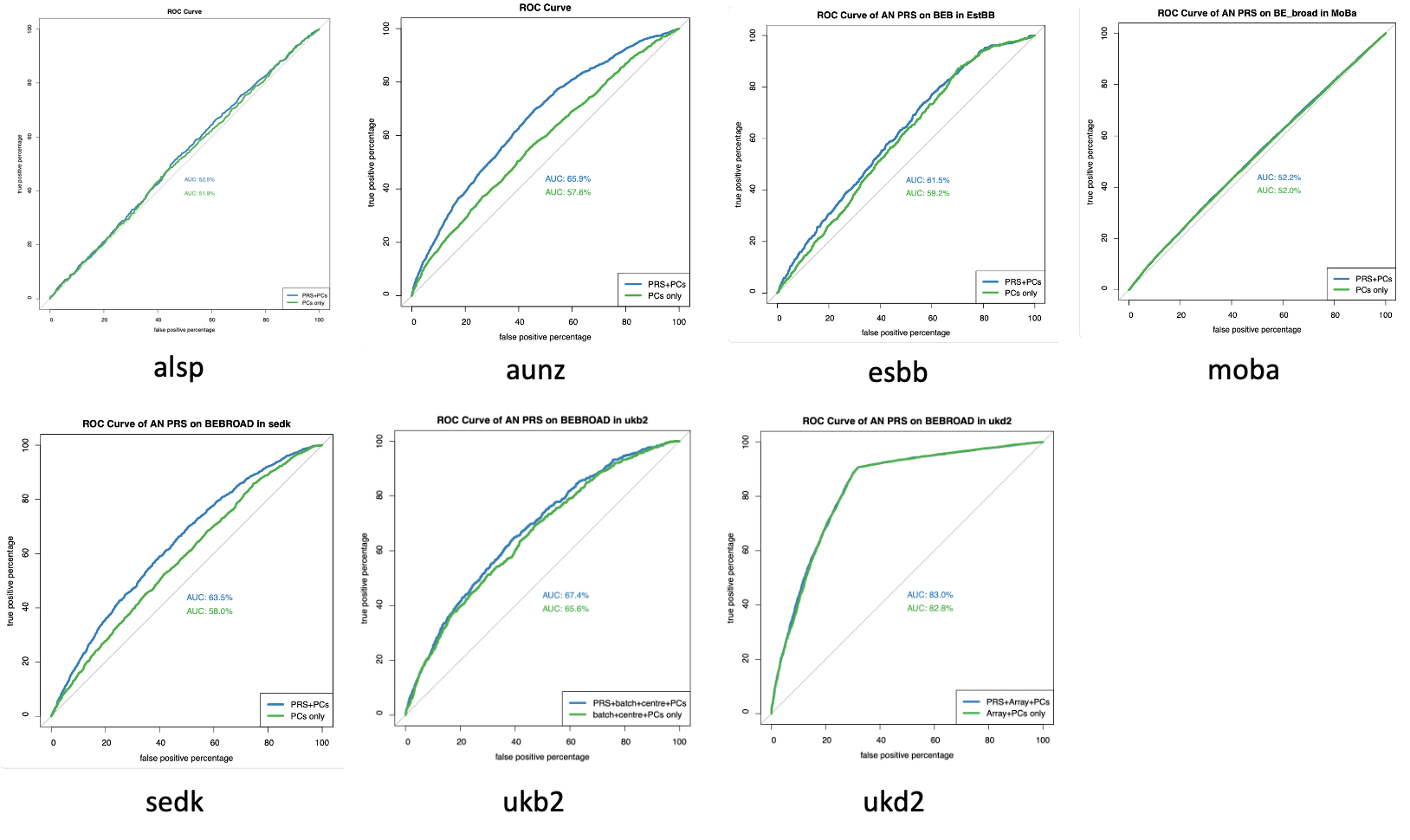

#### Supplementary Figure 23i: Decile plots and area under the receiver operating characteristic curve for the association of AN PRS with BE-NARROW

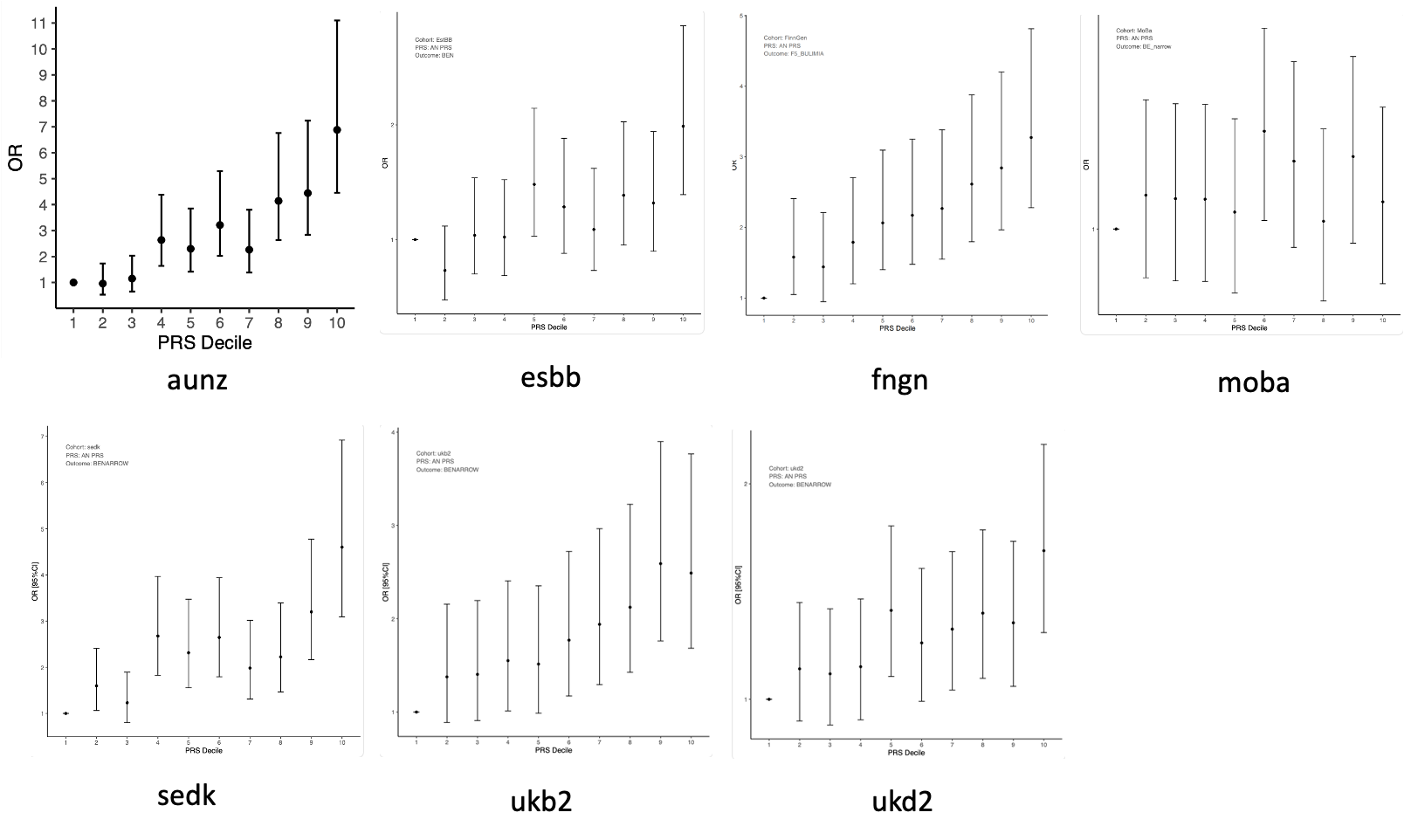

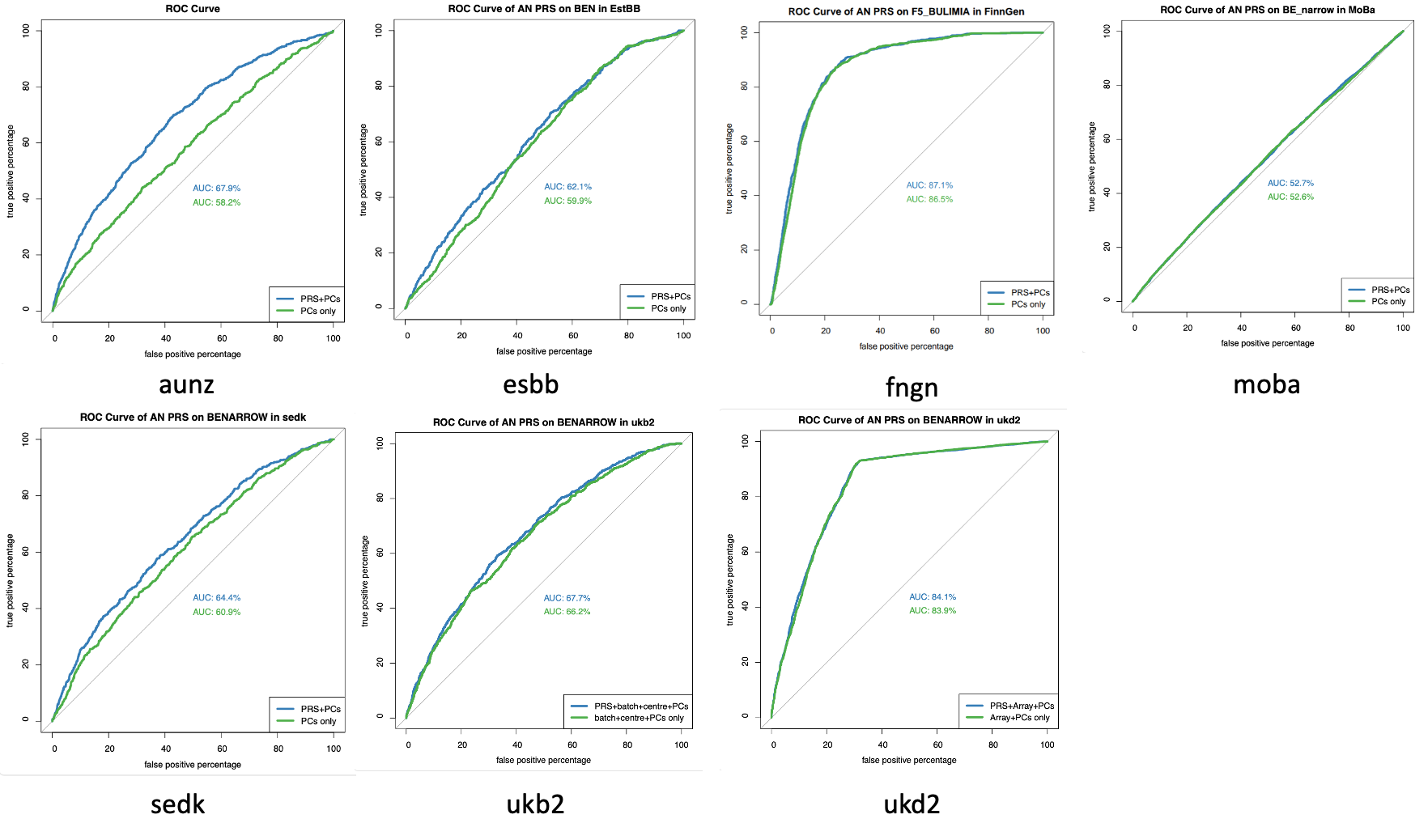

### Supplementary Figure 24: Cross-sex polygenic scoring.

Decile plots depict the odds ratio (OR) of each decile of female-based polygenic risk scores of BE-BROAD on the BE-BROAD outcome in males, and female-based polygenic risk scores of AN on the AN outcome in males, with the lowest decile serving as the reference group. The receiver operating characteristic (ROC) curve is a graphical illustration of the performance of a regression model to predict the binary outcome (i.e., case/control status of BE-BROAD or AN here). If a regression model has a ROC curve overlapping with the diagonal grey line in the ROC curve plot, it indicates that the regression model is no better than chance to differentiate cases and controls with an area under the curve (AUC) of 0.5. An AUC greater than 0.5 suggests that the prediction performance of a model is better than chance and an AUC of 1 indicates perfect performance. In each ROC curve plot, ROC curves of two models are plotted: one for the baseline model (e.g., genetic principal components (PCs) as predictor variables) and one for the PRS model (i.e., both PRS and baseline predictors are included as predictor variables). The precision-recall (PR) curve is another graphical plot to examine predictive performance and performs more accurate evaluation for imbalance datasets. In contrast to the ROC curve which plots sensitivity (true positives detected/all true positives) and 1- specificity (false positives detected/all true negatives), the PR curve plots precision (true positives detected/all detected positives) and recall (same as sensitivity). PR curves are also plotted for both baseline and PRS models, with a greater PR AUC value indicating better predictive performance.

#### Supplementary Figure 24a: Decile plots, area under the receiver operating characteristic curve, and area under the precision-recall curve for the association of female BE-BROAD PRS with BE-BROAD in males

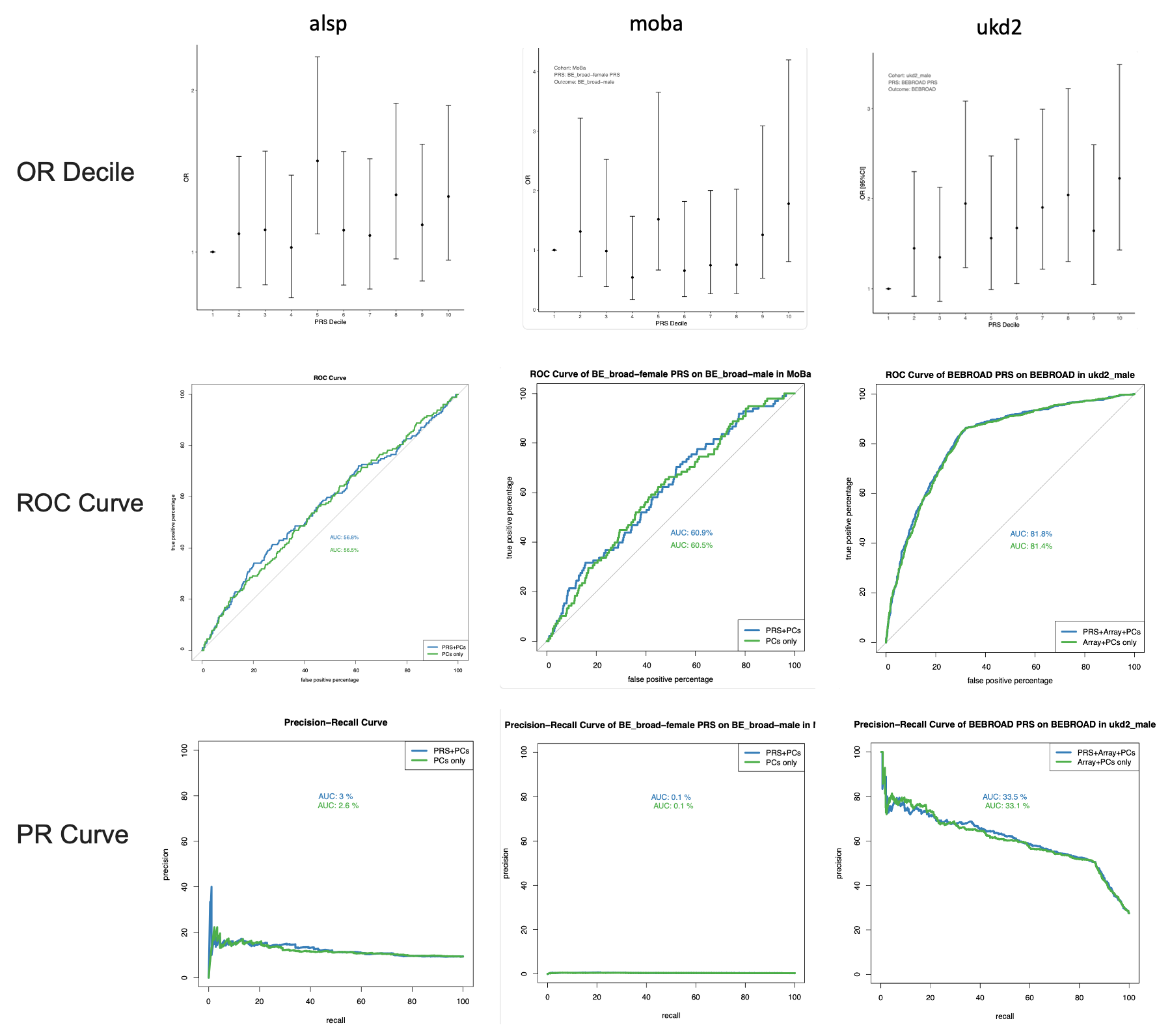

#### Supplementary Figure 24b: Decile plots, area under the receiver operating characteristic curve, and area under the precision-recall curve for the association of female AN PRS with AN in males

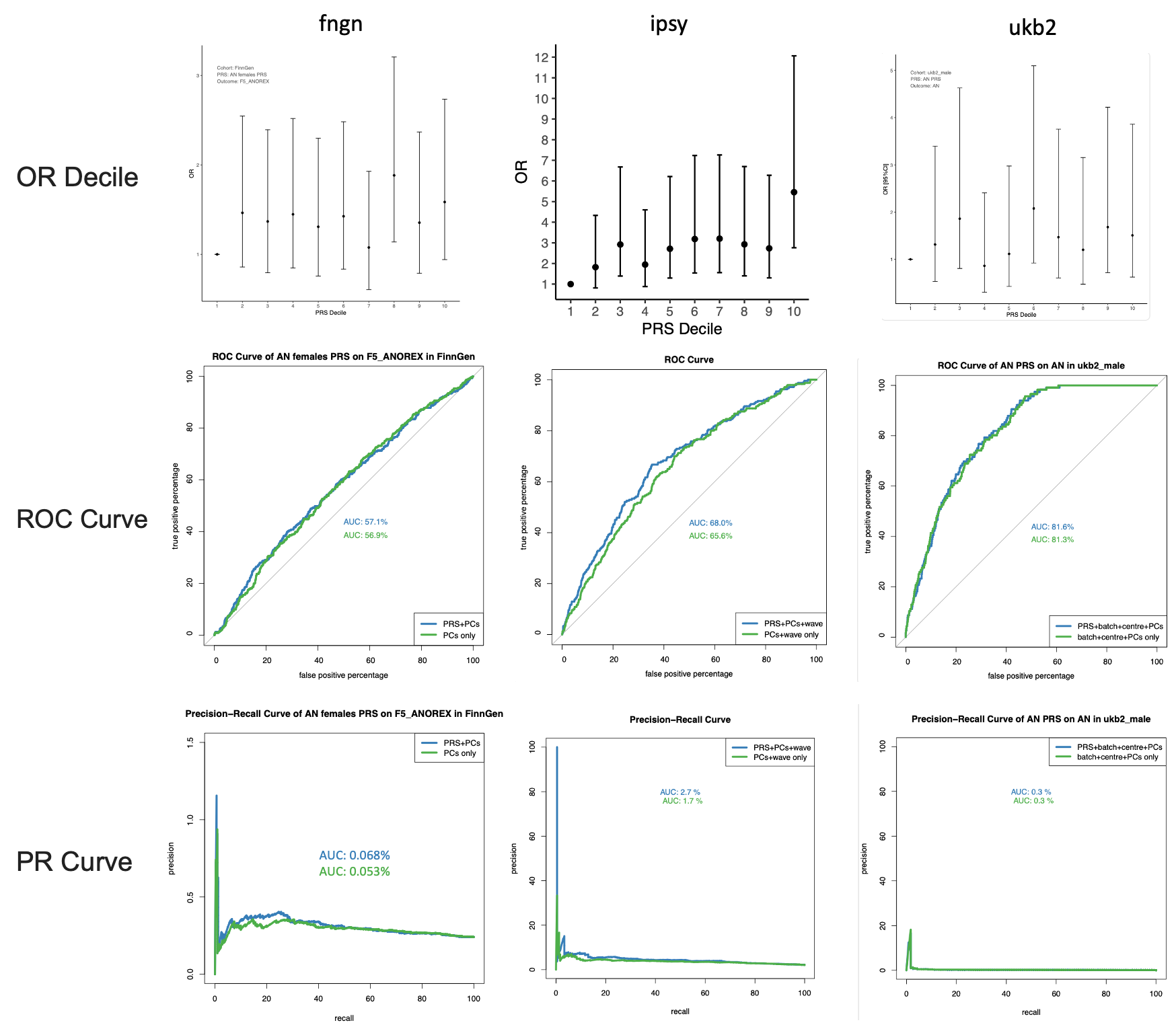

### Supplementary Figure 25: Polygenic scoring across comorbid AN and BE-BROAD (AN_BEB), BE-BROAD-only, and AN-only subgroups.

The plots depict PRS differences (i.e., beta values) for AN PRS, BE-BROAD PRS, and BE-NARROW PRS between each subgroup and the control group across four study cohorts (aunz, sedk, ukb2, ukd2), adjusting for genetic principal components (PCs). Overall, the comorbid AN_BEB and AN-only groups exhibit similar high average AN PRS, with the AN-only group having the lowest BE PRSs. The BE-BROAD-only group has the lowest AN PRS but similarly high BE PRSs as the AN_BEB group.

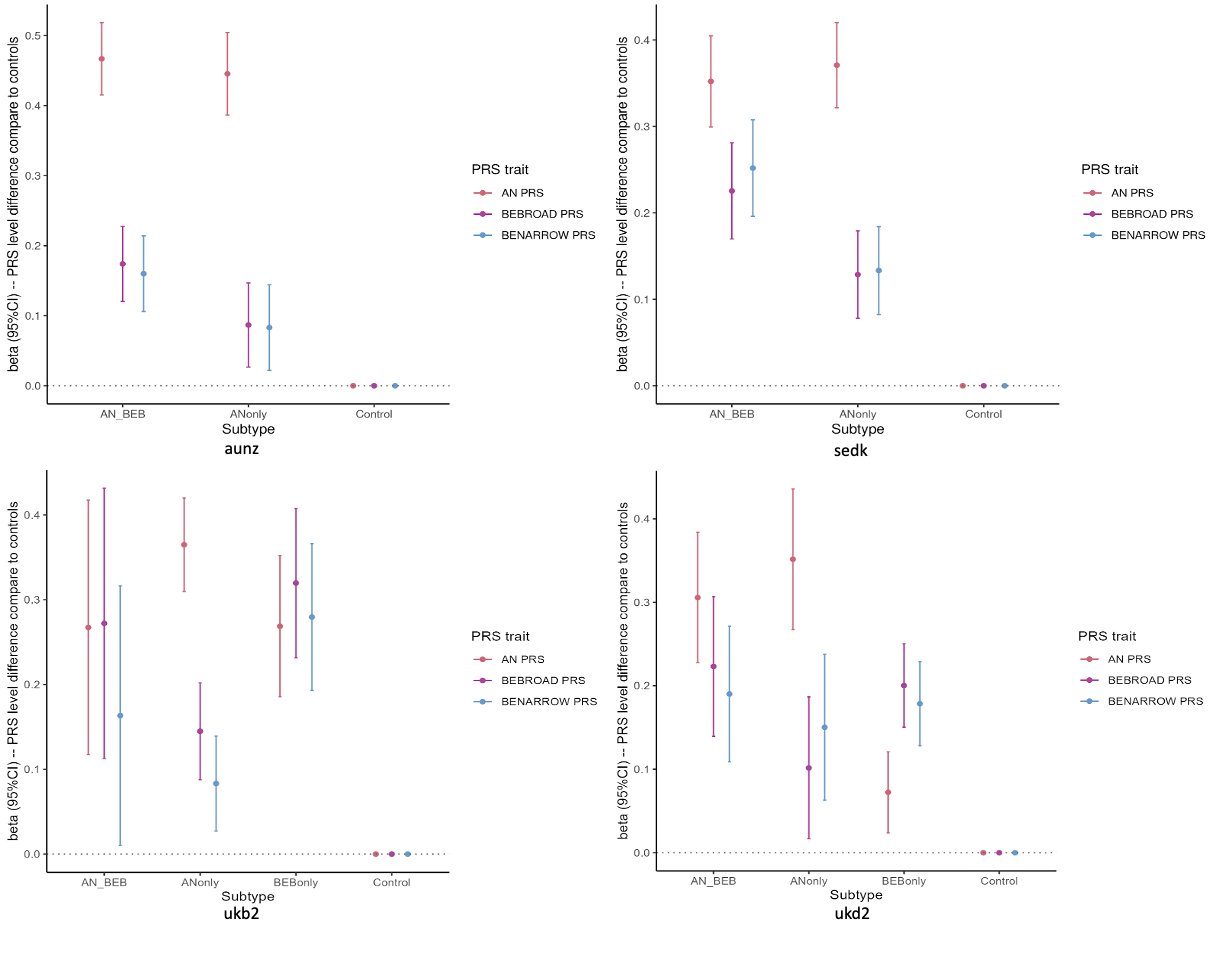

### Supplementary Figure 26: Graphic representation of the GWAS-by-subtraction model.

Circles represent the latent (anthropometric and non-anthropometric) genetic components of body mass index and binge eating. Squares represent the observed SNP estimates obtained from the GWAS summary statistics of body mass index and binge eating (broadly defined; BE-BROAD). Single-headed arrows indicate linear regressions pointing from the independent variable to the dependent variable, and double-headed arrows indicate variance or covariance relationships.

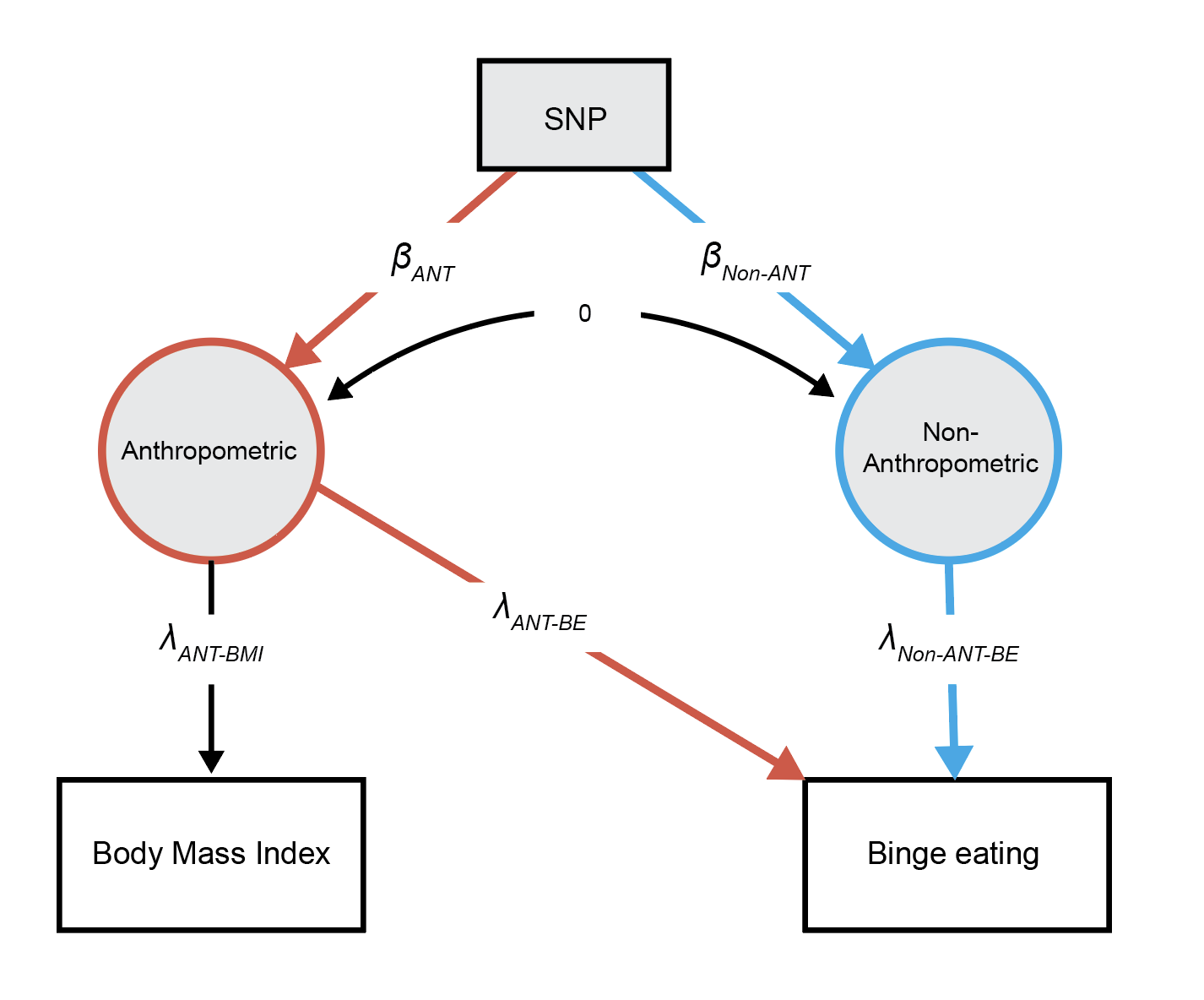

### Supplementary Figure 27: Forest and scatter plots from Mendelian randomisation analyses with BE-BROAD.

(a, b) BE-BROAD acting on BMI; (c, d) *NonBMI* BE-BROAD acting on BMI; (e, f) BMI acting on BE-BROAD; (g, h) BMI acting on *NonBMI* BE-BROAD. Plots a, c, e, and g show the effect size (beta) and standard error of different Mendelian randomisation analyses of a given exposure on a given outcome. Plots b, d, f, and h show the effect (on the beta scale) and standard error of the effect of each variant (dots) on both exposure and outcome, with regression slopes from different Mendelian randomisation analyses.

#### Supplementary Figure 27a

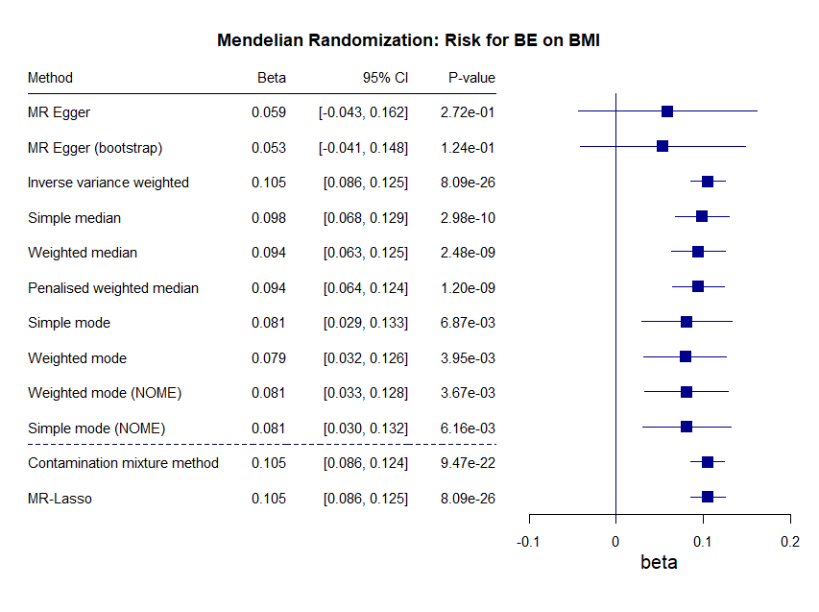

#### Supplementary Figure 27b

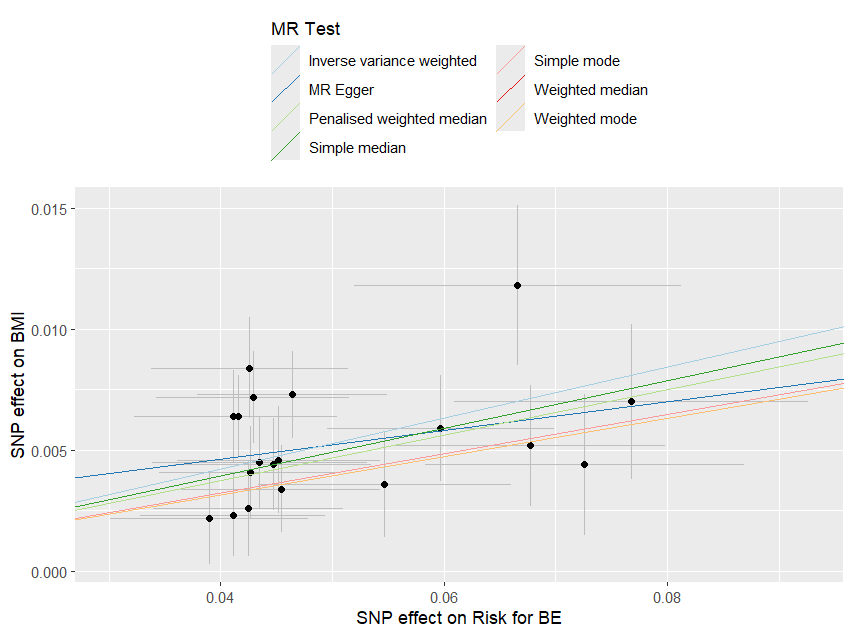

#### Supplementary Figure 27c

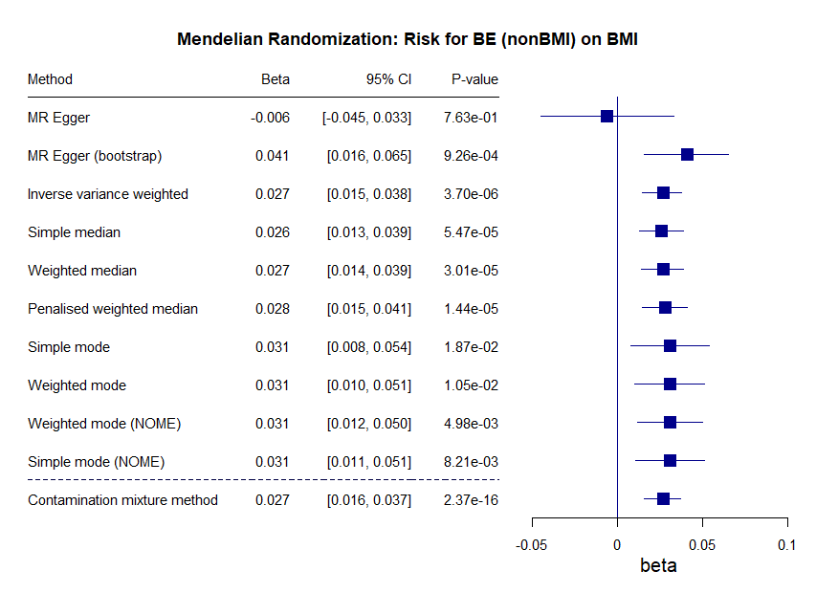

#### Supplementary Figure 27d

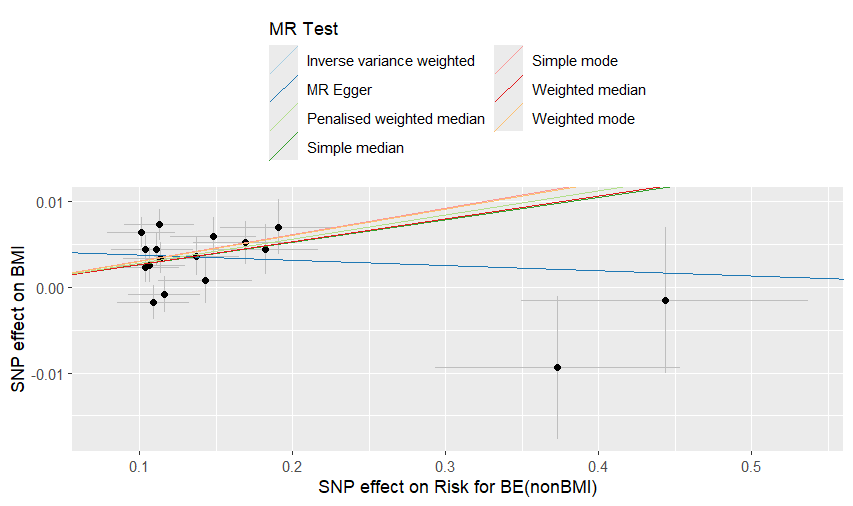

#### Supplementary Figure 27e

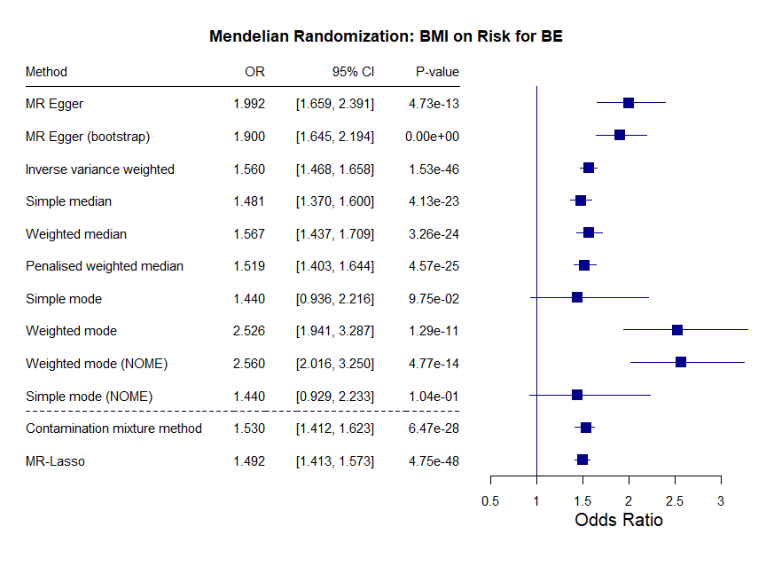

#### Supplementary Figure 27f

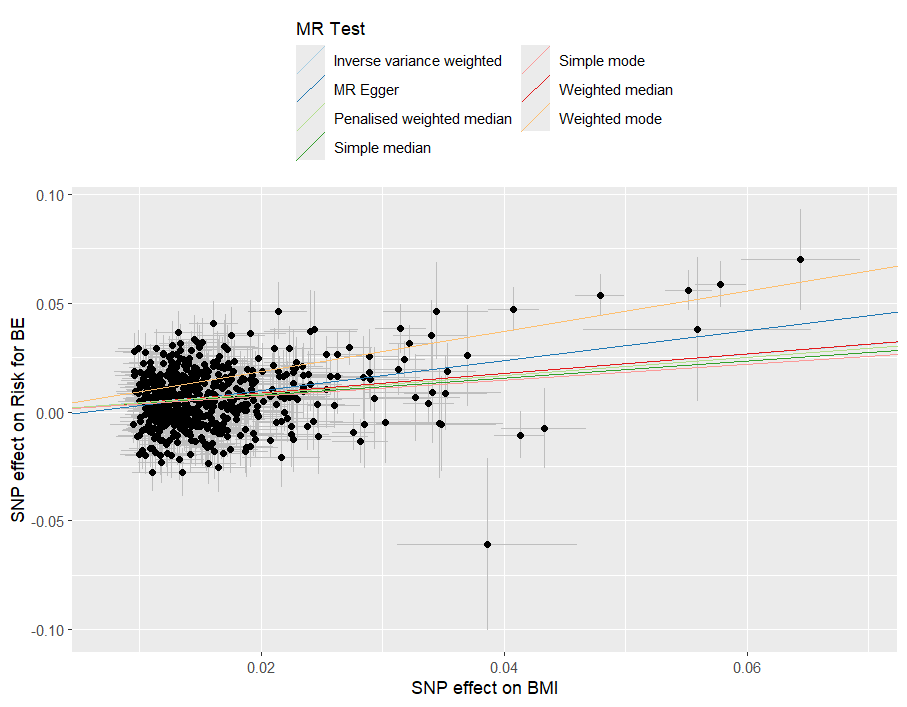

#### Supplementary Figure 27g

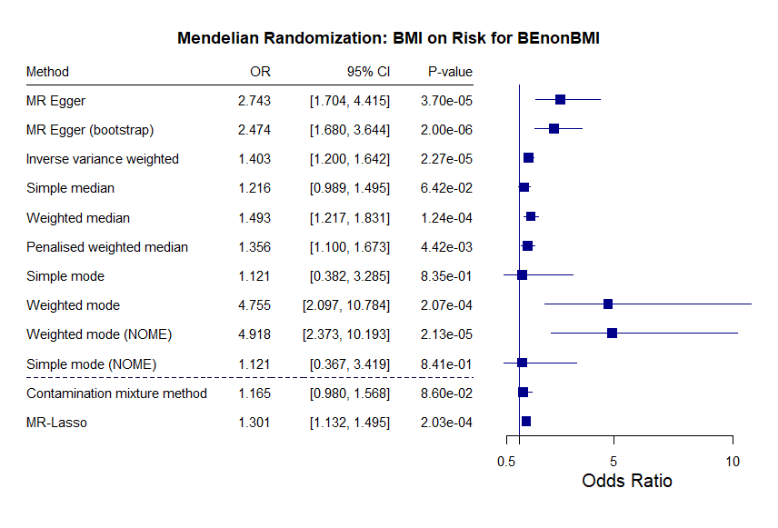

#### Supplementary Figure 27h

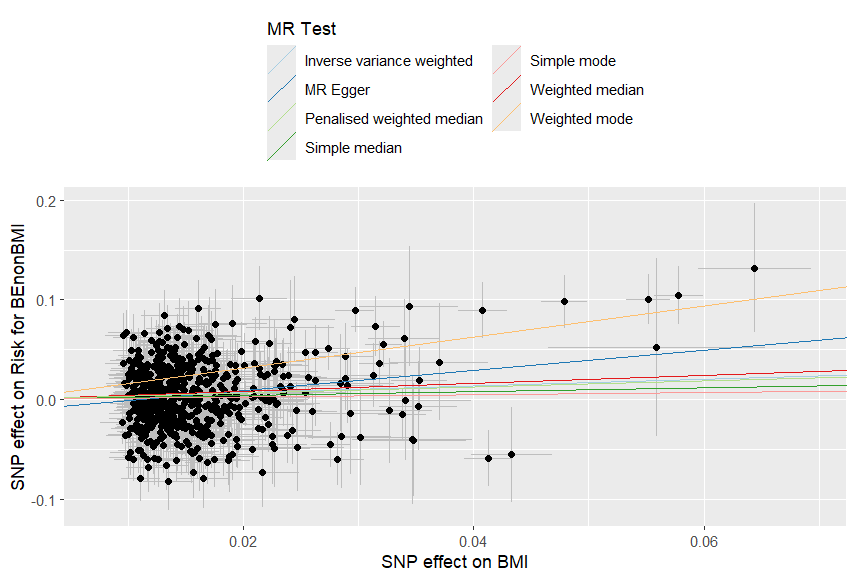

### Supplementary Figure 28: Forest and scatter plots from Mendelian randomisation analyses with AN.

(a, b) AN acting on BMI; (c, d) *NonBMI* AN acting on BMI; (e, f) BMI acting on AN; (g, h) BMI acting on *NonBMI* AN. Plots a, c, e, and g show the effect size (beta) and standard error of different Mendelian randomisation analyses of a given exposure on a given outcome. Plots b, d, f, and h show the effect (on the beta scale) and standard error of the effect of each variant (dots) on both exposure and outcome, with regression slopes from different Mendelian randomisation analyses.

#### Supplementary Figure 28a

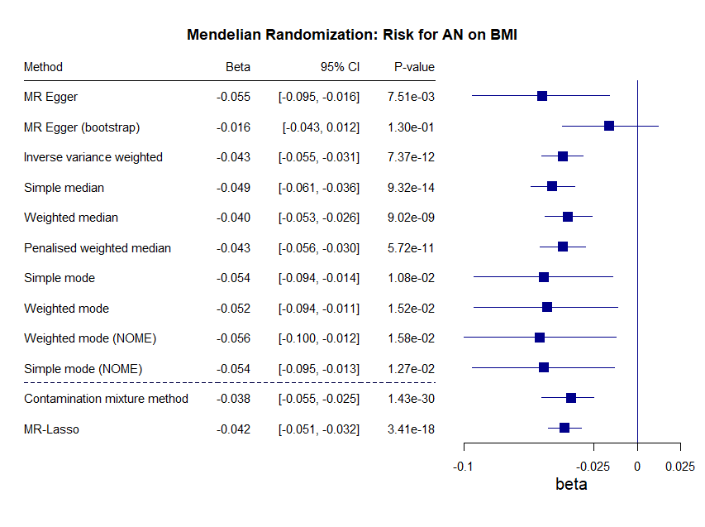

#### Supplementary Figure 28b

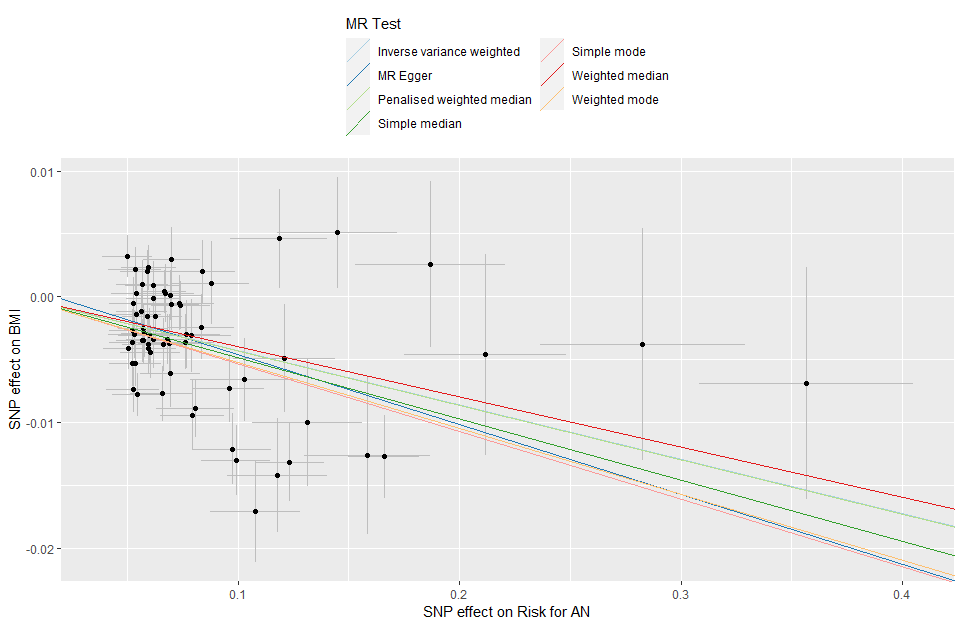

#### Supplementary Figure 28c

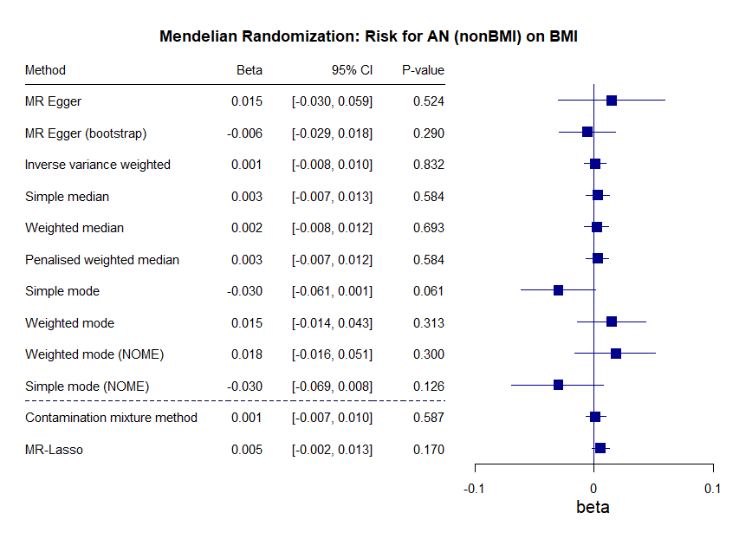

#### Supplementary Figure 28d

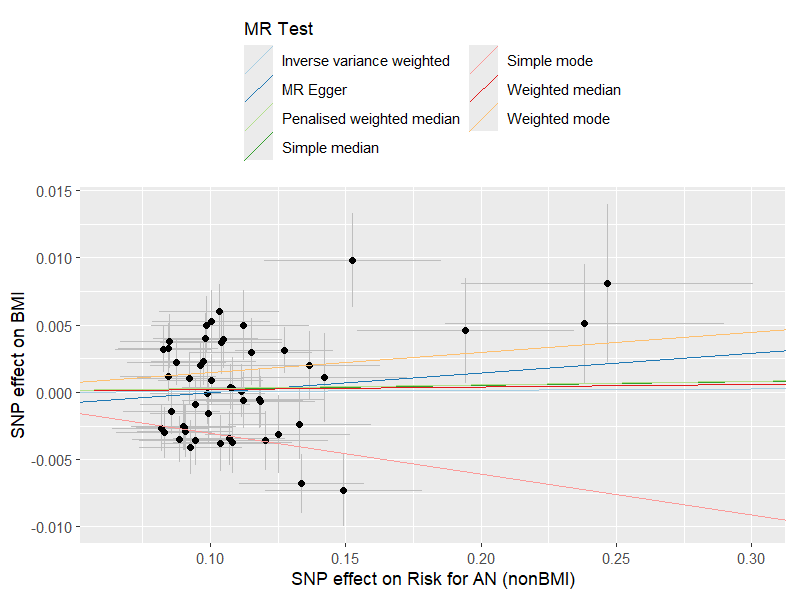

#### Supplementary Figure 28e

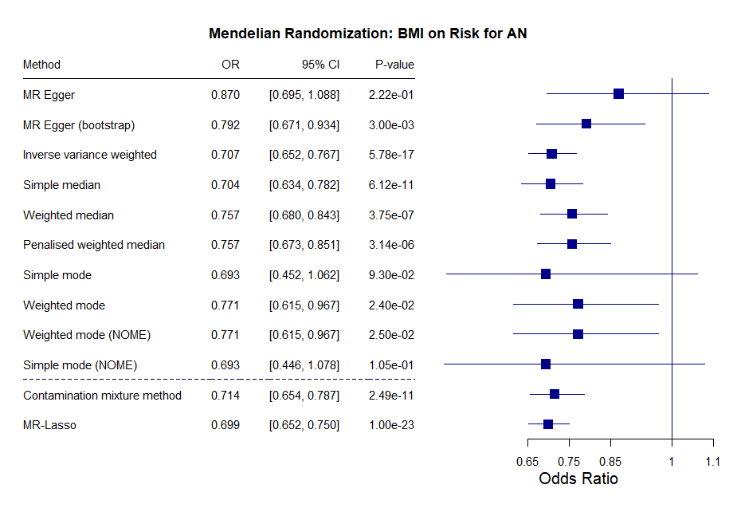

#### Supplementary Figure 28f

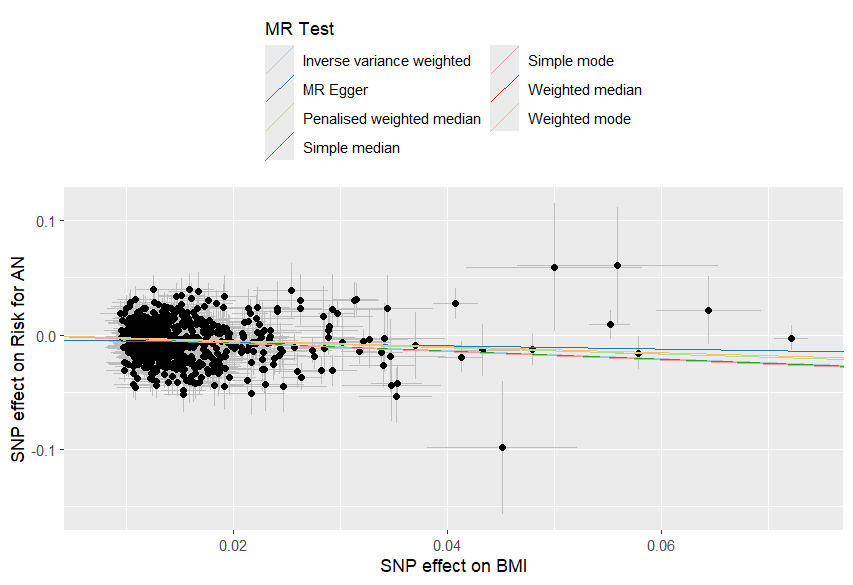

#### Supplementary Figure 28g

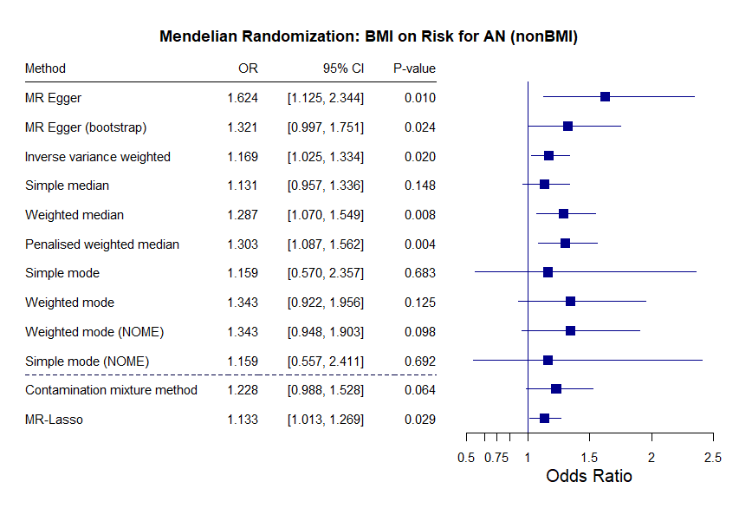

#### Supplementary Figure 28h

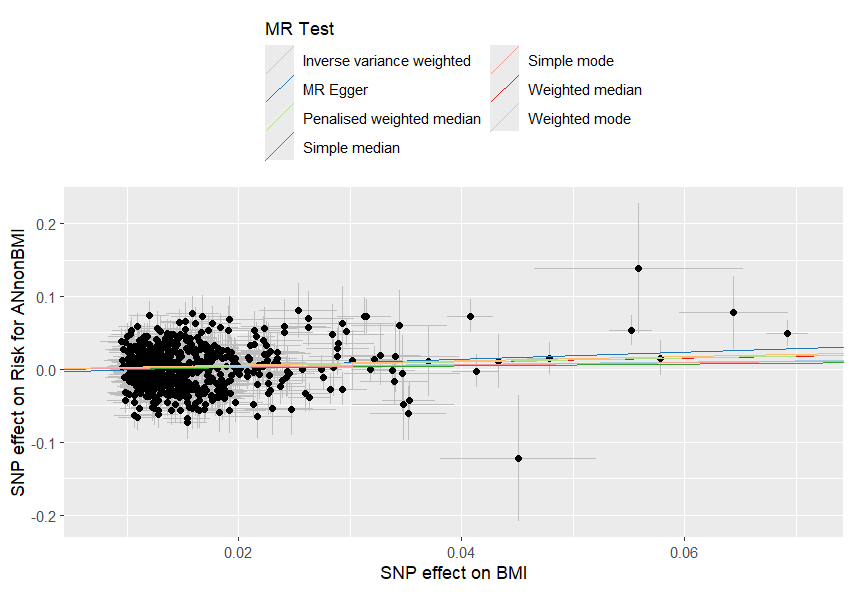
